## Supplemental Content for "Incidence of Long-term Post-acute Sequelae of SARS-CoV-2 Infection Related to Pain and Other Symptoms: A Living Systematic Review and Meta-analysis"

**eAppendix 1.** Literature Search Strategy

**eAppendix 2.** R code

**eAppendix 3.** The system of the Newcastle-Ottawa quality assessment scale in cohort studies

**eTable 1.** Summary of studies included in the meta-analysis

**eTable 2.** The Results of the Newcastle-Ottawa Quality Assessment Scale for Included Studies

**eFigure 1a-e.** Forest Plot, Bubble Plots (follow-up period, age and sex) and Funnel Plot on Abdominal Pain

**eFigure 2a-e.** Forest Plot, Bubble Plots (follow-up period, age and sex) and Funnel Plot on Arthralgia

**eFigure 3a-e.** Forest Plot, Bubble Plots (follow-up period, age and sex) and Funnel Plot on Chest Pain

**eFigure 4a-e.** Forest Plot, Bubble Plots (follow-up period, age and sex) and Funnel Plot on Ear Pain

**eFigure 5a-e.** Forest Plot, Bubble Plots (follow-up period, age and sex) and Funnel Plot on Headache

**eFigure 6a-e.** Forest Plot, Bubble Plots (follow-up period, age and sex) and Funnel Plot on Myalgia

**eFigure 7a-e.** Forest Plot, Bubble Plots (follow-up period, age and sex) and Funnel Plot on Neuralgia

**eFigure 8a-e.** Forest Plot, Bubble Plots (follow-up period, age and sex) and Funnel Plot on Sore Throat

**eFigure 9a-e.** Forest Plot, Bubble Plots (follow-up period, age and sex) and Funnel Plot on Ageusia

**eFigure 10a-e.** Forest Plot, Bubble Plots (follow-up period, age and sex) and Funnel Plot on Alopecia

**eFigure 11a-e.** Forest Plot, Bubble Plots (follow-up period, age and sex) and Funnel Plot on Anorexia

**eFigure 12a-e.** Forest Plot, Bubble Plots (follow-up period, age and sex) and Funnel Plot on Anosmia

**eFigure 13a-e.** Forest Plot, Bubble Plots (follow-up period, age and sex) and Funnel Plot on Anxiety

**eFigure 14a-e.** Forest Plot, Bubble Plots (follow-up period, age and sex) and Funnel Plot on Chills

**eFigure 15a-e.** Forest Plot, Bubble Plots (follow-up period, age and sex) and Funnel Plot on Confusion

**eFigure 16a-e.** Forest Plot, Bubble Plots (follow-up period, age and sex) and Funnel Plot on Cough

**eFigure 17a-e.** Forest Plot, Bubble Plots (follow-up period, age and sex) and Funnel Plot on Depression

**eFigure 18a-e.** Forest Plot, Bubble Plots (follow-up period, age and sex) and Funnel Plot on Diarrhea

**eFigure 19a-e.** Forest Plot, Bubble Plots (follow-up period, age and sex) and Funnel Plot on Dyspnea

**eFigure 20a-e.** Forest Plot, Bubble Plots (follow-up period, age and sex) and Funnel Plot on Fatigue

**eFigure 21a-e.** Forest Plot, Bubble Plots (follow-up period, age and sex) and Funnel Plot on Fever

**eFigure 22a-e.** Forest Plot, Bubble Plots (follow-up period, age and sex) and Funnel Plot on Insomnia

**eFigure 23a-e.** Forest Plot, Bubble Plots (follow-up period, age and sex) and Funnel Plot on Memory impairment

**eFigure 24a-e.** Forest Plot, Bubble Plots (follow-up period, age and sex) and Funnel Plot on Nasal blockage

**eFigure 25a-e.** Forest Plot, Bubble Plots (follow-up period, age and sex) and Funnel Plot on Nausea

**eFigure 26a-e.** Forest Plot, Bubble Plots (follow-up period, age and sex) and Funnel Plot on Palpitation

**eFigure 27a-e.** Forest Plot, Bubble Plots (follow-up period, age and sex) and Funnel Plot on Rhinorrhea

**eFigure 28a-e.** Forest Plot, Bubble Plots (follow-up period, age and sex) and Funnel Plot on Sneezing

**eFigure 29a-e.** Forest Plot, Bubble Plots (follow-up period, age and sex) and Funnel Plot on Sputum

**eFigure 30a-e.** Forest Plot, Bubble Plots (follow-up period, age and sex) and Funnel Plot on Vertigo (Dizziness)

**eFigure 31a-e.** Forest Plot, Bubble Plots (follow-up period, age and sex) and Funnel Plot on Vomiting

**eFigure 32a-e.** Forest Plot, Bubble Plots (follow-up period, age and sex) and Funnel Plot on Weakness

**eFigure 33a-e.** Forest Plot, Bubble Plots (follow-up period, age and sex) and Funnel Plot on Weight loss

### eAppendix1. Literature Search Strategy

1) **PubMed**

“long COVID”[All Fields] OR “long COVID-19”[All Fields] OR “long-haul covid”[All Fields] OR “Chronic COVID syndrome”[All Fields] OR “Post-COVID-19 syndrome”[All Fields] OR (“Long-term complications”[All Fields] AND “COVID”[All Fields]) OR (“long-term consequences”[All Fields] AND ”COVID”[All Fields]) OR (“long-term sequelae”[All Fields] AND “COVID”[All Fields])

2) **EMBASE**

(‘long covid’ OR ‘long covid -19’ OR ‘long-haul covid’ OR ‘Chronic covid syndrome’ OR ‘Post-covid -19 syndrome’ OR (‘Long-term complications’AND ‘covid’) OR (‘long-term consequences’ AND ‘covid’) OR (‘long-term sequelae’ AND ‘covid’)

3) **Scopus**

( "long COVID" OR "long COVID-19" OR "long-haul covid" OR "Chronic COVID syndrome" OR "Post-COVID-19 syndrome" OR ( "Long-term complications" AND "COVID" ) OR ( "long-term consequences" AND "COVID" ) )

4) **CHINAL**

long COVID [TI] OR long COVID-19 [TI] OR long-haul covid [TI] OR Chronic COVID syndrome [TI] OR Post-COVID-19 syndrome [TI] OR (Long-term complications of COVID [TI]) OR (long-term consequences of COVID [TI]) OR (long-term sequelae of COVID [TI])

5) **MedRχiv and BioRχiv**

( "long COVID" OR "long COVID-19" OR "long-haul covid" OR "Chronic COVID syndrome" OR "Post-COVID-19 syndrome" OR ( "Long-term complications" AND "COVID" ) OR ( "long-term consequences" AND "COVID" ) )

**eAppendix 2. R code**

```
library(meta)
# meta-analysis of proportions, inverse variance method, DerSimonian-Laird estimator for tau^2, Logit
#transformation
meta<-metaprop(n, N, data=Abdominal_pain,studlab =paste(Author),method="Inverse",
               comb.fixed = F,comb.random = T, method.tau="DL", sm="PLOGIT")
meta
forest(meta)

# Mixed-Effects model meta-regression for followup, Age and Sex as covariates
metareg1<-metareg(meta,followup)
metareg1
metareg2<-metareg(meta,meanAge)
metareg2
metareg3<-metareg(meta, maleGender)
metareg3

bubble1<-bubble(metareg1)
bubble2<-bubble(metareg2)
bubble3<-bubble(metareg3)

# Plot to assess funnel plot asymmetry
funnel(meta)
# Egger's linear regression test of funnel plot asymmetry
metabias(meta, method.bias = "linreg", k.min=3)
```

### eAppendix 3. The system of Newcastle-Ottawa quality assessment scale in cohort studies

Note: A study can be awarded a maximum of one star for each numbered item within the Selection and Outcome categories. A maximum of two stars (*) can be given for Comparability

**Selection**

1) Representativeness of the exposed cohort
a) truly representative of the average _______________ (describe) in the community *
b) somewhat representative of the average ______________ in the community *
c) selected group of users eg nurses, volunteers
d) no description of the derivation of the cohort

2) Selection of the non exposed cohort
a) drawn from the same community as the exposed cohort *
b) drawn from a different source
c) no description of the derivation of the non exposed cohort

3) Ascertainment of exposure
a) secure record (eg surgical records) *
b) structured interview *
c) written self report
d) no description

4) Demonstration that outcome of interest was not present at start of study
a) yes *
b) no

**Comparability**

1) Comparability of cohorts on the basis of the design or analysis
a) study controls for _____________ (select the most important factor) *
b) study controls for any additional factor * (This criteria could be modified to indicate specific control for a second important factor.)

**Outcome**

1) Assessment of outcome

a) independent blind assessment *

b) record linkage *

c) self report

d) no description

2) Was follow-up long enough for outcomes to occur

a) yes (select an adequate follow up period for outcome of interest) *

b) no

3) Adequacy of follow up of cohorts

a) complete follow up - all subjects accounted for *

b) subjects lost to follow up unlikely to introduce bias - small number lost - > 80 % (select an adequate %) follow up, or description provided of those lost) *

c) follow up rate < 80% (select an adequate %) and no description of those lost

d) no statement

**eTable 1. Summary of studies included in the meta-analysis**

| First author | Year published | Location | Patinet setting | Sample size (n) | Diagnotic criteria of SARS-CoV-2 | Respiratory support | Age (mean or median) | Sex, % male | Follow-up period, (maximum) (month) | Patient follow-up (n/total) |
|---|---|---|---|---|---|---|---|---|---|---|
| Arnold, D[19] | 2020 | UK | Non-hospitalized | 110 | Positive PCR and clinico-radiological diagnosis | Oxygen alone, CPAP or IV | 60 | 56 | 3 | 110/110 |
| Boscolo-Rizzo, P[20] | 2020 | Italy | Hospitalized | 187 | PCR | NS | 56 | 44.9 | 1 | 187/202 |
| Carfi, A[21] | 2020 | Italy | Hospitalized | 143 | RT-PCR | Oxygen alone, CPAP or IV | 56.5 | 62.9 | 2 | 143/143 |
| Carvalho-Schneider, C[22] | 2021 | France | Hospitalized and non-hospitalized | 150 | RT-PCR | NS | 48.8 | 56 | 2 | 130/293 |
| Cheng, DO[23] | 2020 | UK | Hospitalized | 109 | PCR and clinical symptoms | NS | 73 | 55.8 | 2.3 | 109/1946 |

| | | | | | | | | | | |
|---|---|---|---|---|---|---|---|---|---|---|
| Chiesa-Estomb, CN[24] | 2020 | Spain | Hospitalized and non-hospitalized | 751 | RT-PCR and serology | NS | 41 | 36.5 | 1.6 | 751/1231 |
| Cirulli, E[25] | 2020 | USA | Hospitalized and non-hospitalized | 357 | No description | NS | 56 | 35.9 | 3 | 216/357 |
| Davis, HE[26] | 2020 | 56 countries | Hopitalized and non-hospitalized | 3762 | PCR, antigen, antibody positive | NS | 50.5 | 78.9 | 7 | 3762/3762 |
| Dennis, A[27] | 2020 | UK | Hospitalized and non-hospitalized | 201 | RT-PCR, serology and symptoms | NS | 44 | 30.3 | 5.3 | 201/unreported |
| Eiros, R[28] | 2020 | Spain | Hopitalized and non-hospitalized | 139 | RT-PCR and serology | Oxygen | 52 | 28.1 | 2.8 | 139/142 |
| Geortz, YNJ[29] | 2020 | Netherlands | Hopitalized and non-hospitalized | 2113 | PCR and CT | NS | 47 | 14.7 | 3.2 | 2113/2113 |
| Halpin, SJ[30] | 2020 | UK | Hospitalized | 100 | PCR positive | Oxygen alone, CPAP or IV | 66.7 | 54 | 2 | 100/191 |

| | | | | | | | | | | |
|---|---|---|---|---|---|---|---|---|---|---|
| Huang, C[31] | 2021 | China | Hospitalized | 1733 | SARS-CoV2 antibody | NS | 57 | 52 | 6.6 | 1733/2469 |
| Klein, H[32] | 2020 | Israel | Hopitalized and non-hospitalized | 112 | RT-PCR positive | NS | 35 | 64.3 | 6 | 112/114 |
| Lavoto, A[33] | 2020 | Italy | except for ICU | 121 | Swab PCR positive | NS | 46.7 | 40.5 | 1.4 | 121/121 |
| Mandal, S[34] | 2020 | UK | Hospitalized | 384 | Swab PCR positive | Oxygen alone, CPAP or IV | 59.9 | 62 | 1.5 | 384/479 |
| Moradian, ST[35] | 2021 | Iran | Hospitalized | 200 | RT-PCR | NS | 55.6 | 80 | 1.5 | 200/300 |
| Neto, DB[36] | 2020 | Brazil | Hospitalized | 545 | RT-PCR | NS | 37.7 | 36.3 | 4 | 545/669 |
| Petersen, M[37] | 2020 | Denmark | Non-hospitalized | 180 | RT-PCR of an oropharyngeal swab | NS | 39.9 | 45.6 | 7 | 180/180 |
| Pilotto, A[38] | 2020 | Italy | Hospitalized | 165 | NS | Oxygen alone, CPAP or IV | 64.8 | 24 | 6 | 165/208 |

| | | | | | | | | | | |
|---|---|---|---|---|---|---|---|---|---|---|
| Poncet-Megemont, L[39] | 2020 | France | Hospitalized and non-hospitalized | 139 | PCR and chest CT | NS | 48.5 | 37.4 | 1.1 | 139/180 |
| Rahmani, H[40] | 2020 | Iran | Hospitalized | 173 | PCR , clinical data and chest CT | Oxygen alone, CPAP or IV | 60 | 53.1 | 1.9 | 173/213 |
| Savarraj, JPJ[41] | 2020 | USA | Hospitalized | 48 | RT-PCR | NS | 50 | 34.3 | 3 | 48/140 |
| Salmon-Ceron, D[42] | 2021 | France | Hopitalized and non-hospitalized | 70 | positive PCR and serology | Oxygen alone, CPAP or IV | 45 | 78.6 | 2 | 70/70 |
| Stavem, K[43] | 2020 | Norway | Non-hospitalized | 451 | PCR | NS | 49.8 | 44 | 4.2 | 451/451 |
| Sudre, C[44] | 2020 | UK, US, Sweden | NS | 4182 | PCR | NS | 42 | 28.5 | 2 | 4182/4182 |
| Tenforde, MW[45] | 2020 | US | Non-hospitalized | 270 | RT-PCR positive | NS | 39.6 | 48.1 | 1 | 270/274 |
| Tomasoni, D[46] | 2020 | Italy | Hospitalized | 105 | NS | Oxygen alone, CPAP or IV | 55 | 73.3 | 3 | 105/105 |

| | | | | | | | | | | |
|---|---|---|---|---|---|---|---|---|---|---|
| Townsend, L[47] | 2020 | Ireland | Hospitalized | 128 | RT-PCR | NS | 49.5 | 53.9 | 1.5 | 128/223 |
| Wang, X[48] | 2020 | China | Hospitalized | 131 | NS | NS | 49 | 45 | 1 | 131/147 |
| Weerahandi, H[49] | 2020 | USA | Hospitalized | 152 | laboratory-confirmed | Oxygen | 62 | 63 | 1.3 | 152/161 |
| Wu, C[50] | 2020 | China | Hospitalized | 370 | RT-PCR | NS | 50.5 | 54.9 | 0.8 | 370/370 |
| Xiong, Q[51] | 2020 | China | Hospitalized | 538 | according to WHO guidance | NS | 52 | 45.5 | 3.6 | 538/2641 |
| Yan, N[52] | 2020 | China | Hospitalized | 337 | RT-PCR positive | NS | 44 | 45.7 | 0.5 | 296/337 |
| Zhao, Y[53] | 2020 | China | Hospitalized | 55 | RT-PCR positive | Oxygen | 47.8 | 58.2 | 3 | 55/55 |

RT-PCR: Reverse Transcription Polymerase Chain Reaction, CPAP: Continuous Positive Airway Pressure, IV: Invasive Ventilation, NS:not specified.

**eTable 2. The Results of the Newcastle-Ottawa Quality Assessment Scale for Included Studies**

| First author | 1) Representativeness | 2) None exposed cohort | 3) Ascertainment of exposure | 4) Demonstration | 5) Comparability | 6) Assessment of outcome | 7) Was follow-up long enough | 8) Adequacy of follow-up | Score |
|---|---|---|---|---|---|---|---|---|---|
| Arnold, D[19] | a | NA | a | b | NA | a | a | a | 6 |
| Boscolo-Rizzo, P[20] | a | NA | b | b | NA | c | a | b | 5 |
| Carfi, A[21] | a | NA | a | a | NA | a | a | a | 6 |
| Carvalho-Schneider, C[22] | a | NA | b | a | NA | a | a | a | 6 |
| Cheng, DO[23] | a | NA | b | a | NA | a | a | a | 6 |
| Chiesa-Estomb, CN[24] | a | NA | b | a | NA | a | a | b | 6 |
| Cirulli, E[25] | a | NA | b | b | NA | c | a | a | 5 |
| Davis, HE[26] | b | NA | b | b | NA | c | a | a | 5 |

| | | | | | | | | | |
|---|---|---|---|---|---|---|---|---|---|
| Dennis, A[27] | a | NA | a | a | NA | c | a | d | 3 |
| Eiros, R[28] | c | NA | b | a | NA | b | a | b | 5 |
| Geortz, YNJ[29] | b | NA | c | a | NA | c | a | d | 3 |
| Halpin, SJ[30] | a | NA | b | a | NA | c | a | c | 4 |
| Huang, C[31] | a | NA | a | a | NA | a | a | c | 5 |
| Klein, H[32] | a | NA | b | a | NA | c | a | b | 5 |
| Lavoto, A[33] | c | NA | a | a | NA | c | a | d | 3 |
| Mandal, S[34] | a | NA | b | a | NA | c | a | b | 5 |
| Moradian, ST[35] | a | NA | b | a | NA | a | a | a | 6 |
| Neto, DB[36] | b | NA | c | b | NA | c | a | b | 4 |
| Petersen, M[37] | a | NA | a | b | NA | c | a | a | 5 |
| Pilotto, A[38] | a | NA | b | a | NA | a | a | a | 6 |
| Poncet-Megemont, L[39] | a | NA | b | a | NA | c | a | b | 5 |
| Rahmani, H[40] | b | NA | a | a | NA | a | a | b | 6 |

| | | | | | | | | | |
|---|---|---|---|---|---|---|---|---|---|
| Savarraj, JPJ[41] | a | NA | b | a | NA | c | a | b | 5 |
| Salmon-Ceron, D[42] | a | NA | a | a | NA | b | a | b | 6 |
| Stavem, K[43] | a | NA | a | b | NA | c | a | a | 5 |
| Sudre, C[44] | a | NA | b | b | NA | c | a | a | 5 |
| Tenforde, MW[45] | b | NA | b | a | NA | c | a | b | 5 |
| Tomasoni, D[46] | a | NA | b | b | NA | a | a | a | 6 |
| Townsend, L[47] | a | NA | b | a | NA | a | a | a | 6 |
| Wang, X[48] | a | NA | a | b | NA | a | a | b | 6 |
| Weerahandi, H[49] | b | NA | b | a | NA | c | a | b | 5 |
| Wu, C[50] | a | NA | b | a | NA | d | a | b | 6 |
| Xiong, Q[51] | a | NA | b | a | NA | c | a | b | 5 |
| Yan, N[52] | a | NA | b | a | NA | c | b | b | 5 |
| Zhao, Y[53] | a | NA | a | a | NA | b | a | a | 6 |

NA: not applicable.

### eFigure 1a. Forest Plot on Abdominal Pain

| Study | Events | Total | Proportion | 95%-CI | Weight |
|---|---|---|---|---|---|
| Arnold, D | 2 | 1[illegible]0 | 0.02 | [0.00; 0.06] | 6.5% |
| Boscolo-Rizzo, P | 2 | [illegible]37 | 0.01 | [0.00; 0.06] | 4.6% |
| Davis, HE | 1505 | 3762 | 0.40 | [0.38; 0.42] | 13.6% |
| Dennis, A | 108 | 201 | 0.54 | [0.47; 0.61] | 13.1% |
| Eiros, R | 6 | 13[illegible] | 0.04 | [0.02; 0.09] | 9.9% |
| Klein, H | 1 | [illegible]12 | 0.01 | [0.00; 0.05] | 4.3% |
| Salmon-Ceron, D | 3 | 7[illegible] | 0.04 | [0.01; 0.12] | 7.8% |
| Stavem, K | 14 | 4[illegible] | 0.03 | [0.02; 0.05] | 11.8% |
| Sudre, C | 1056 | 4182 | 0.25 | [0.24; 0.27] | 13.6% |
| Tenforde, MW | 41 | 270 | 0.15 | [0.11; 0.20] | 12.8% |
| **Random effects model** | | **9484** | **0.11** | **[0.07; 0.16]** | **100.0%** |

Heterogeneity: $I^2$ = 98%, □$^2$ = 0.4681, $p$ < 0.01

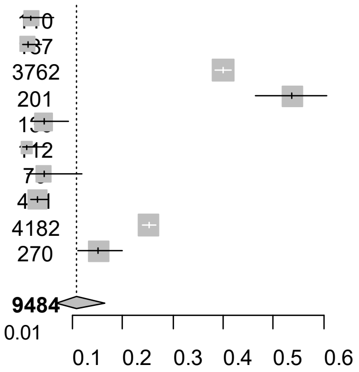


Point sizes are proportional to an inverse of the precision of the estimates and bar correspond to 95% confidence intervals.

**eFigure 1b. Bubble Plots (follow-up period) on Abdominal Pain**

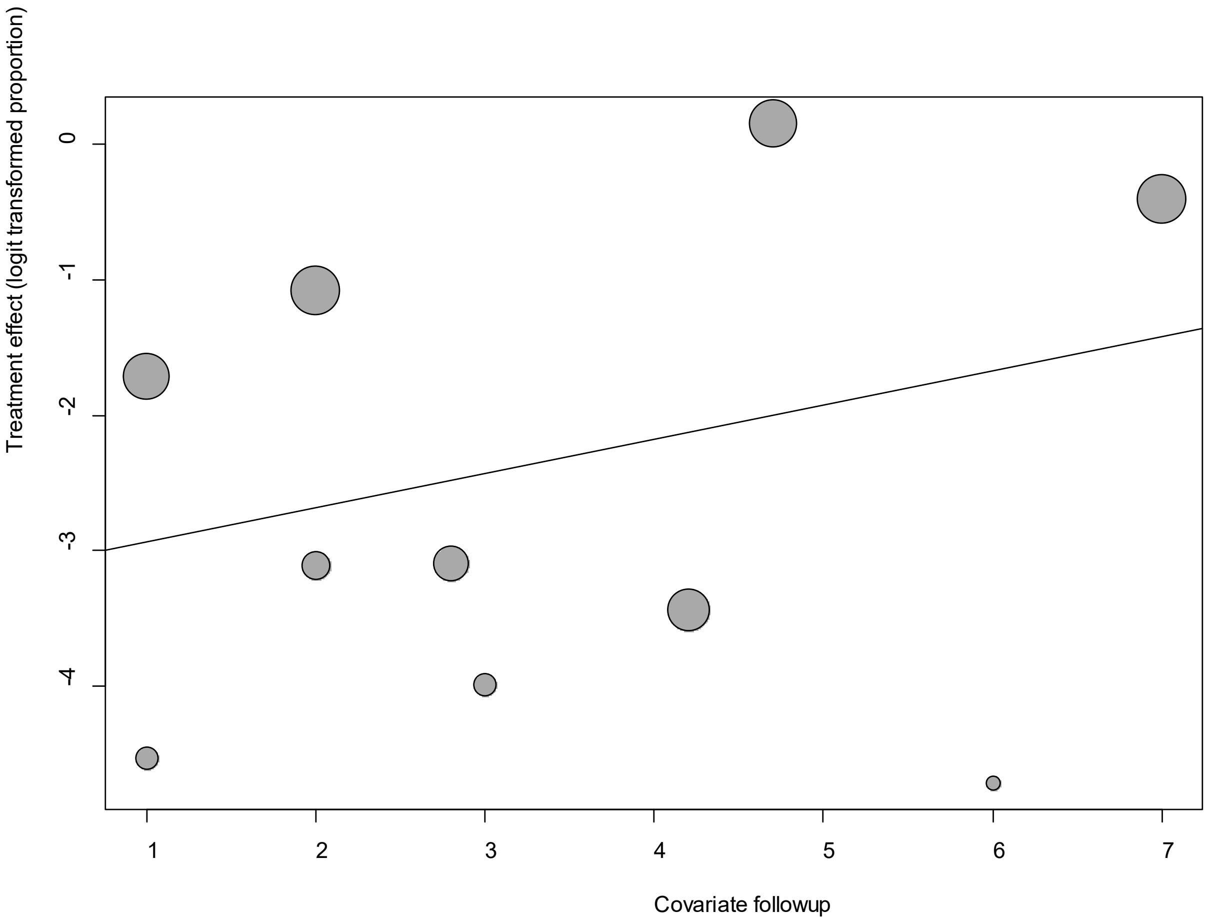


Point sizes are proportional to an inverse of the precision of the estimates.

The regression coefficient was 0.25 (95% confidence interval:-0.12-0.62).(*P*=0.18).

**eFigure 1c. Bubble Plots (age) on Abdominal Pain**

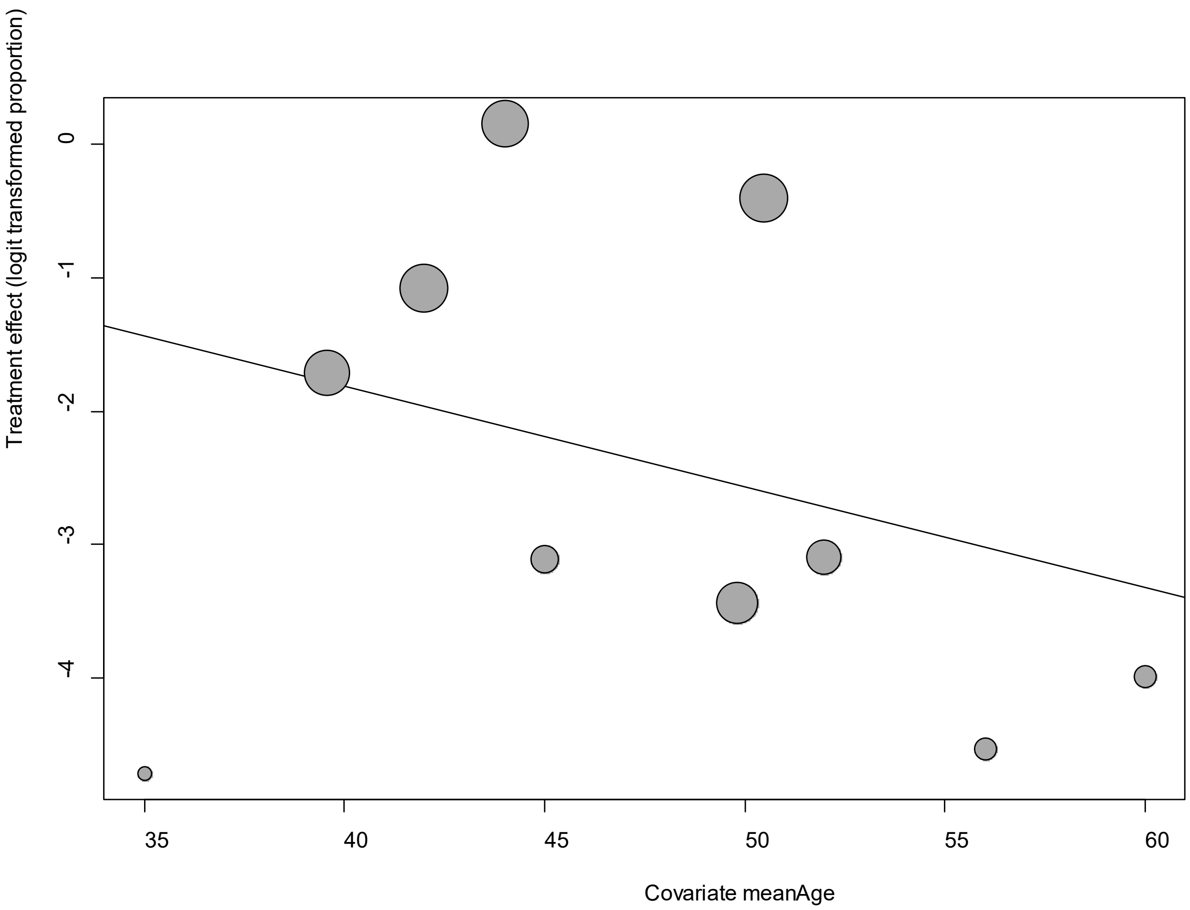


Point sizes are proportional to an inverse of the precision of the estimates.

The regression coefficient was -0.08 (95% confidence interval:-0.19-0.04).(*P*=0.20).

**eFigure 1d. Bubble Plots (sex) on Abdominal Pain**

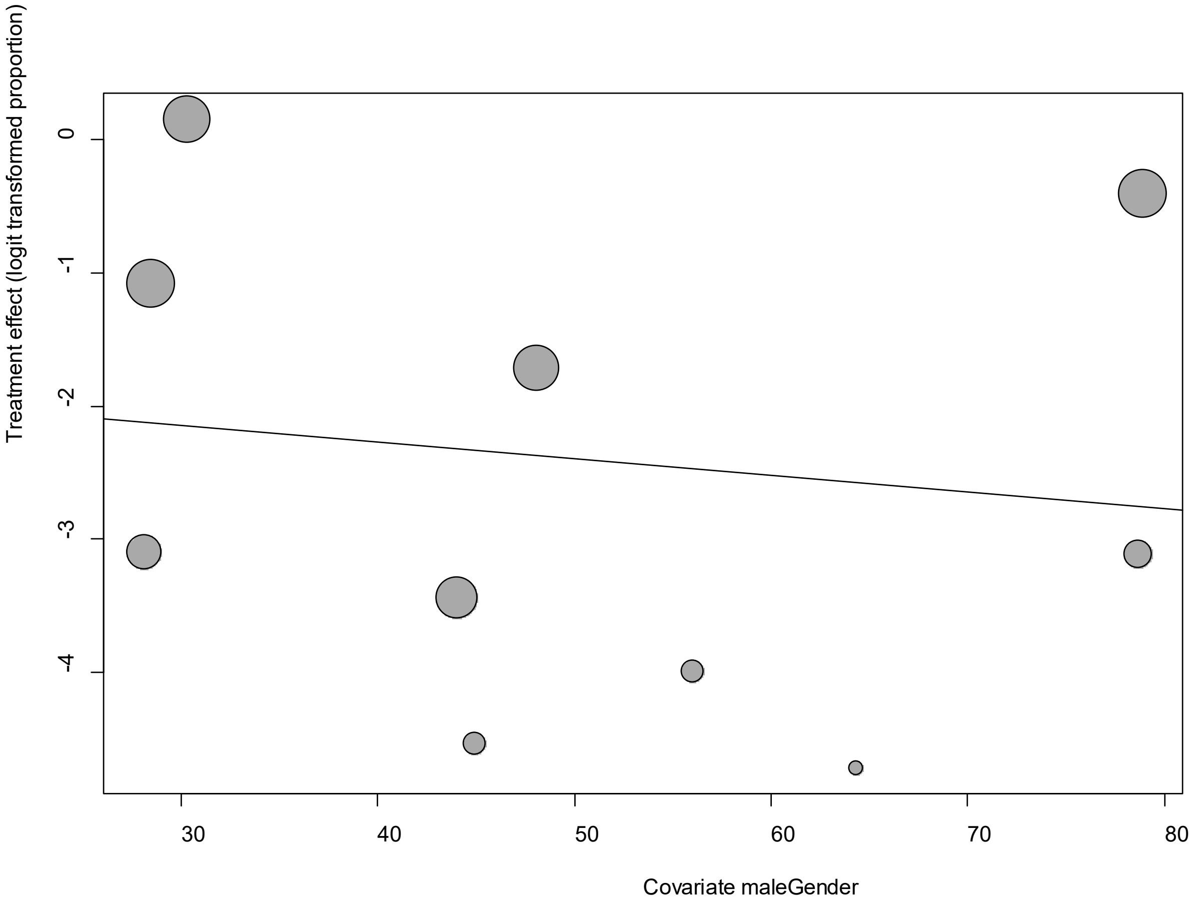


Point sizes are proportional to an inverse of the precision of the estimates.

The regression coefficient was -0.01 (95% confidence interval:--0.06-0.03).(*P*=0.59).

**eFigure 1e. Funnel Plot of studies reporting Abdominal Pain**

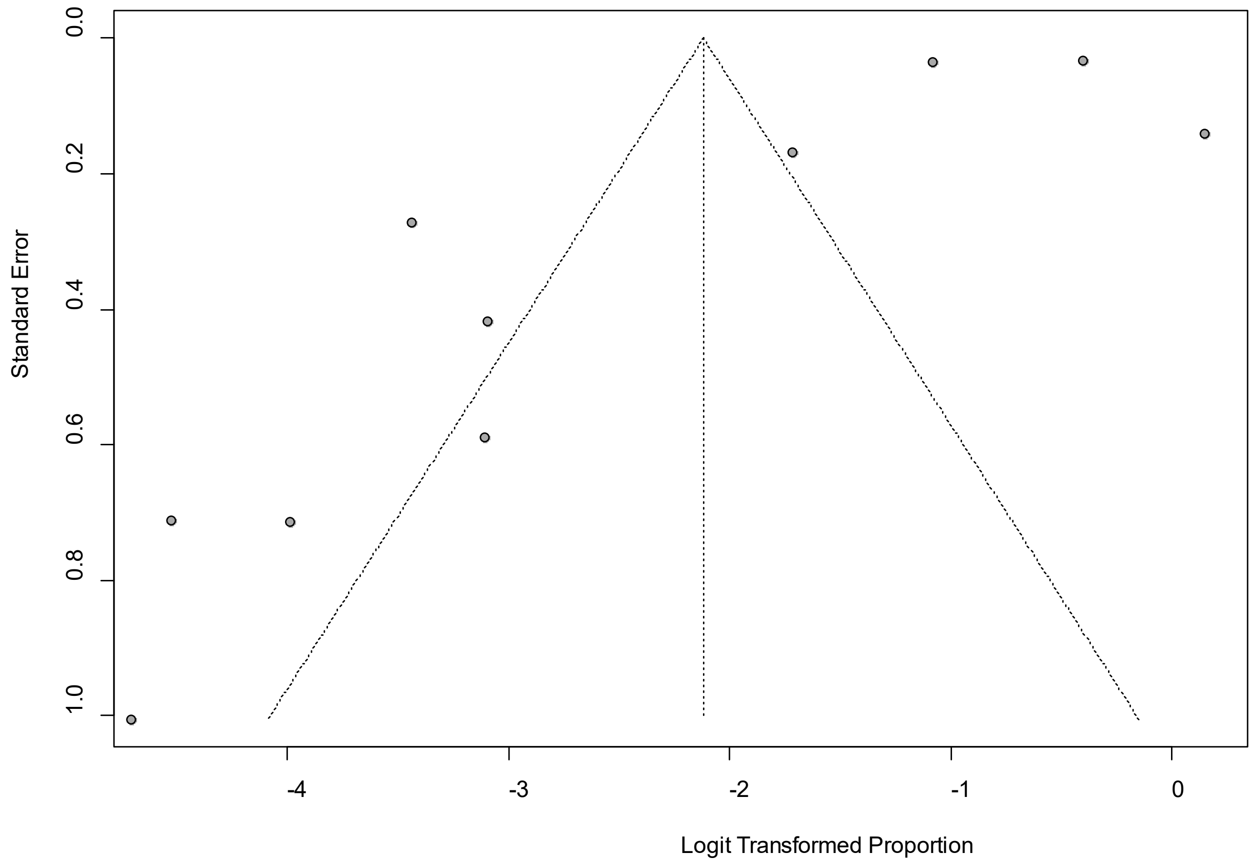


Egger's *P* was 0.09, indicating the presence of publication bias.

**eFigure 2a. Forest Plot on Arthralgia**

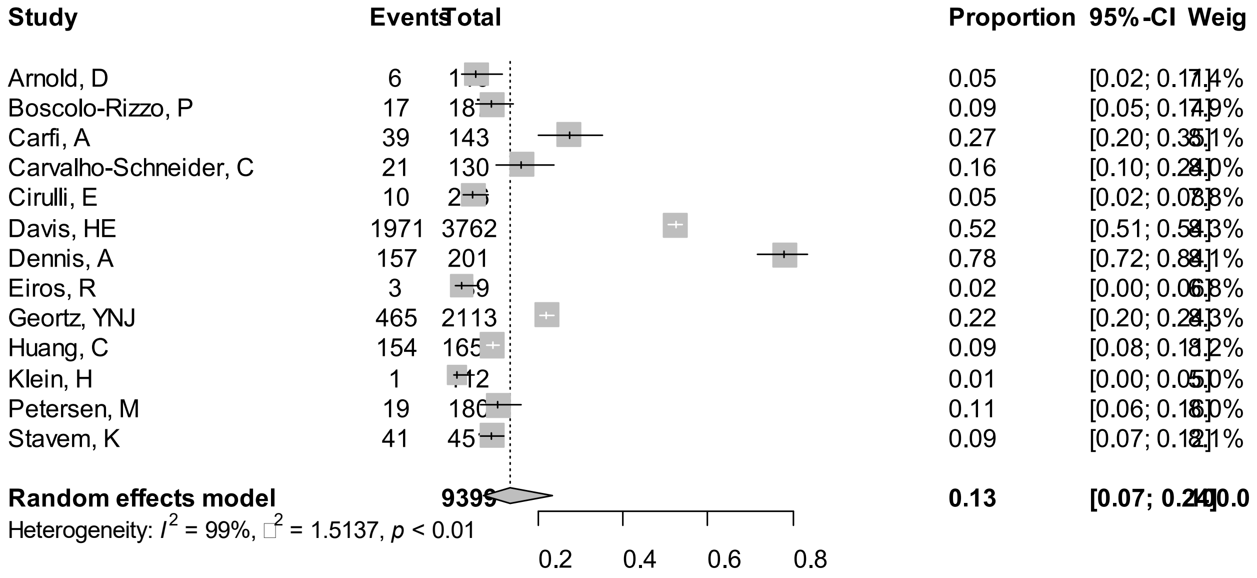


Point sizes are proportional to an inverse of the precision of the estimates and bar correspond to 95% confidence intervals.

**eFigure 2b. Bubble Plots (follow-up period) on Arthralgia**

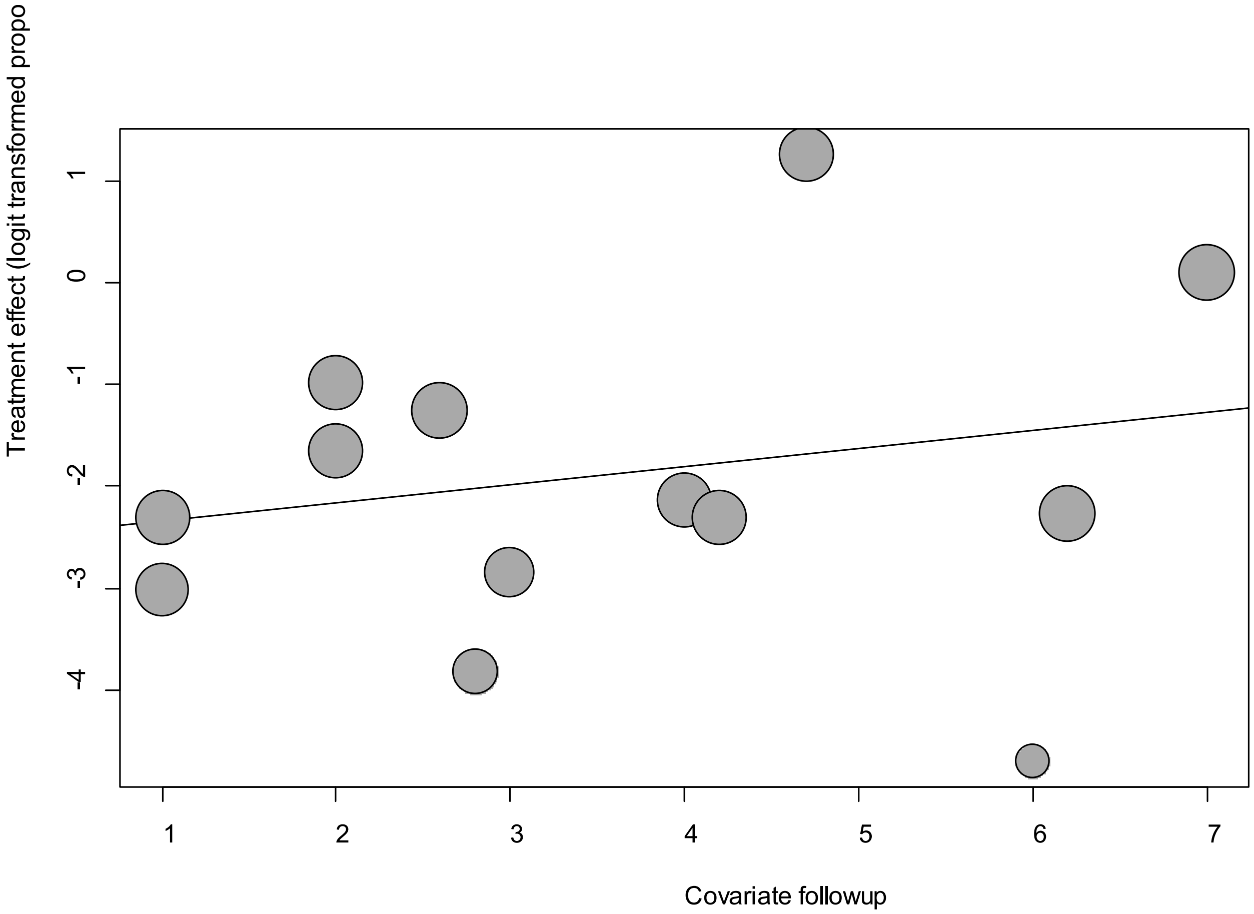


Point sizes are proportional to an inverse of the precision of the estimates.

The regression coefficient was 0.18 (95% confidence interval:-0.22-0.57).(*P*=0.37).

**eFigure 2c. Bubble Plots (age) on Arthralgia**

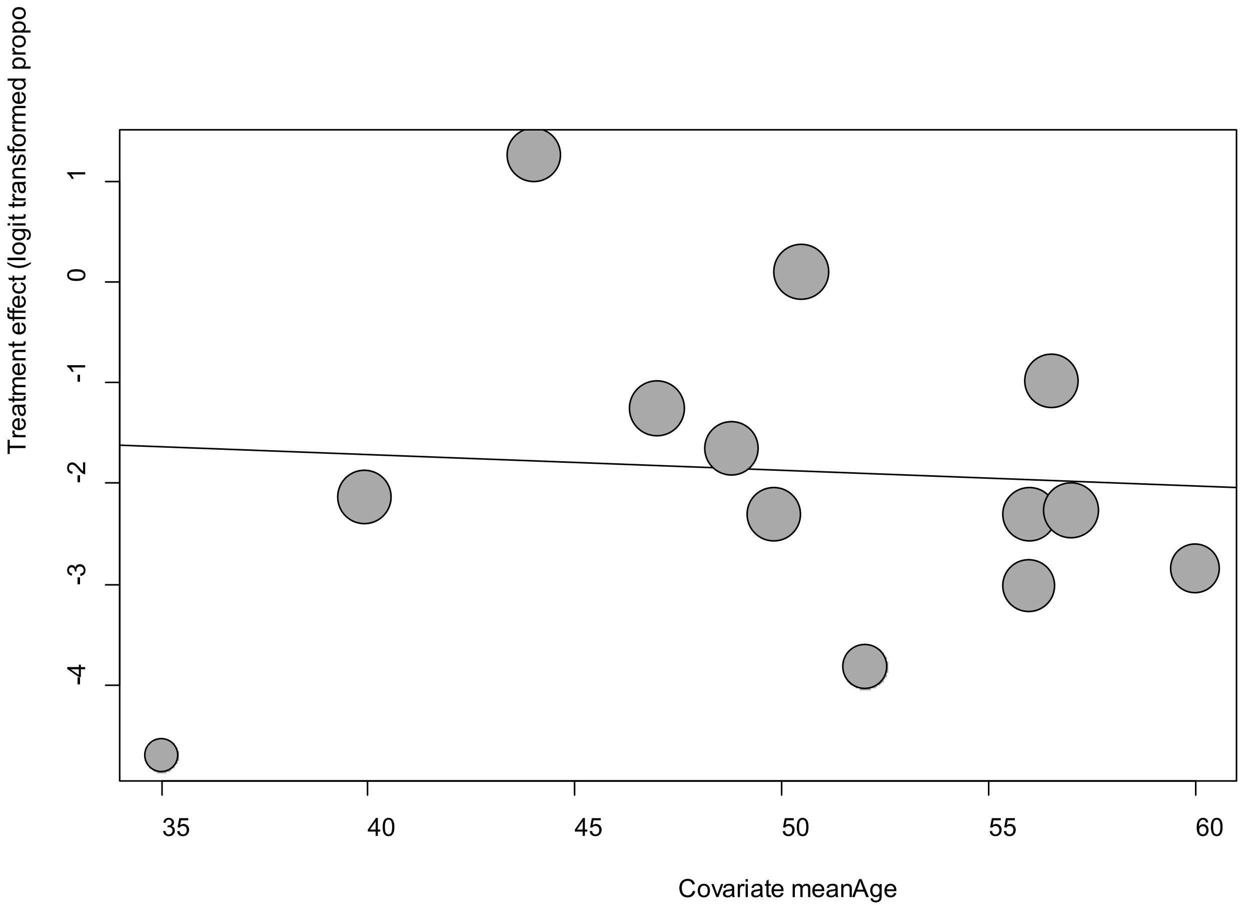


Point sizes are proportional to an inverse of the precision of the estimates.

The regression coefficient was -0.02 (95% confidence interval:-0.13-0.09).(*P*=0.78).

**eFigure 2d. Bubble Plots (sex) on Arthralgia**

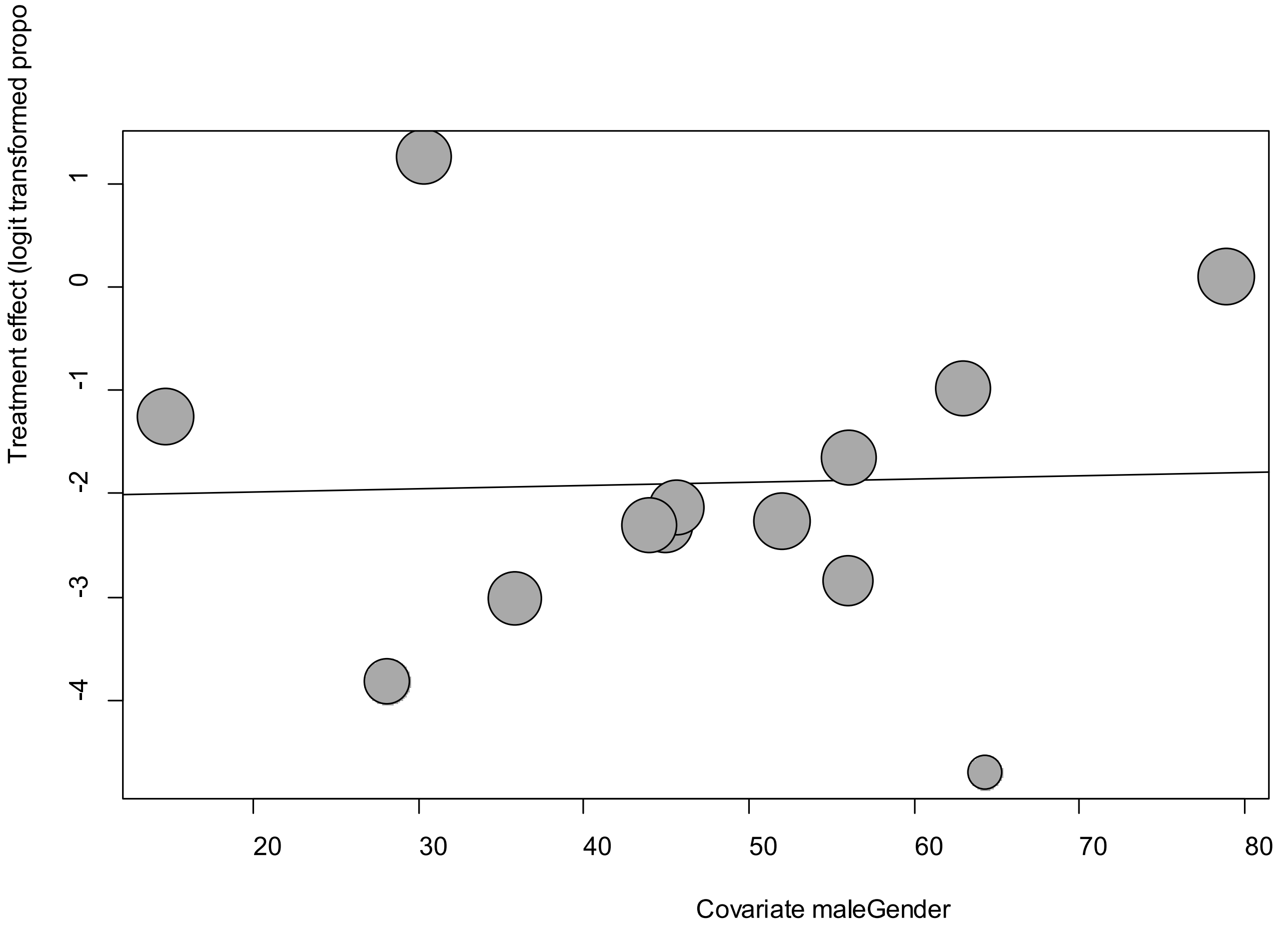


Point sizes are proportional to an inverse of the precision of the estimates.

The regression coefficient was 0.003 (95% confidence interval:-0.05-0.05).(*P*=0.90).

**eFigure 2e. Funnel Plot of studies reporting on Arthralgia**

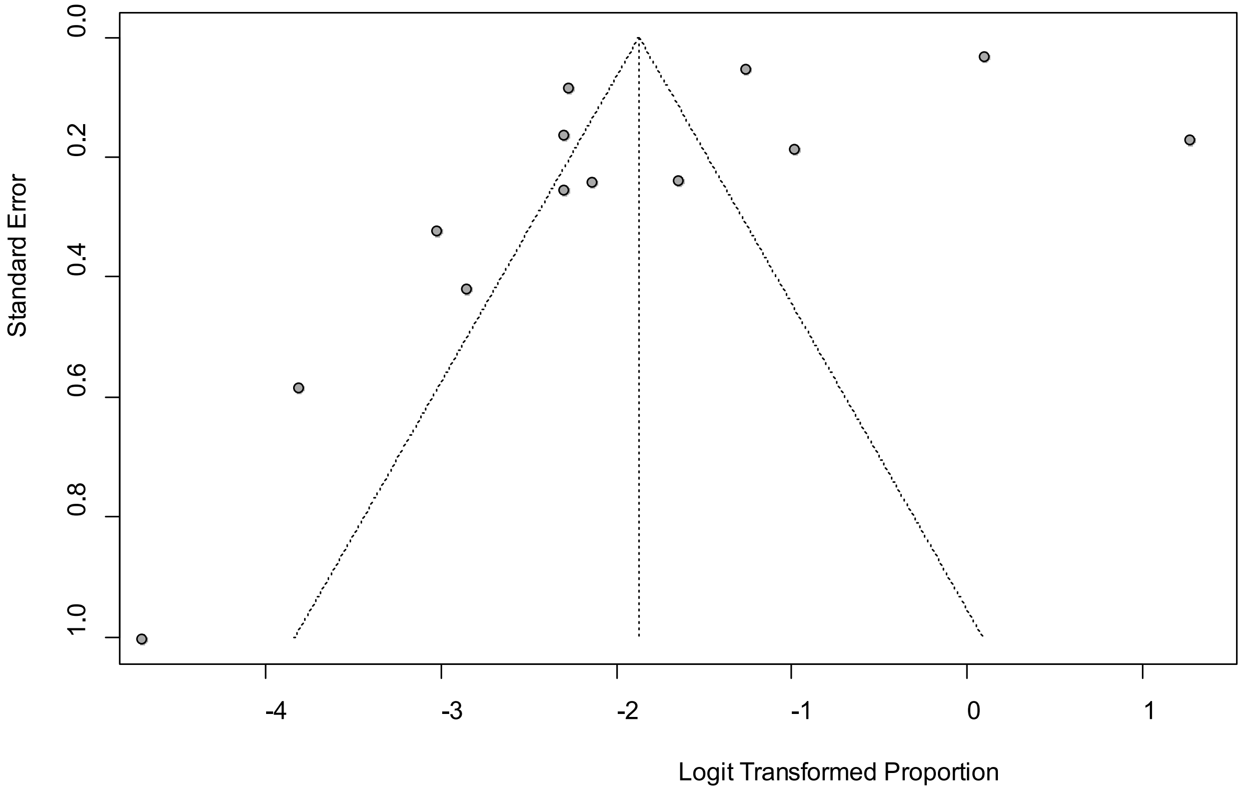


Egger's *P* was 0.9, indicating the absence of publication bias.

**eFigure 3a. Forest Plot on Chest Pain**

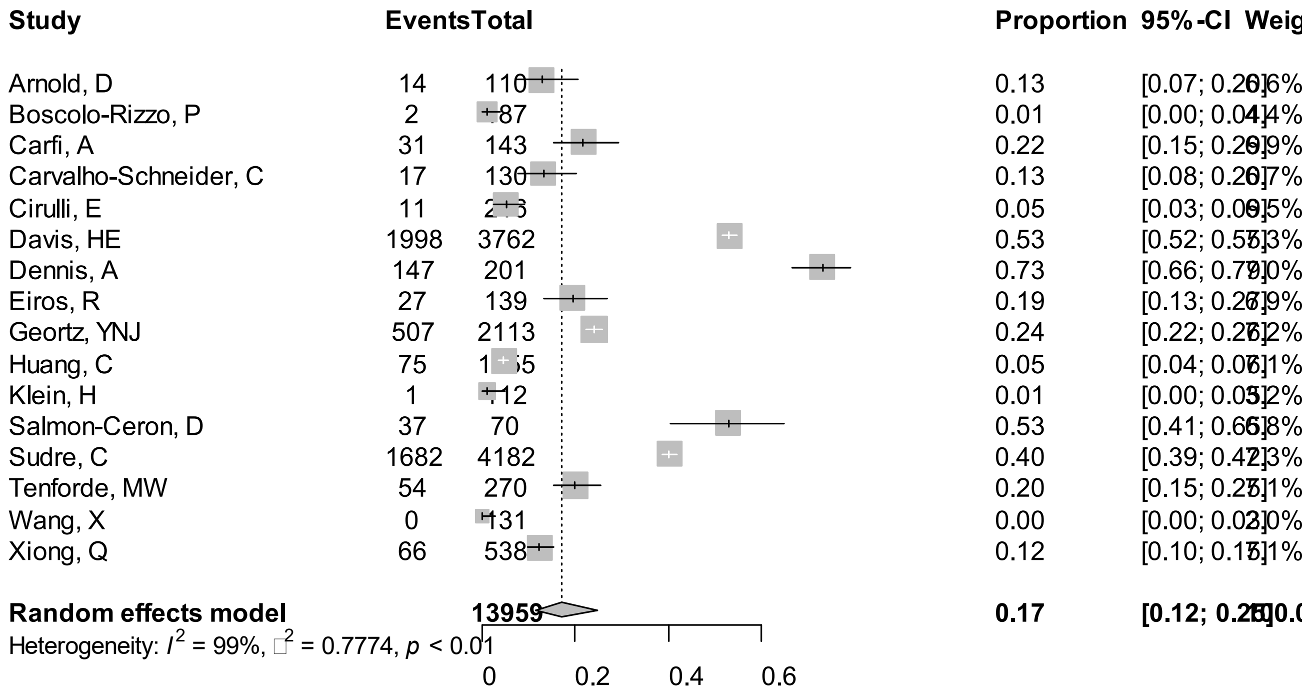


Point sizes are proportional to an inverse of the precision of the estimates and bar correspond to 95% confidence intervals.

**eFigure 3b. Bubble Plots (follow-up period) on Chest Pain**

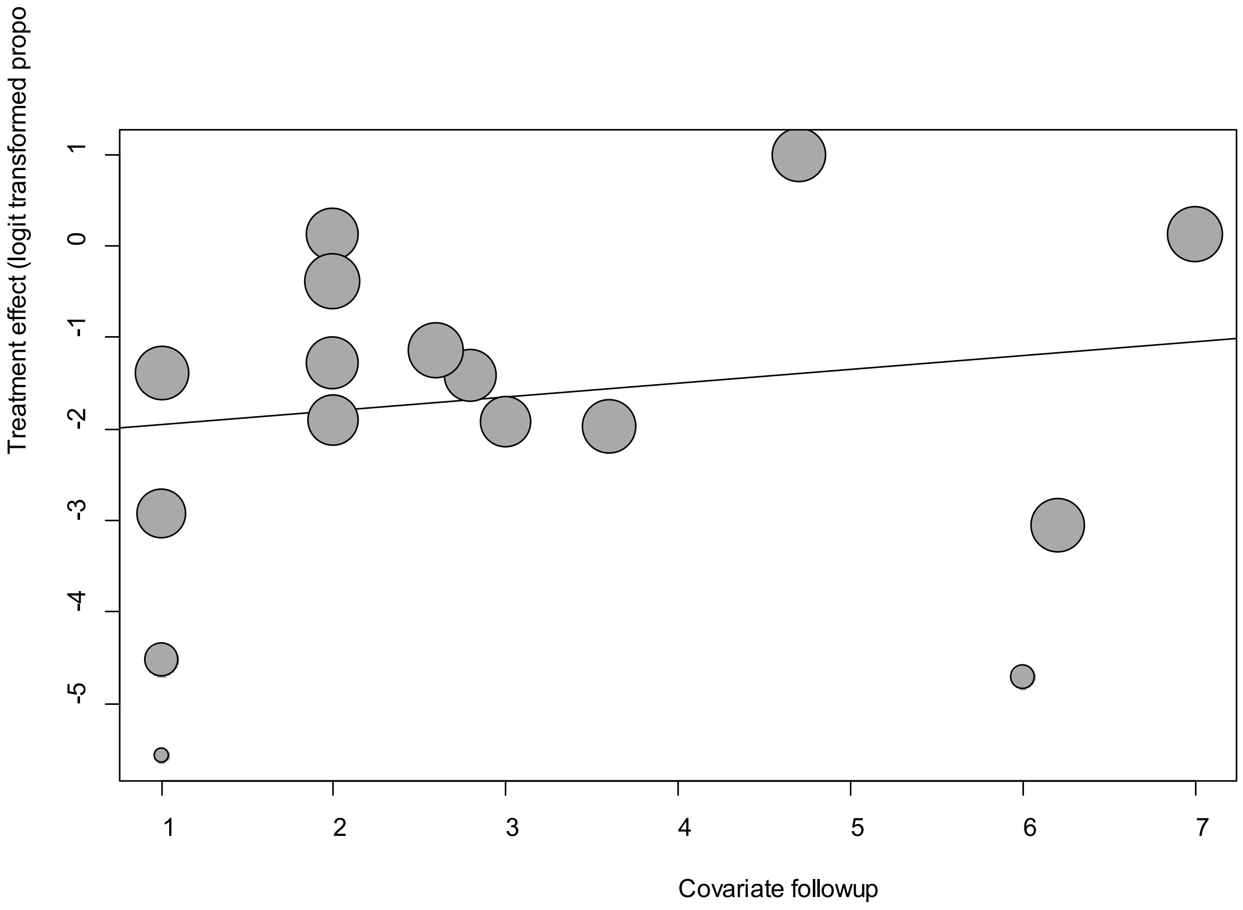


Point sizes are proportional to an inverse of the precision of the estimates.

The regression coefficient was 0.15 (95% confidence interval:-0.15-0.45).(*P*=0.34).

**eFigure 3c. Bubble Plots (age) on Chest Pain**

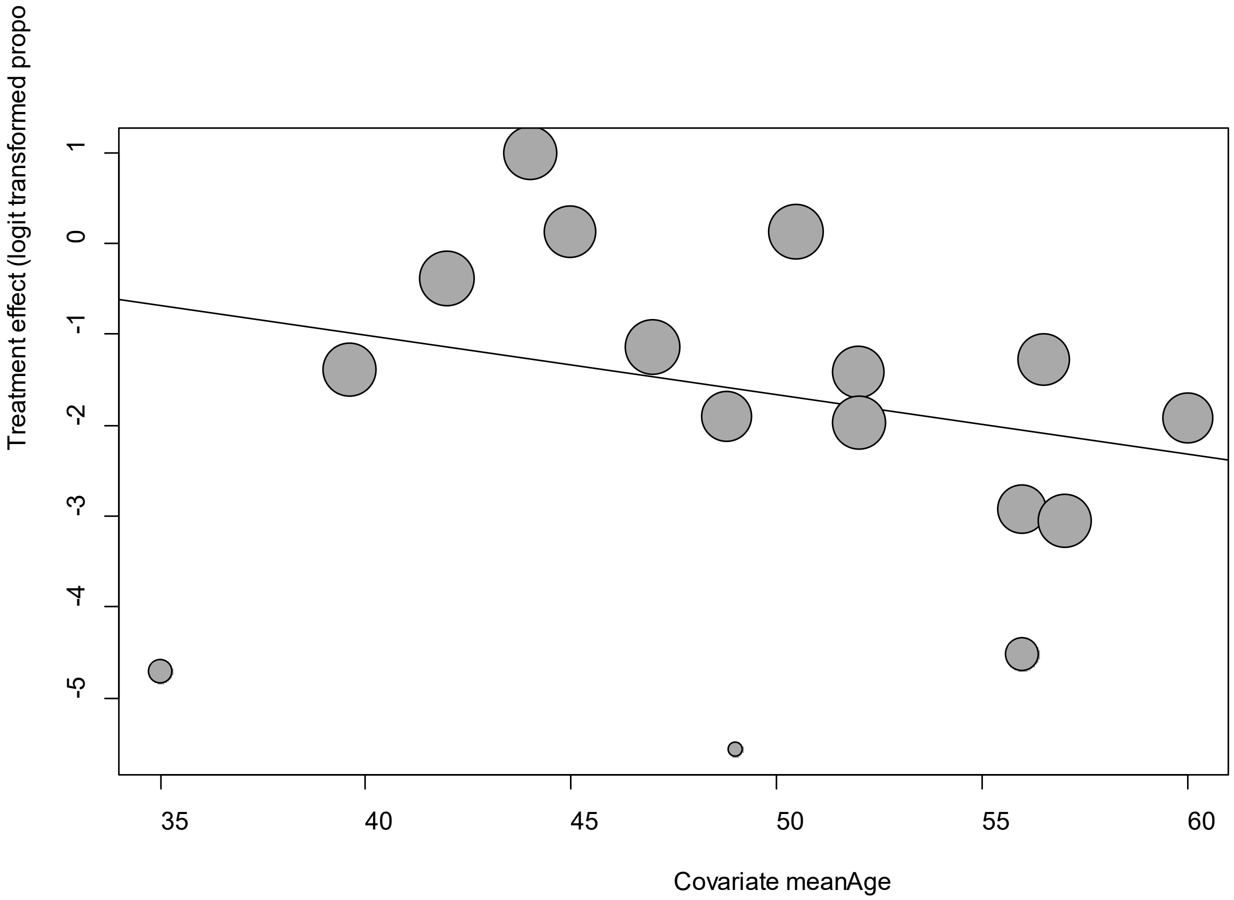


Point sizes are proportional to an inverse of the precision of the estimates.

The regression coefficient was -0.07 (95% confidence interval:-0.15-0.02).(*P*=0.14).

**eFigure 3d. Bubble Plots (sex) on Chest Pain**

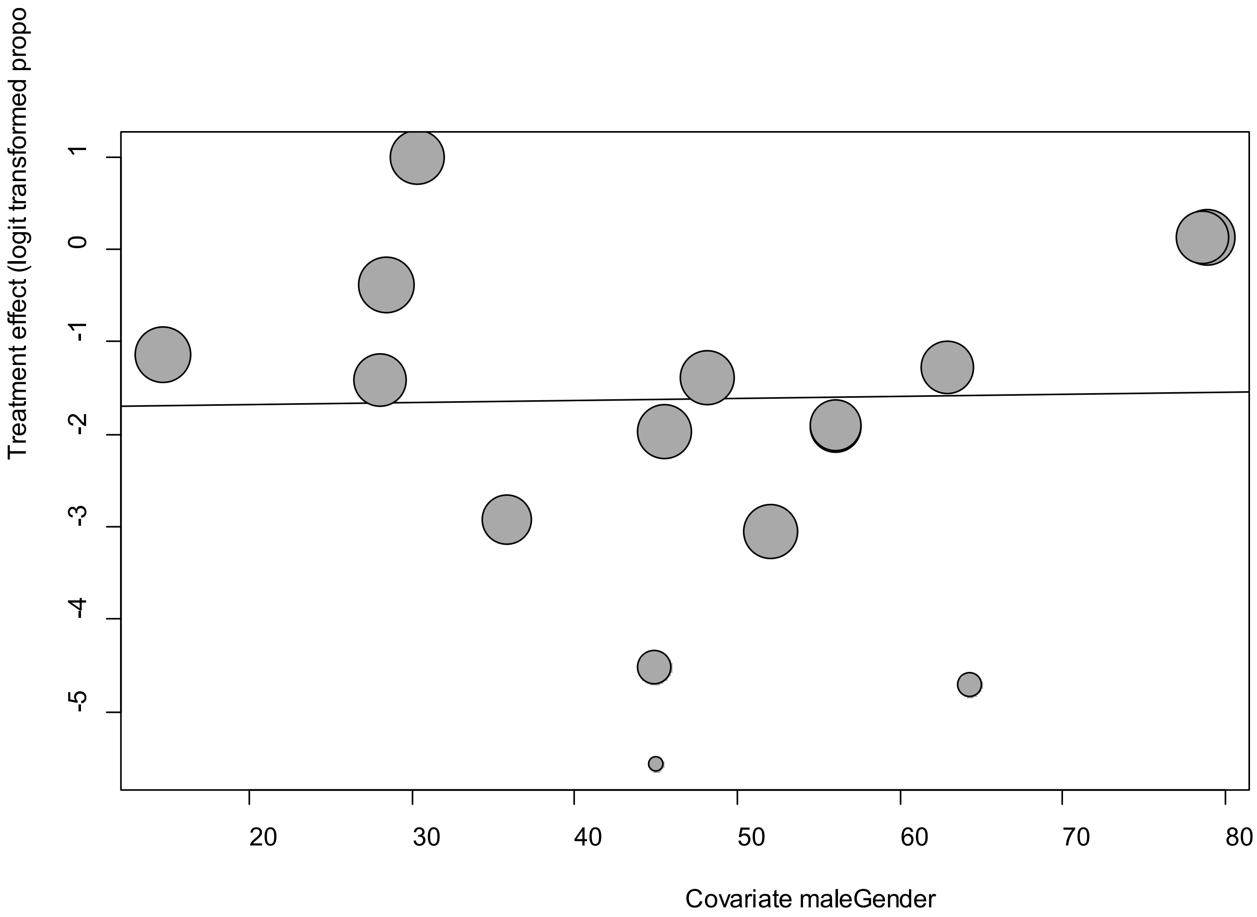


Point sizes are proportional to an inverse of the precision of the estimates.

The regression coefficient was -0.002 (95% confidence interval:-0.027-0.03).(*P*=0.88).

**eFigure 3e. Funnel Plot of studies reporting on Chest Pain**

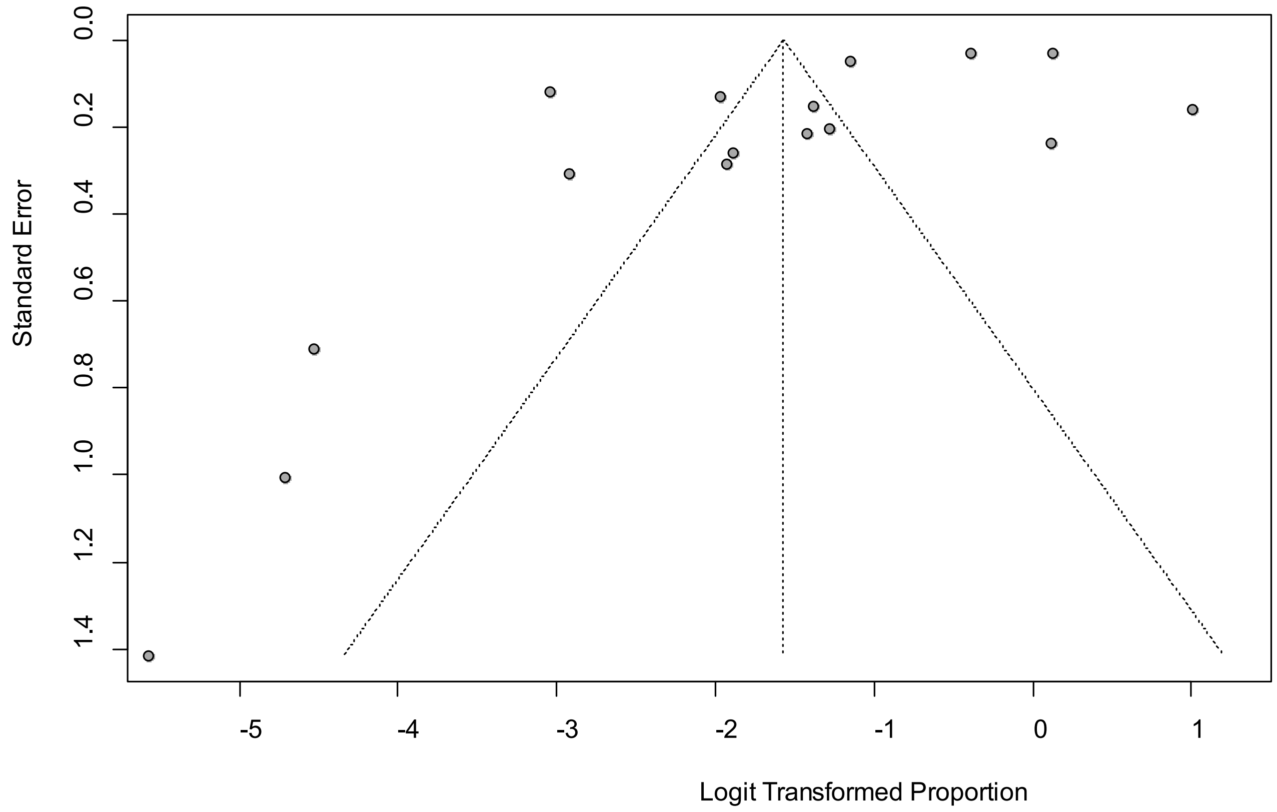


Egger's *P* was 0.03, indicating the presence of publication bias.

**eFigure 4a. Forest Plot on Ear Pain**

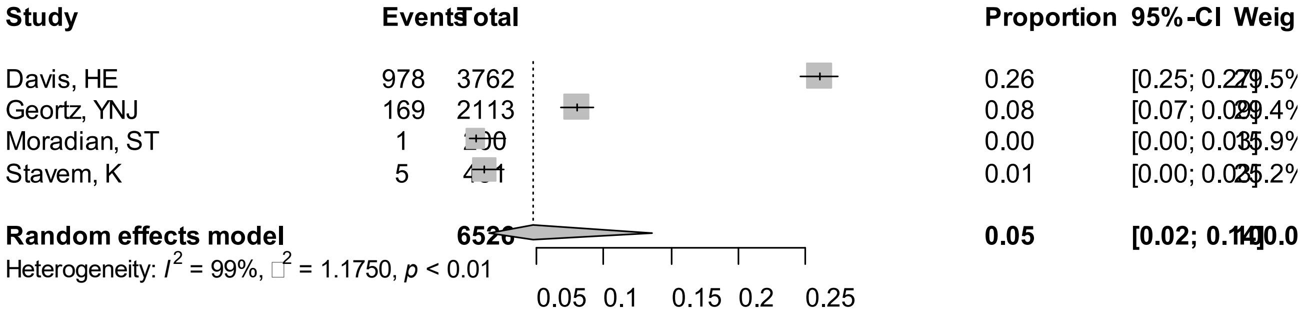


Point sizes are proportional to an inverse of the precision of the estimates and bar correspond to 95% confidence intervals.

**eFigure 4b. Bubble Plots (follow-up period) on Ear Pain**

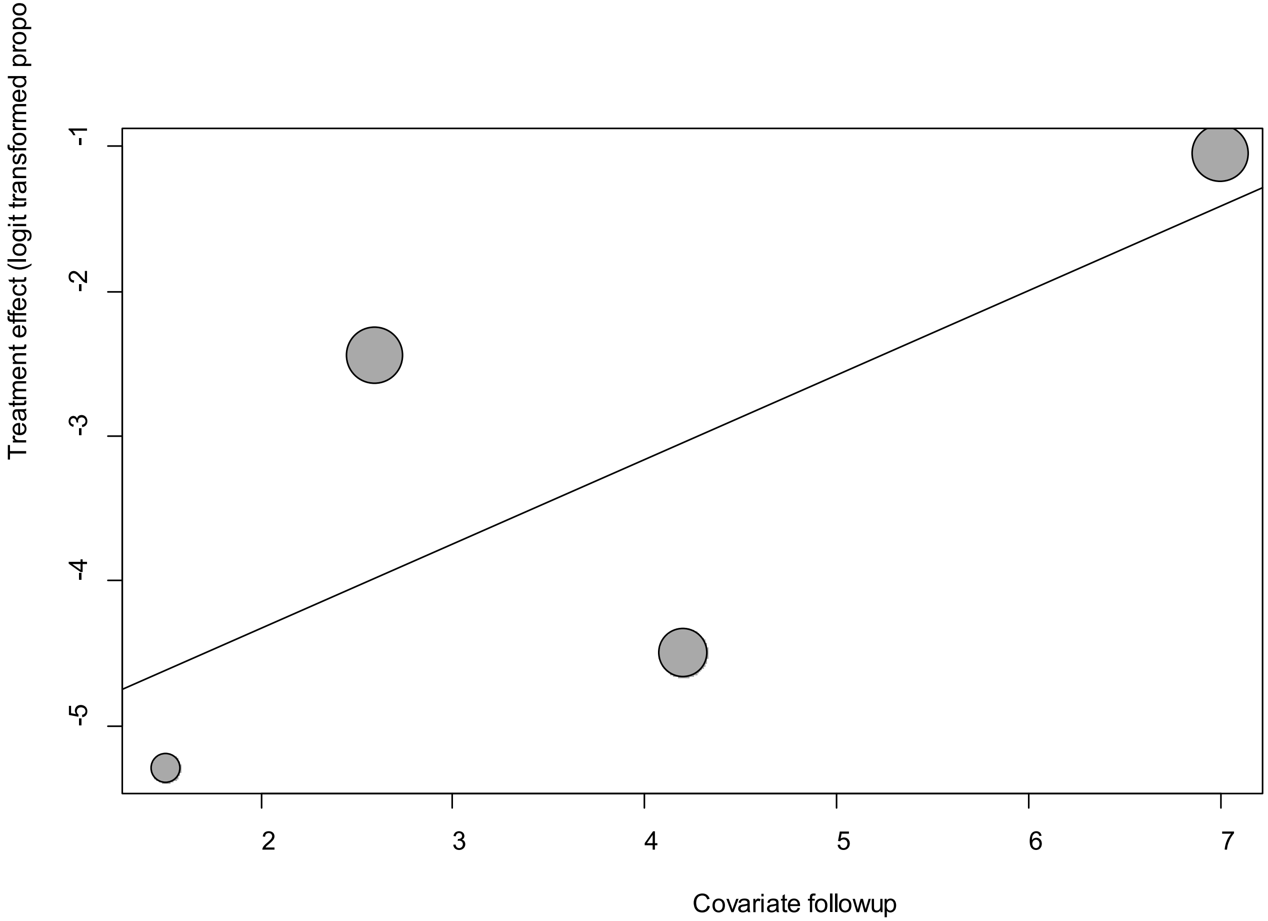


Point sizes are proportional to an inverse of the precision of the estimates.

The regression coefficient was 0.58 (95% confidence interval:-0.35-1.51).(*P*=0.22).

**eFigure 4c. Bubble Plots (age) on Ear Pain**

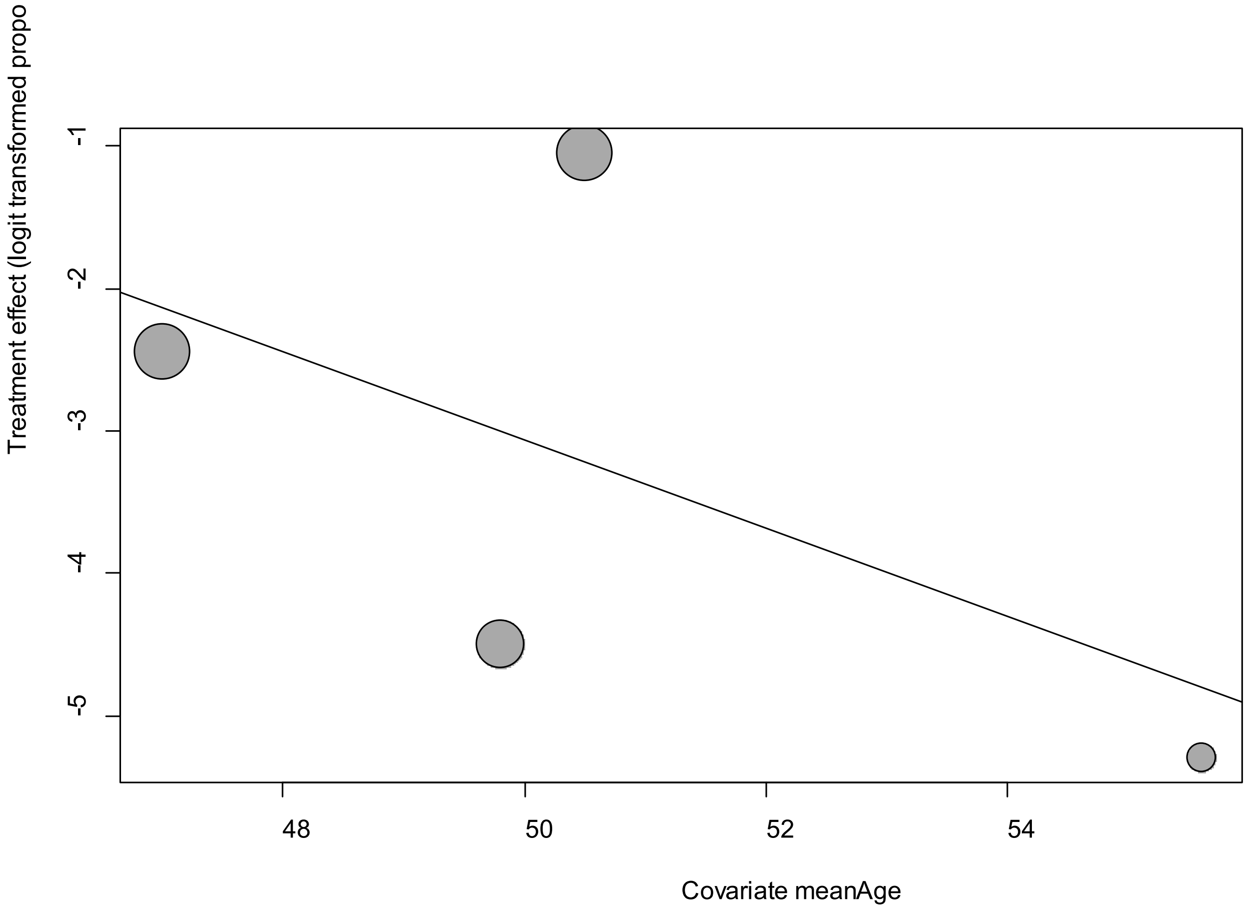


Point sizes are proportional to an inverse of the precision of the estimates.

The regression coefficient was -0.31 (95% confidence interval:-1.06-0.44).(*P*=0.42).

**eFigure 4d. Bubble Plots (sex) on Ear Pain**

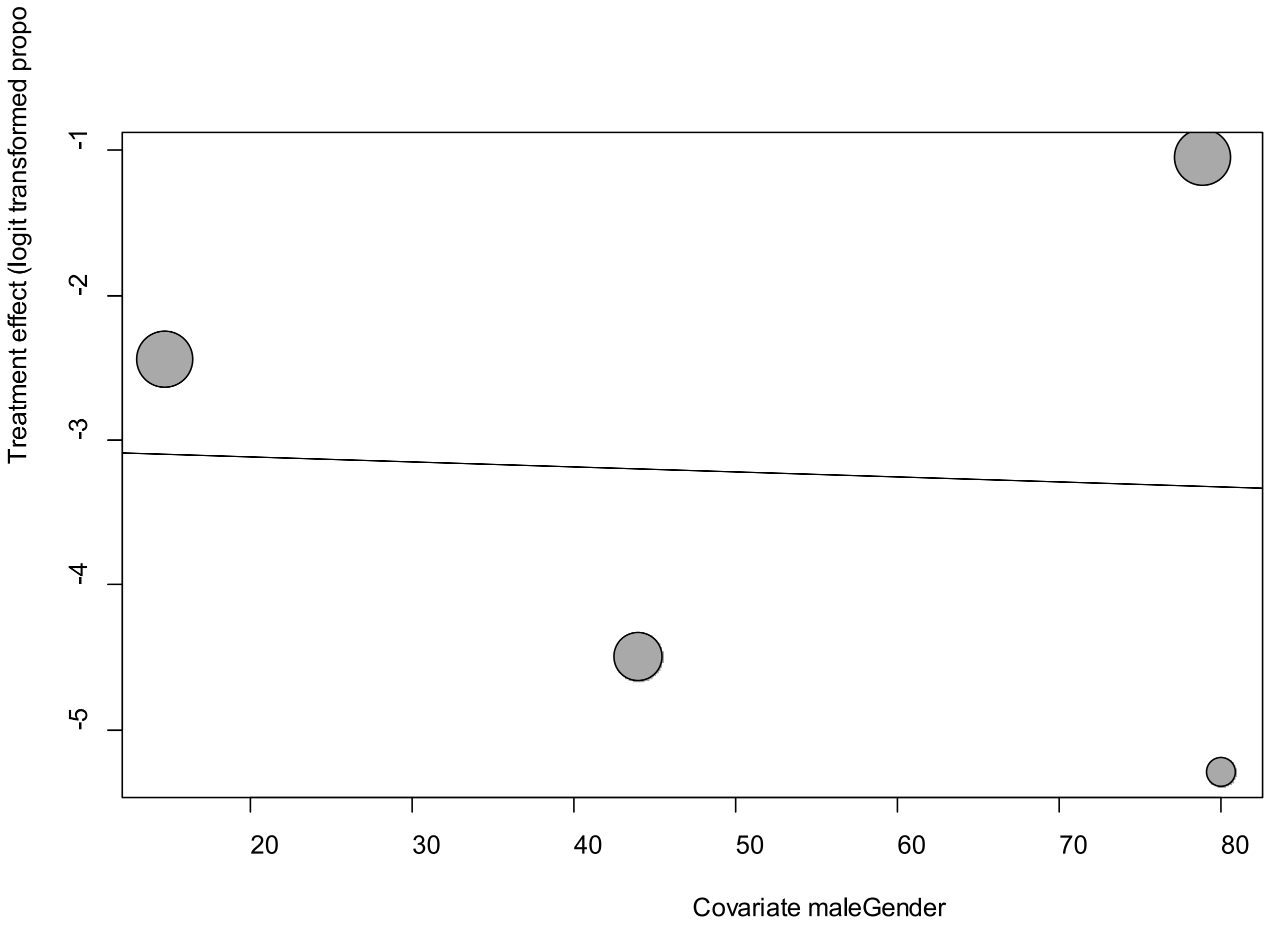


Point sizes are proportional to an inverse of the precision of the estimates.

The regression coefficient was -0.003 (95% confidence interval:-0.090-0.08).(*P*=0.94).

**eFigure 4e. Funnel Plot of studies reporting on Ear Pain**

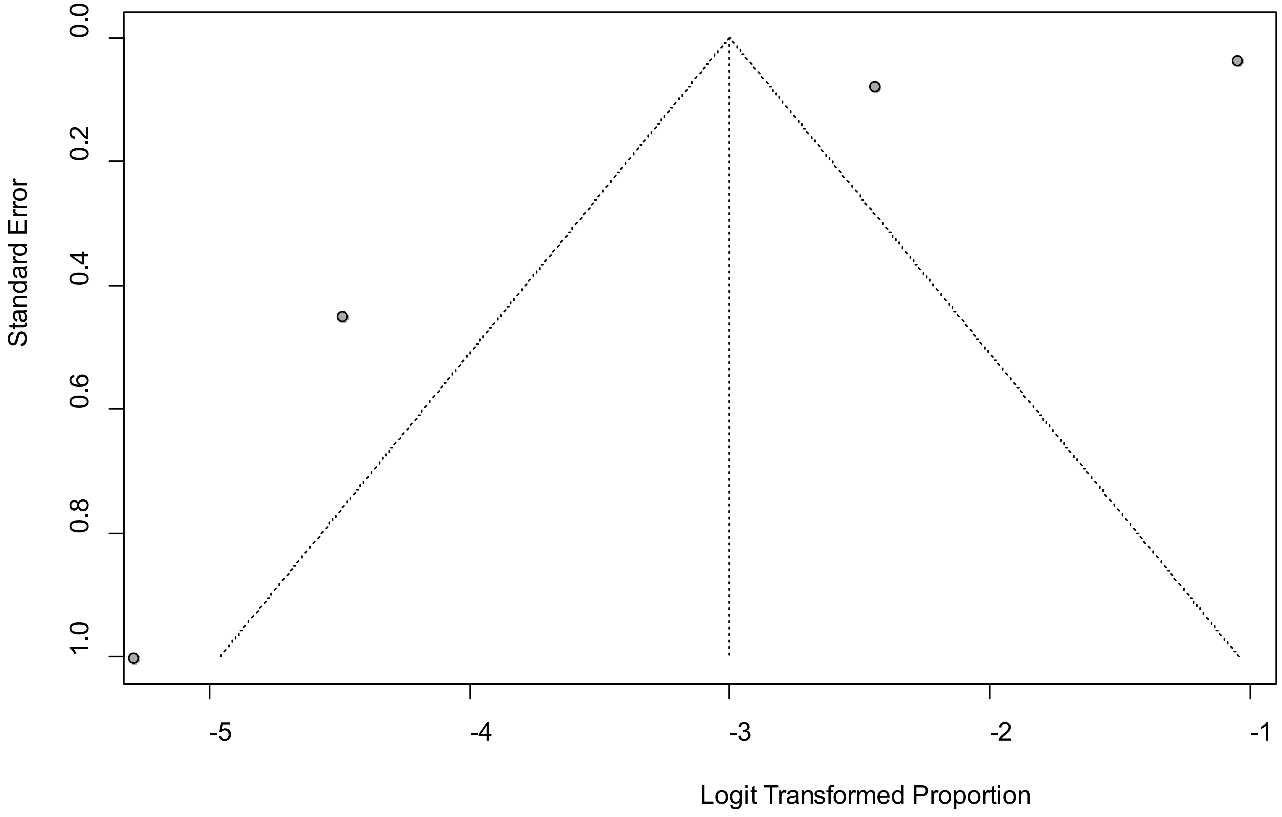


Egger's *P* was 0.28, indicating the absence of publication bias.

**eFigure 5a. Forest Plot on Headache**

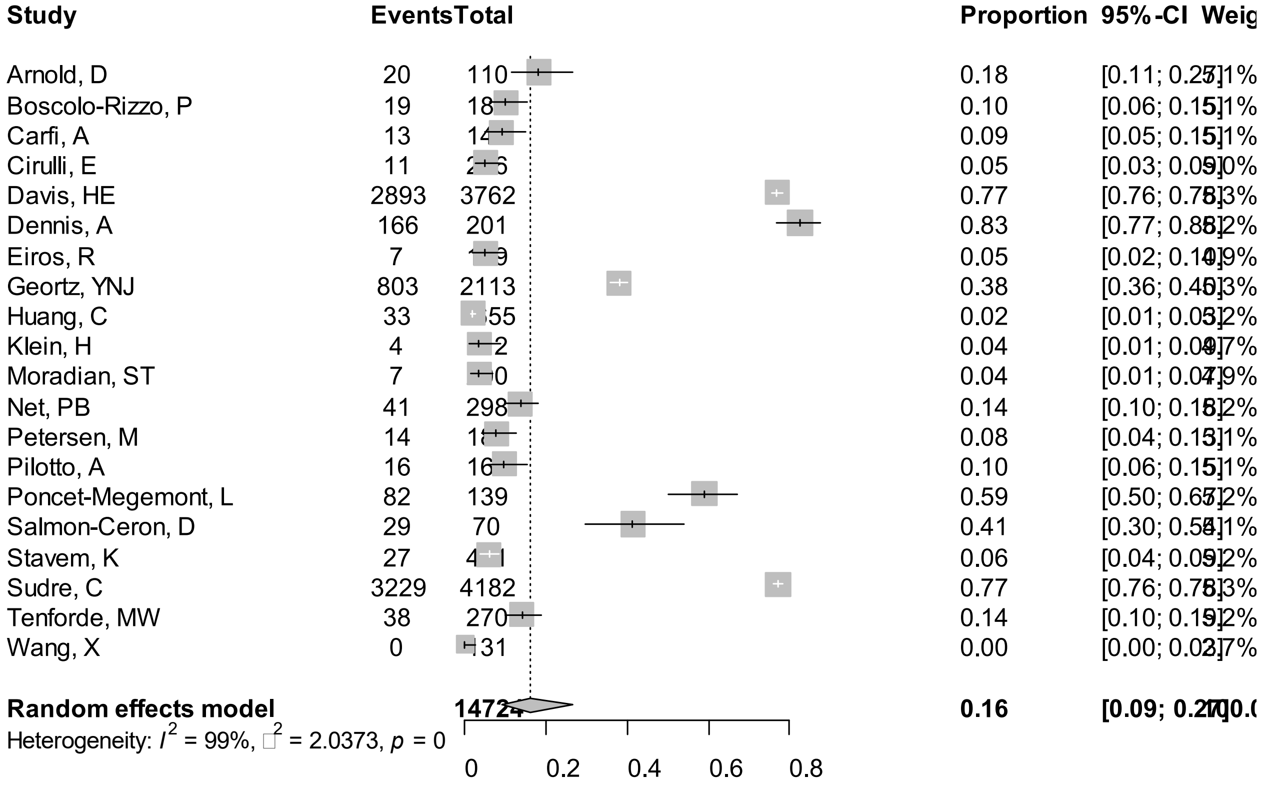


Point sizes are proportional to an inverse of the precision of the estimates and bar correspond to 95% confidence intervals.

**eFigure 5b. Bubble Plots (follow-up period) on Headache**

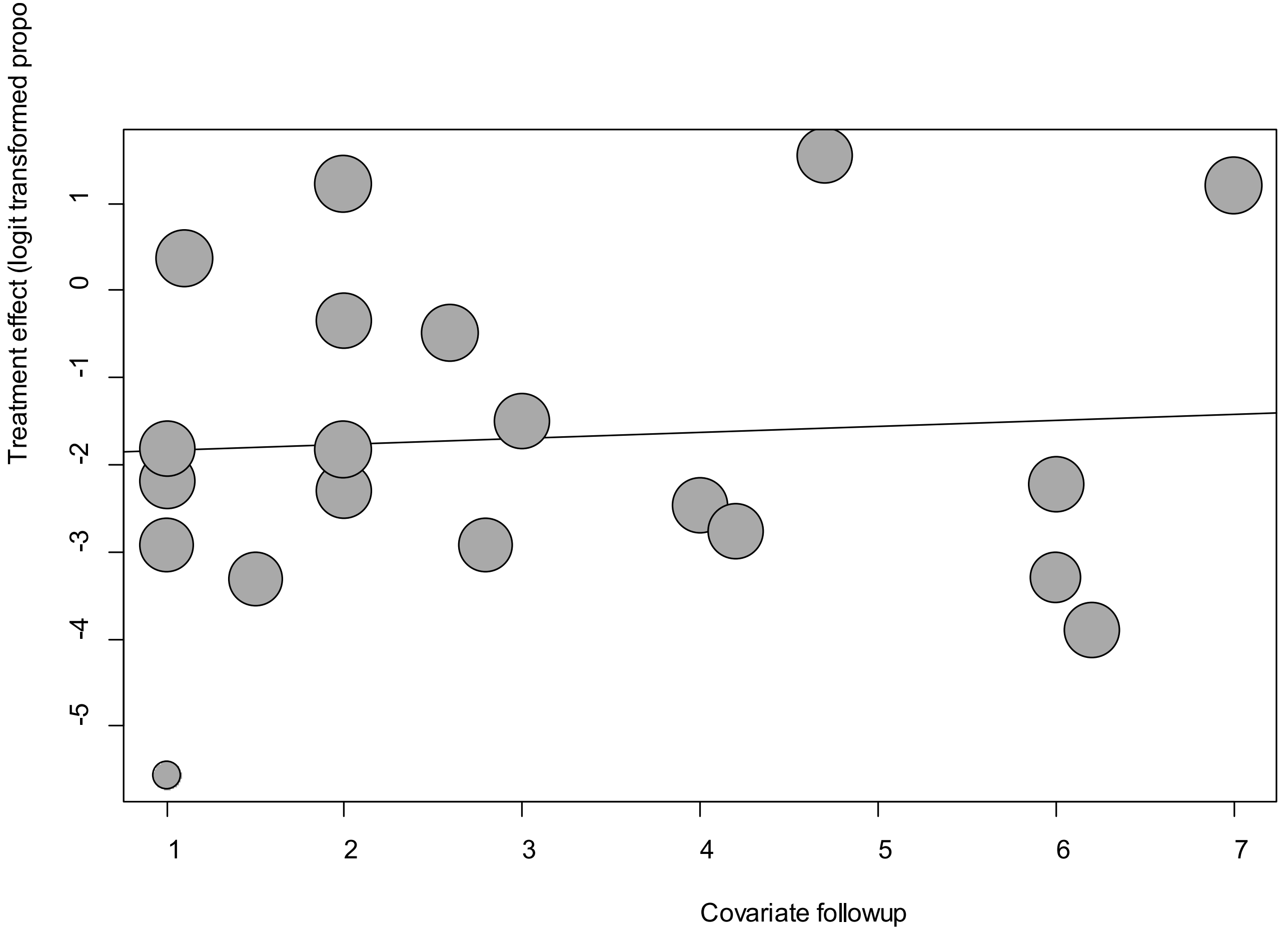


Point sizes are proportional to an inverse of the precision of the estimates.

The regression coefficient was 0.07 (95% confidence interval:-0.33-0.47).(*P*=0.74).

**eFigure 5c. Bubble Plots (age) on Headache**

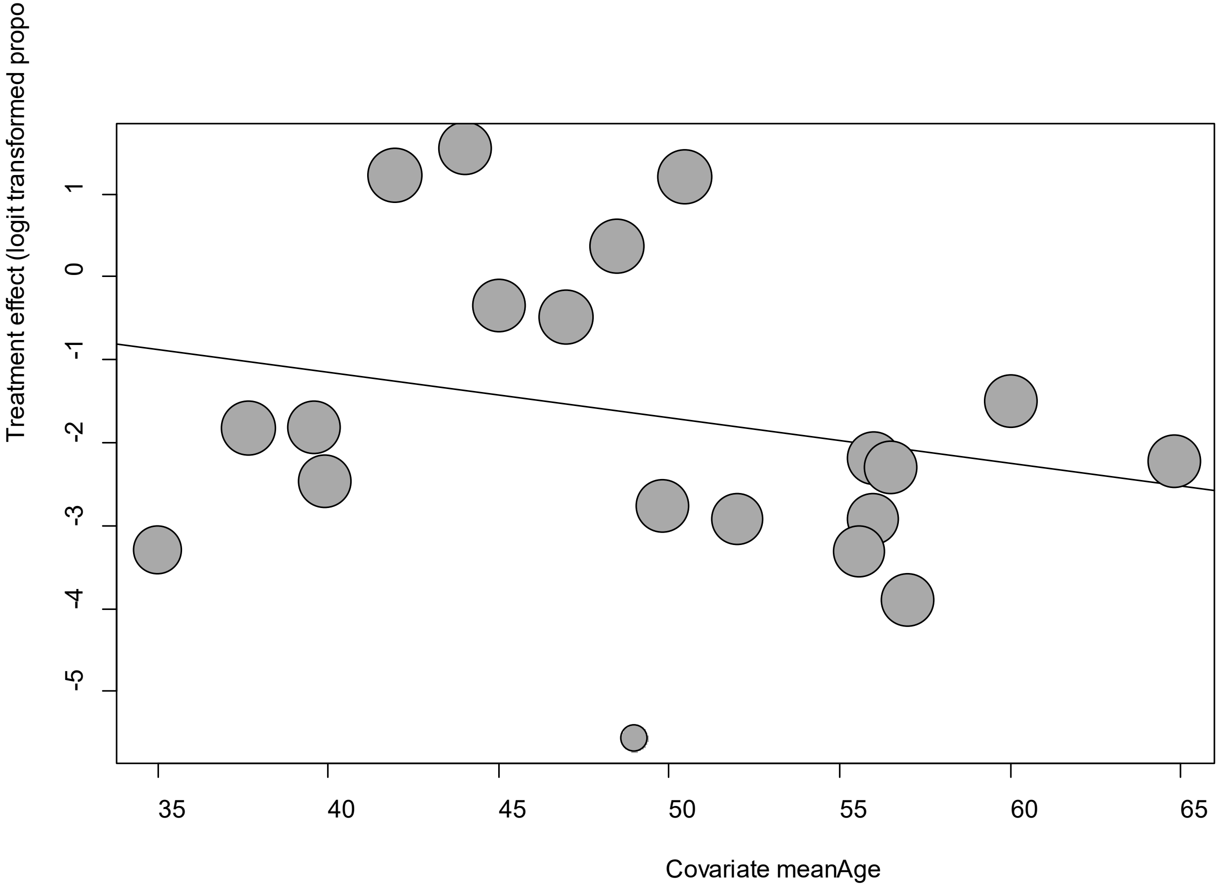


Point sizes are proportional to an inverse of the precision of the estimates.

The regression coefficient was 0.07 (95% confidence interval:-0.33-0.47).(*P*=0.74).

**eFigure 5d. Bubble Plots (sex) on Headache**

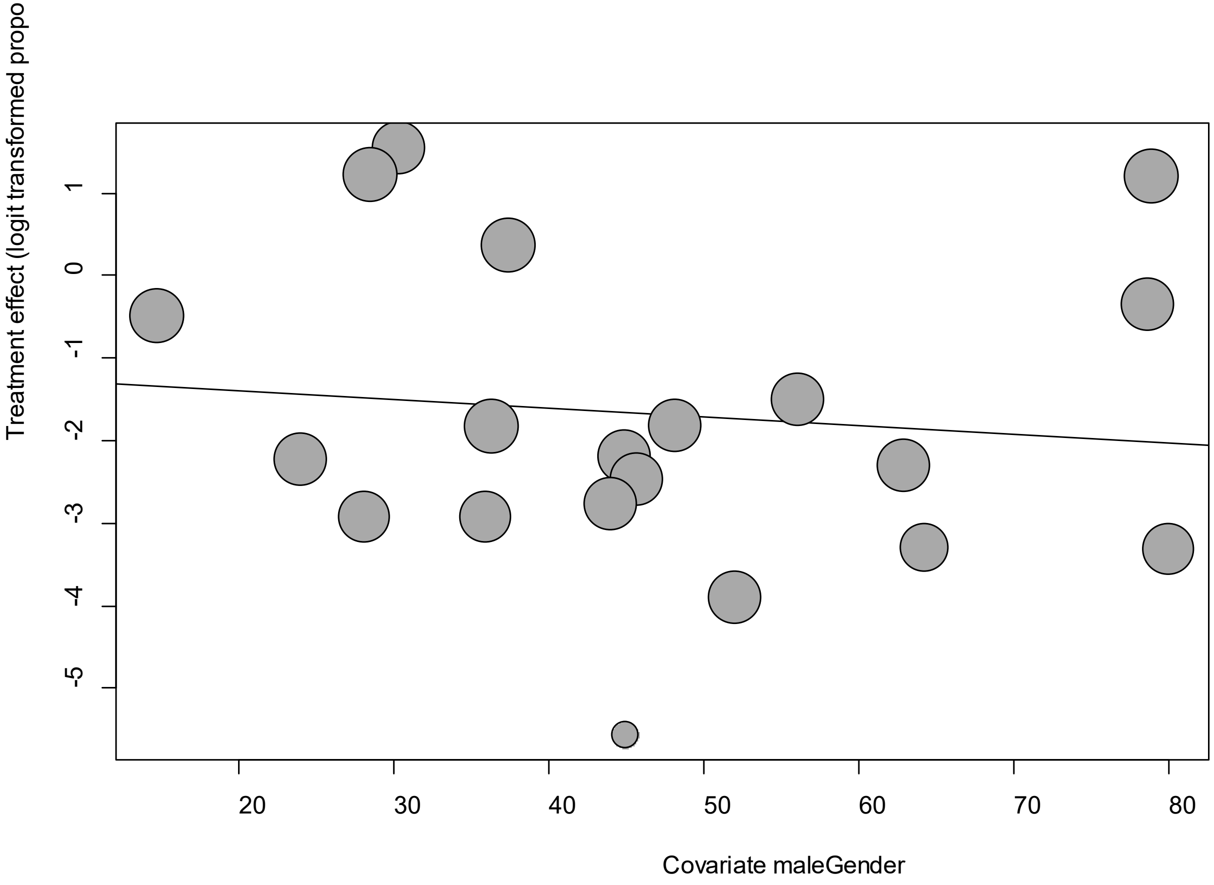


Point sizes are proportional to an inverse of the precision of the estimates.

The regression coefficient was -0.01 (95% confidence interval:-0.05-0.03).(*P*=0.62).

**eFigure 5e. Funnel Plot of studies reporting on Headache**

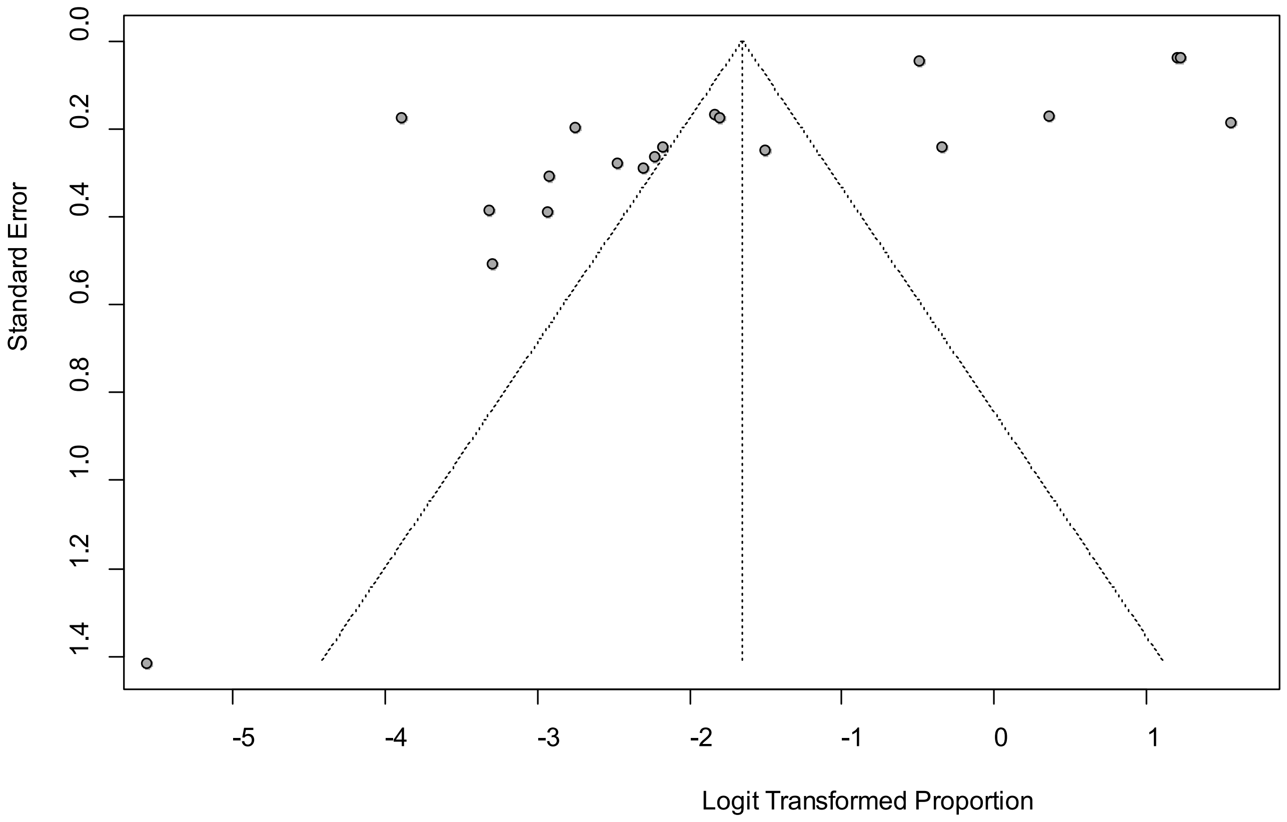


Egger's *P* was 0.0006, indicating the presence of publication bias.

### eFigure 6a. Forest Plot on Myalgia

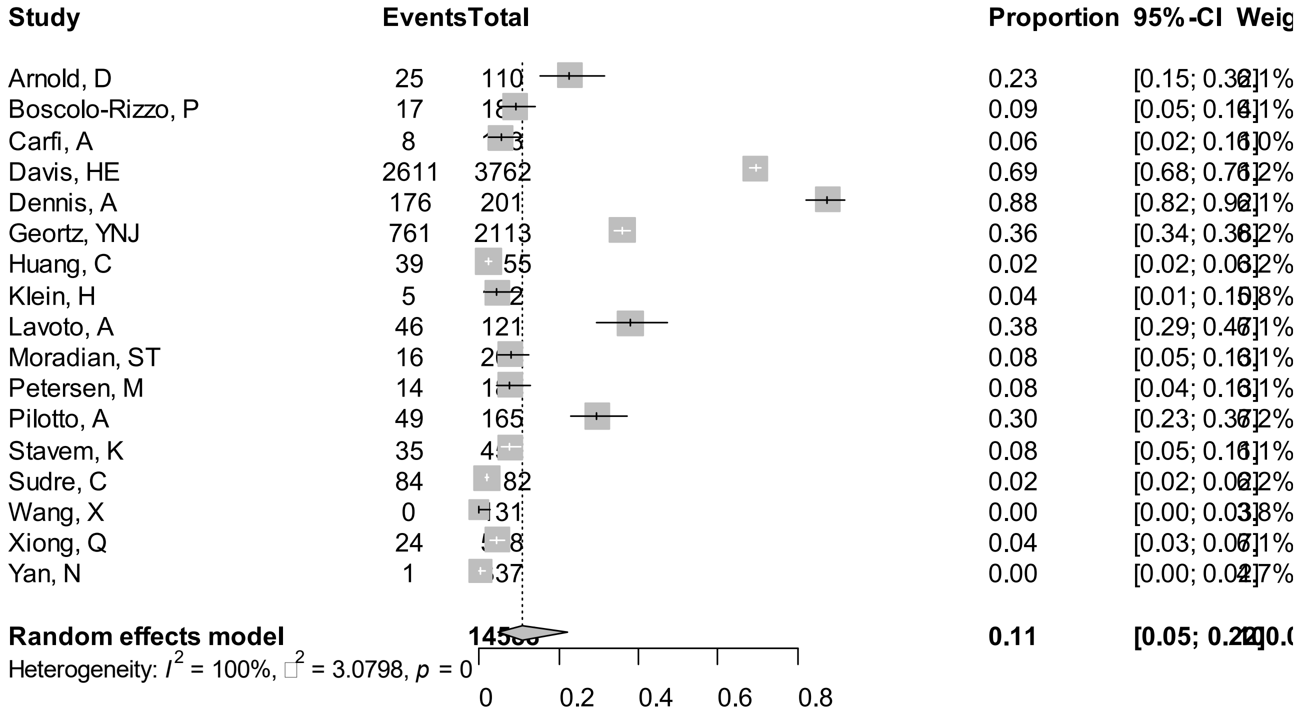


Point sizes are proportional to an inverse of the precision of the estimates and bar correspond to 95% confidence intervals.

**eFigure 6b. Bubble Plots (follow-up period) on Myalgia**

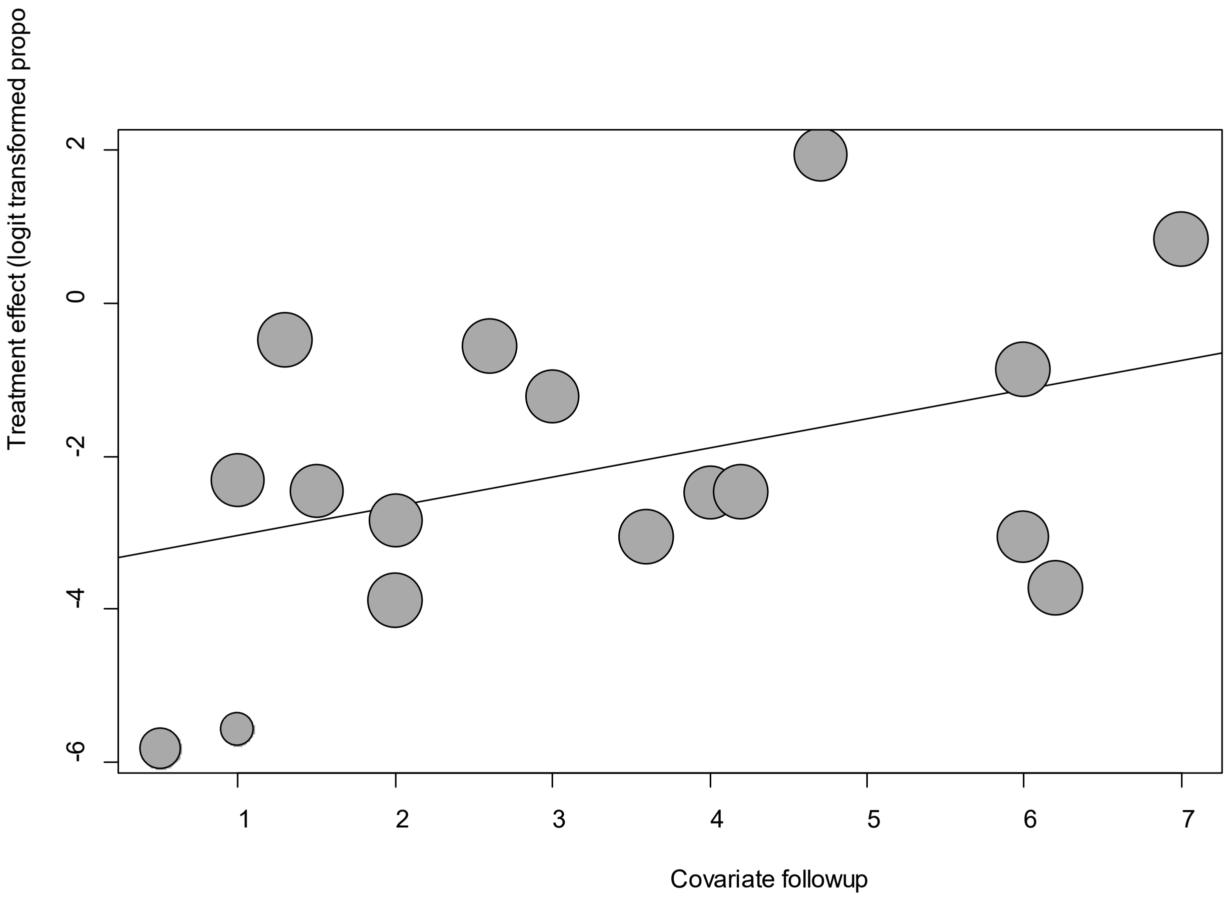


Point sizes are proportional to an inverse of the precision of the estimates.

The regression coefficient was 0.38 (95% confidence interval:--0.07-0.83).(*P*=0.09).

**eFigure 6c. Bubble Plots (age) on Myalgia**

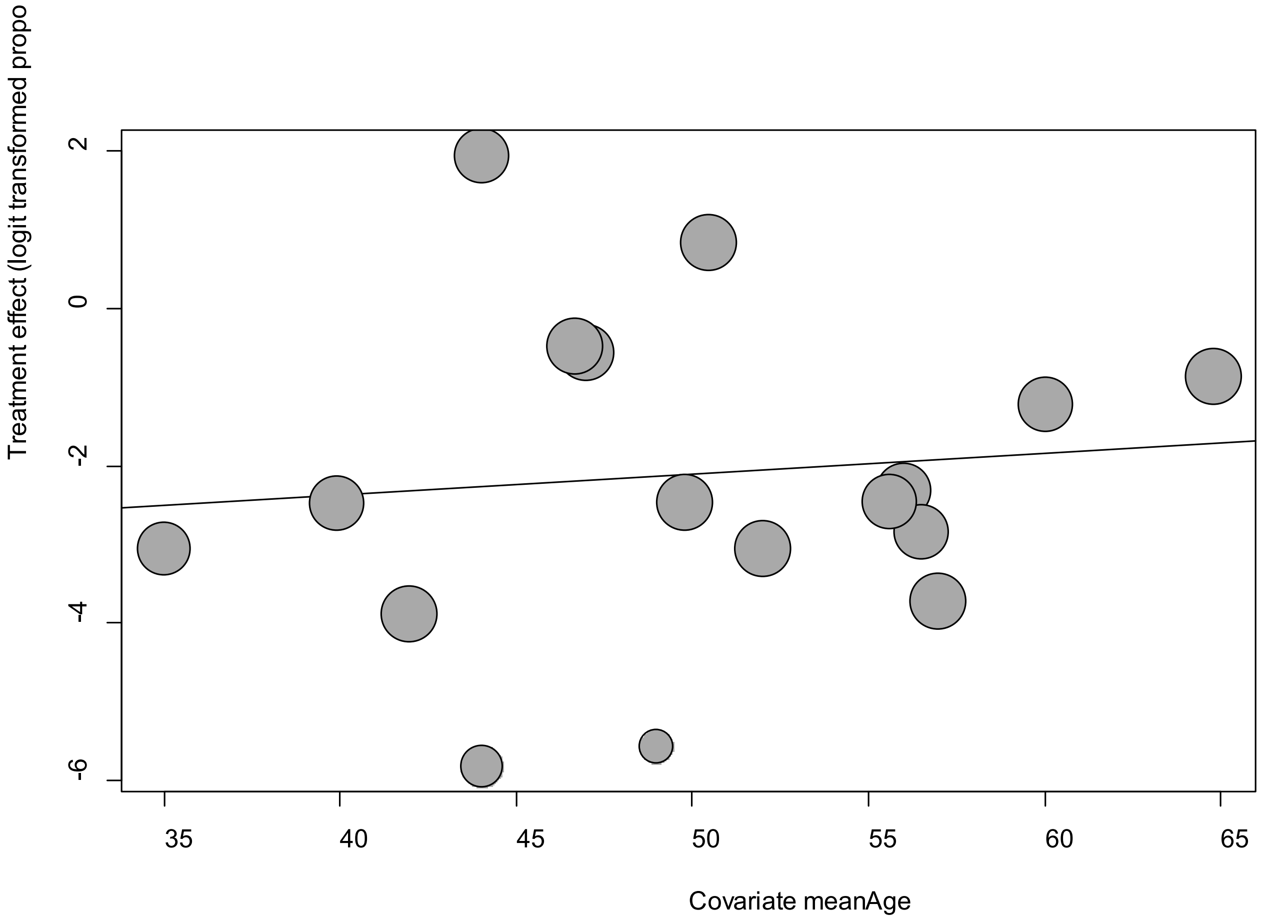


Point sizes are proportional to an inverse of the precision of the estimates.

The regression coefficient was 0.03 (95% confidence interval:-0.09-0.14).(*P*=0.66).

**eFigure 6d. Bubble Plots (sex) on Myalgia**

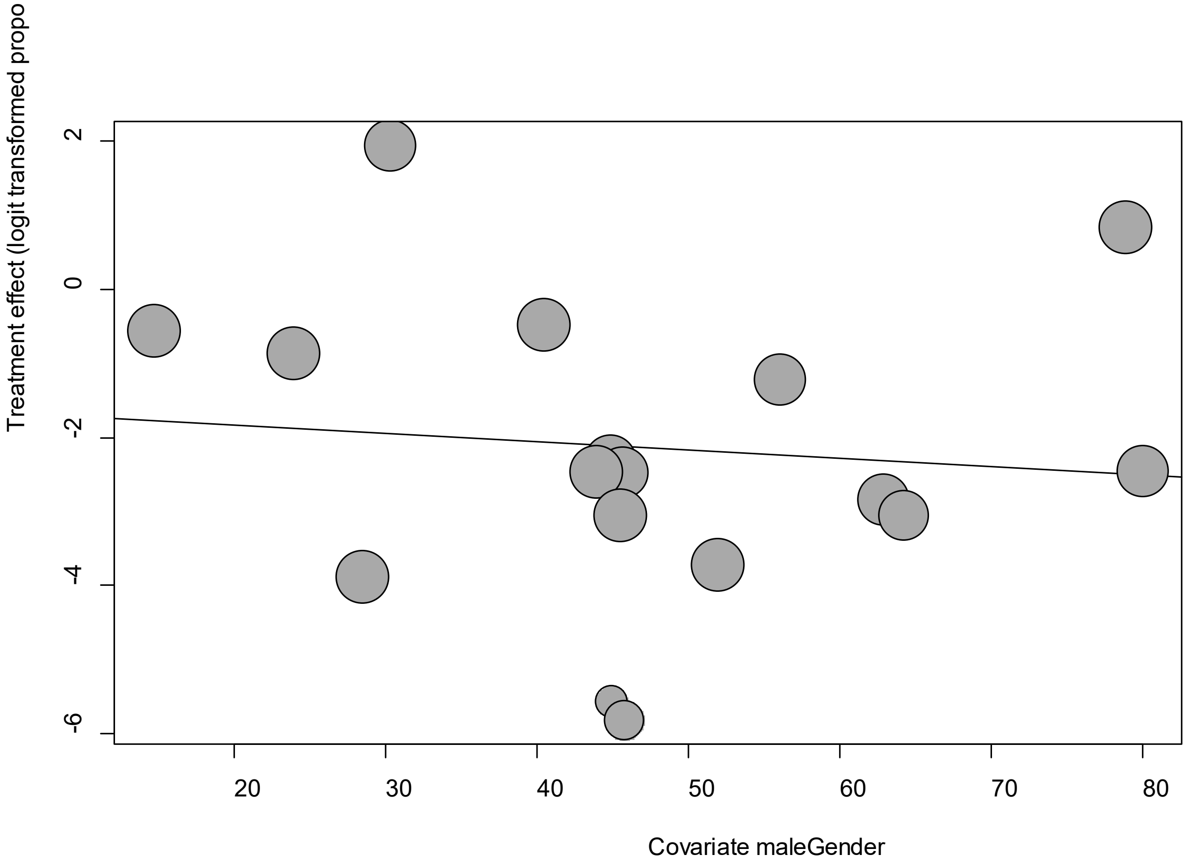


Point sizes are proportional to an inverse of the precision of the estimates.

The regression coefficient was -0.01 (95% confidence interval:-0.07-0.05).(*P*=0.72).

**eFigure 6e. Funnel Plot of studies reporting on Myalgia**

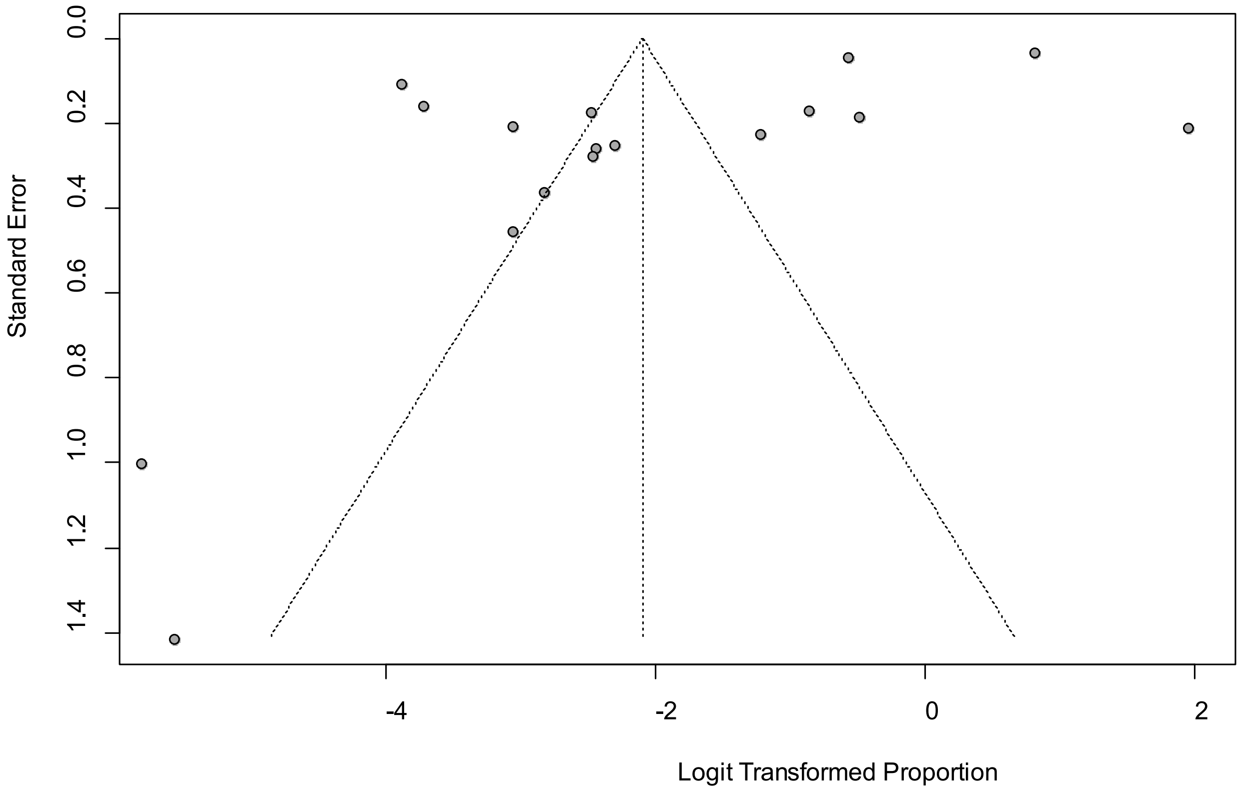


Egger's *P* was 0.011, indicating the presence of publication bias.

**eFigure 7a. Forest Plot on Neuralgia**

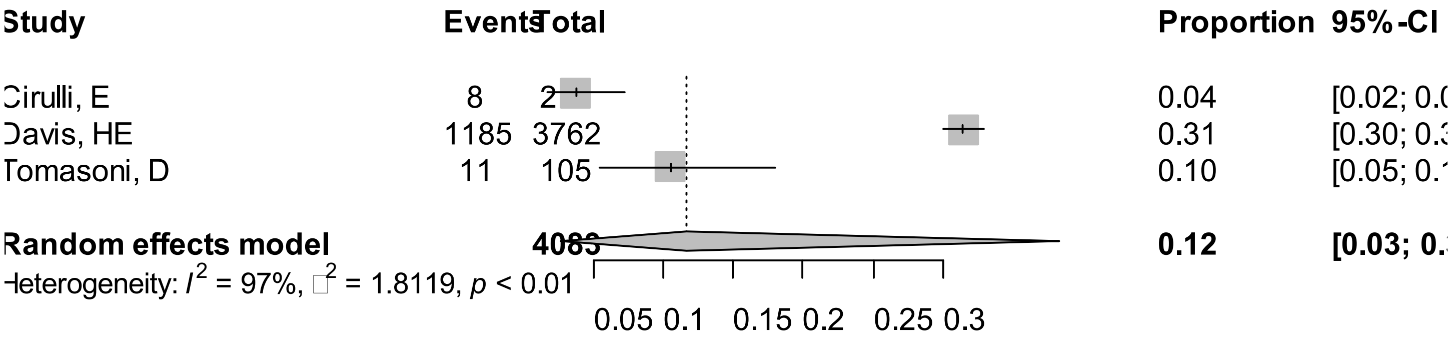


Point sizes are proportional to an inverse of the precision of the estimates and bar correspond to 95% confidence intervals.

**eFigure 7b. Bubble Plots (follow-up period) on Neuralgia**

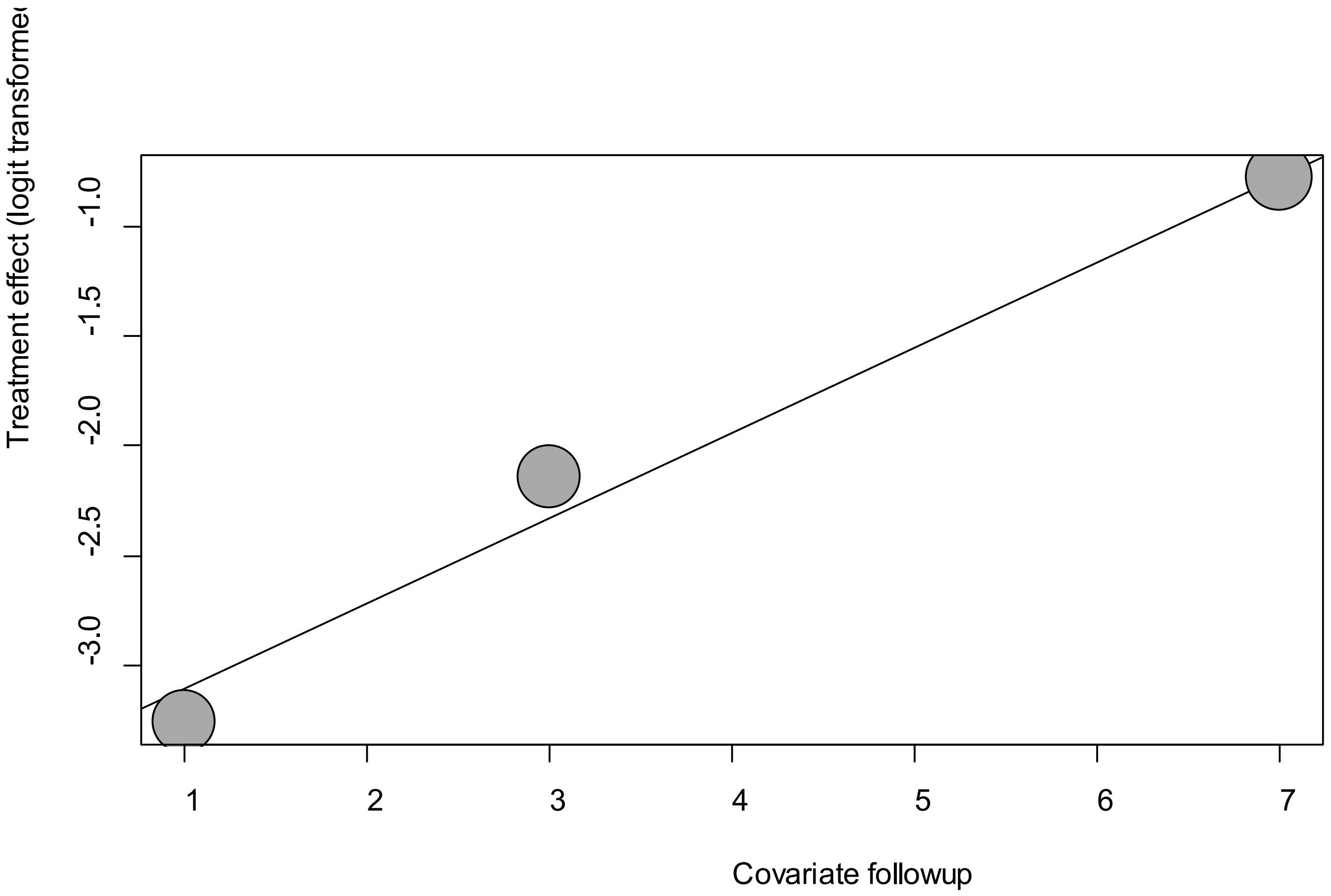


Point sizes are proportional to an inverse of the precision of the estimates.

The regression coefficient was 0.39 (95% confidence interval:0.29-0.48).(*P*<0.001).

**eFigure 7c. Bubble Plots (age) on Neuralgia**

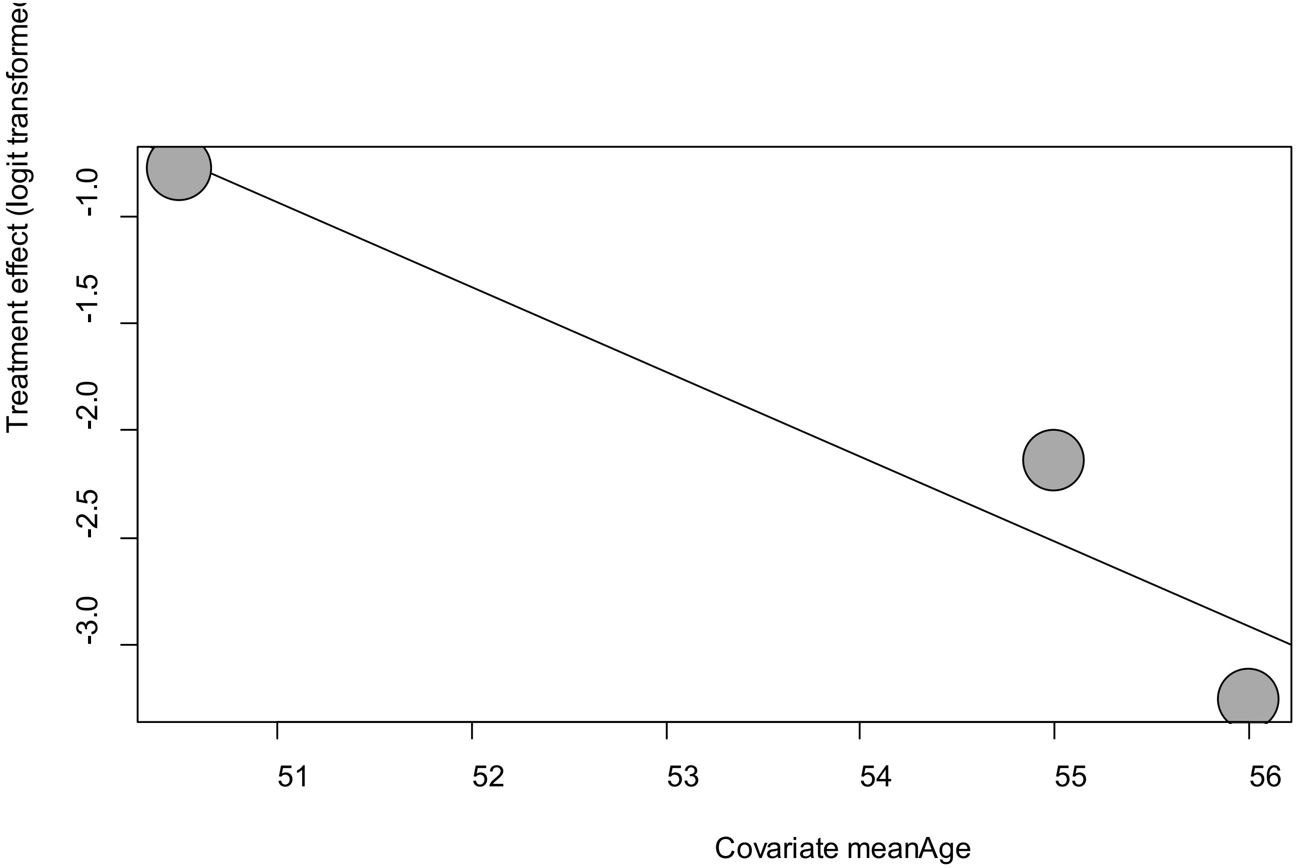


Point sizes are proportional to an inverse of the precision of the estimates.

The regression coefficient was -0.40 (95% confidence interval:-0.60—0.19).(*P*=0.0002).

**eFigure 7d. Bubble Plots (sex) on Neuralgia**

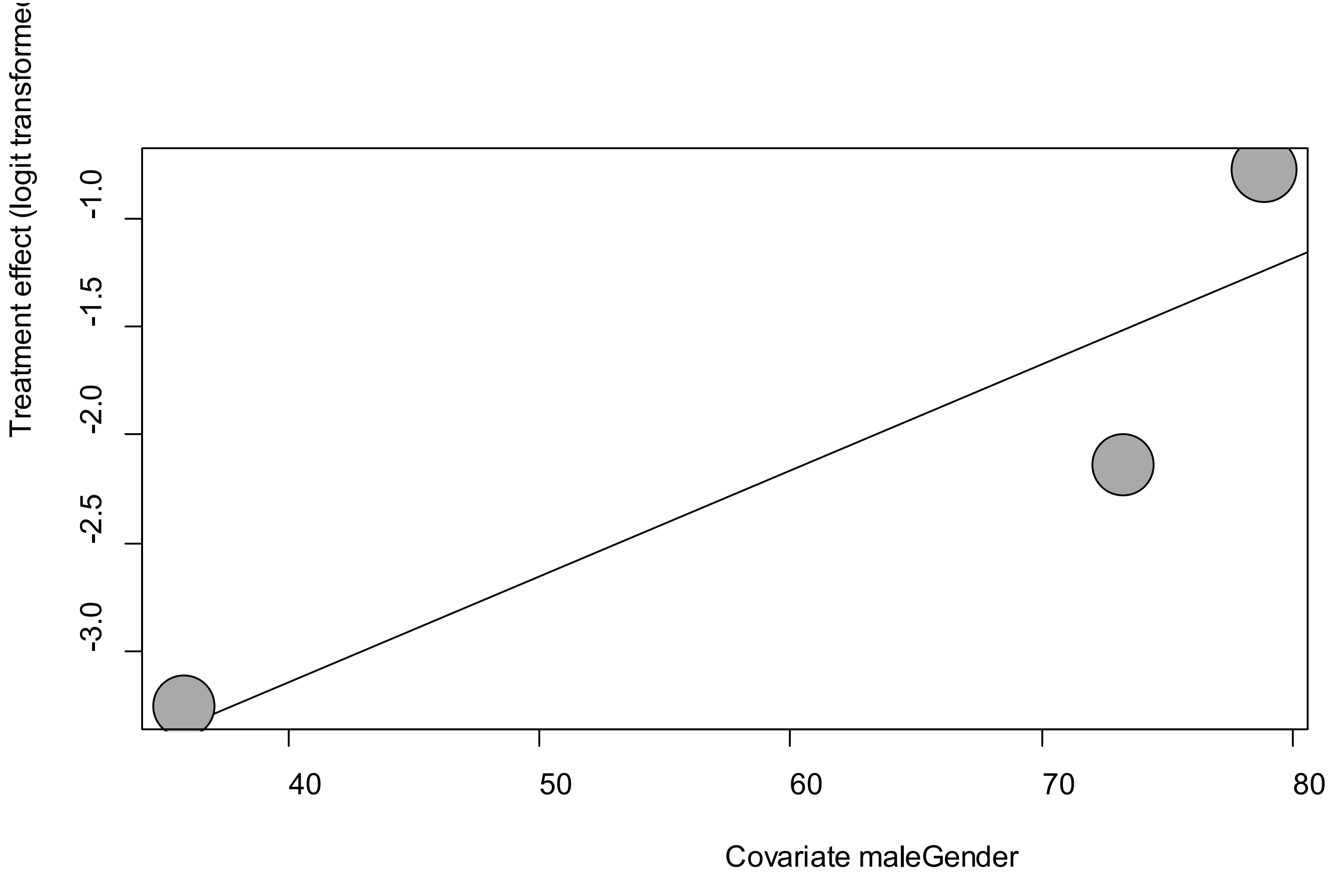


Point sizes are proportional to an inverse of the precision of the estimates.

The regression coefficient was 0.005 (95% confidence interval:0.001-0.10).(*P*=0.045).

**eFigure 7e. Funnel Plot of studies reporting on Neuralgia**

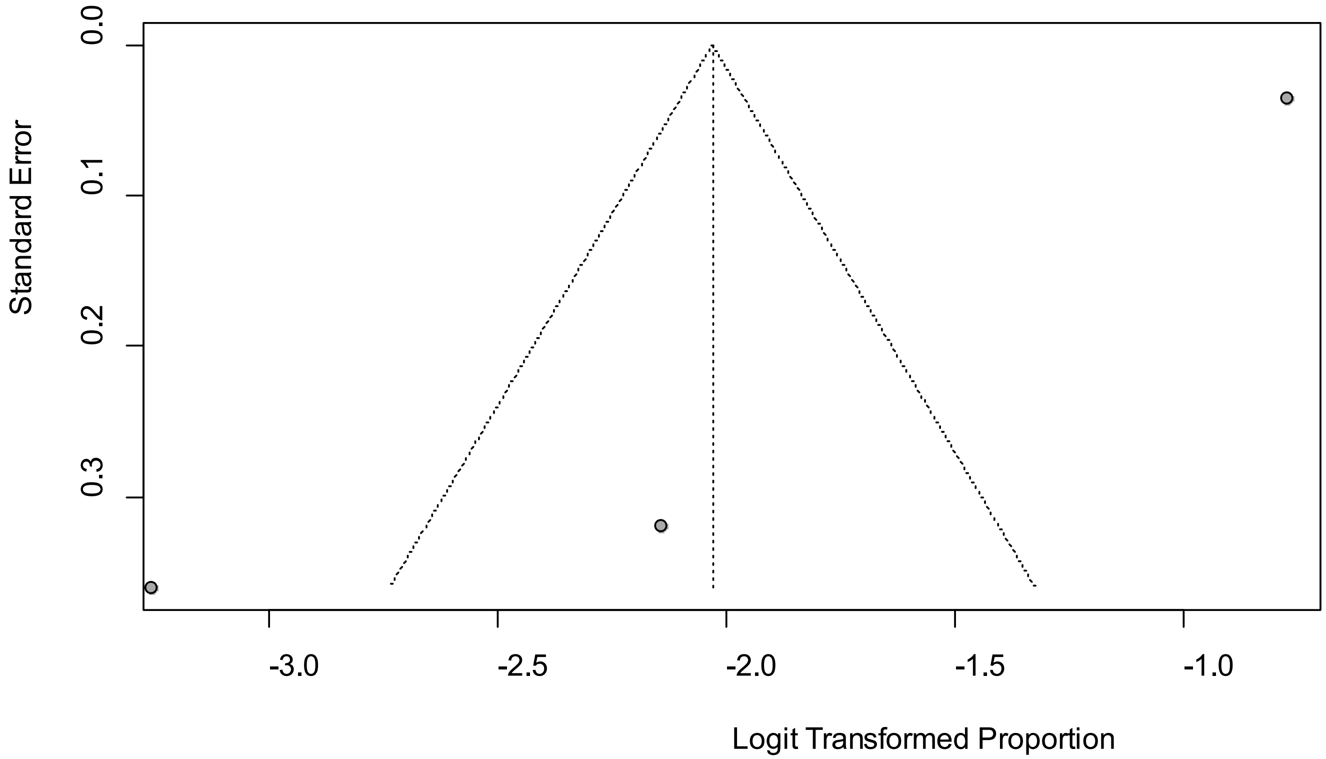


Egger’s *P* was 0.14, indicating the absence of publication bias.

### eFigure 8a. Forest Plot on Sore Throat

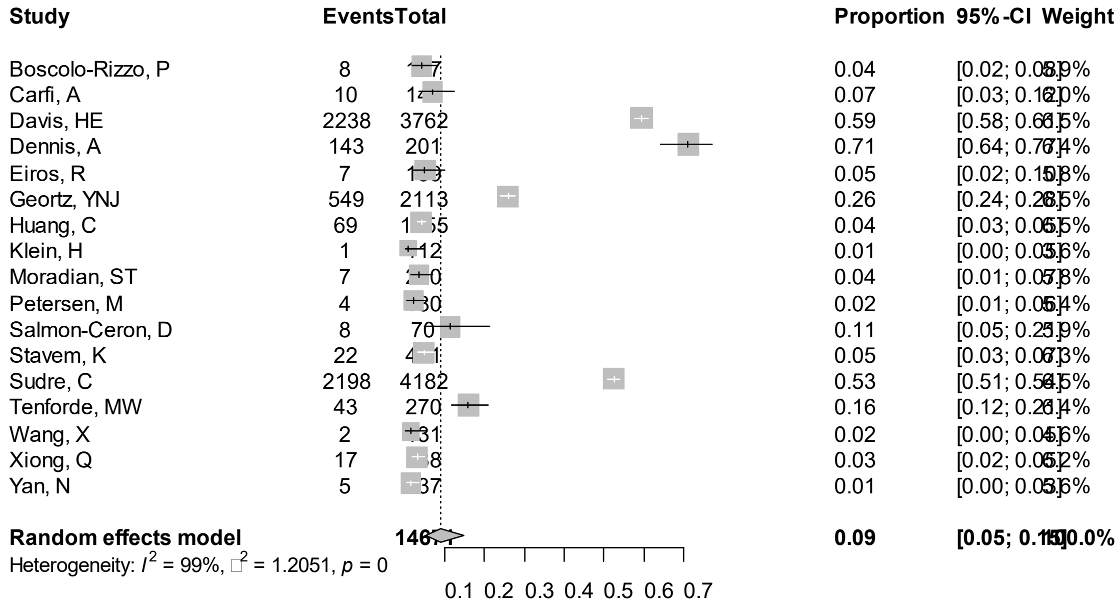


Point sizes are proportional to an inverse of the precision of the estimates and bar correspond to 95% confidence intervals.

### eFigure 8b. Bubble Plots (follow-up period) on Sore Throat

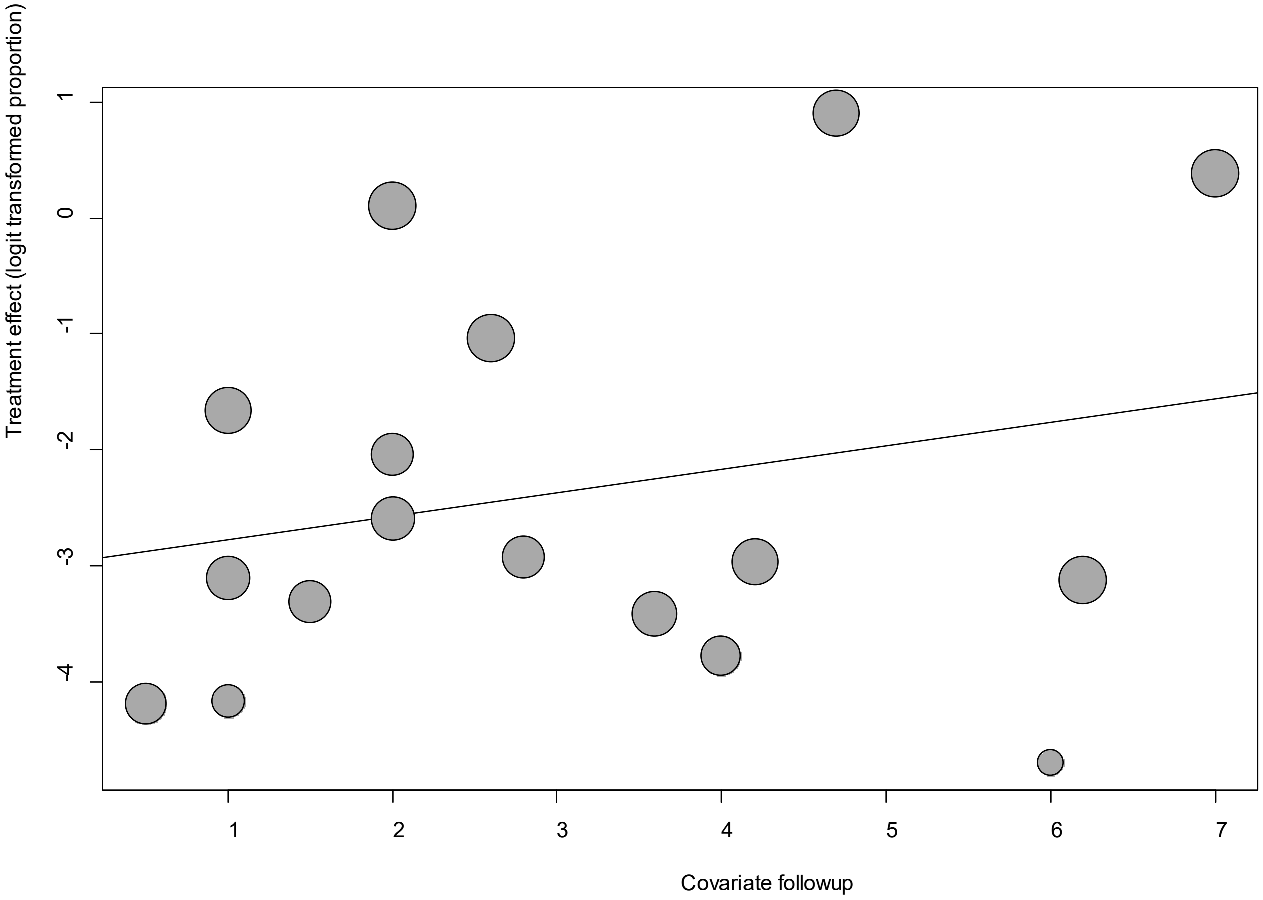


Point sizes are proportional to an inverse of the precision of the estimates.

The regression coefficient was 0.20 (95% confidence interval:-0.17-0.58).(*P*=0.29).

**eFigure 8c. Bubble Plots (age) on Sore Throat**

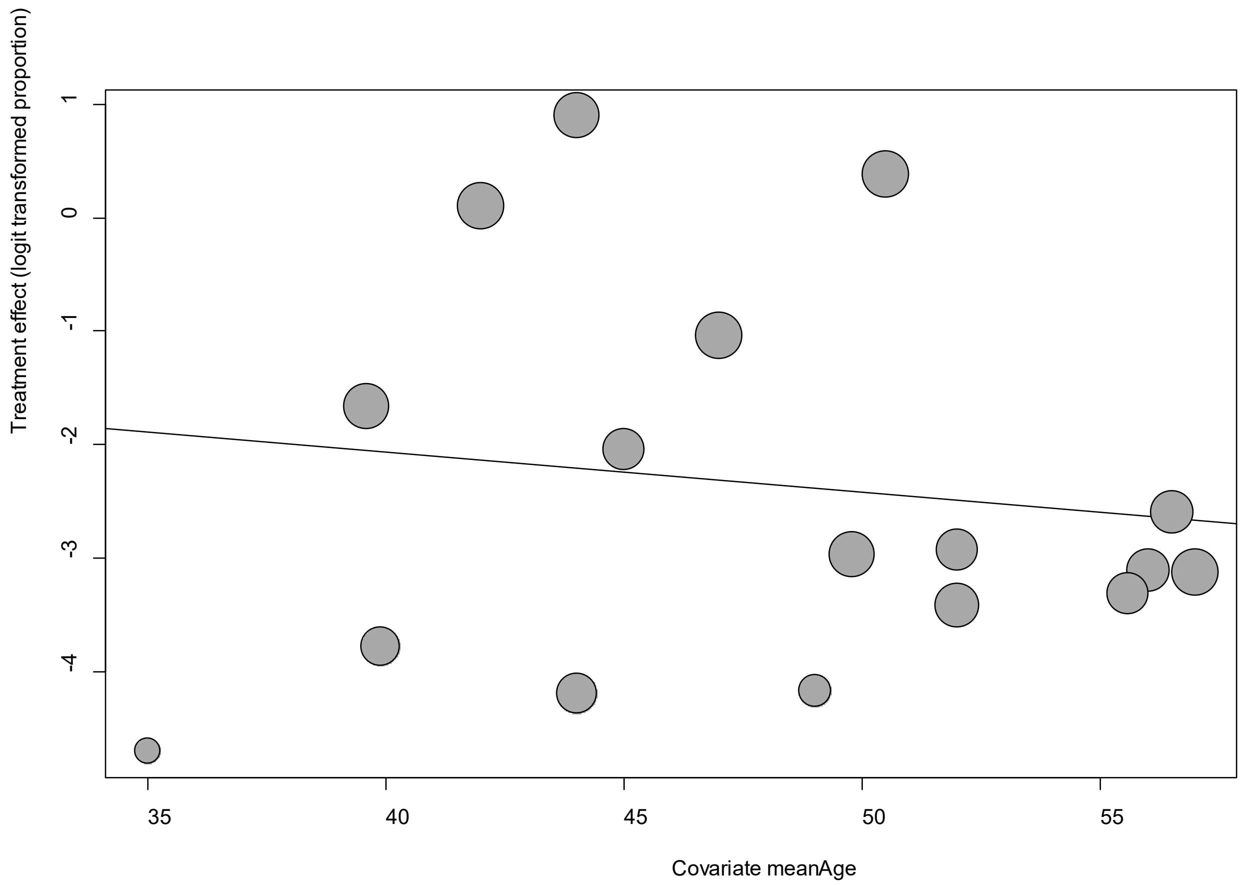


Point sizes are proportional to an inverse of the precision of the estimates.

The regression coefficient was -0.04 (95% confidence interval:-0.15-0.08).(*P*=0.53).

### eFigure 8d. Bubble Plots (sex) on Sore Throat

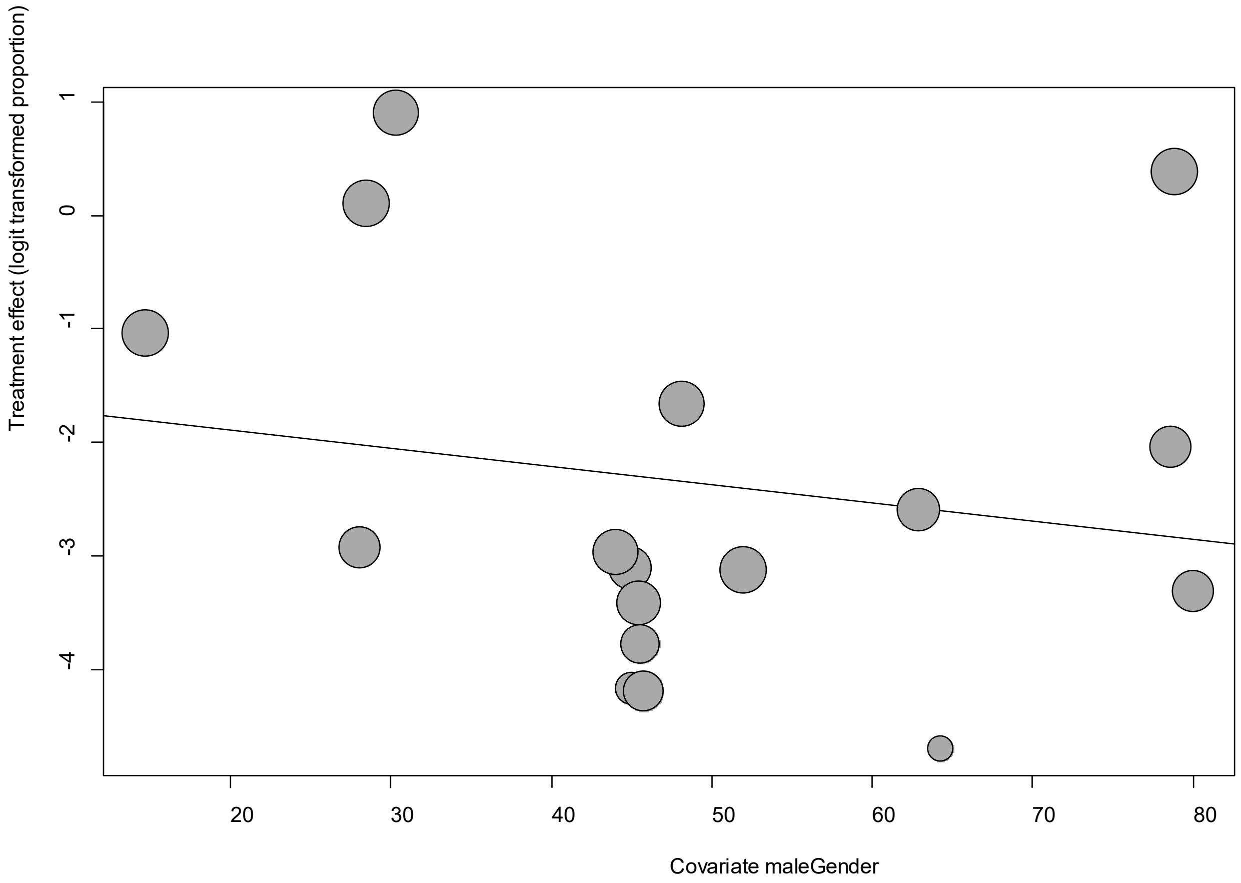


Point sizes are proportional to an inverse of the precision of the estimates.

The regression coefficient was -0.02 (95% confidence interval: -0.05-0.02).(*P*=0.40).

**eFigure 8e. Funnel Plot of studies reporting on Sore Throat**

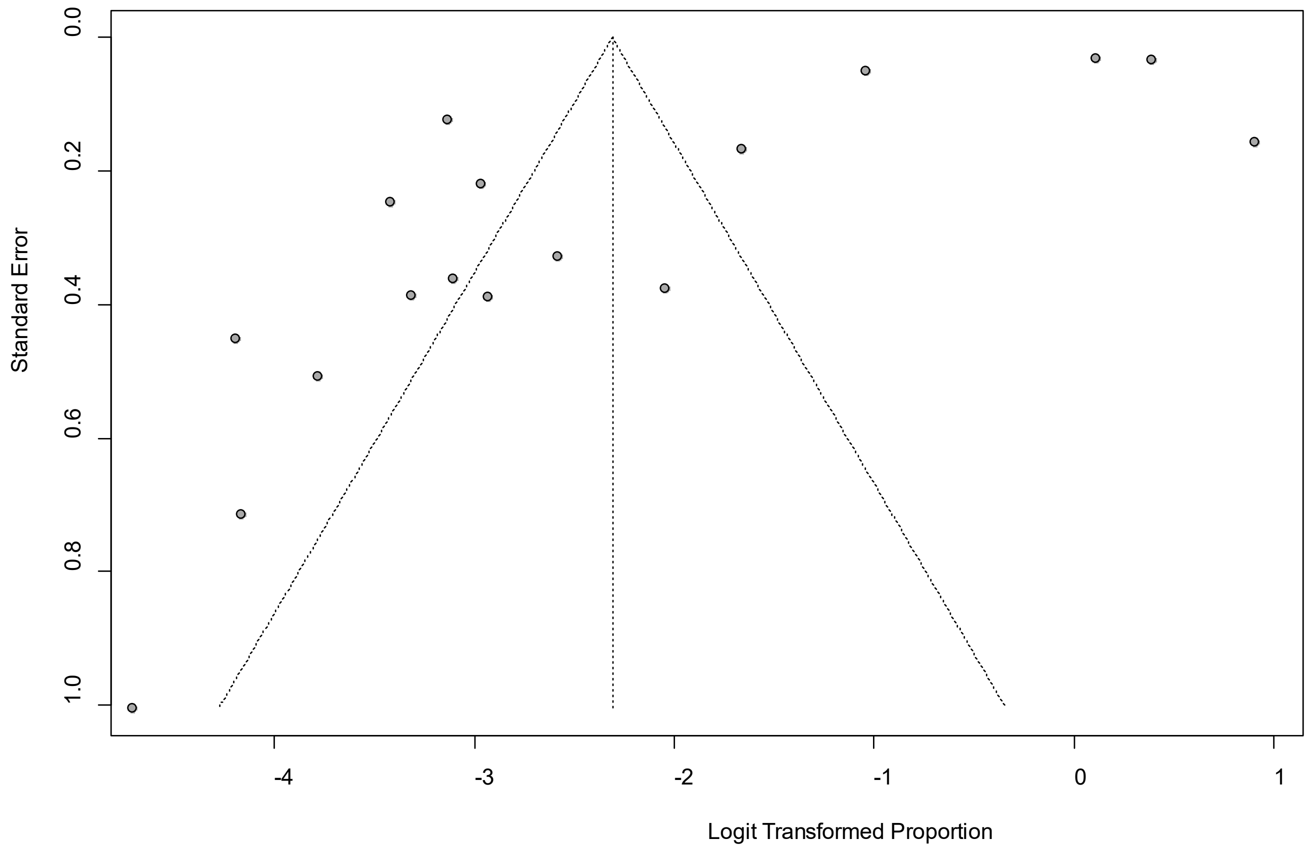


Egger's *P* was 0.40, indicating the absence of publication bias.

**eFigure 9a. Forest Plot on Ageusia**

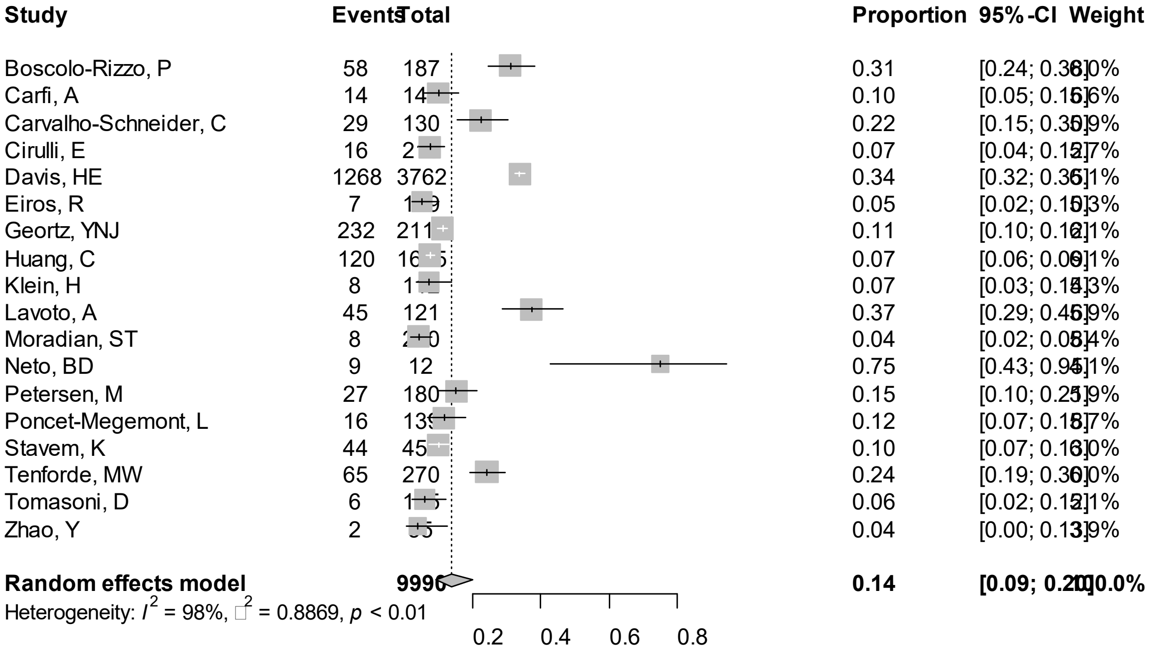


Point sizes are proportional to an inverse of the precision of the estimates and bar correspond to 95% confidence intervals.

### eFigure 9b. Bubble Plots (follow-up period) on Ageusia

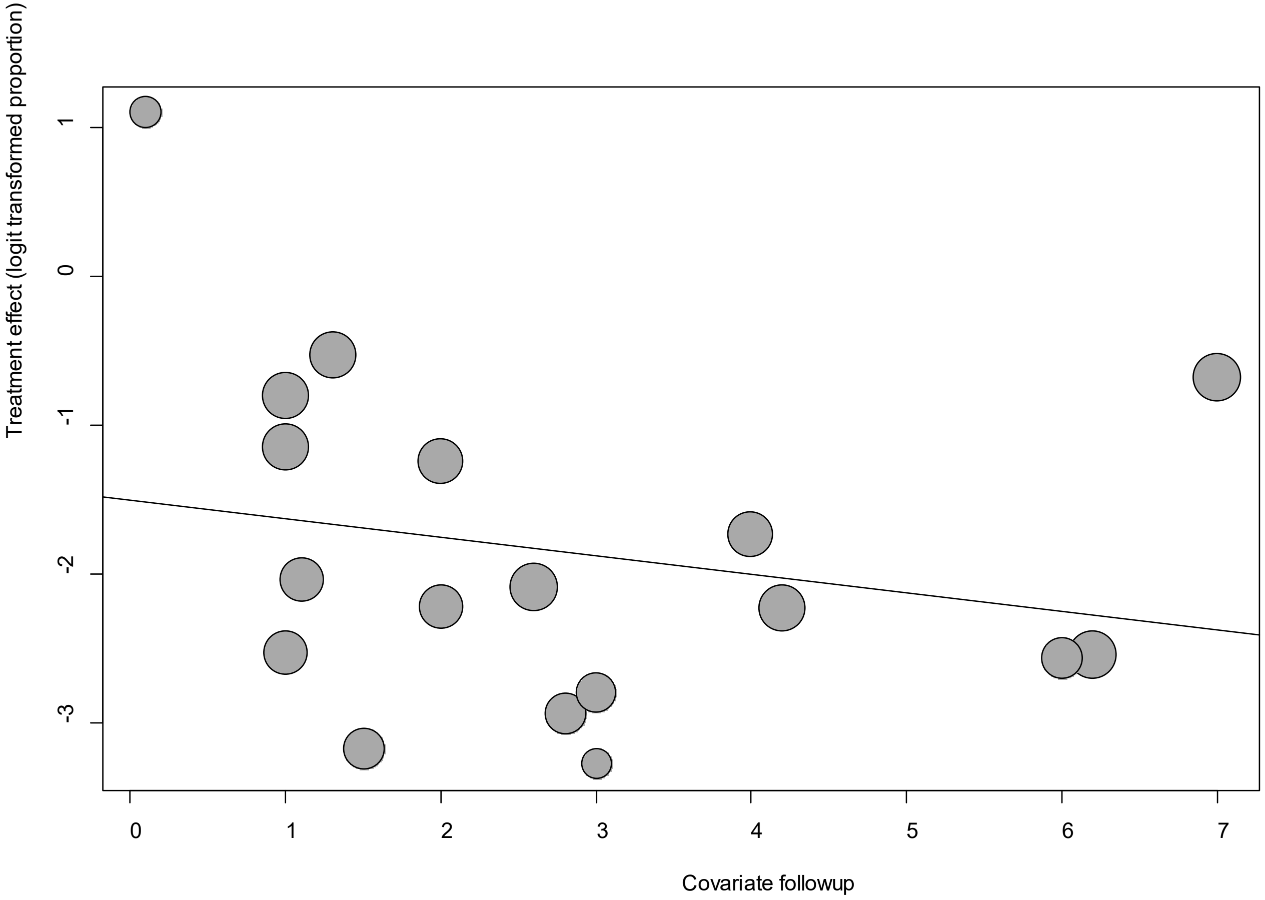


Point sizes are proportional to an inverse of the precision of the estimates.

The regression coefficient was -0.12 (95% confidence interval:-0.37-0.12).(*P*=0.32).

**eFigure 9c. Bubble Plots (age) on Ageusia**

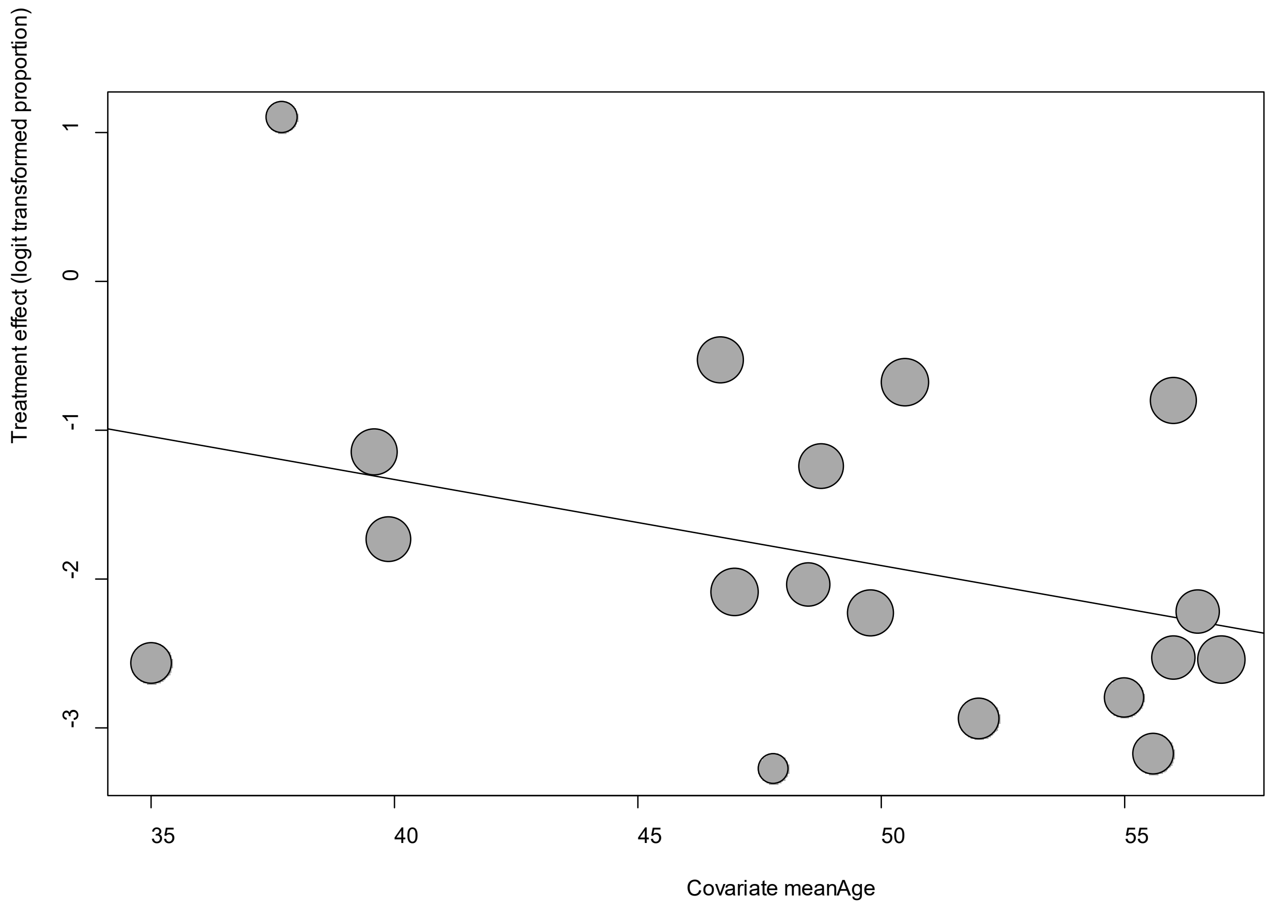


Point sizes are proportional to an inverse of the precision of the estimates.

The regression coefficient was -0.06 (95% confidence interval:-0.13-0.01).(*P*=0.11).

**eFigure 9d. Bubble Plots (sex) on Ageusia**

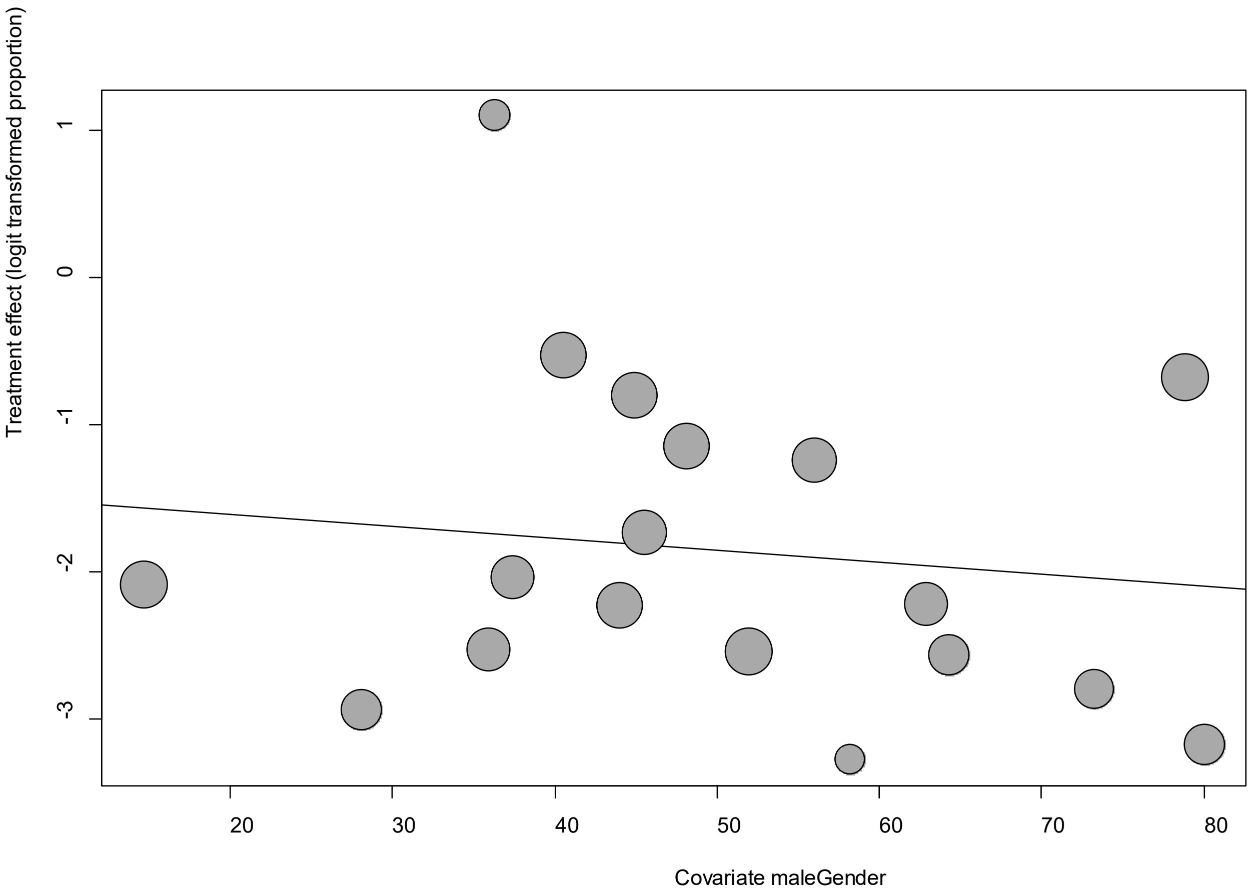


Point sizes are proportional to an inverse of the precision of the estimates.

The regression coefficient was -0.08 (95% confidence interval:-0.03-0.02).(*P*=0.51).

**eFigure 9e. Funnel Plot of studies reporting on Ageusia**

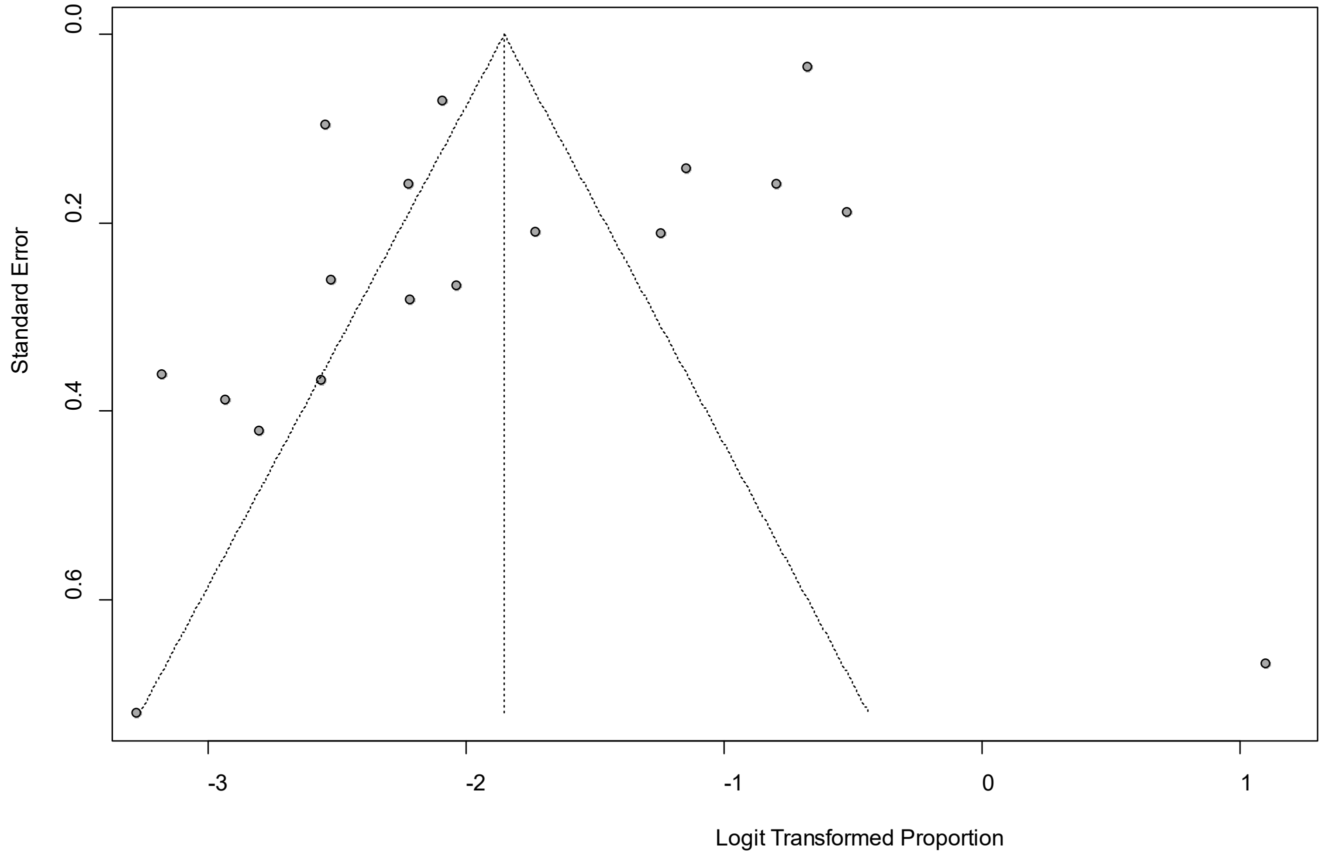


Egger's *P* was 0.03, indicating the absence of publication bias.

**eFigure 10a. Forest Plot on Alopecia**

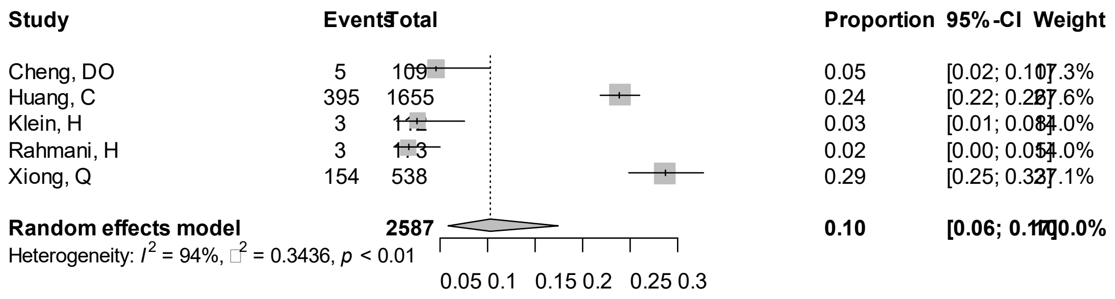


Point sizes are proportional to an inverse of the precision of the estimates and bar correspond to 95% confidence intervals.

**eFigure 10b. Bubble Plots (follow-up period) on Alopecia**

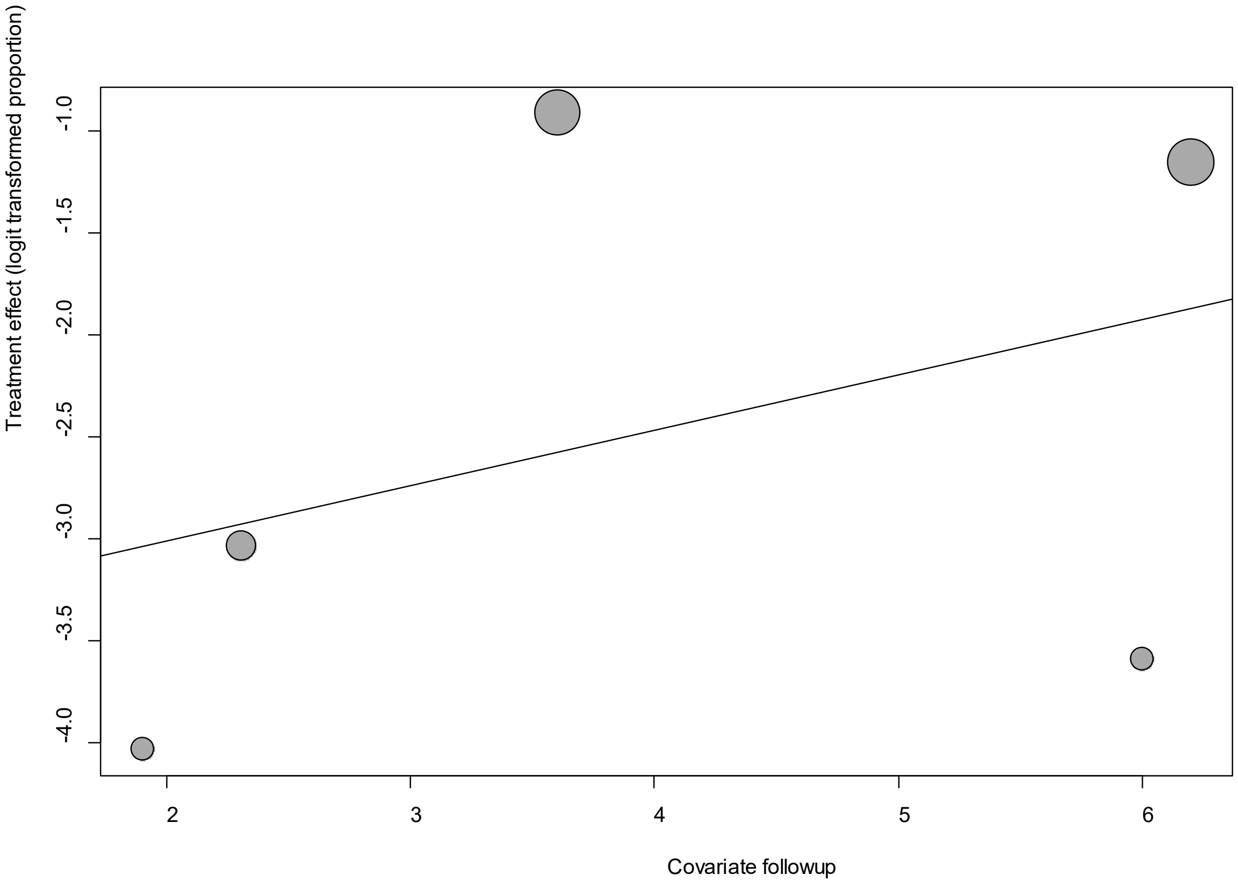


Point sizes are proportional to an inverse of the precision of the estimates.

The regression coefficient was 0.27 (95% confidence interval:-0.46-1.00).(*P*=0.46).

**eFigure 10c. Bubble Plots (age) on Alopecia**

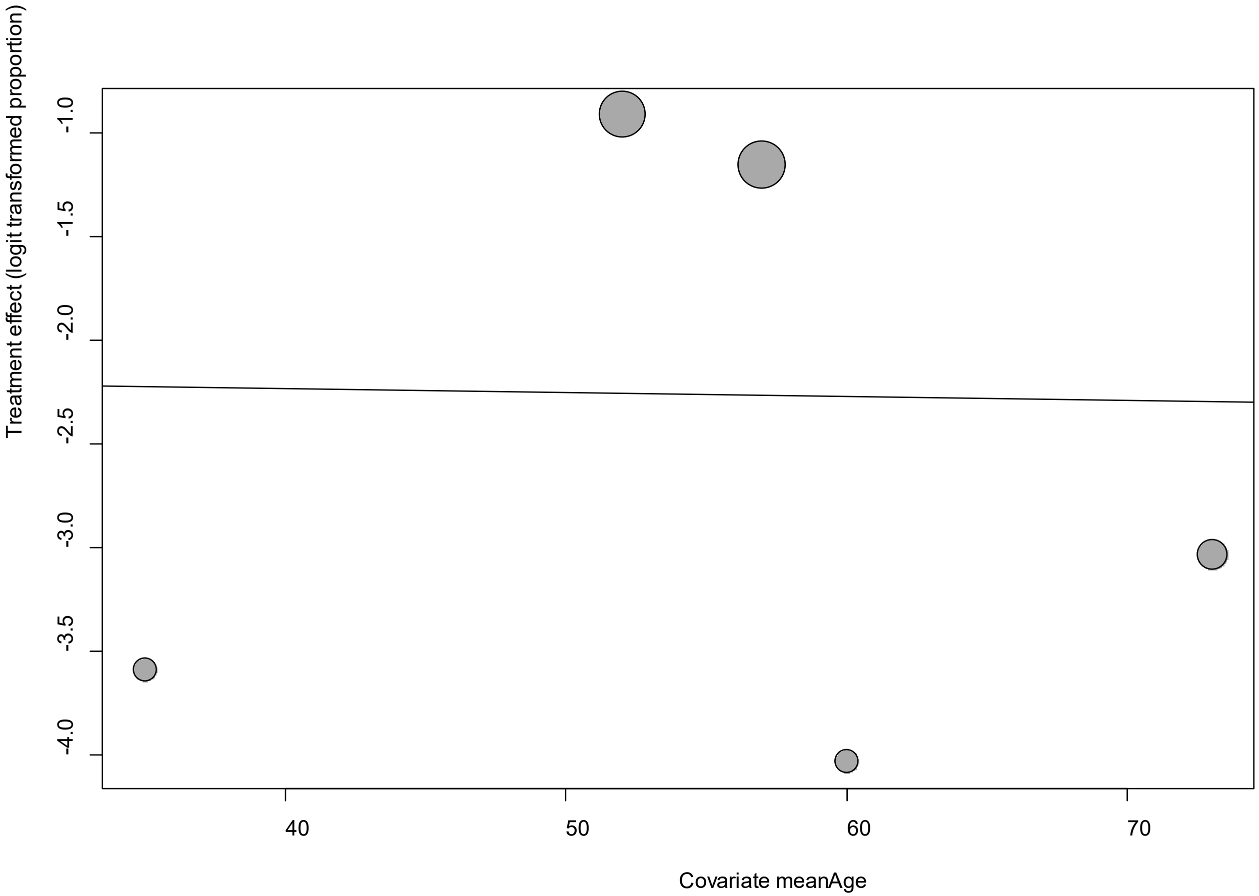


Point sizes are proportional to an inverse of the precision of the estimates.

The regression coefficient was -0.002 (95% confidence interval:-0.07-0.06).(*P*=0.95).

**eFigure 10d. Bubble Plots (sex) on Alopecia**

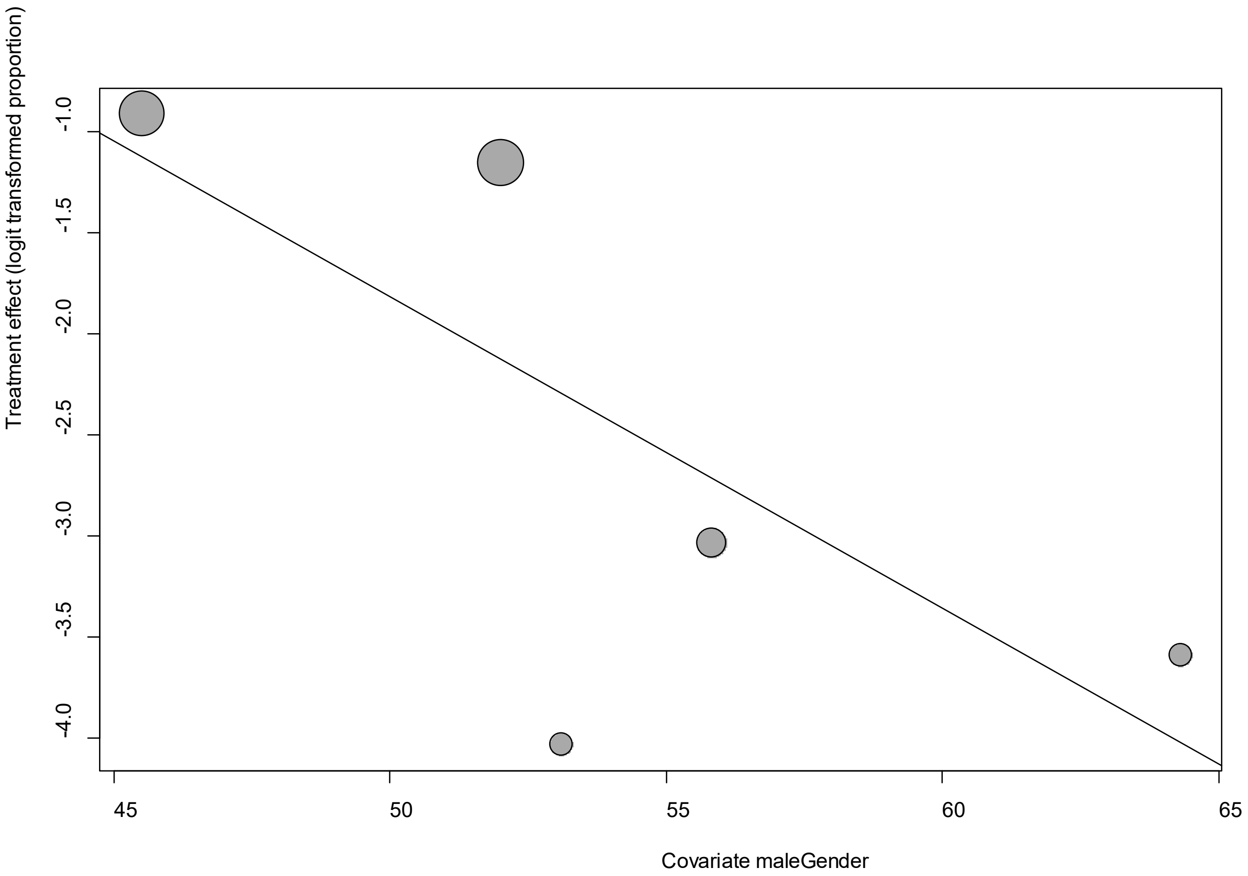


Point sizes are proportional to an inverse of the precision of the estimates.

The regression coefficient was -0.15 (95% confidence interval:-0.29—0.01).(*P*=0.03).

**eFigure 10e. Funnel Plot of studies reporting on Alopecia**

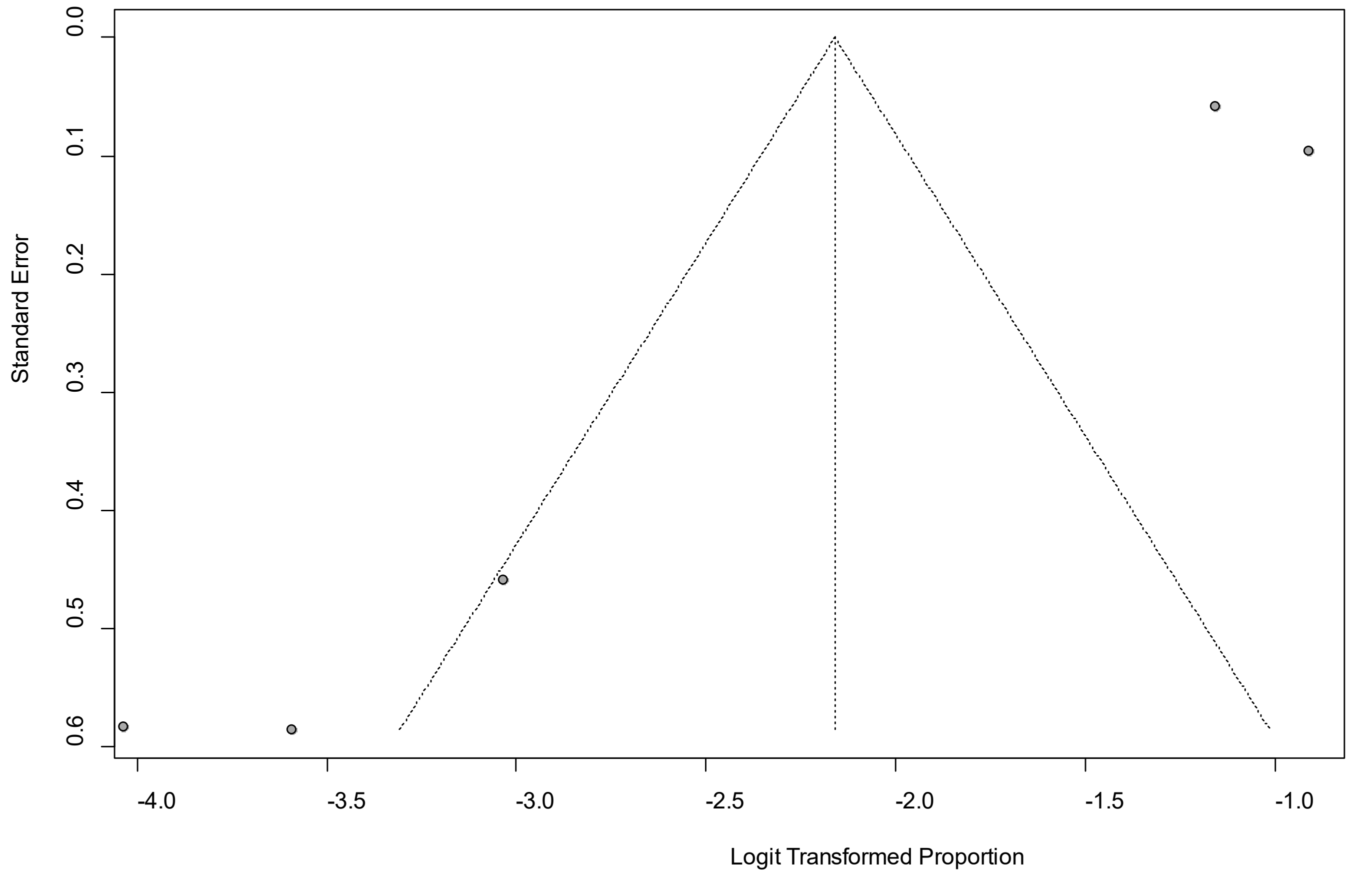


Egger's *P* was 0.052, indicating the presence of publication bias.

### eFigure 11a. Forest Plot on Anorexia

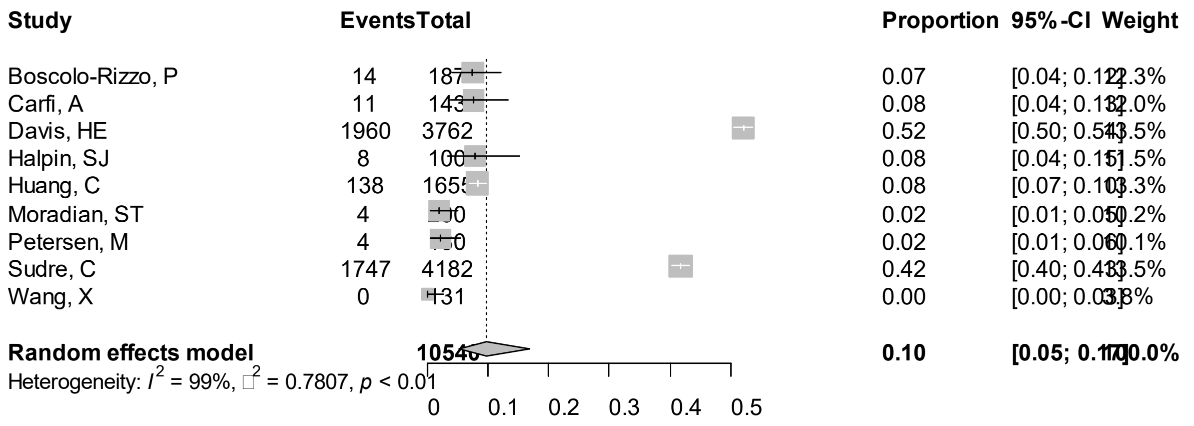


Point sizes are proportional to an inverse of the precision of the estimates and bar correspond to 95% confidence intervals.

**eFigure 11b. Bubble Plots (follow-up period) on Anorexia**

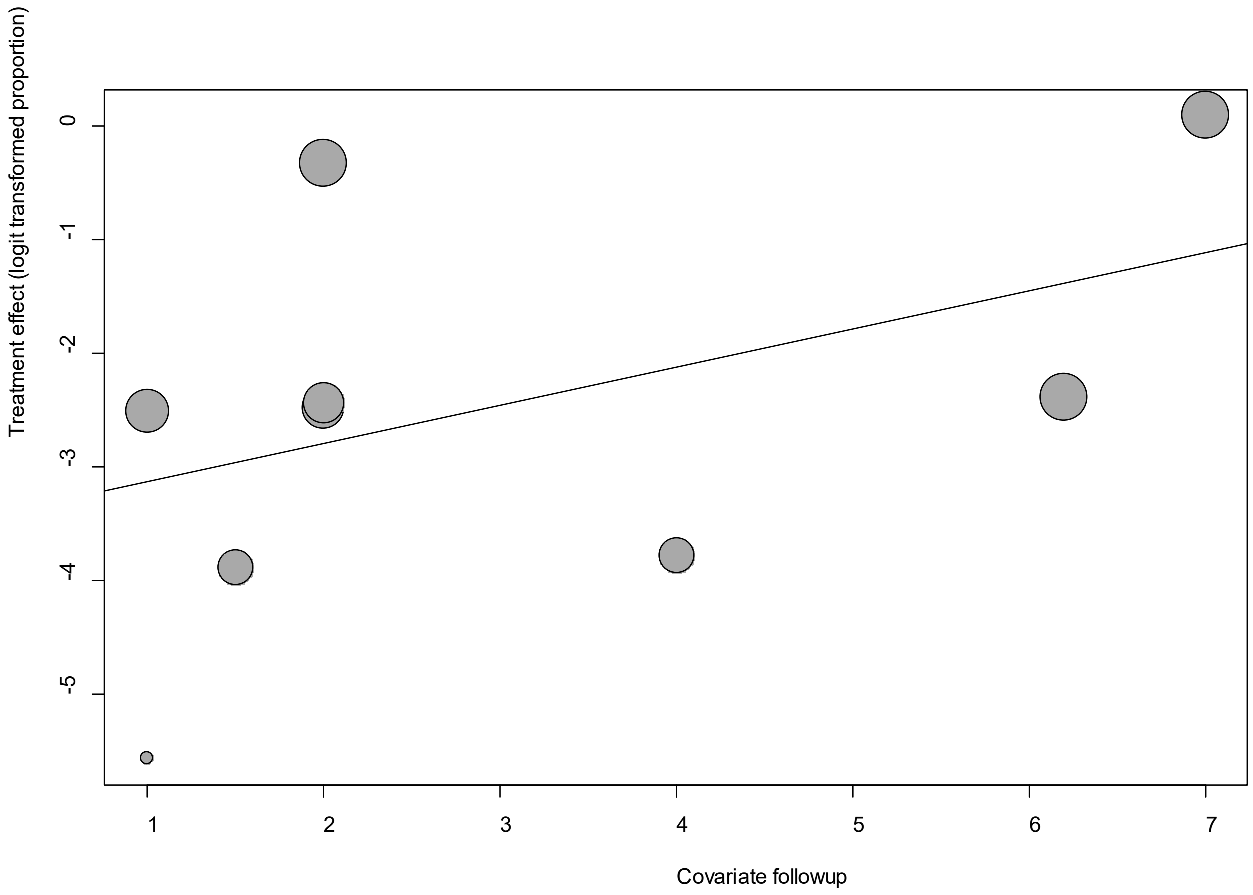


Point sizes are proportional to an inverse of the precision of the estimates.

The regression coefficient was 0.34 (95% confidence interval:-0.23-0.90).(*P*=0.24).

**eFigure 11c. Bubble Plots (age) on Anorexia**

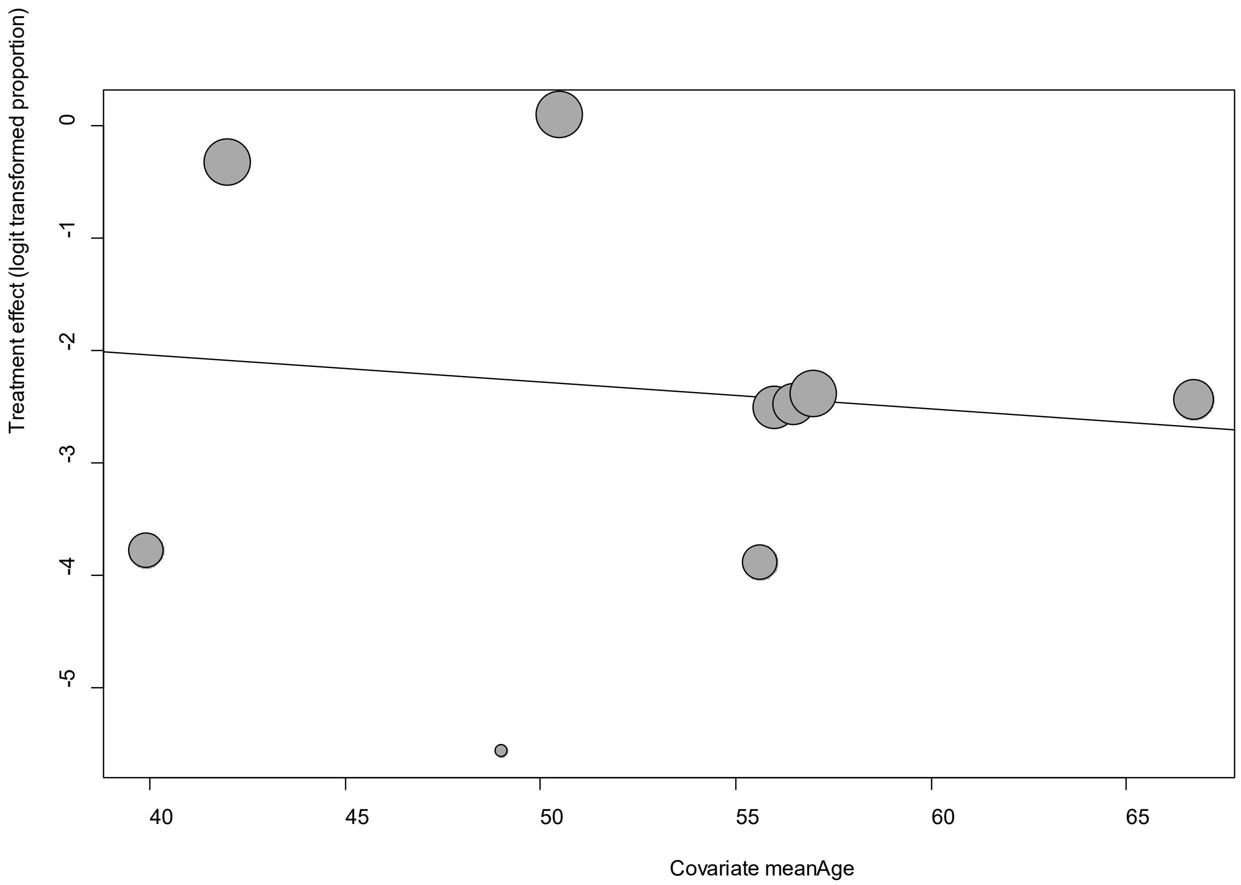


Point sizes are proportional to an inverse of the precision of the estimates.

The regression coefficient was -0.023 (95% confidence interval:-0.14-0.09).(*P*=0.68).

**eFigure 11d. Bubble Plots (sex) on Anorexia**

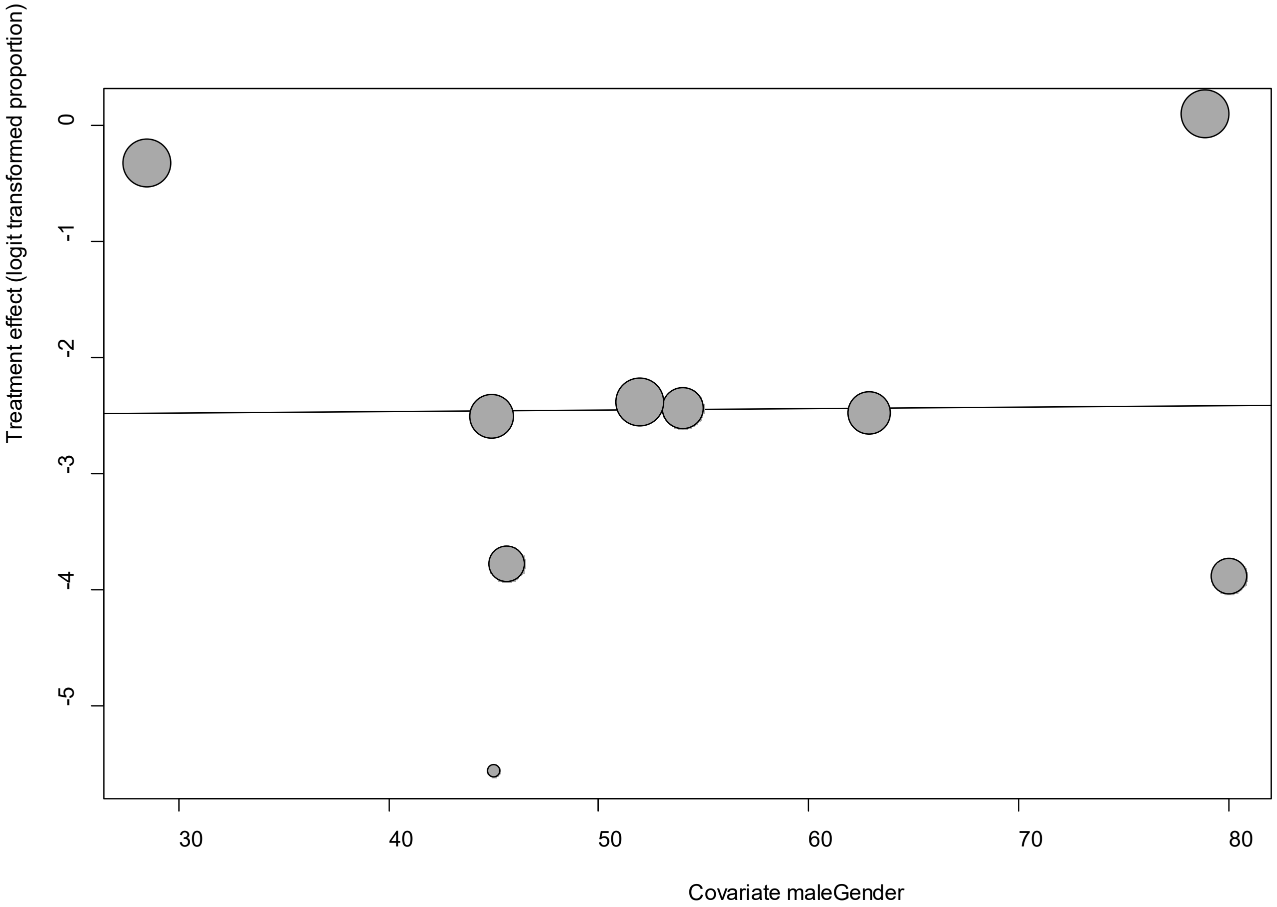


Point sizes are proportional to an inverse of the precision of the estimates.

The regression coefficient was 0.0013 (95% confidence interval:-0.08-0.08).(*P*=0.97).

**eFigure 11e. Funnel Plot of studies reporting on Anorexia**

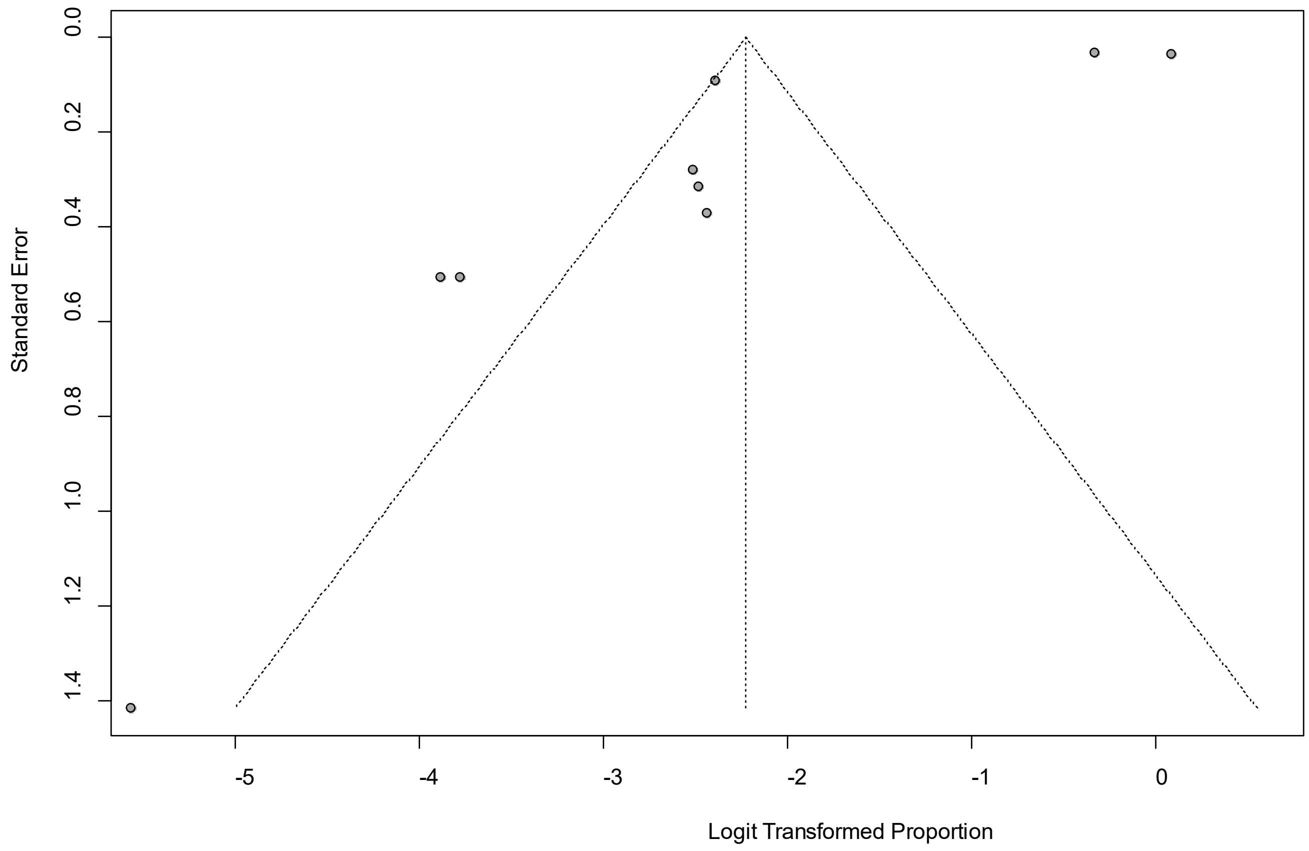


Egger's *P* was 0.036, indicating the presence of publication bias.

### eFigure 12a. Forest Plot on Anosmia

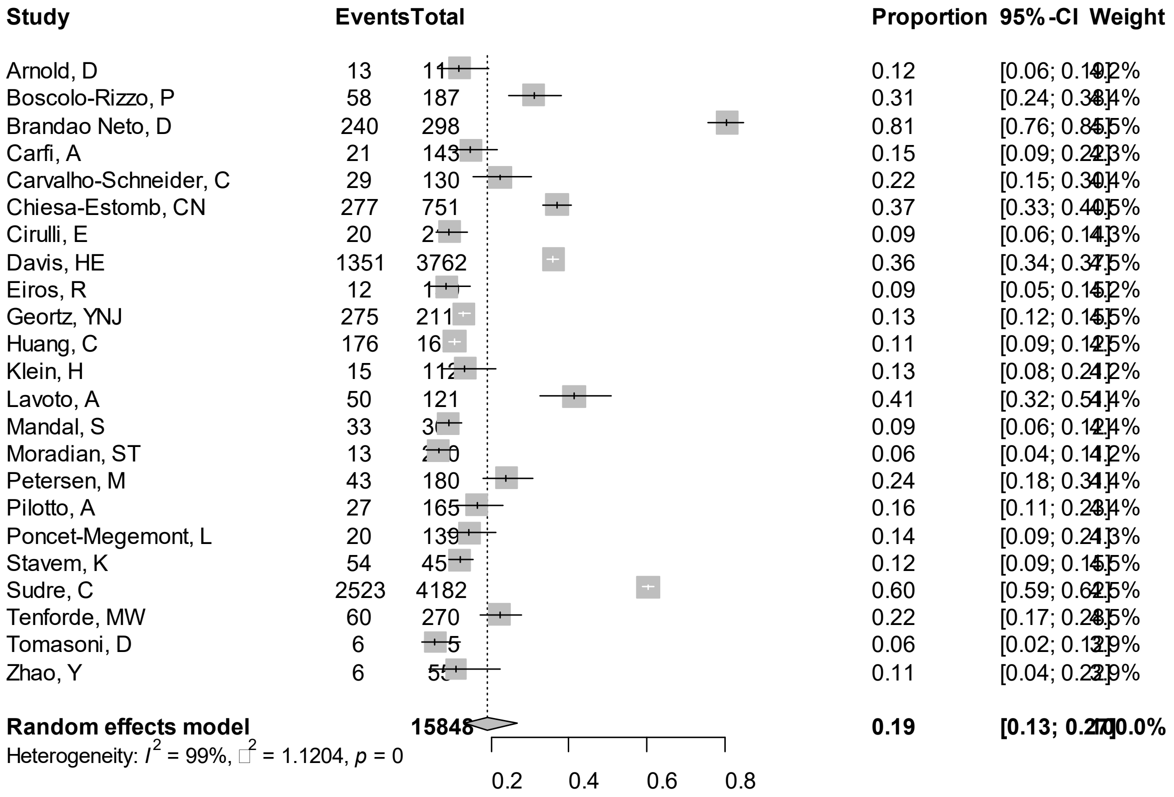


Point sizes are proportional to an inverse of the precision of the estimates and bar correspond to 95% confidence intervals.

**eFigure 12b. Bubble Plots (follow-up period) on Anosmia**

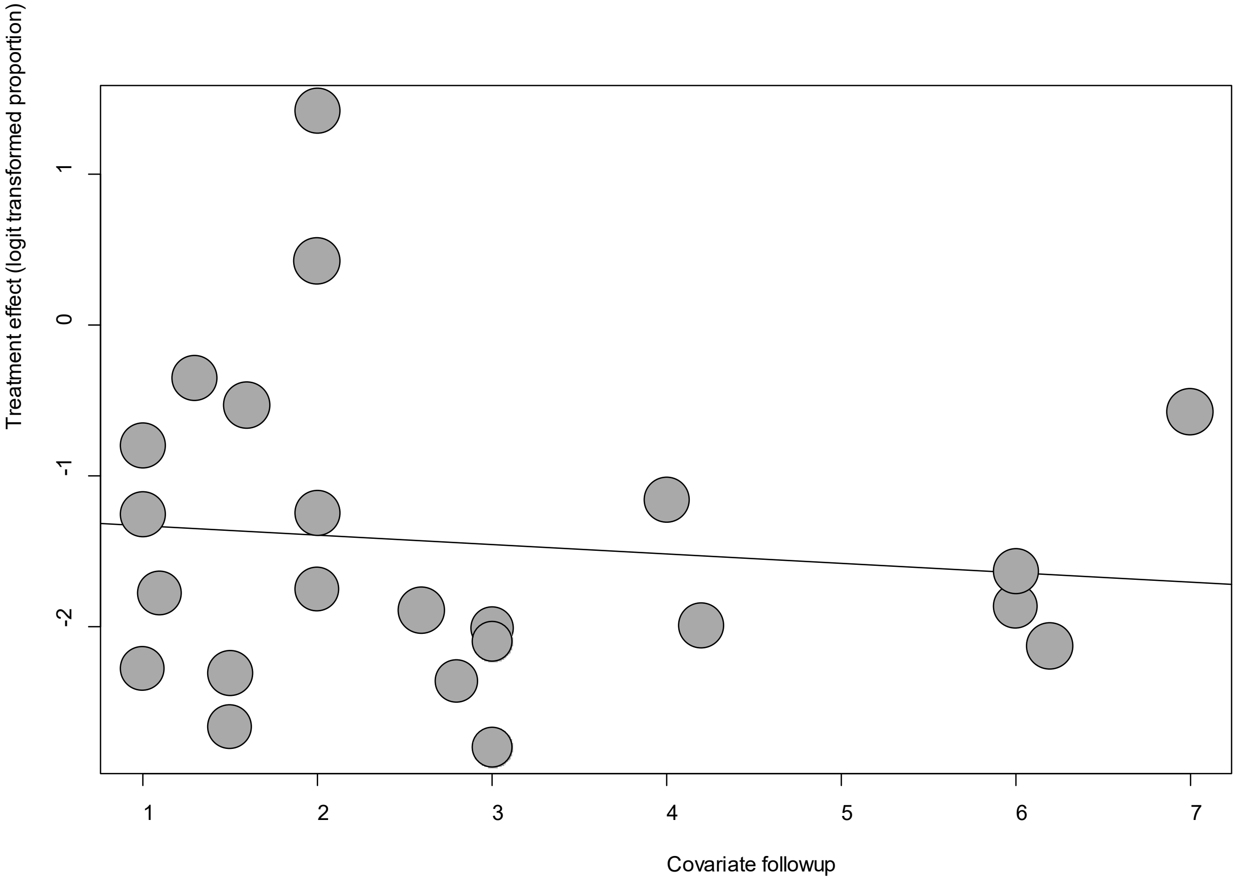


Point sizes are proportional to an inverse of the precision of the estimates.

The regression coefficient was -0.06 (95% confidence interval:-0.34-0.22).(*P*=0.67).

### eFigure 12c. Bubble Plots (age) on Anosmia

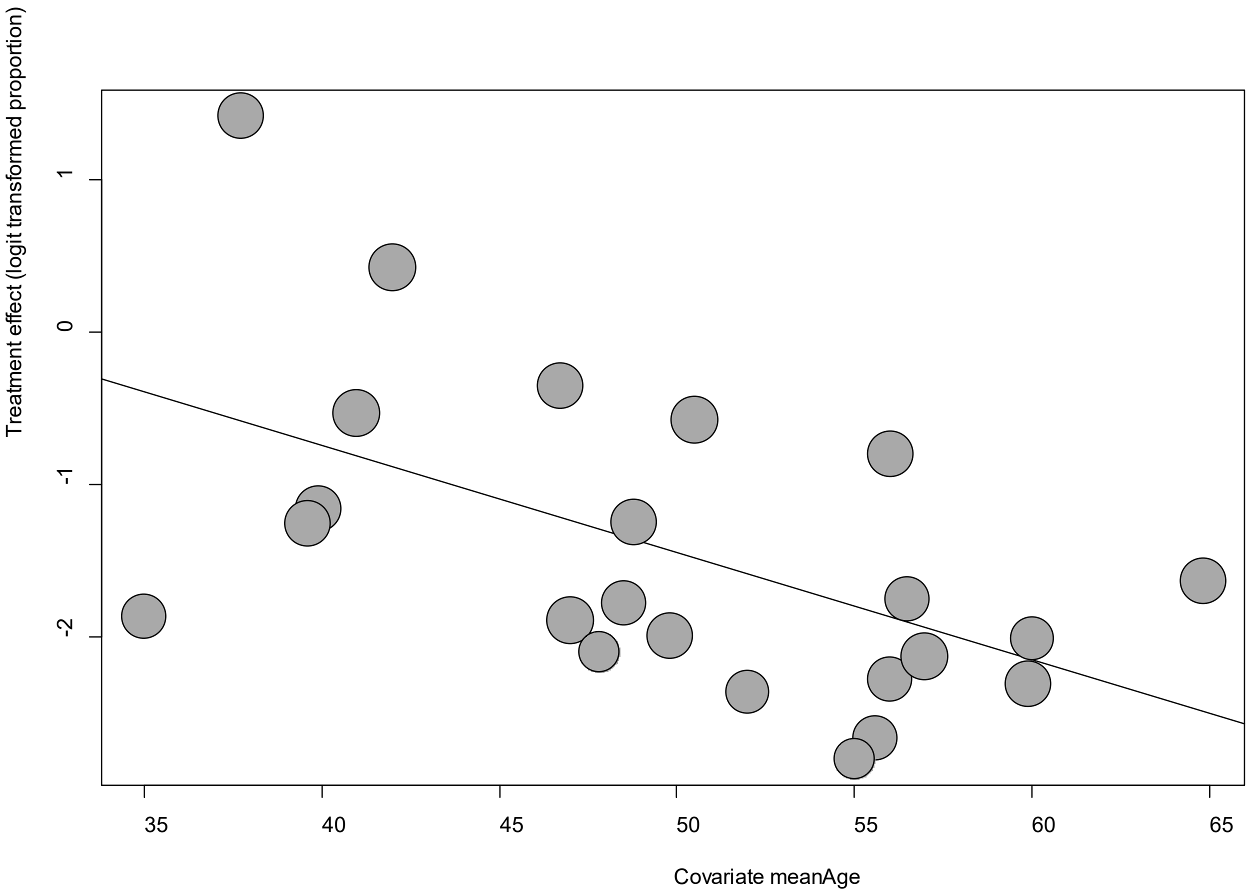


Point sizes are proportional to an inverse of the precision of the estimates.

The regression coefficient was -0.07 (95% confidence interval:-0.11—0.02).(*P*=0.003).

**eFigure 12d. Bubble Plots (sex) on Anosmia**

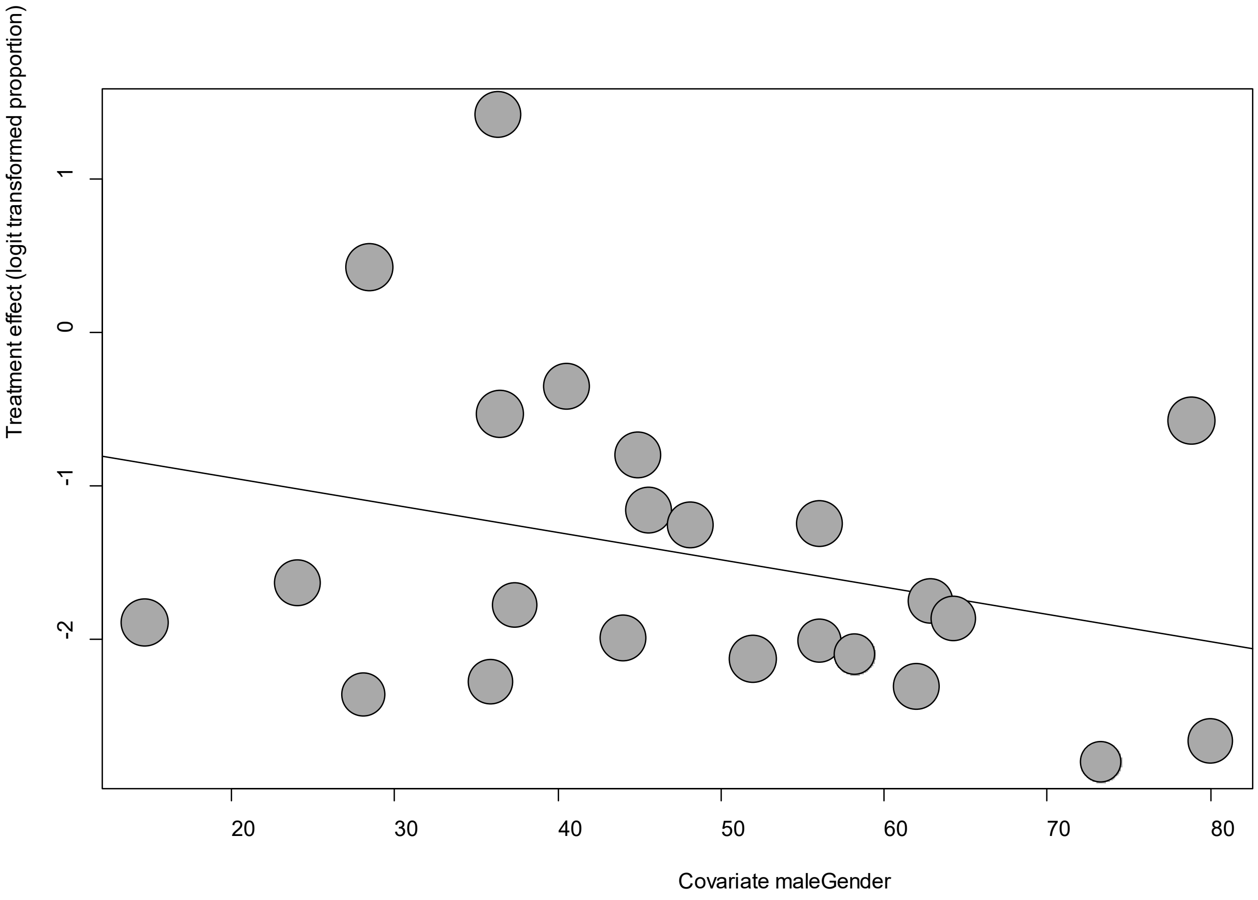


Point sizes are proportional to an inverse of the precision of the estimates.

The regression coefficient was -0.018 (95% confidence interval:-0.05-0.01).(*P*=0.26).

**eFigure 12e. Funnel Plot of studies reporting on Anosmia**

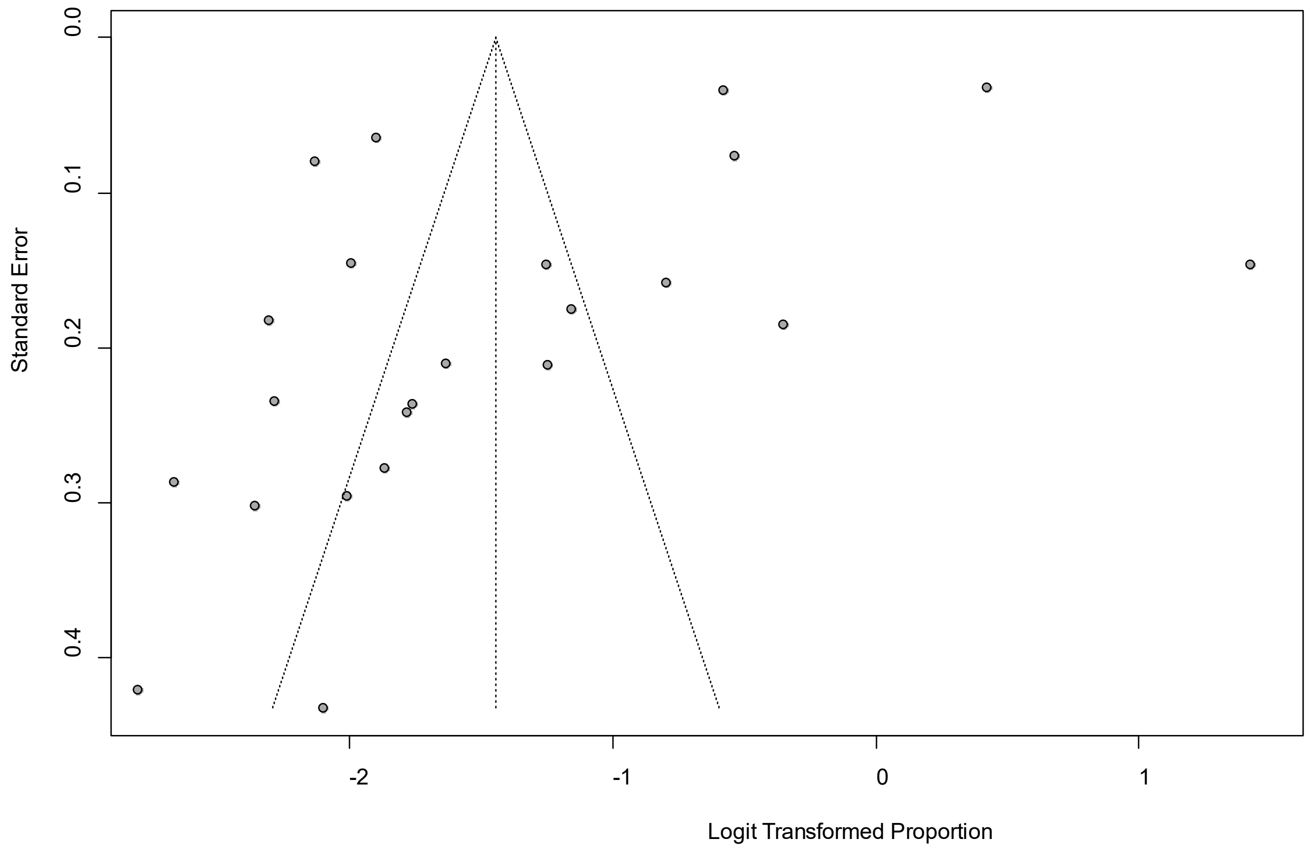


Egger's *P* was 0.008, indicating the presence of publication bias.

### eFigure 13a. Forest Plot on Anxiety

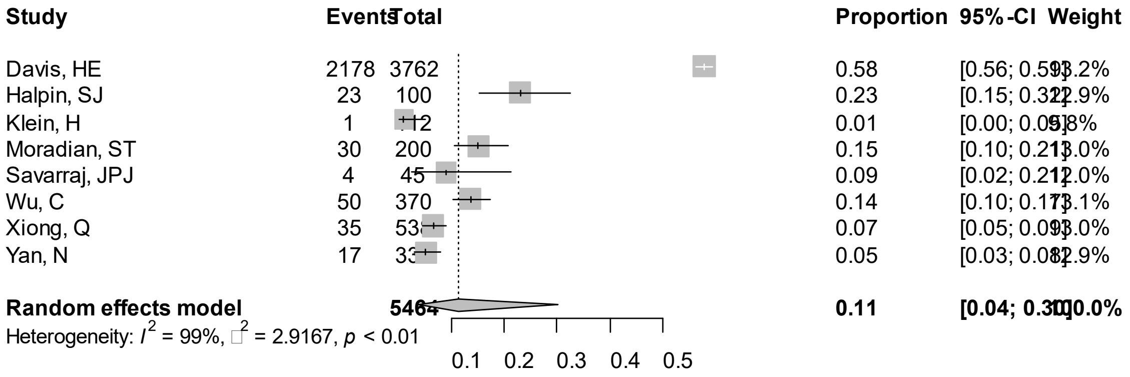


Point sizes are proportional to an inverse of the precision of the estimates and bar correspond to 95% confidence intervals.

### eFigure 13b. Bubble Plots (follow-up period) on Anxiety

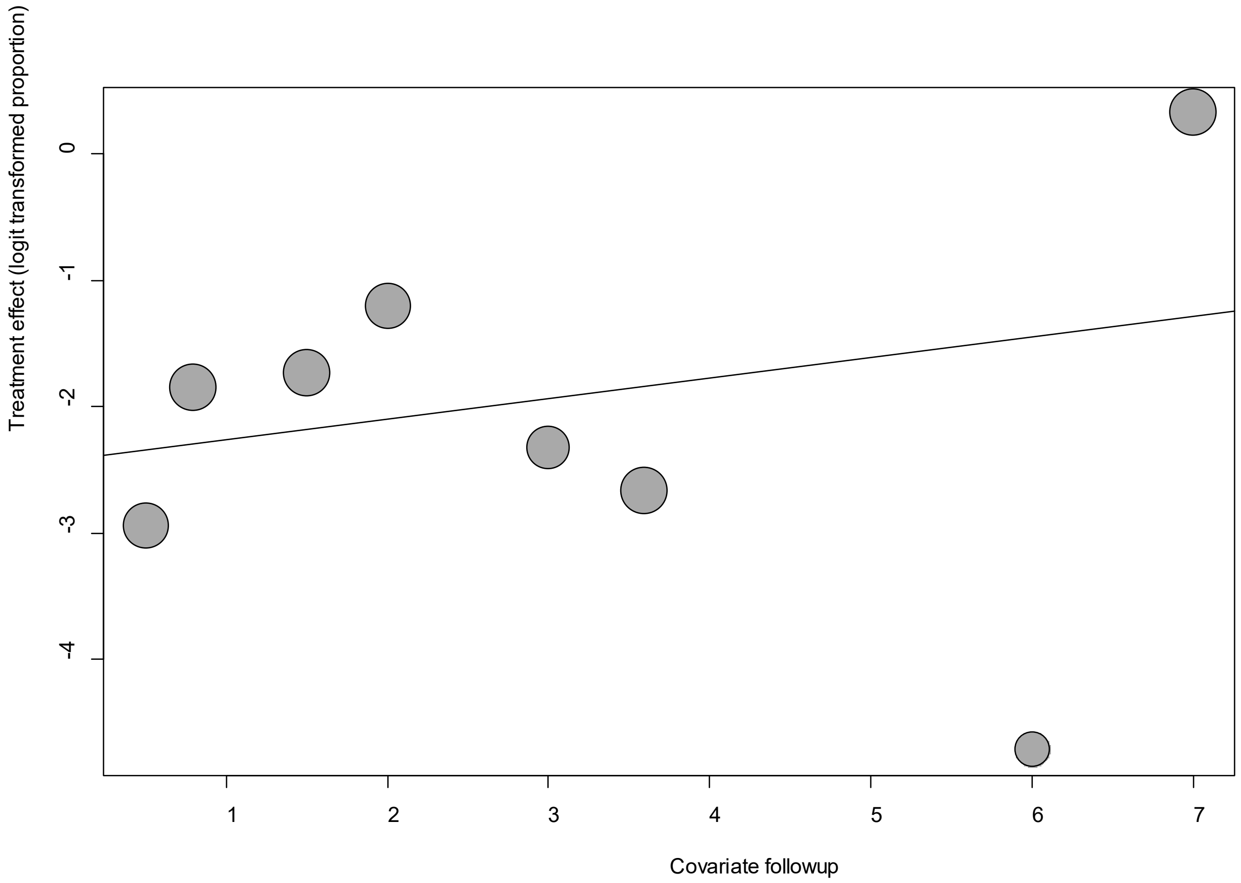


Point sizes are proportional to an inverse of the precision of the estimates.

The regression coefficient was 0.16 (95% confidence interval:-0.17-0.50).(*P*=0.34).

**eFigure 13c. Bubble Plots (age) on Anxiety**

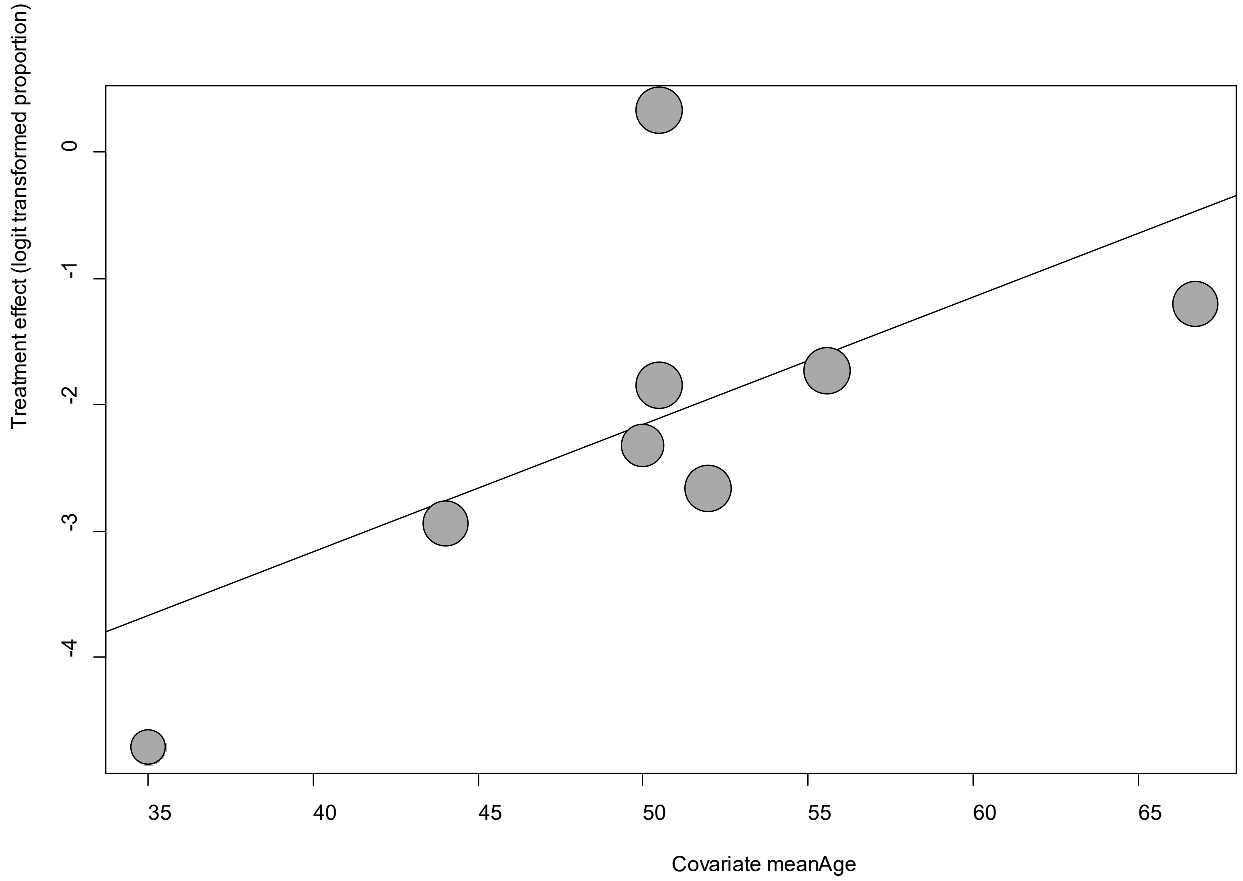


Point sizes are proportional to an inverse of the precision of the estimates.

The regression coefficient was 0.101 (95% confidence interval:-0.06--.26).(*P*=0.21).

**eFigure 13d. Bubble Plots (sex) on Anxiety**

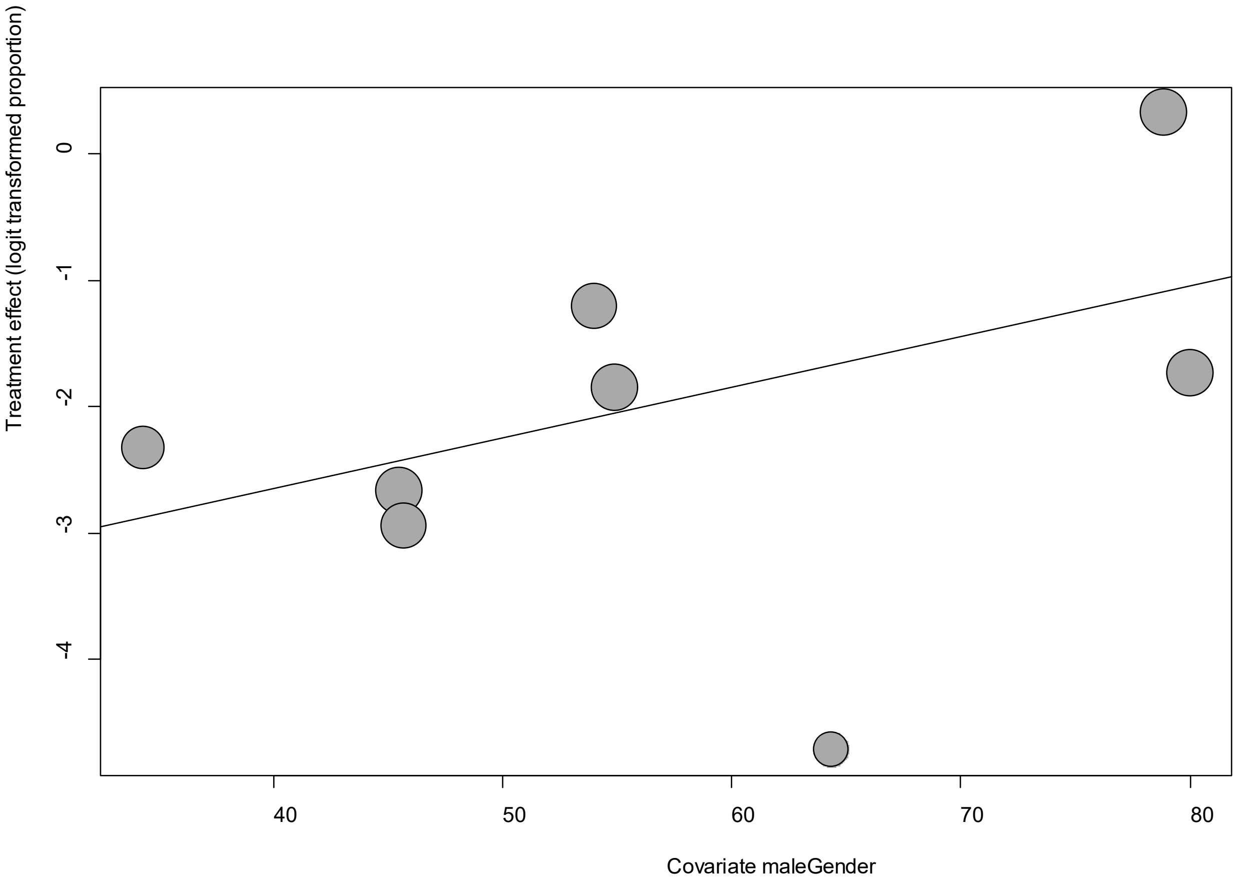


Point sizes are proportional to an inverse of the precision of the estimates.

The regression coefficient was 0.004 (95% confidence interval:-0.007-0.009).(*P*=0.09).

**eFigure 13e. Funnel Plot of studies reporting on Anxiety**

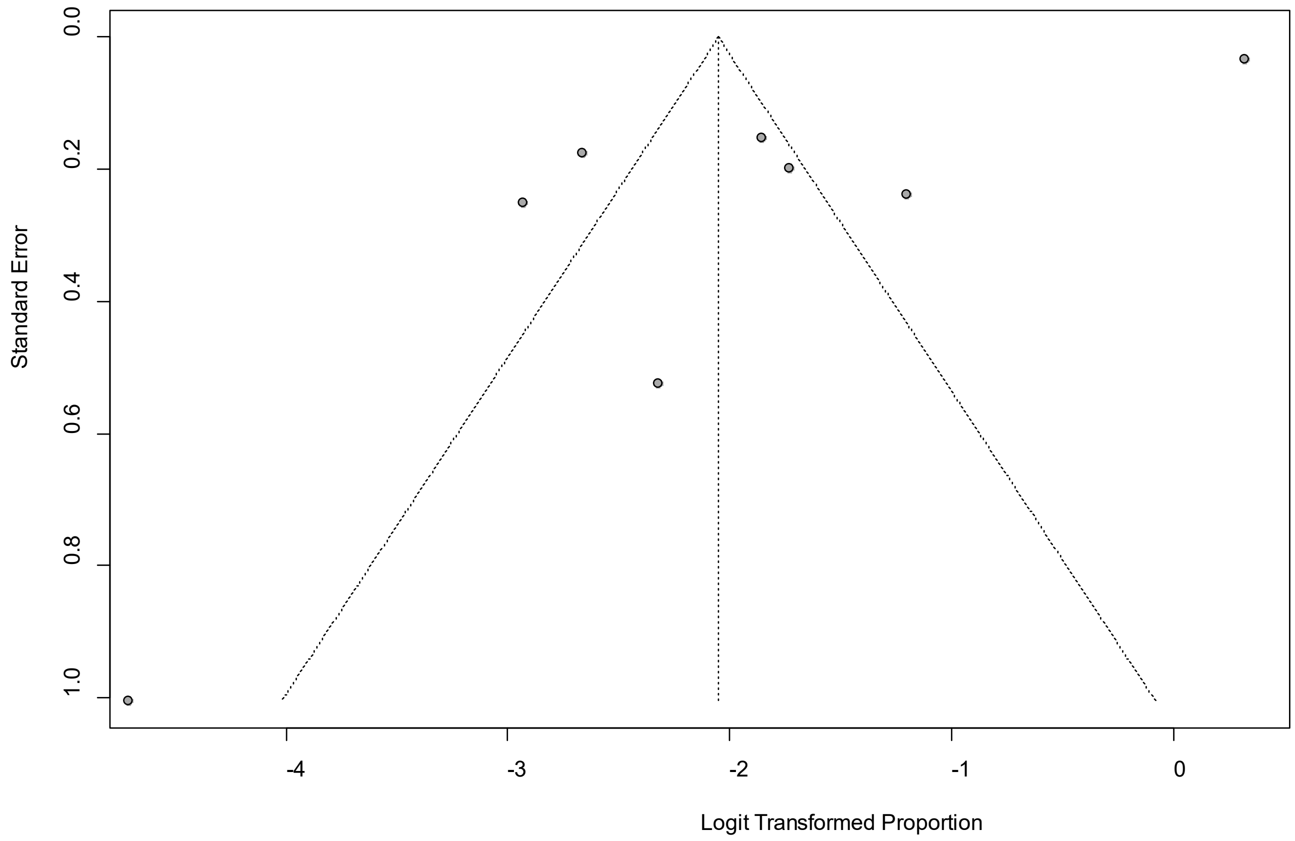


Egger's *P* was 0.004, indicating the presence of publication bias.

**eFigure 14a. Forest Plot on Chills**

| Study | Events | Total | Proportion | 95%-CI | Weight |
|---|---|---|---|---|---|
| Davis, HE | 1516 | 3762 | 0.40 | [0.39; 0.42] | 17.6% |
| Moradian, ST | 1 | [illegible]00 | 0.00 | [0.00; 0.03] | 14.1% |
| Petersen, M | 9 | 180 | 0.05 | [0.02; 0.09] | 17.1% |
| Stavem, K | 4 | [illegible]51 | 0.01 | [0.00; 0.02] | 16.6% |
| Tenforde, MW | 11 | 27[illegible] | 0.04 | [0.02; 0.07] | 17.2% |
| Xiong, Q | 25 | 538 | 0.05 | [0.03; 0.07] | 17.4% |
| **Random effects model** | | **54[illegible]** | **0.04** | **[0.01; 0.18]** | **100.0%** |

Heterogeneity: $I^2 = 99\%$, $\tau^2 = 4.0474$, $p < 0.01$

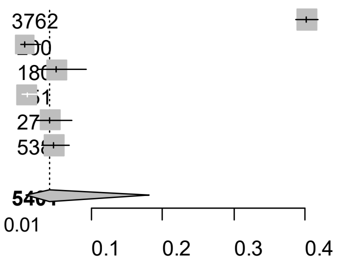


Point sizes are proportional to an inverse of the precision of the estimates and bar correspond to 95% confidence intervals.

**eFigure 14b. Bubble Plots (follow-up period) on Chills**

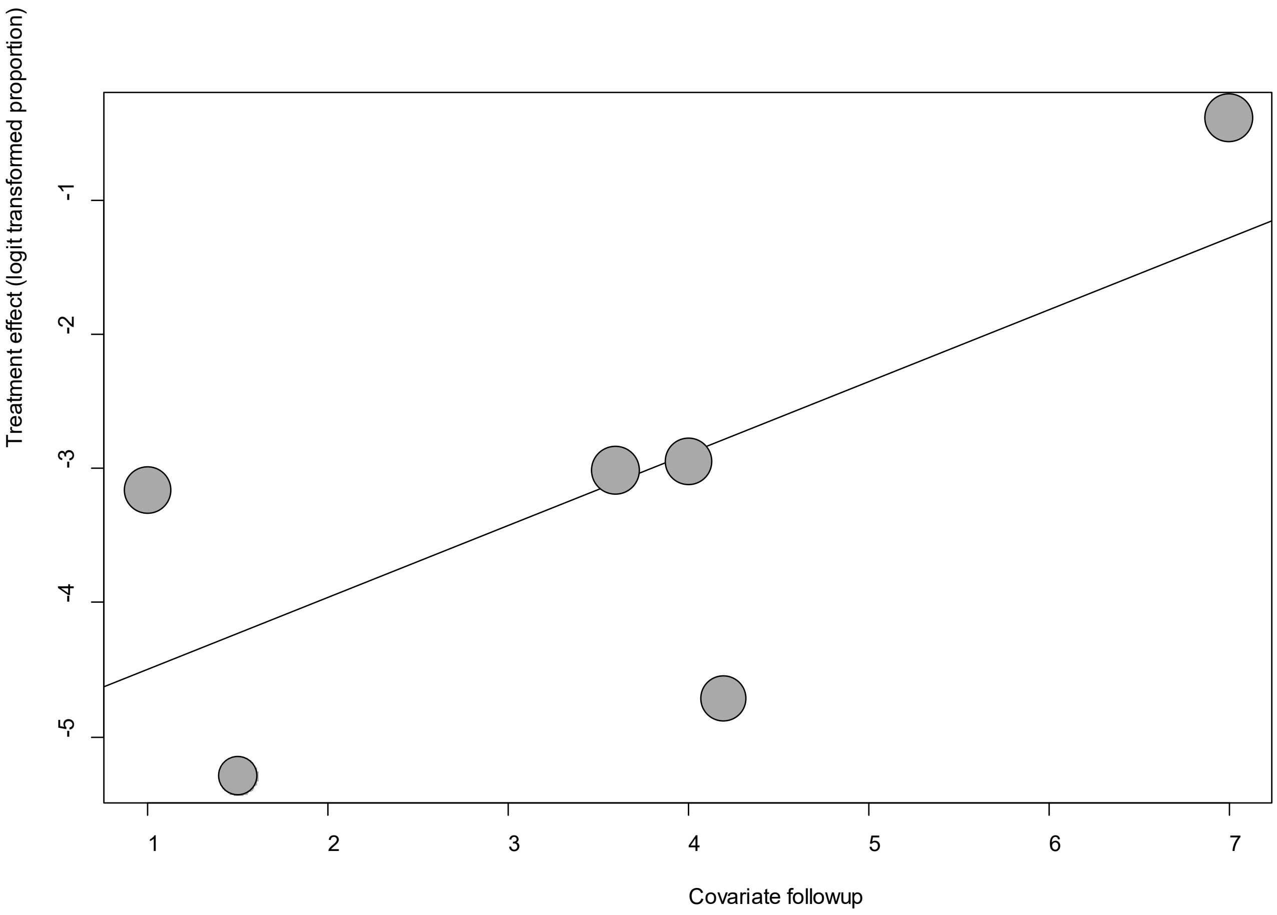


Point sizes are proportional to an inverse of the precision of the estimates.

The regression coefficient was 0.54 (95% confidence interval:0.07-1.00).(*P*=0.02).

**eFigure 14c. Bubble Plots (age) on Chills**

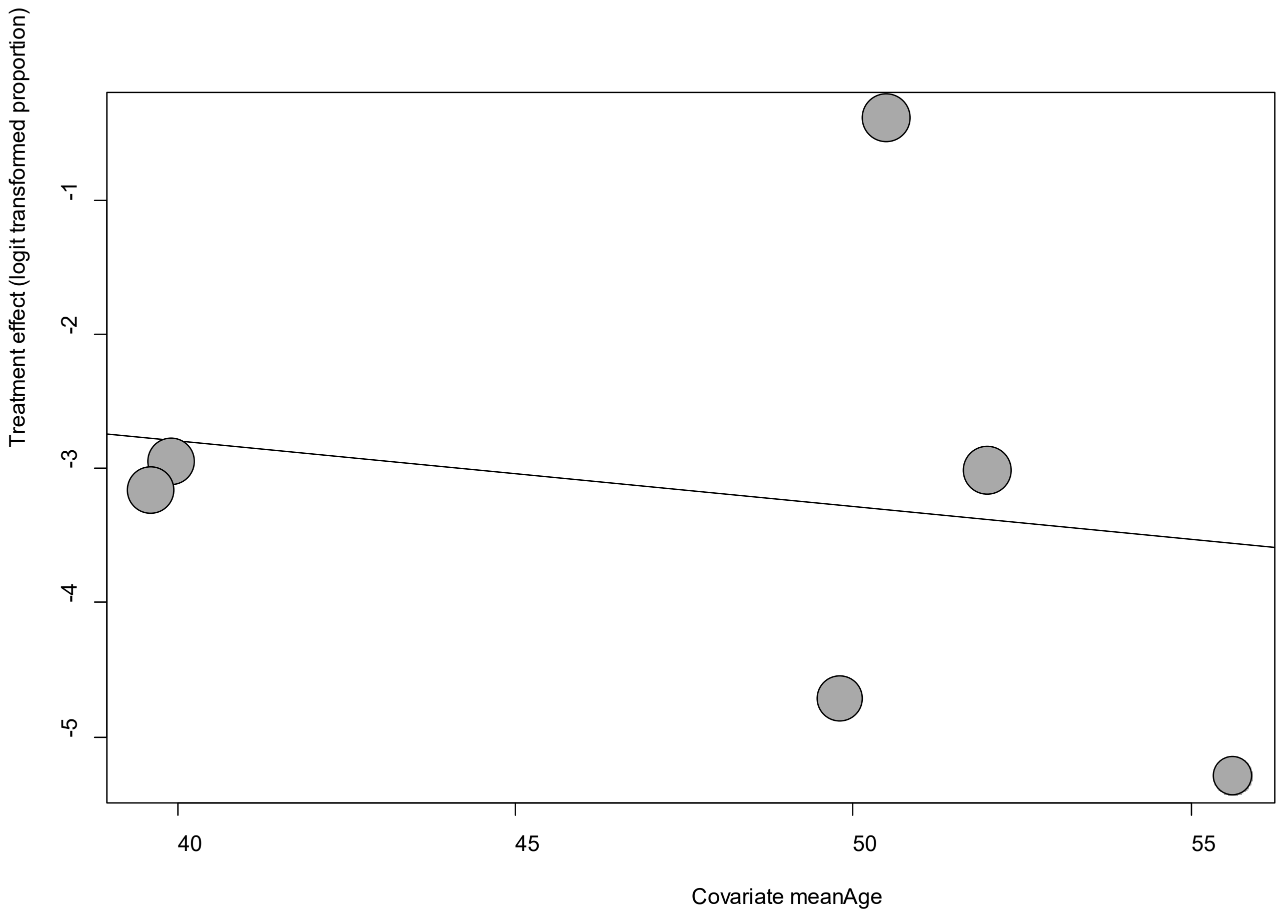


Point sizes are proportional to an inverse of the precision of the estimates.

The regression coefficient was -0.05 (95% confidence interval:-0.33-0.23).(*P*=0.73).

**eFigure 14d. Bubble Plots (sex) on Chills**

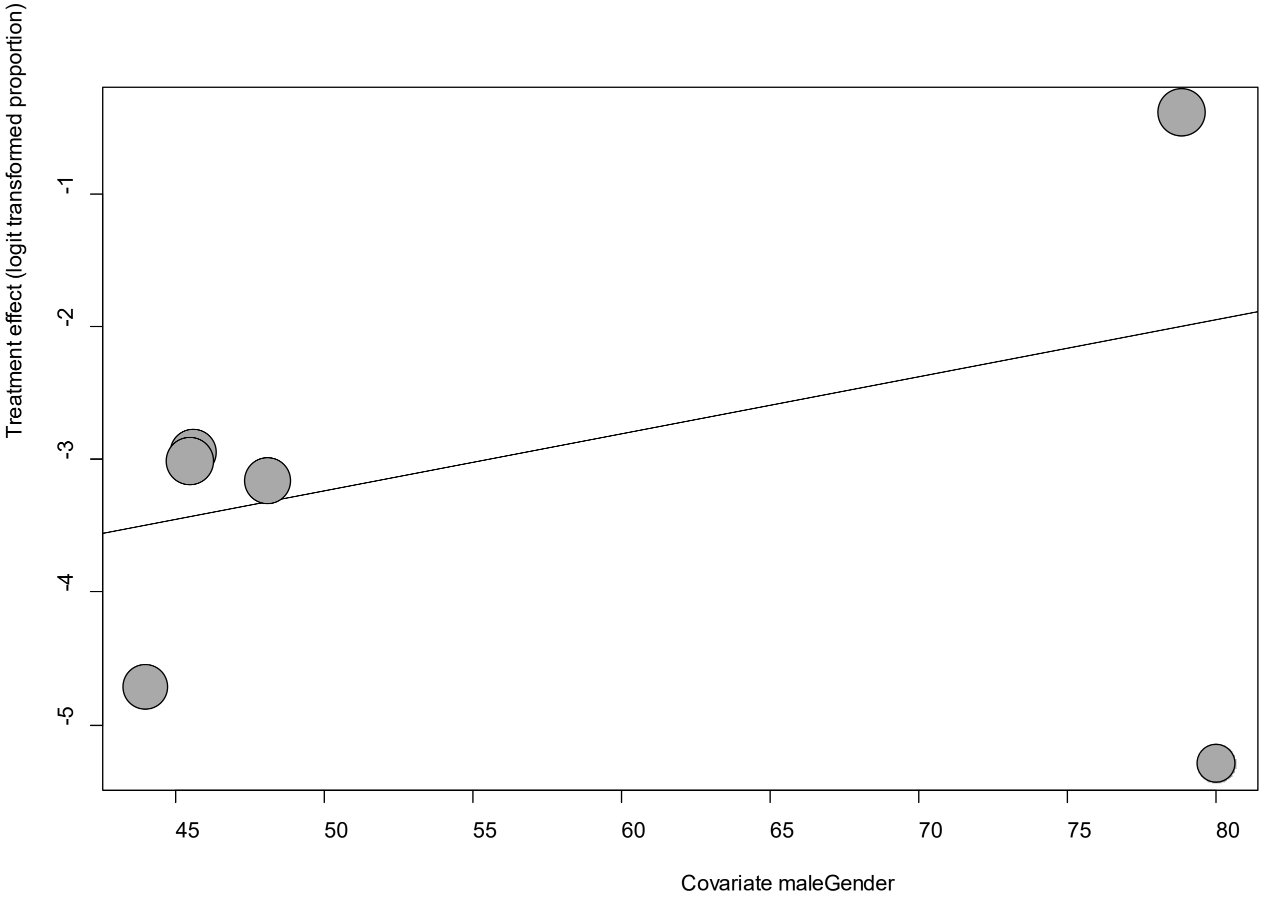


Point sizes are proportional to an inverse of the precision of the estimates.

The regression coefficient was 0.04 (95% confidence interval:-0.01-0.1).(*P*=0.13).

**eFigure 14e. Funnel Plot of studies reporting on Chills**

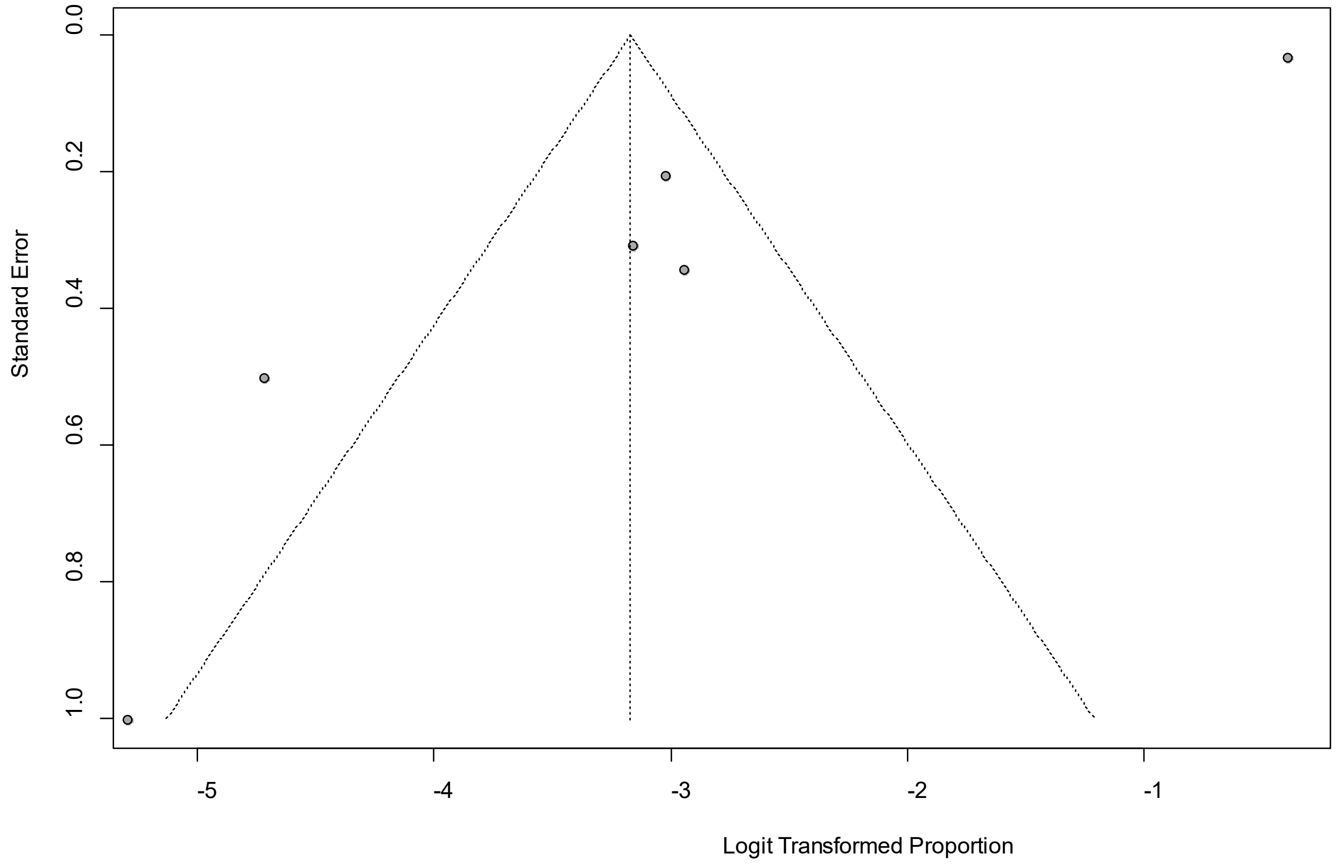


Egger's *P* was 0.005, indicating the presence of publication bias.

### eFigure 15a. Forest Plot on Confusion

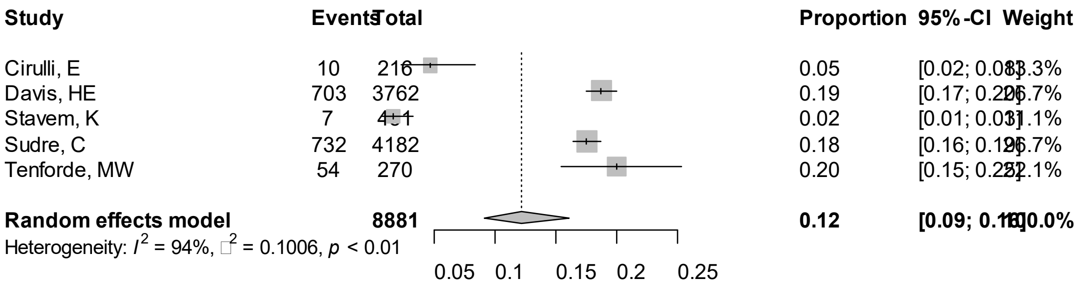


Point sizes are proportional to an inverse of the precision of the estimates.

**eFigure 15b. Bubble Plots (follow-up period) on Confusion**

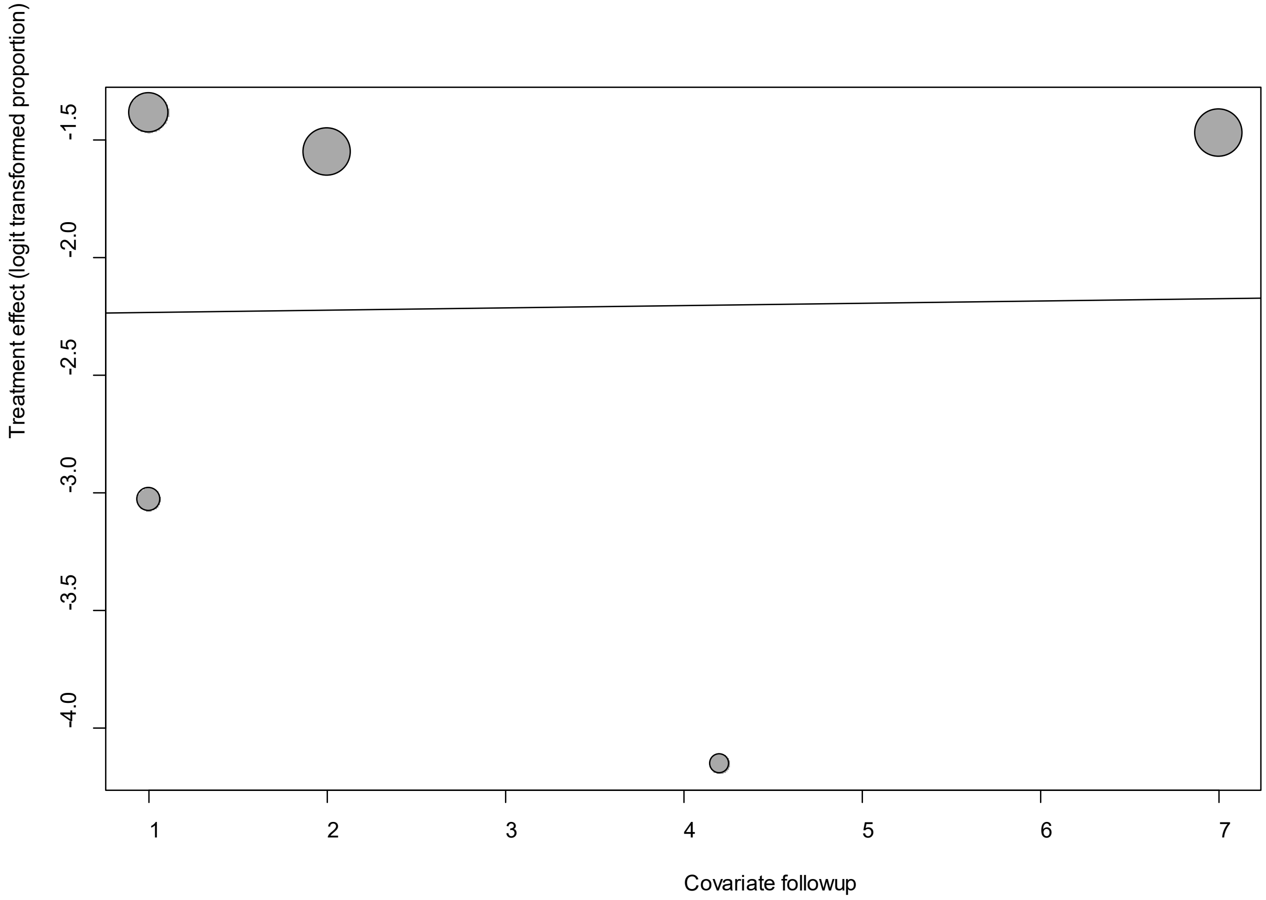


Point sizes are proportional to an inverse of the precision of the estimates.

The regression coefficient was 0.01 (95% confidence interval:-0.27-0.29).(*P*=0.94).

### eFigure 15c. Bubble Plots (age) on Confusion

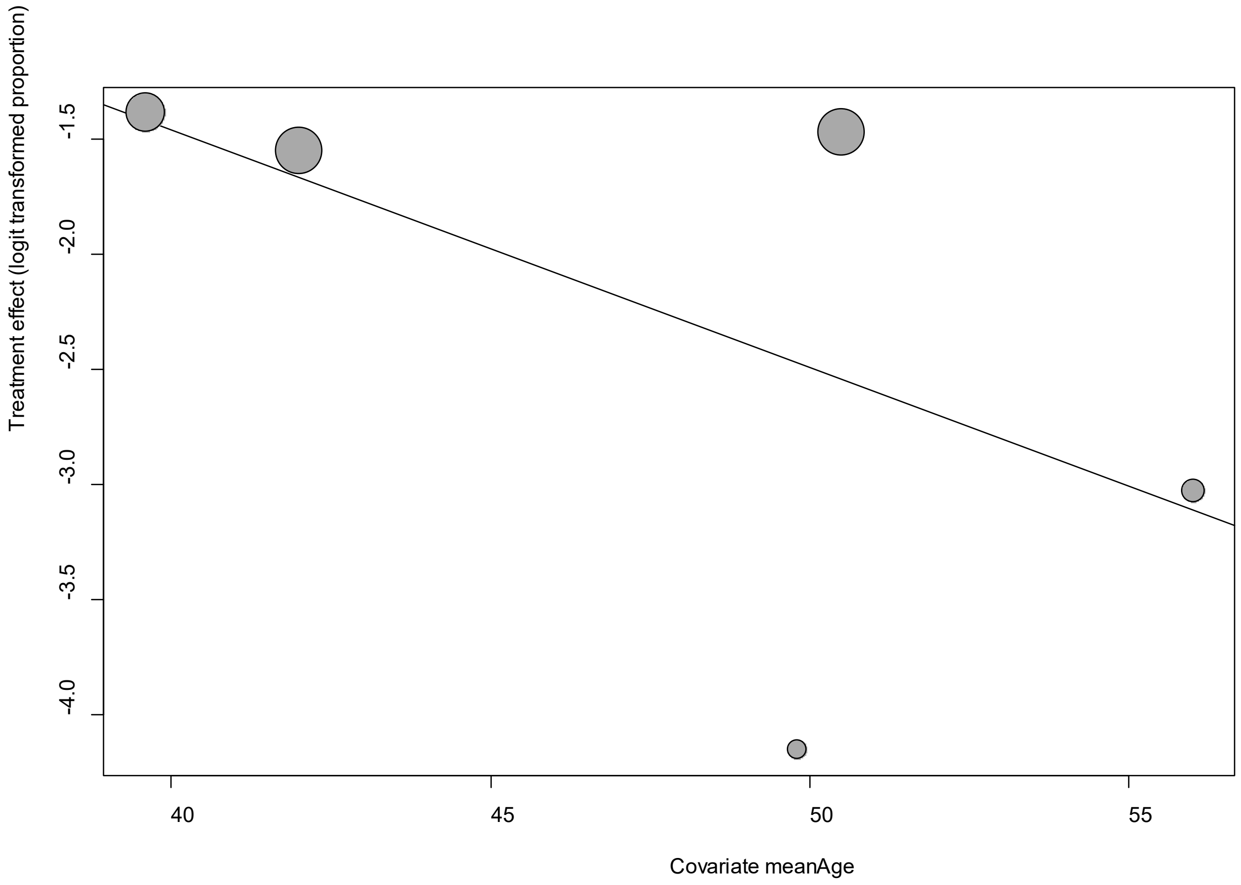


Point sizes are proportional to an inverse of the precision of the estimates.

The regression coefficient was -0.10 (95% confidence interval:-0.21-0.006).(*P*=0.05).

**eFigure 15d. Bubble Plots (sex) on Confusion**

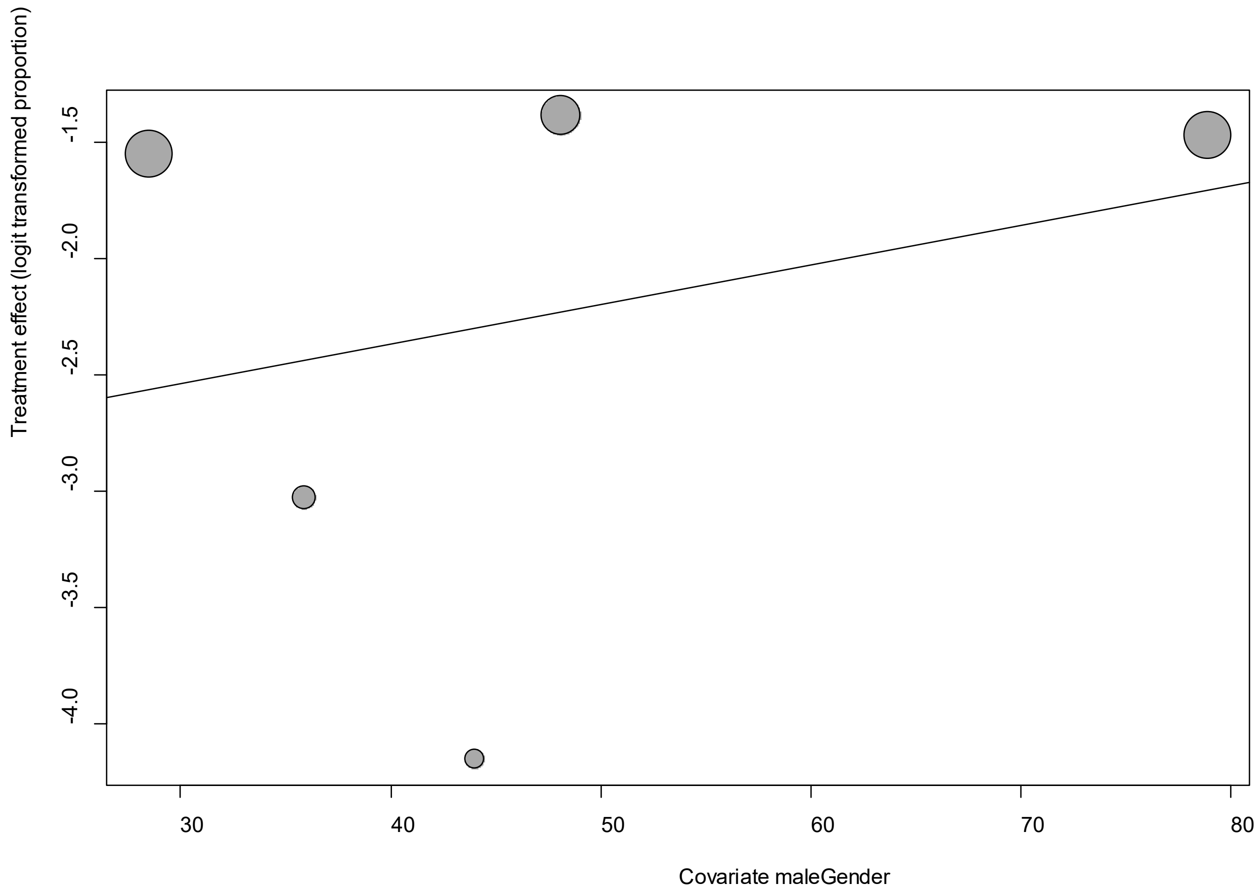


Point sizes are proportional to an inverse of the precision of the estimates.

The regression coefficient was 0.02 (95% confidence interval:-0.03-0.06).(*P*=0.44).

**eFigure 15e. Funnel Plot of studies reporting on Confusion**

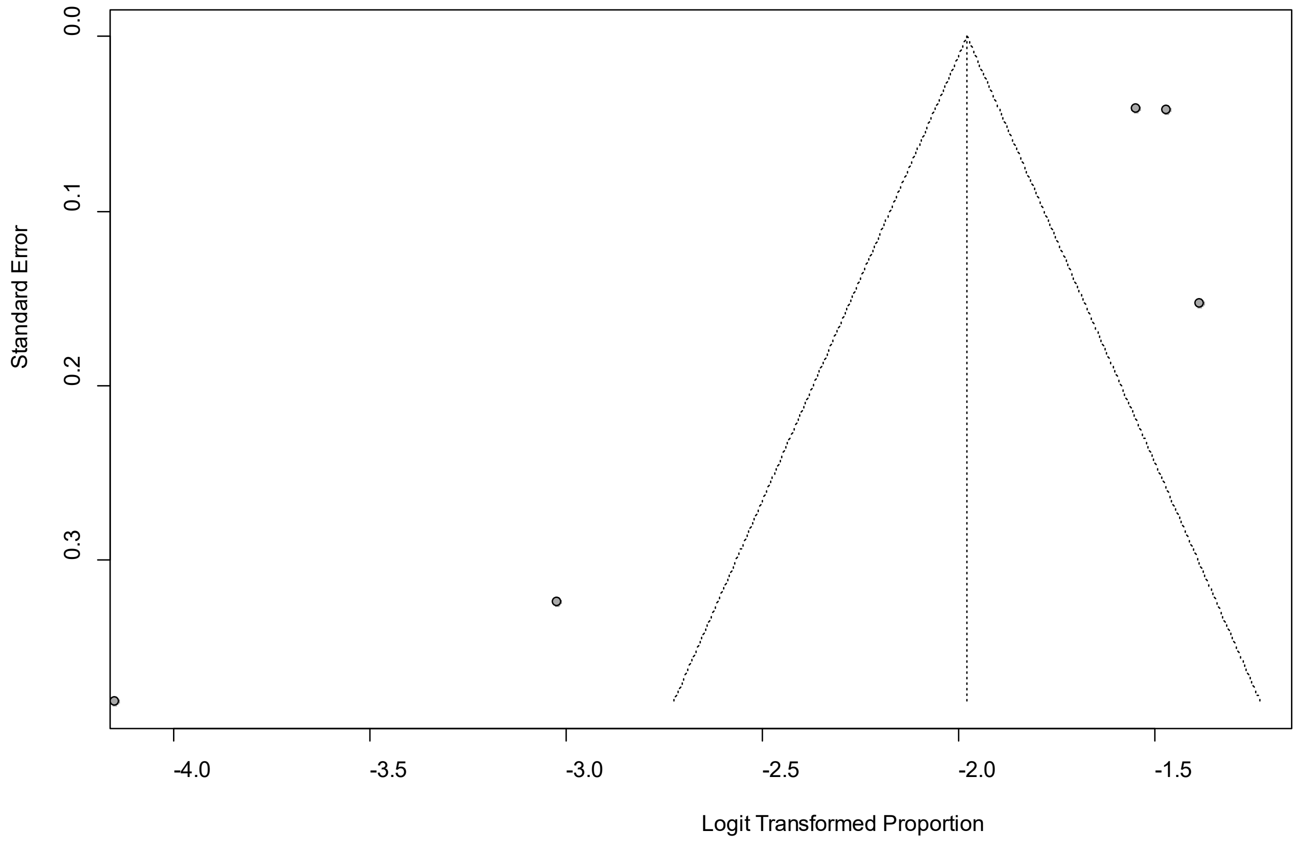


Egger's *P* was 0.12, indicating the presence of publication bias.

### eFigure 16a. Forest Plot on Cough

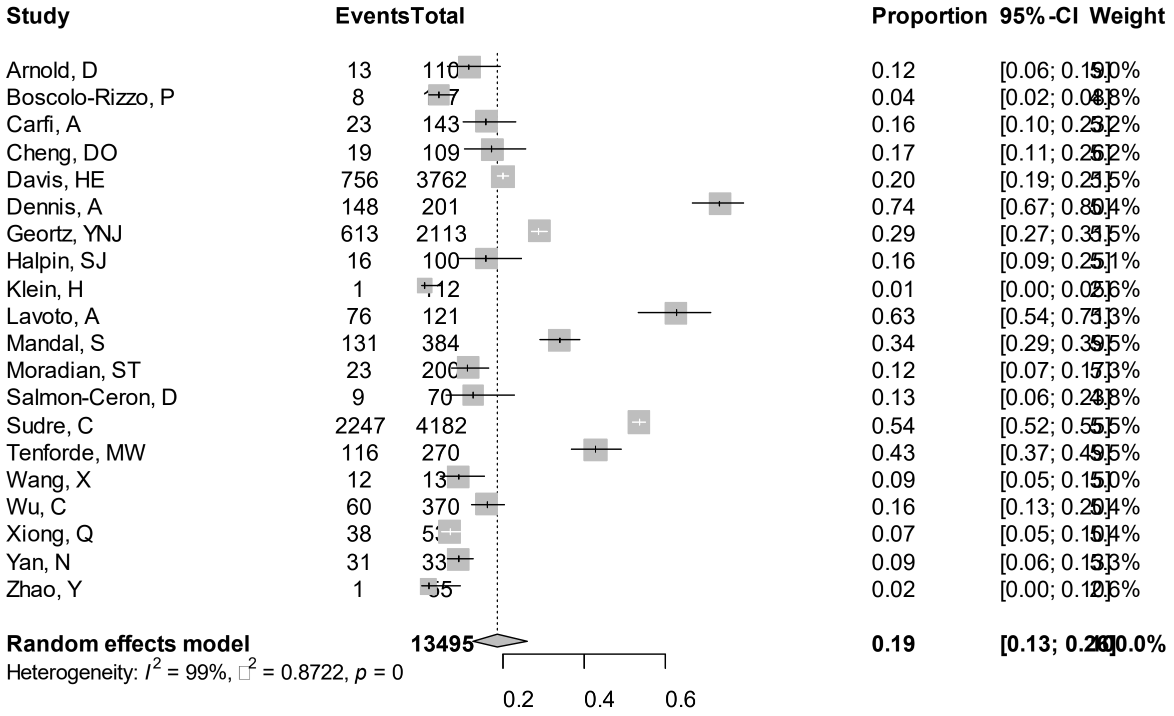


Point sizes are proportional to an inverse of the precision of the estimates.

**eFigure 16b. Bubble Plots (follow-up period) on Cough**

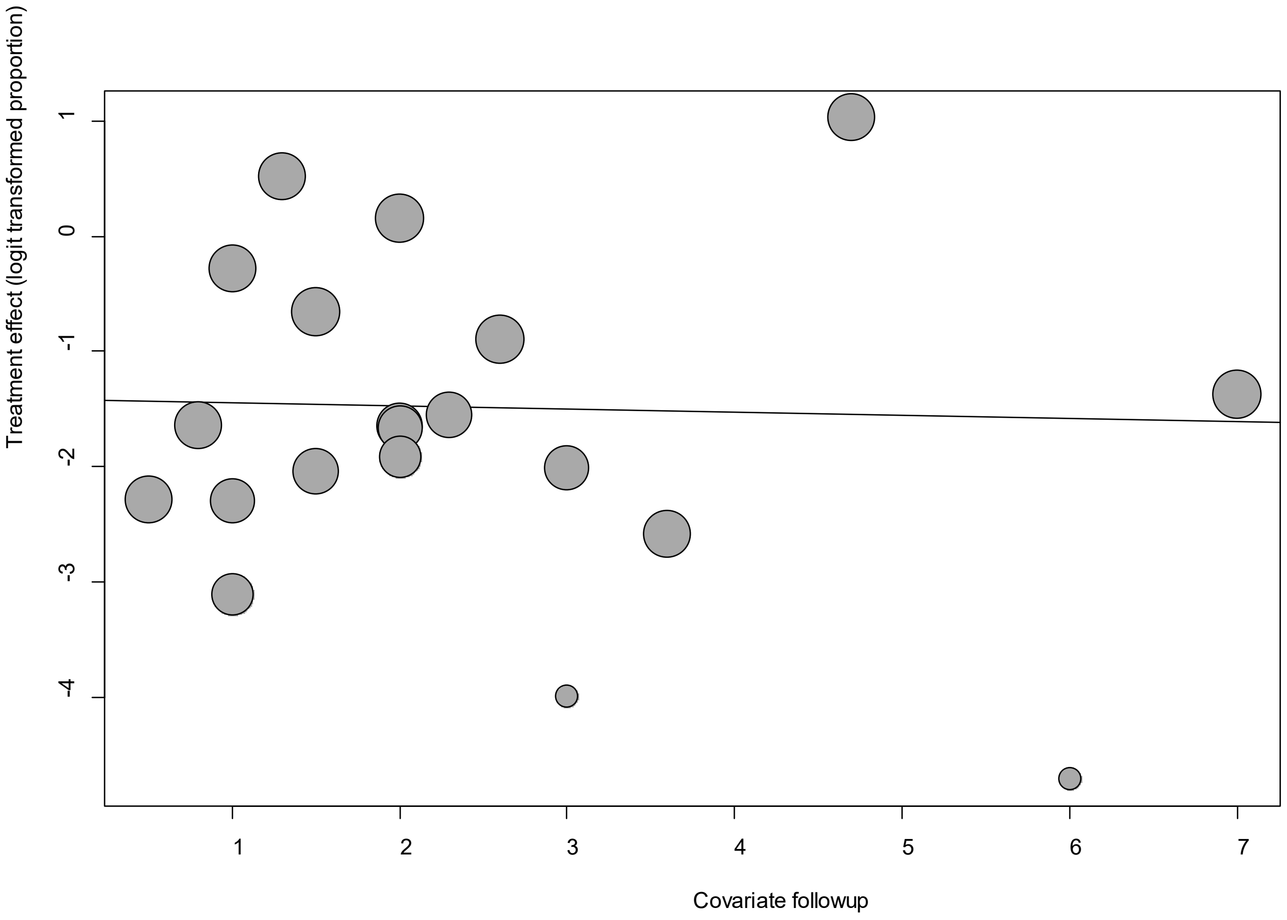


Point sizes are proportional to an inverse of the precision of the estimates.

The regression coefficient was -0.027 (95% confidence interval:-0.30-0.24).(*P*=0.84).

**eFigure 16c. Bubble Plots (age) on Cough**

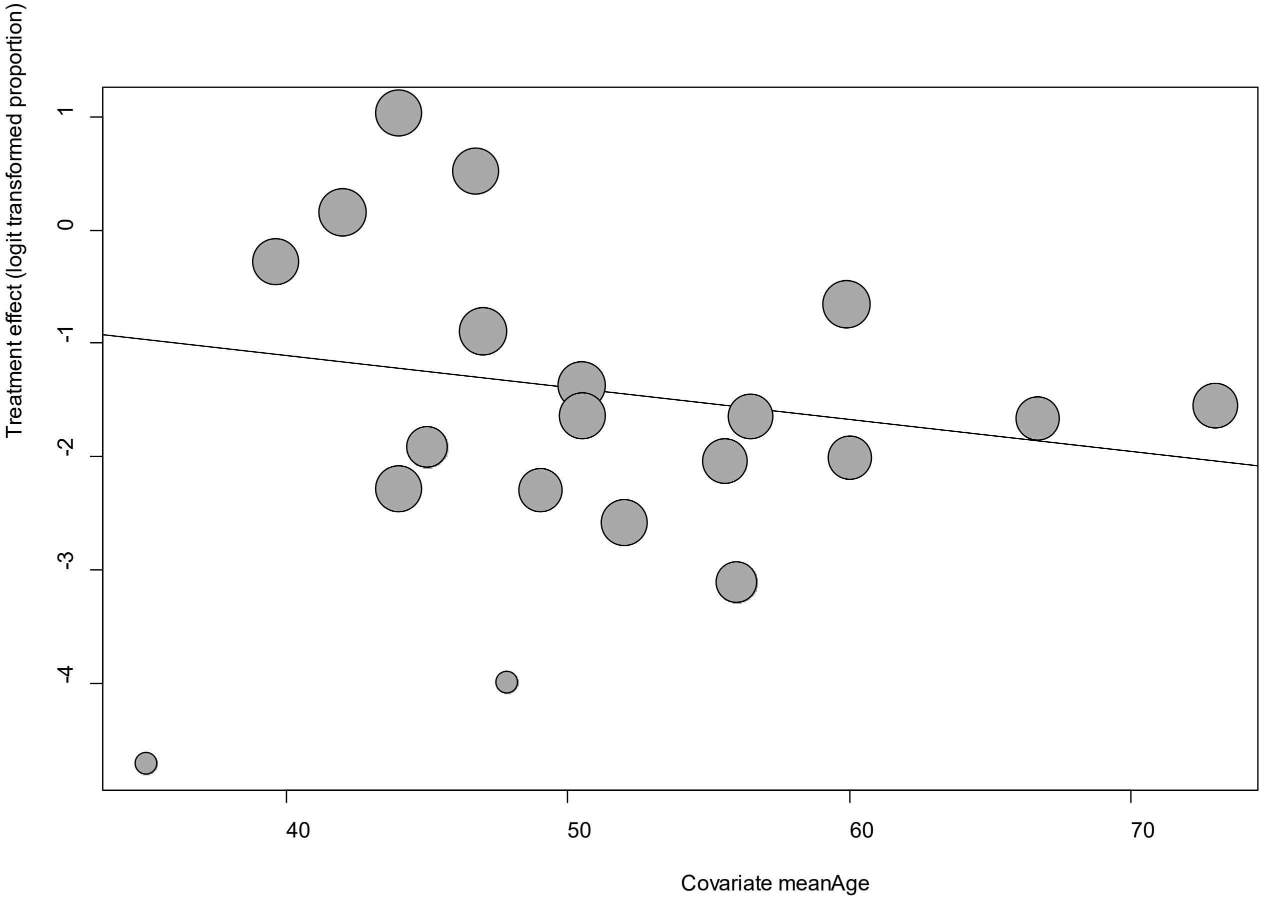


Point sizes are proportional to an inverse of the precision of the estimates.

The regression coefficient was -0.028 (95% confidence interval:-0.07-0.01).(*P*=0.16).

**eFigure 16d. Bubble Plots (sex) on Cough**

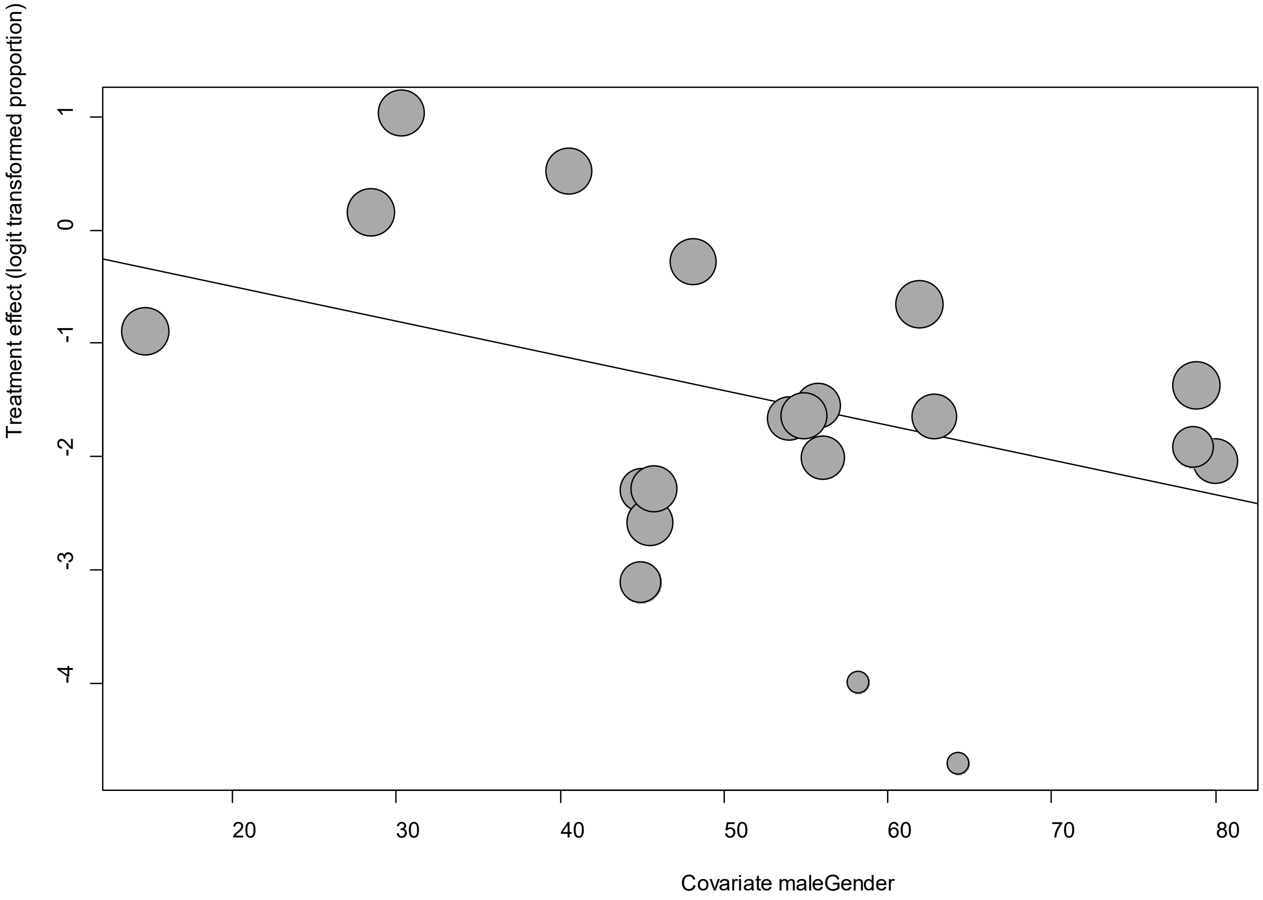


Point sizes are proportional to an inverse of the precision of the estimates.

The regression coefficient was -0.03 (95% confidence interval:-0.06—0.006).(*P*=0.02).

### eFigure 16e. Funnel Plot of studies reporting on Cough

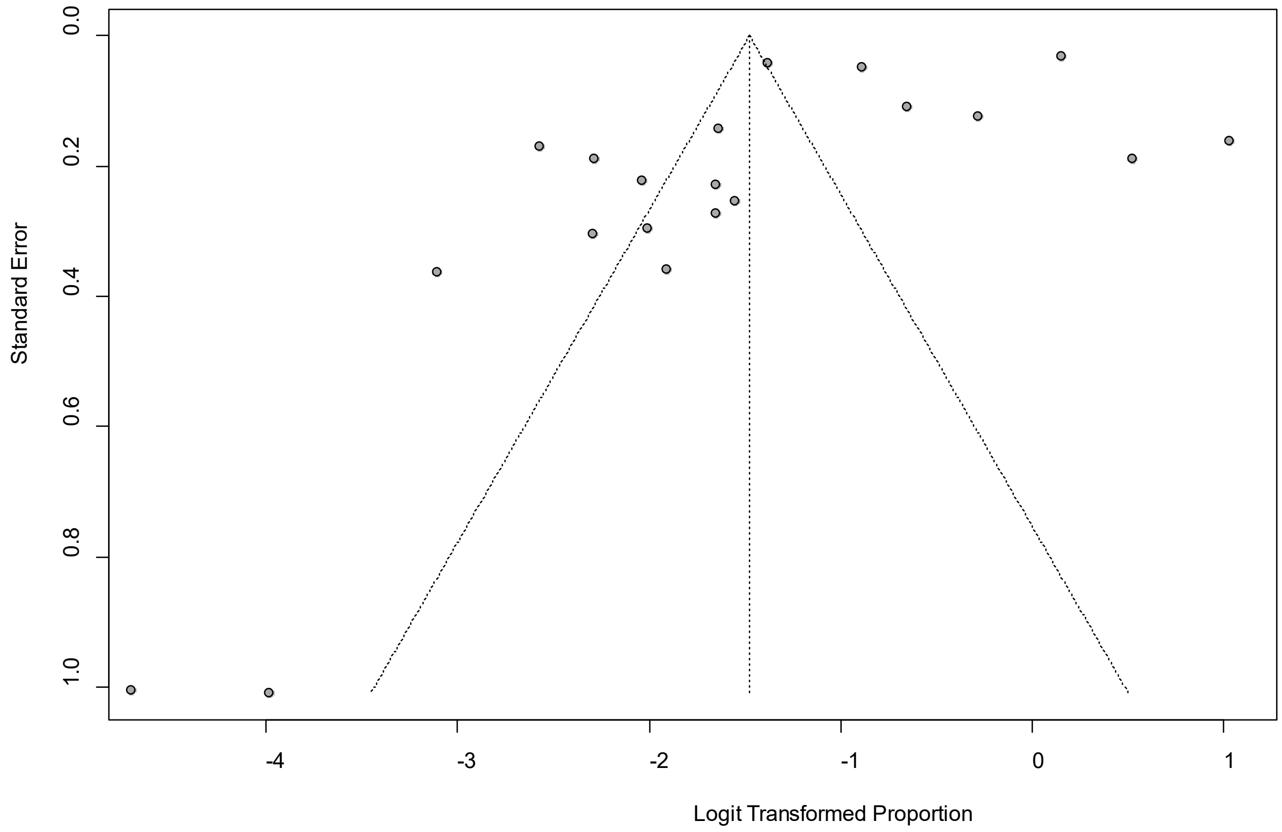


Egger's *P* was 0.06, indicating the presence of publication bias.

**eFigure 17a. Forest Plot on Depression**

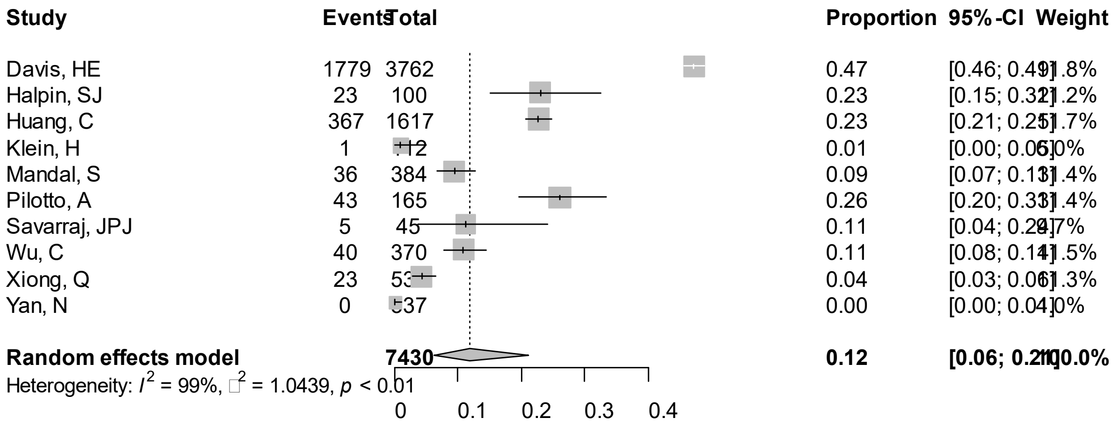


Point sizes are proportional to an inverse of the precision of the estimates.

**eFigure 17b. Bubble Plots (follow-up period) on Depression**

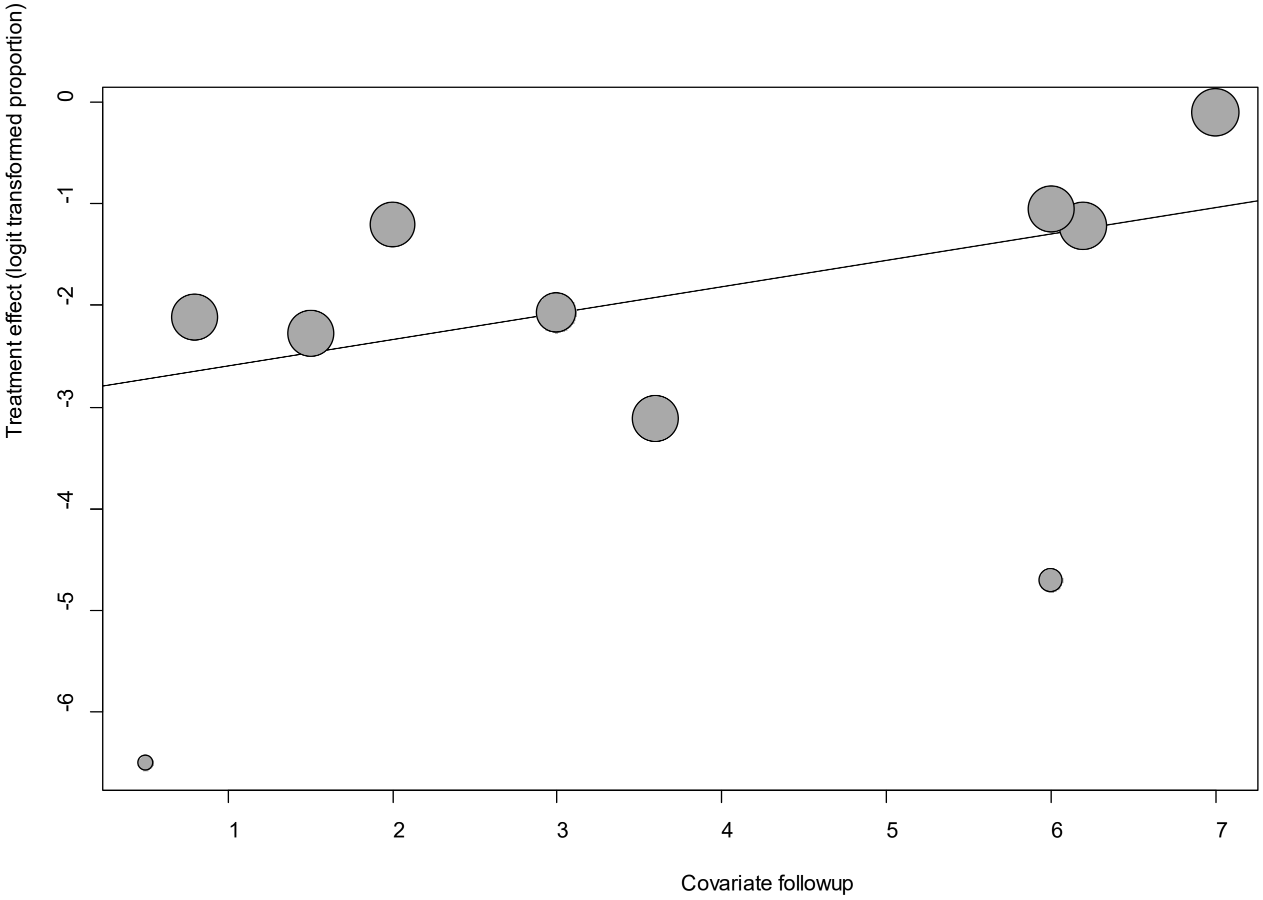


Point sizes are proportional to an inverse of the precision of the estimates.

The regression coefficient was 0.26 (95% confidence interval:0.05-0.47).(*P*=0.015).

**eFigure 17c. Bubble Plots (age) on Depression**

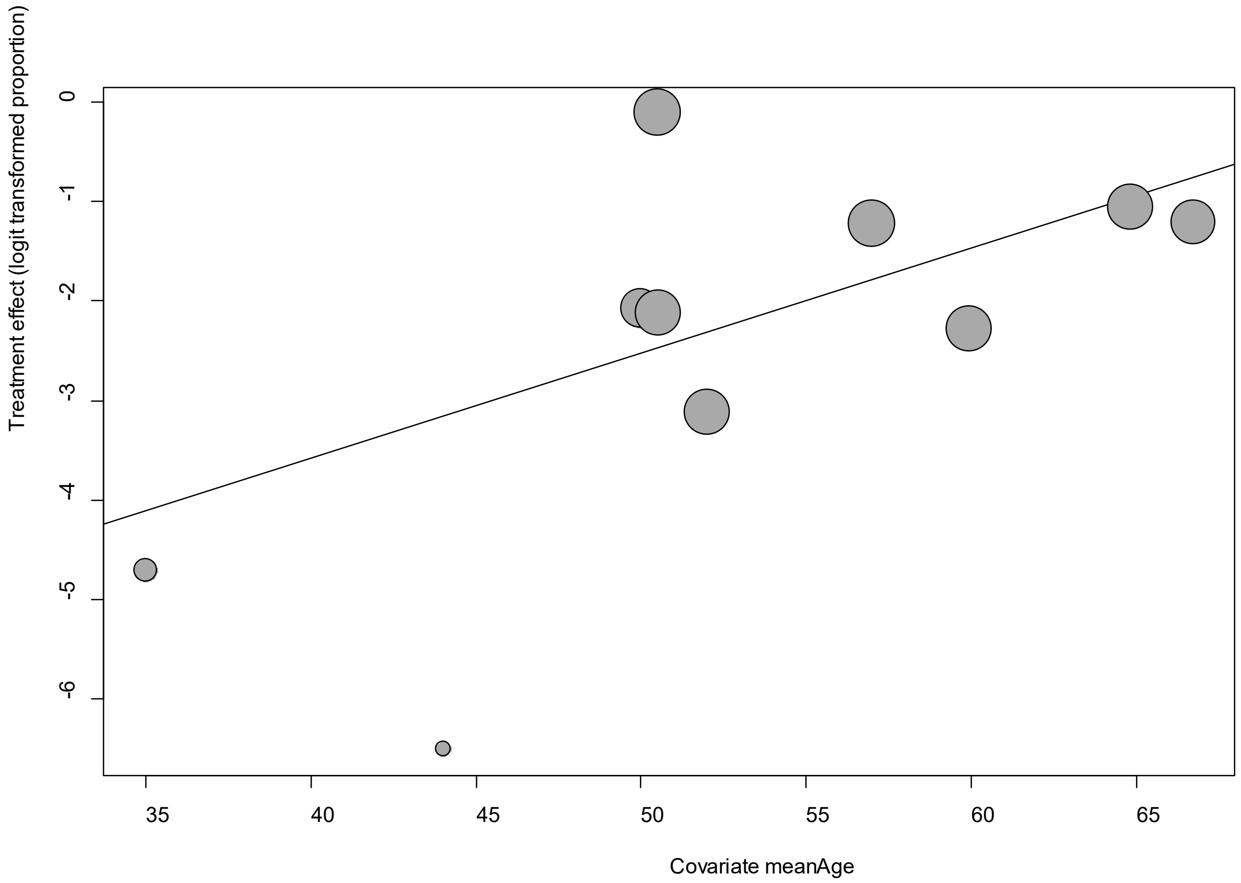


Point sizes are proportional to an inverse of the precision of the estimates.

The regression coefficient was 0.11 (95% confidence interval:0.02-0.19).(*P*=0.02).

**eFigure 17d. Bubble Plots (sex) on Depression**

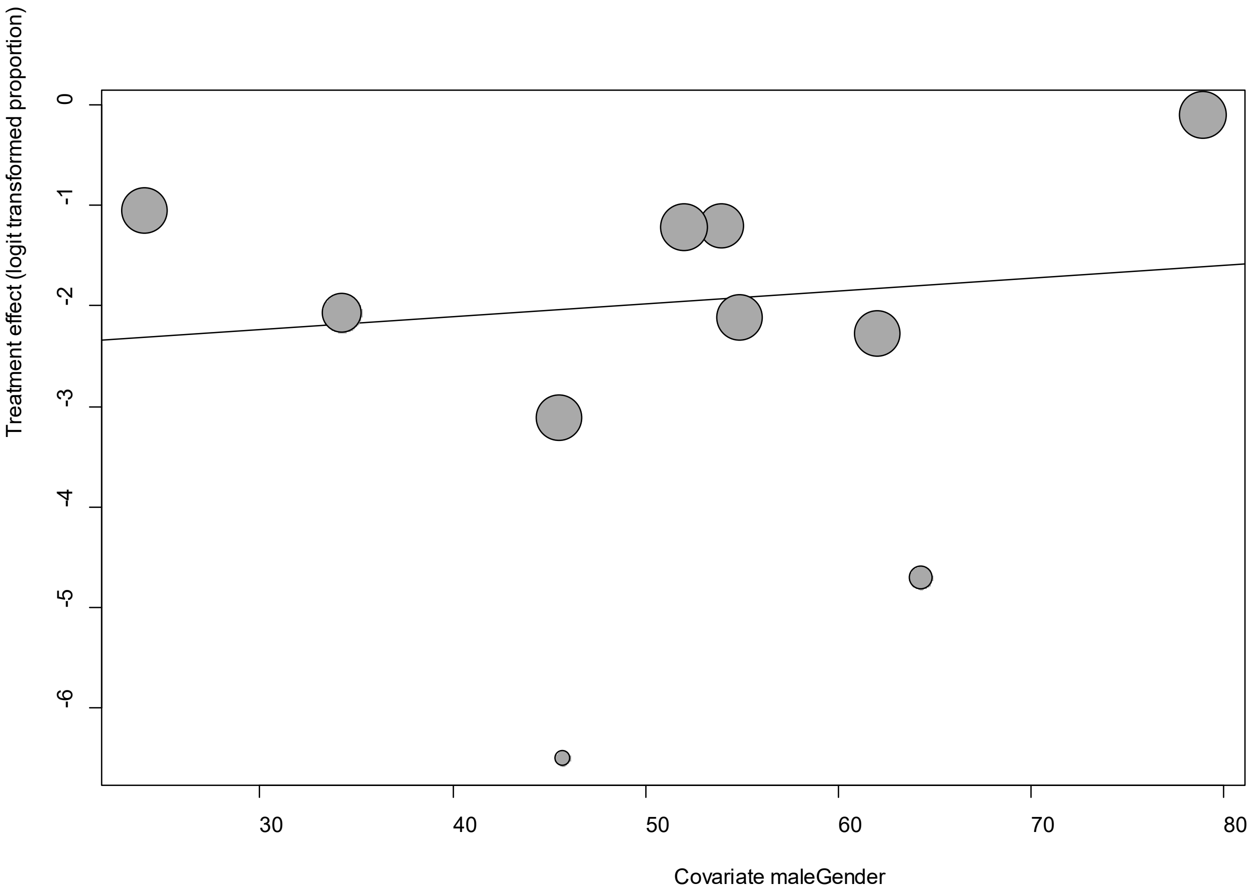


Point sizes are proportional to an inverse of the precision of the estimates.

The regression coefficient was 0.013 (95% confidence interval:-0.03-0.05).(*P*=0.53).

**eFigure 17e. Funnel Plot of studies reporting on Depression**

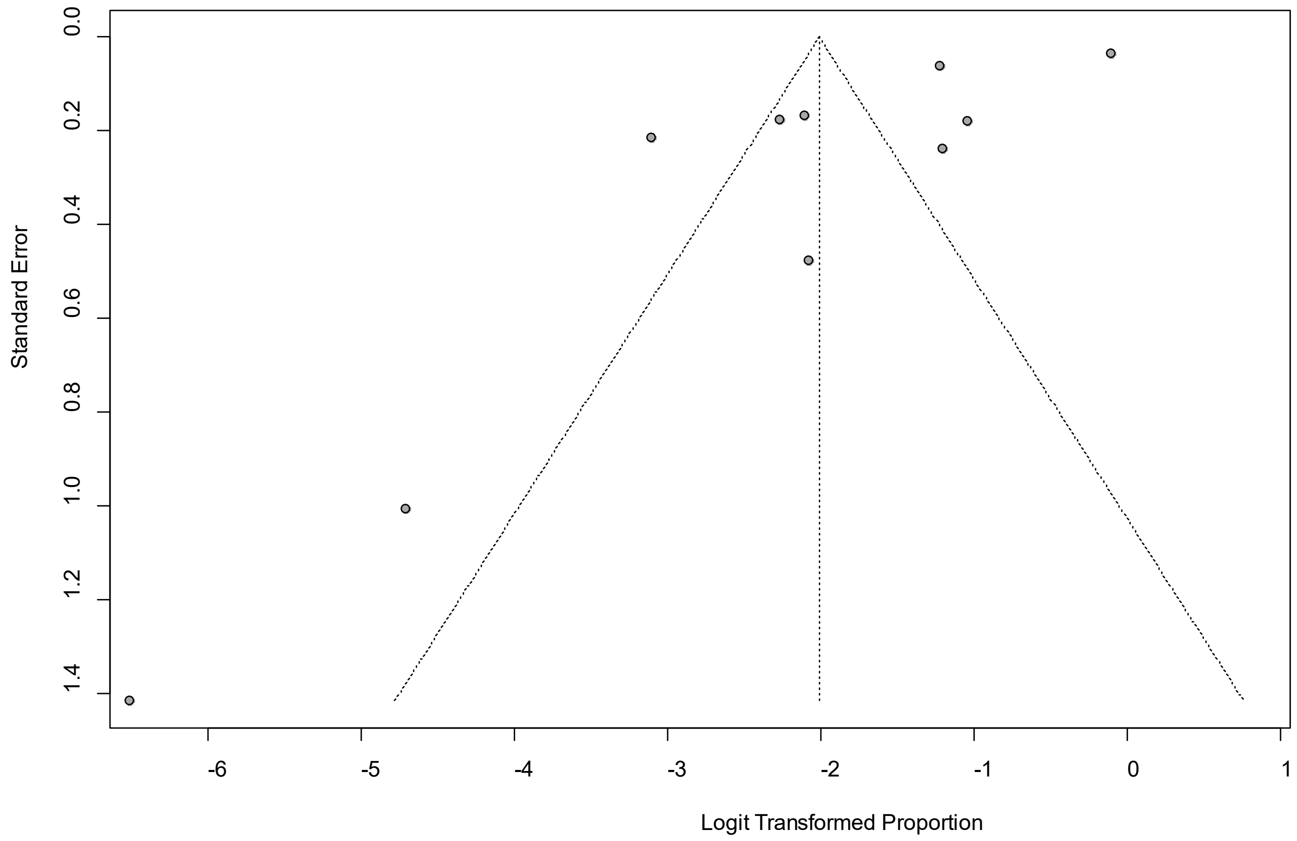


Egger's *P* was 0.01, indicating the presence of publication bias.

**eFigure 18a. Forest Plot on Diarrhea**

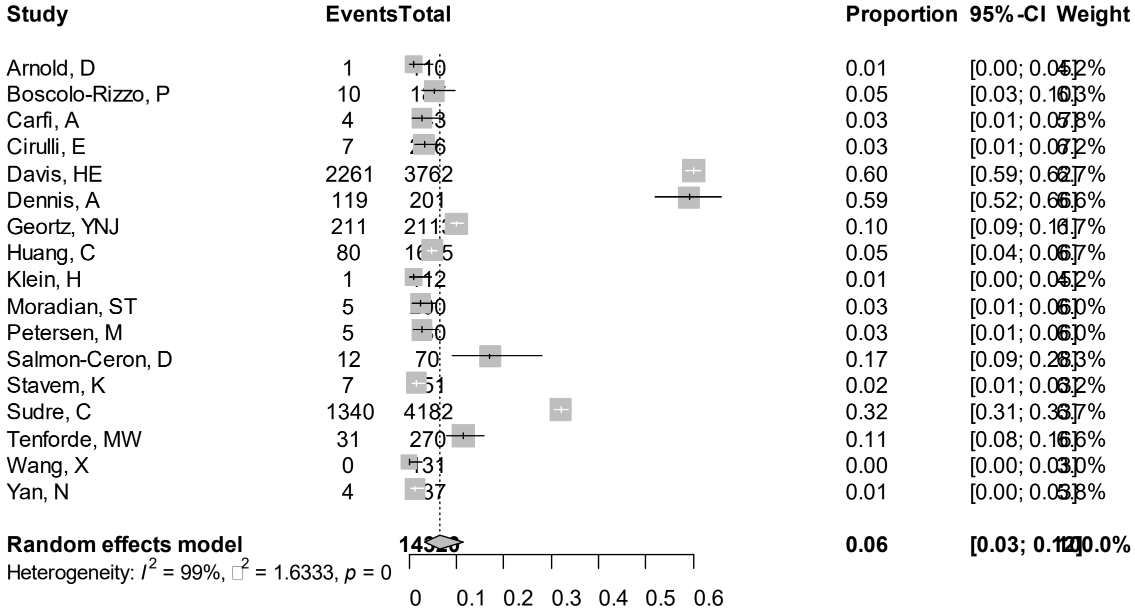


Point sizes are proportional to an inverse of the precision of the estimates.

**eFigure 18b. Bubble Plots (follow-up period) on Diarrhea**

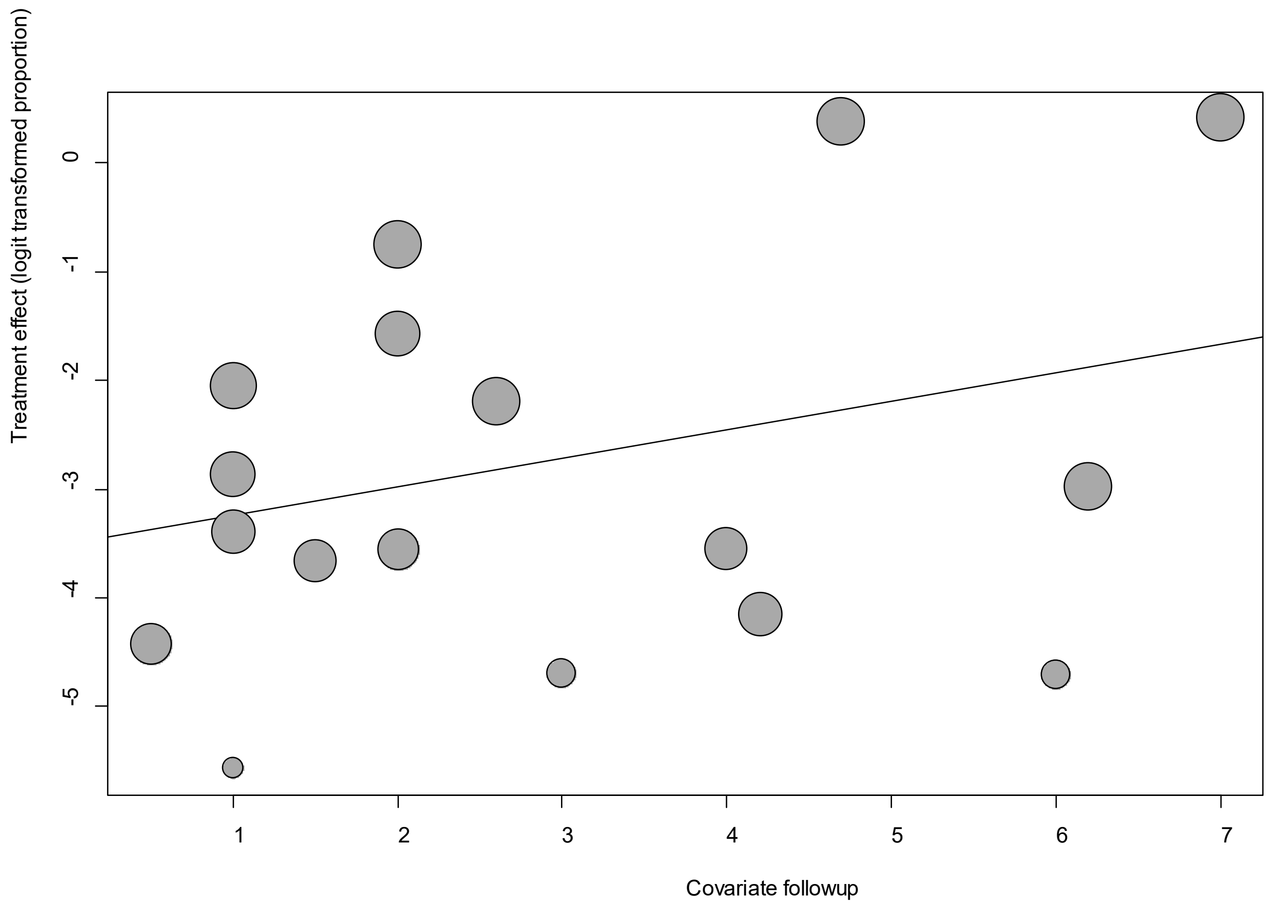


Point sizes are proportional to an inverse of the precision of the estimates.

The regression coefficient was 0.26 (95% confidence interval:-0.12-0.64).(*P*=0.18).

**eFigure 18c. Bubble Plots (age) on Diarrhea**

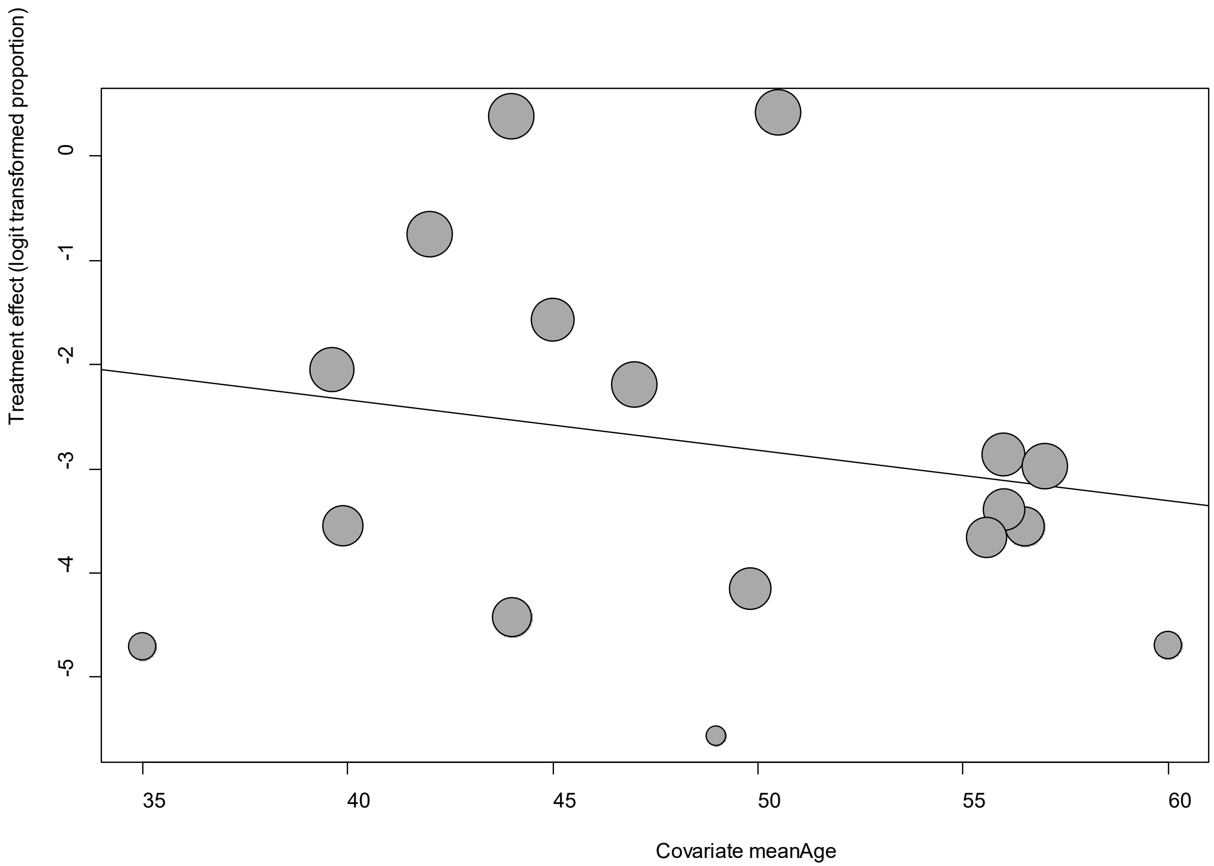


Point sizes are proportional to an inverse of the precision of the estimates.

The regression coefficient was -0.05 (95% confidence interval:-0.17-0.07).(*P*=0.44).

**eFigure 18d. Bubble Plots (sex) on Diarrhea**

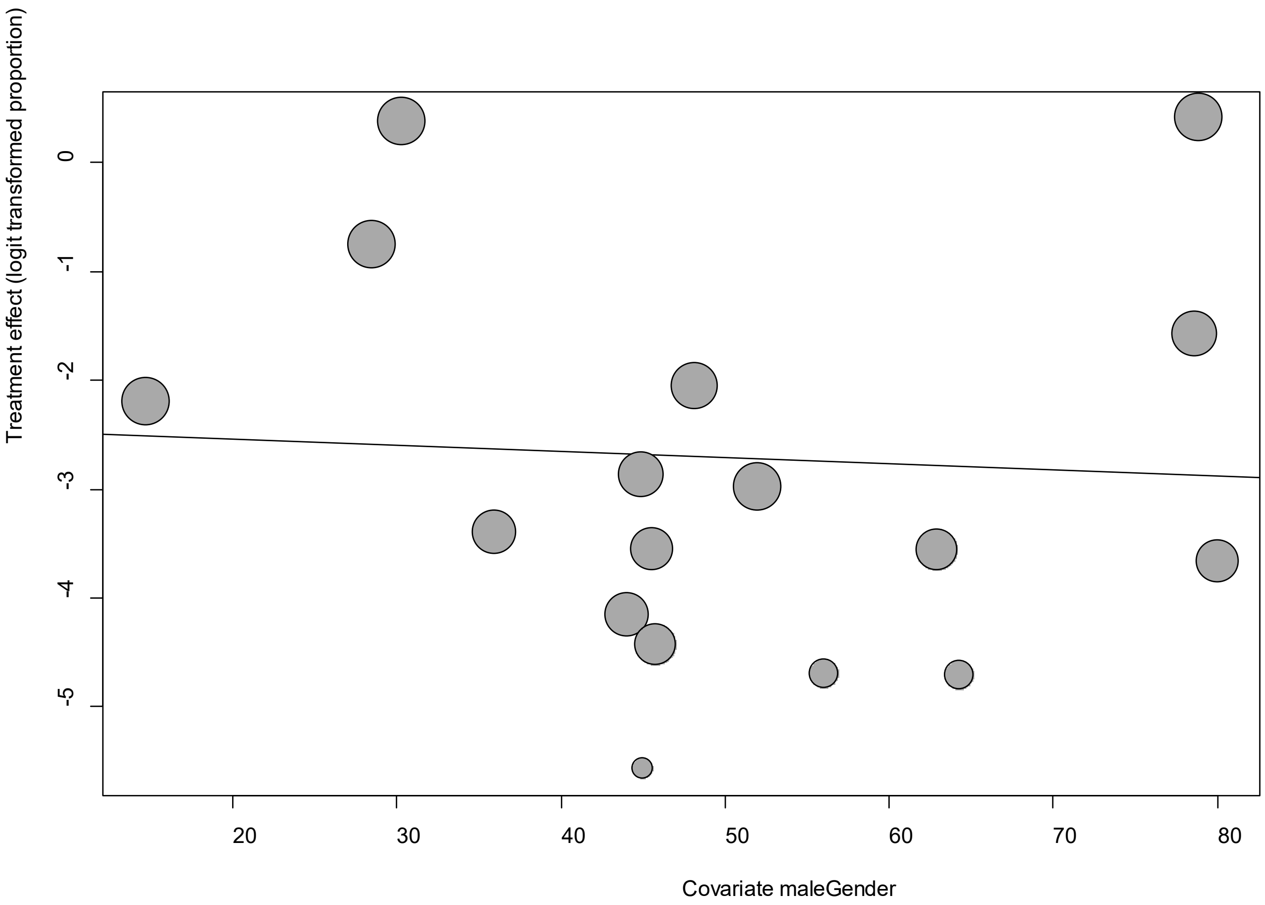


Point sizes are proportional to an inverse of the precision of the estimates.

The regression coefficient was -0.006 (95% confidence interval:-0.04-0.03).(*P*=0.78).

**eFigure 18e. Funnel Plot of studies reporting on Diarrhea**

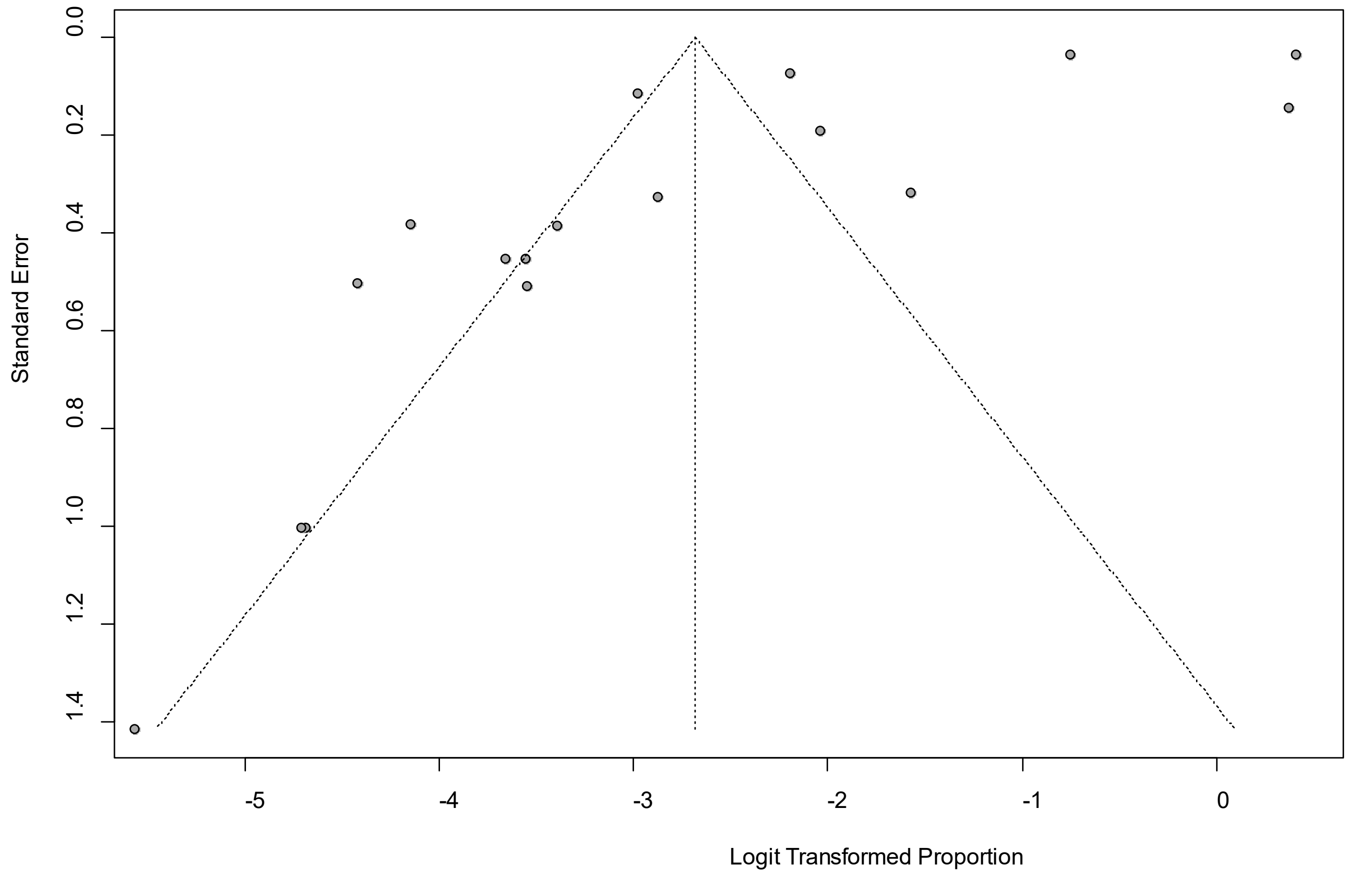


Egger's *P* was 0.02, indicating the presence of publication bias.

### eFigure 19a. Forest Plot on Dyspnea

| Study | Events | Total | Proportion | 95%-CI | Weight |
|---|---|---|---|---|---|
| Arnold, D | 43 | 110 | 0.39 | [0.30; 0.49] | 4.9% |
| Carfi, A | 61 | 143 | 0.43 | [0.34; 0.51] | 4.9% |
| Carvalho-Schneider, C | 10 | 1[illegible] | 0.08 | [0.04; 0.14] | 4.7% |
| Cirulli, E | 20 | 2[illegible] | 0.09 | [0.06; 0.14] | 4.9% |
| Davis, HE | 3499 | 3762 | 0.93 | [0.92; 0.94] | 5.0% |
| Dennis, A | 175 | 201 | 0.87 | [0.82; 0.91] | 4.9% |
| Eiros, R | 36 | 139 | 0.26 | [0.19; 0.34] | 4.9% |
| Geortz, YNJ | 1500 | 2113 | 0.71 | [0.69; 0.73] | 5.0% |
| Halpin, SJ | 50 | 100 | 0.50 | [0.40; 0.60] | 4.9% |
| Klein, H | 10 | 1[illegible] | 0.09 | [0.04; 0.16] | 4.7% |
| Mandal, S | 204 | 384 | 0.53 | [0.48; 0.58] | 5.0% |
| Moradian, ST | 2 | [illegible]00 | 0.01 | [0.00; 0.04] | 3.9% |
| Petersen, M | 10 | [illegible]0 | 0.06 | [0.03; 0.10] | 4.7% |
| Rahmani, H | 6 | [illegible]3 | 0.03 | [0.01; 0.07] | 4.6% |
| Salmon-Ceron, D | 30 | 70 | 0.43 | [0.31; 0.55] | 4.9% |
| Stavem, K | 72 | 451 | 0.16 | [0.13; 0.20] | 5.0% |
| Sudre, C | 1843 | 4182 | 0.44 | [0.43; 0.46] | 5.0% |
| Wang, X | 2 | [illegible]31 | 0.02 | [0.00; 0.05] | 3.9% |
| Weerahandi, H | 113 | 152 | 0.74 | [0.67; 0.81] | 4.9% |
| Yan, N | 7 | [illegible]37 | 0.02 | [0.01; 0.04] | 4.6% |
| Zhao, Y | 8 | 55 | 0.15 | [0.06; 0.27] | 4.6% |
| **Random effects model** | | **13341** | **0.25** | **[0.15; 0.38]** | **100.0%** |

Heterogeneity: $I^2 = 99\%$, $\tau^2 = 1.8530$, $p = 0$

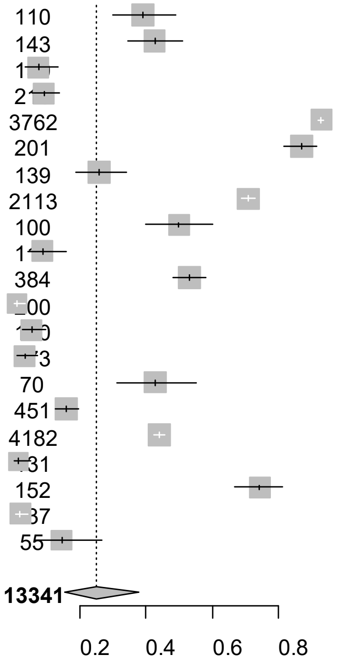


Point sizes are proportional to an inverse of the precision of the estimates.

**eFigure 19b. Bubble Plots (follow-up period) on Dyspnea**

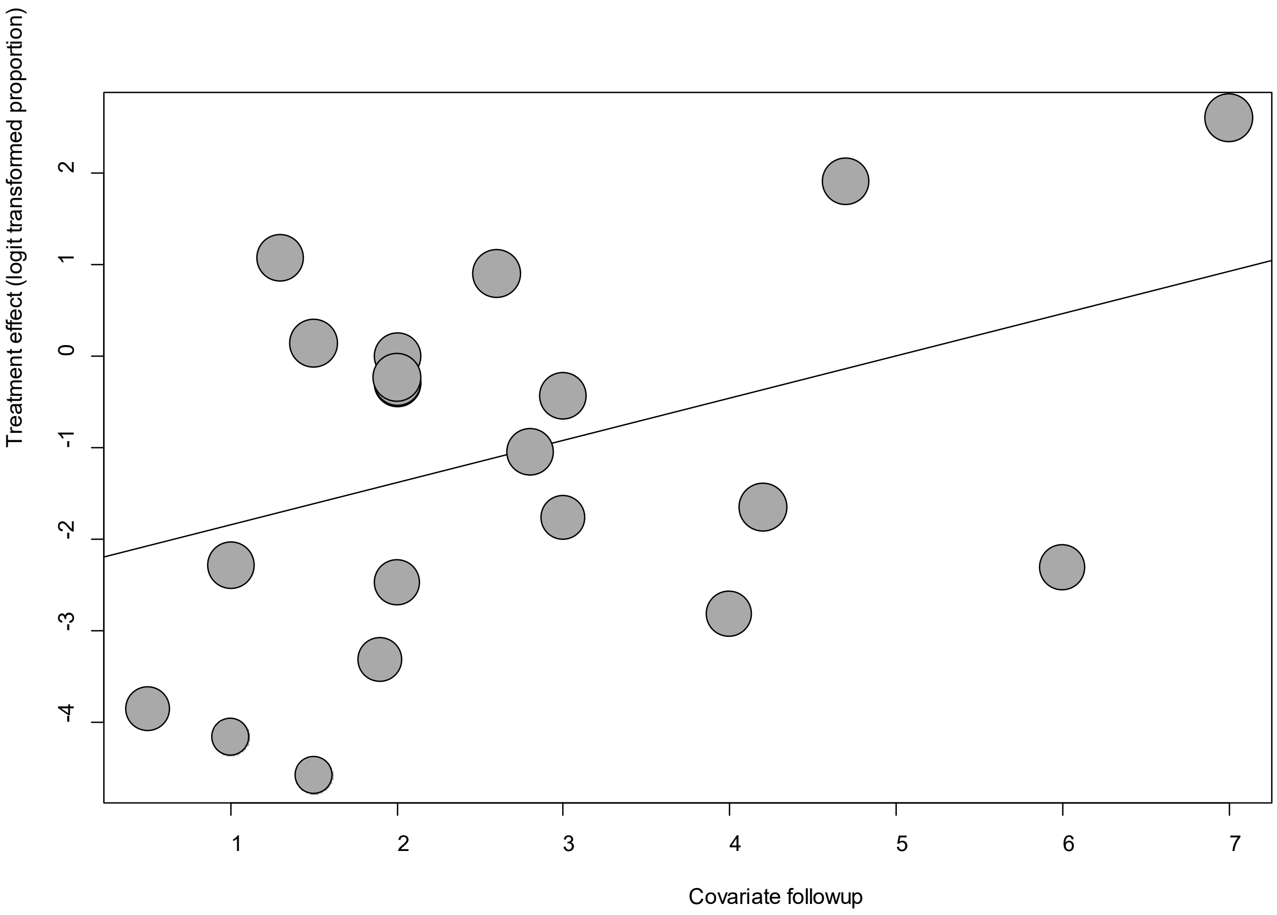


Point sizes are proportional to an inverse of the precision of the estimates.

The regression coefficient was 0.46 (95% confidence interval:0.17-0.76).(*P*=0.002).

**eFigure 19c. Bubble Plots (age) on Dyspnea**

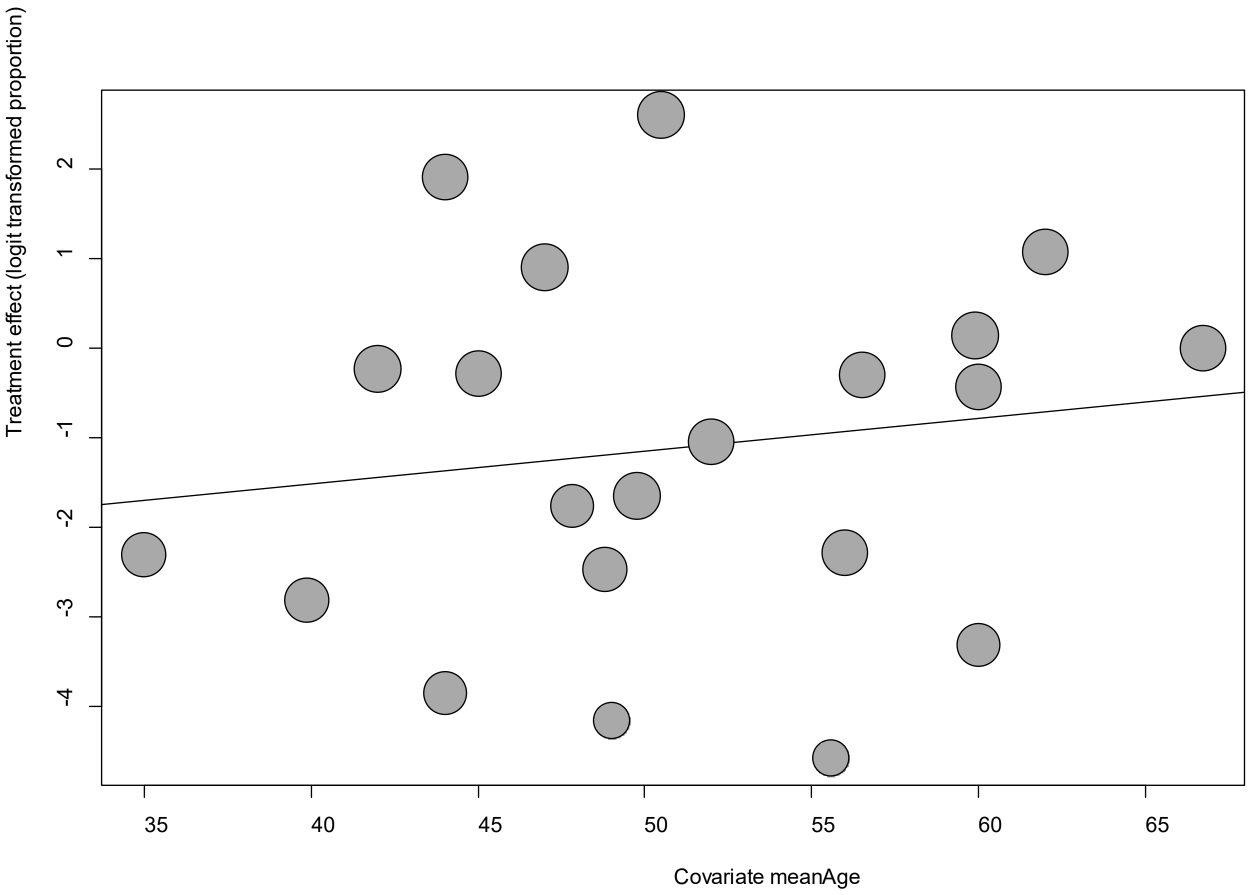


Point sizes are proportional to an inverse of the precision of the estimates.

The regression coefficient was 0.04 (95% confidence interval:-0.05-0.12).(*P*=0.38).

**eFigure 19d. Bubble Plots (sex) on Dyspnea**

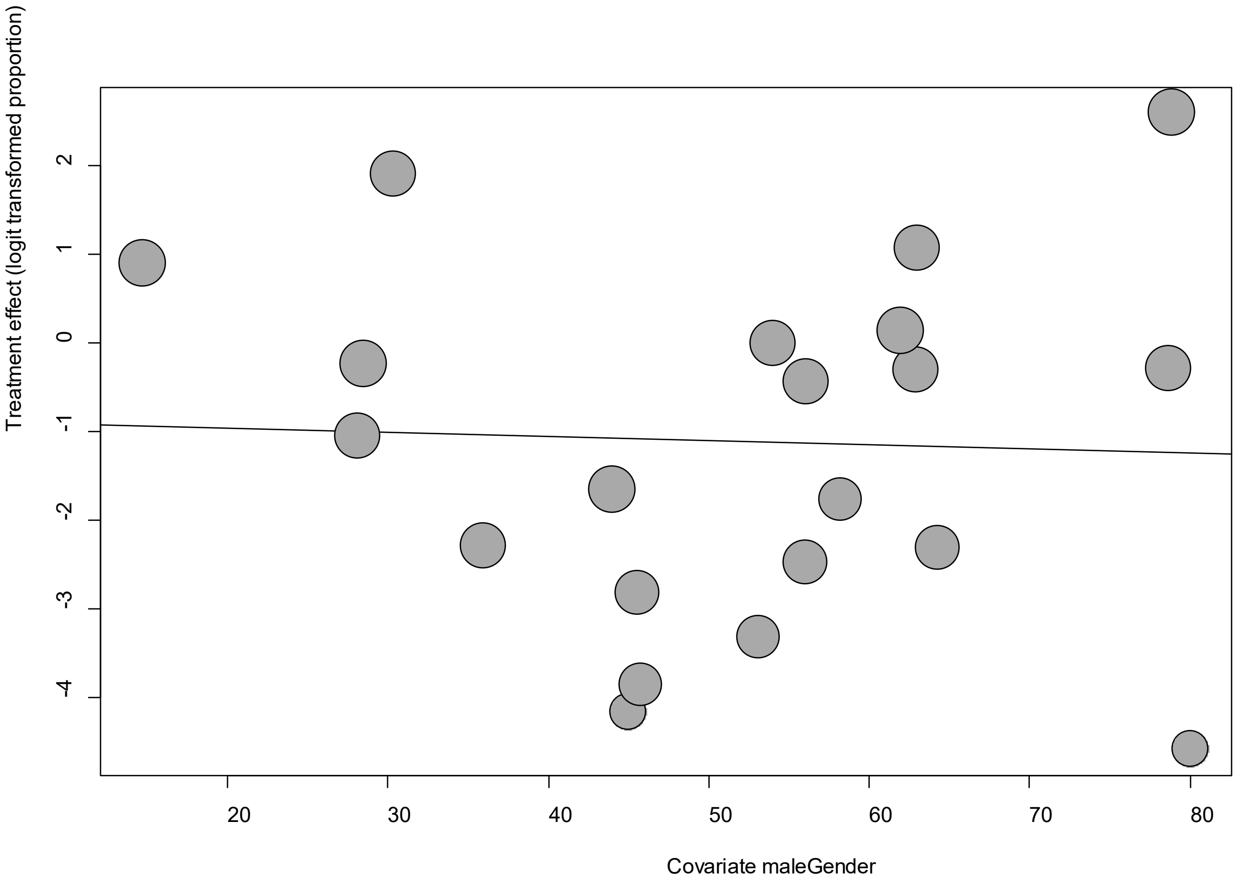


Point sizes are proportional to an inverse of the precision of the estimates.

The regression coefficient was -0.005 (95% confidence interval:-0.04-0.03).(*P*=0.80).

**eFigure 19e. Funnel Plot of studies reporting on Dyspnea**

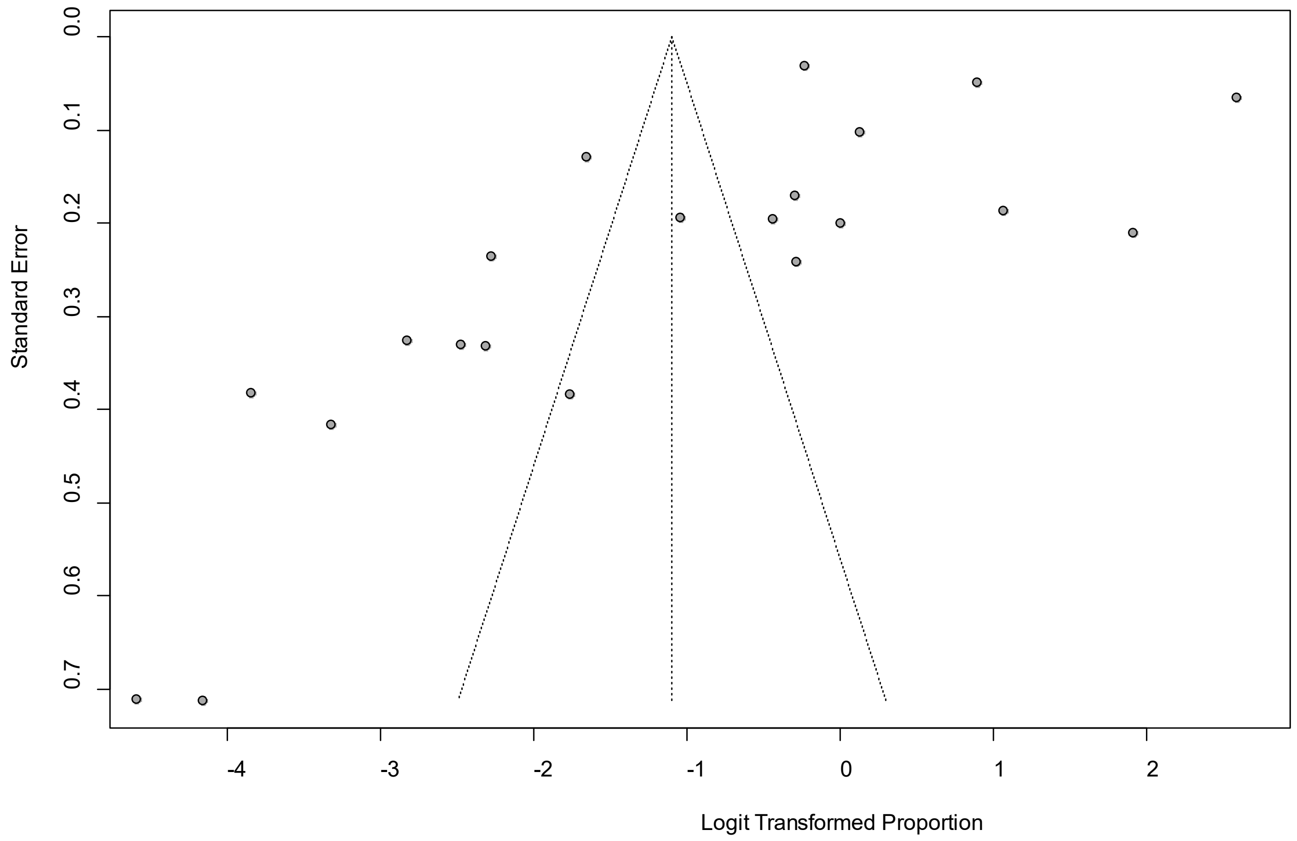


Egger's *P* was 0.13, indicating the presence of publication bias.

### eFigure 20a. Forest Plot on Fatigue

| Study | Events | Total | Proportion | 95%-CI | Weight |
|---|---|---|---|---|---|
| Arnold, D | 43 | 110 | 0.39 | [0.30; 0.49] | 4.0% |
| Boscolo-Rizzo, P | 29 | 187 | 0.16 | [0.11; 0.22] | 3.9% |
| Carfi, A | 75 | 143 | 0.52 | [0.44; 0.61] | 4.0% |
| Cheng, DO | 64 | 109 | 0.59 | [0.49; 0.68] | 4.0% |
| Cirulli, E | 8 | 6 | 0.04 | [0.02; 0.07] | 3.8% |
| Davis, HE | 3694 | 3762 | 0.98 | [0.98; 0.99] | 4.0% |
| Dennis, A | 197 | 201 | 0.98 | [0.95; 0.99] | 3.6% |
| Eiros, R | 37 | 139 | 0.27 | [0.19; 0.35] | 4.0% |
| Geortz, YNJ | 1838 | 2113 | 0.87 | [0.85; 0.88] | 4.0% |
| Halpin, SJ | 64 | 100 | 0.64 | [0.54; 0.73] | 3.9% |
| Huang, C | 1038 | 1655 | 0.63 | [0.60; 0.65] | 4.0% |
| Klein, H | 23 | 112 | 0.21 | [0.13; 0.29] | 3.9% |
| Lavoto, A | 83 | 121 | 0.69 | [0.60; 0.77] | 4.0% |
| Mandal, S | 265 | 384 | 0.69 | [0.64; 0.74] | 4.0% |
| Moradian, ST | 39 | 200 | 0.20 | [0.14; 0.26] | 4.0% |
| Petersen, M | 52 | 180 | 0.29 | [0.22; 0.36] | 4.0% |
| Pilotto, A | 56 | 165 | 0.34 | [0.27; 0.42] | 4.0% |
| Salmon-Ceron, D | 51 | 70 | 0.73 | [0.61; 0.83] | 3.9% |
| Savarraj, JPJ | 19 | 45 | 0.42 | [0.28; 0.58] | 3.8% |
| Sudre, C | 3591 | 4182 | 0.86 | [0.85; 0.87] | 4.0% |
| Tenforde, MW | 95 | 270 | 0.35 | [0.29; 0.41] | 4.0% |
| Townsend, L | 67 | 128 | 0.52 | [0.43; 0.61] | 4.0% |
| Wang, X | 0 | 131 | 0.00 | [0.00; 0.03] | 2.0% |
| Xiong, Q | 152 | 538 | 0.28 | [0.24; 0.32] | 4.0% |
| Yan, N | 5 | 37 | 0.01 | [0.00; 0.03] | 3.6% |
| Zhao, Y | 9 | 55 | 0.16 | [0.08; 0.29] | 3.8% |
| **Random effects model** | | **15653** | **0.45** | **[0.32; 0.59]** | **100.0%** |

Heterogeneity: $I^2$ = 99%, $\square^2$ = 1.9344, $p$ = 0

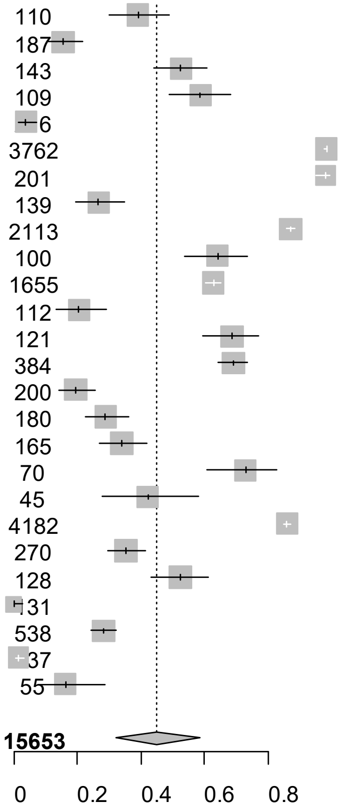


Point sizes are proportional to an inverse of the precision of the estimates.

**eFigure 20b. Bubble Plots (follow-up period) on Fatigue**

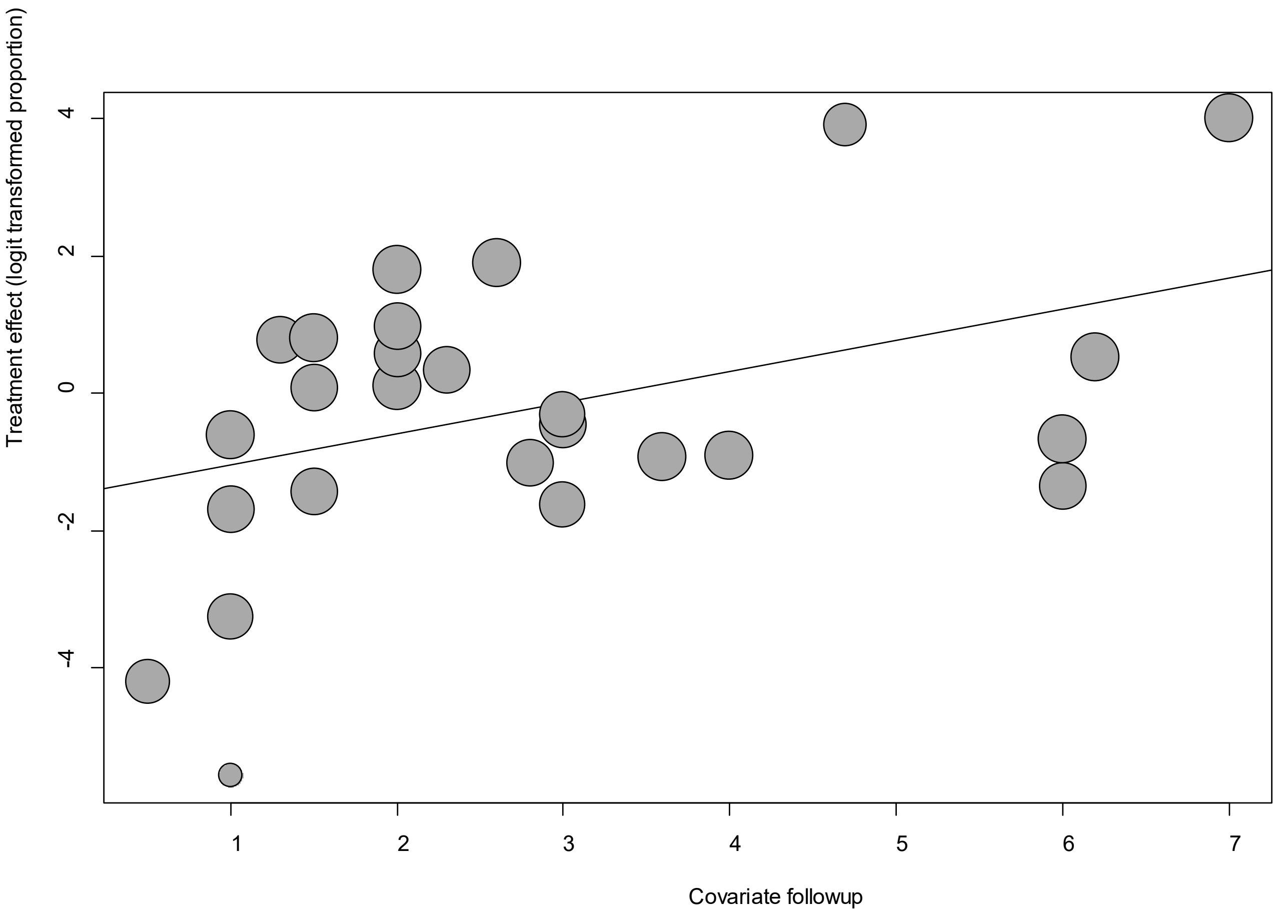


Point sizes are proportional to an inverse of the precision of the estimates.

The regression coefficient was 0.46 (95% confidence interval:0.12-0.79).(*P*=0.007).

### eFigure 20c. Bubble Plots (age) on Fatigue

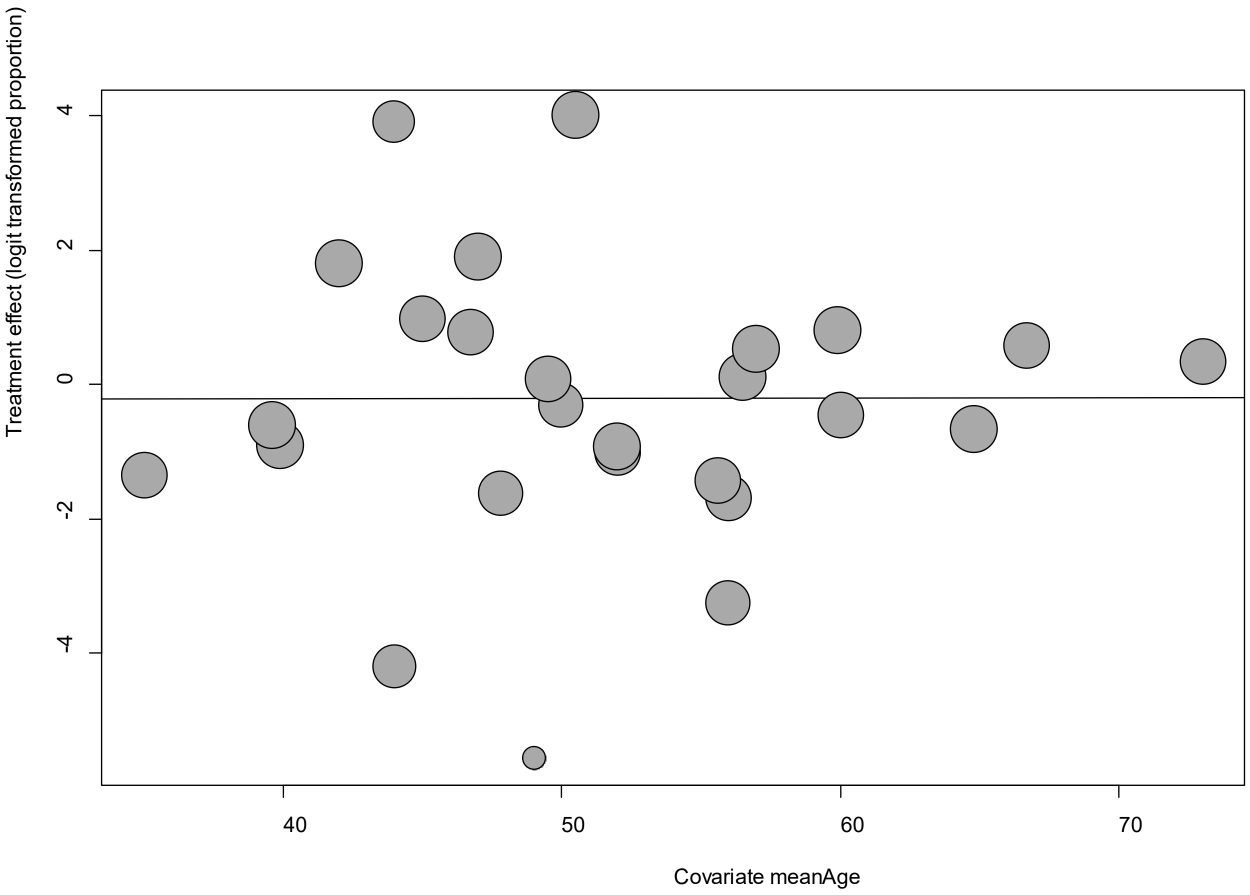


Point sizes are proportional to an inverse of the precision of the estimates.

The regression coefficient was 0.0008 (95% confidence interval:-0.06-0.06).(*P*=0.98).

**eFigure 20d. Bubble Plots (sex) on Fatigue**

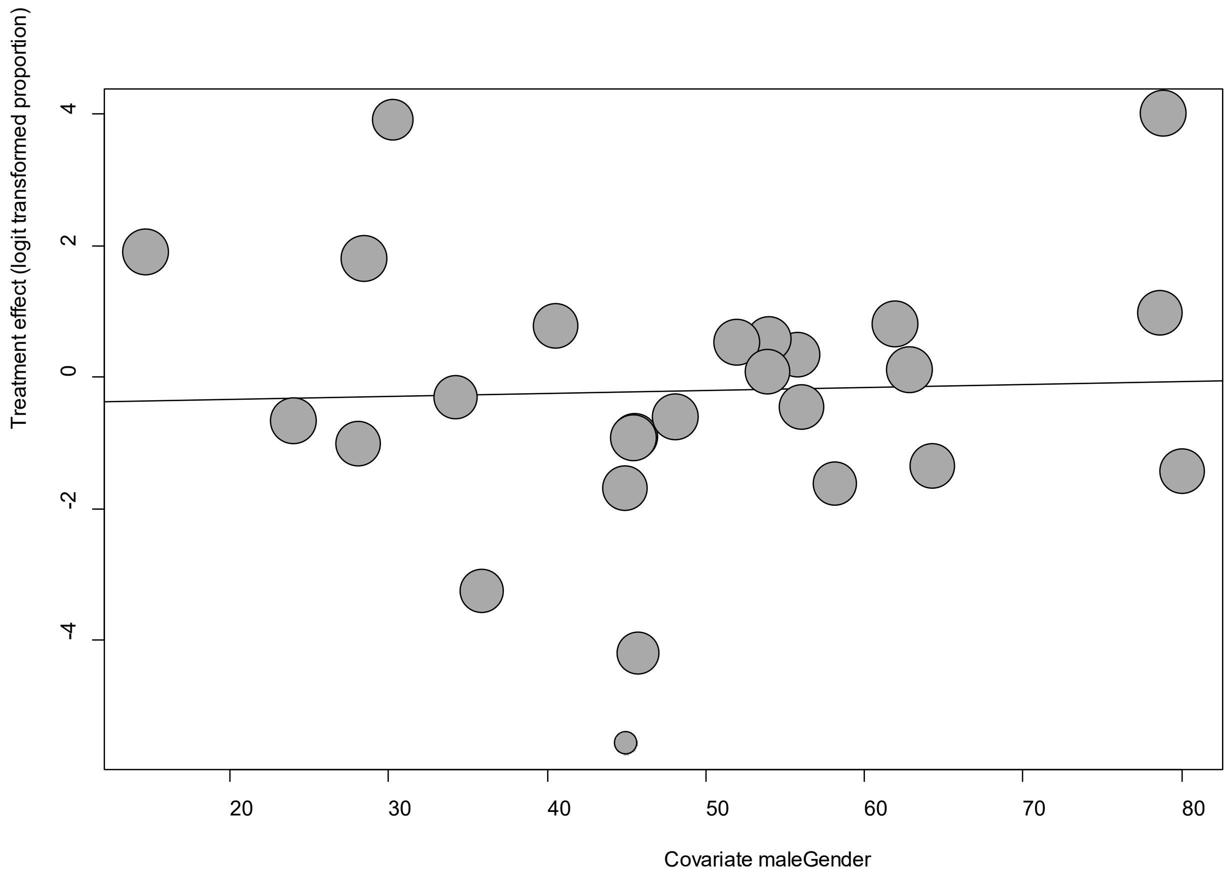


Point sizes are proportional to an inverse of the precision of the estimates.

The regression coefficient was 0.004 (95% confidence interval:-0.03-0.04).(*P*=0.80).

**eFigure 20e. Funnel Plot of studies reporting on Fatigue**

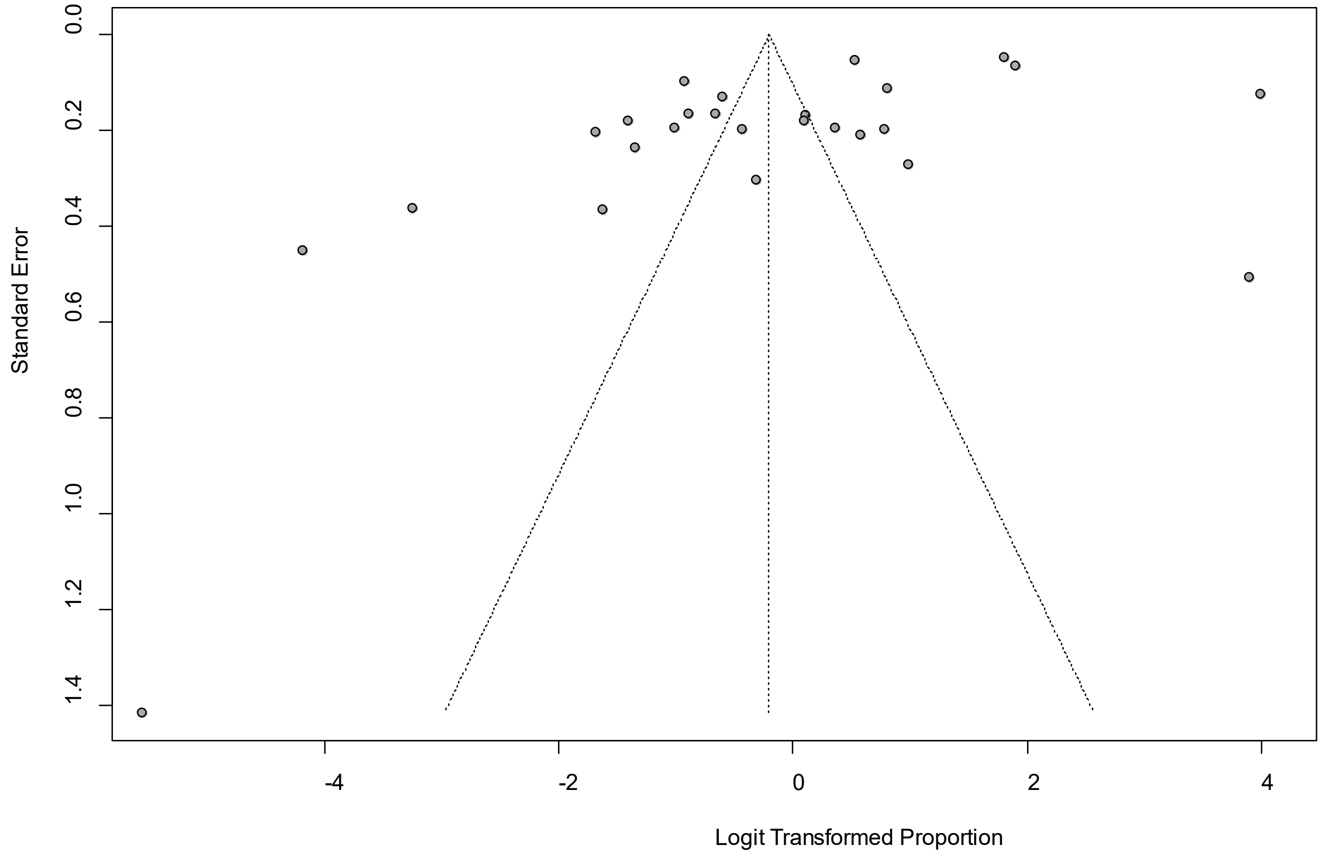


Egger's *P* was 0.006, indicating the presence of publication bias.

### eFigure 21a. Forest Plot on Fever

| Study | Events | Total | Proportion | 95%-CI | Weight |
|---|---|---|---|---|---|
| Arnold, D | 2 | [illegible]10 | 0.02 | [0.00; 0.0[illegible]] | [illegible].3% |
| Boscolo-Rizzo, P | 5 | [illegible]37 | 0.03 | [0.01; 0.0[illegible]] | [illegible].3% |
| Carvalho-Schneider, C | 0 | 130 | 0.00 | [0.00; 0.0[illegible]] | [illegible].3% |
| Davis, HE | 1159 | 3762 | 0.31 | [0.29; 0.3[illegible]] | [illegible].6% |
| Dennis, A | 151 | 201 | 0.75 | [0.69; 0.8[illegible]] | [illegible].3% |
| Geortz, YNJ | 465 | 2113 | 0.22 | [0.20; 0.2[illegible]] | [illegible].6% |
| Huang, C | 2 | [illegible]655 | 0.00 | [0.00; 0.0[illegible]] | [illegible].4% |
| Lavoto, A | 89 | 121 | 0.74 | [0.65; 0.8[illegible]] | [illegible].0% |
| Moradian, ST | 3 | [illegible]00 | 0.01 | [0.00; 0.0[illegible]] | [illegible].3% |
| Petersen, M | 0 | 180 | 0.00 | [0.00; 0.0[illegible]] | [illegible].3% |
| Poncet-Megemont, L | 99 | 139 | 0.71 | [0.63; 0.7[illegible]] | [illegible].1% |
| Stavem, K | 0 | 451 | 0.00 | [0.00; 0.0[illegible]] | [illegible].3% |
| Sudre, C | 1946 | 4182 | 0.47 | [0.45; 0.4[illegible]] | [illegible].6% |
| Tenforde, MW | 8 | [illegible]0 | 0.03 | [0.01; 0.0[illegible]] | [illegible].0% |
| Wang, X | 0 | 131 | 0.00 | [0.00; 0.0[illegible]] | [illegible].3% |
| Yan, N | 0 | 337 | 0.00 | [0.00; 0.0[illegible]] | [illegible].3% |
| **Random effects model** | | **1416[illegible]** | **0.12** | **[0.08; 0.1[illegible]]** | **[illegible]0.0%** |

Heterogeneity: $I^2$ = 98%, [illegible]$^2$ = 0.6266, $p$ < 0.01

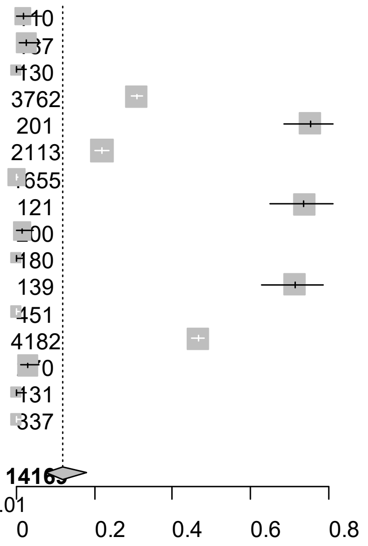


Point sizes are proportional to an inverse of the precision of the estimates.

**eFigure 21b. Bubble Plots (follow-up period) on Fever**

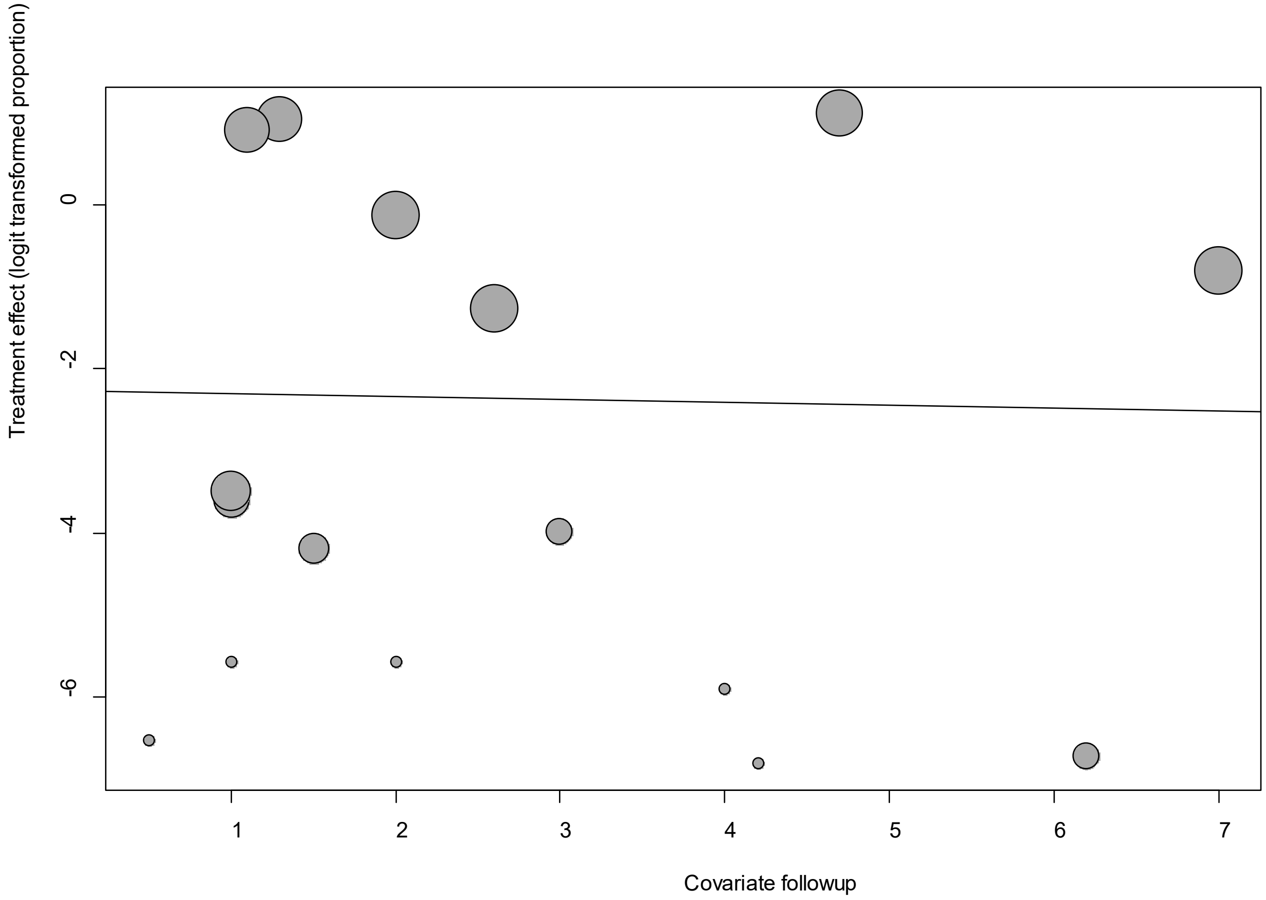


Point sizes are proportional to an inverse of the precision of the estimates.

The regression coefficient was -0.04 (95% confidence interval:-0.35-0.27).(*P*=0.83).

**eFigure 21c. Bubble Plots (age) on Fever**

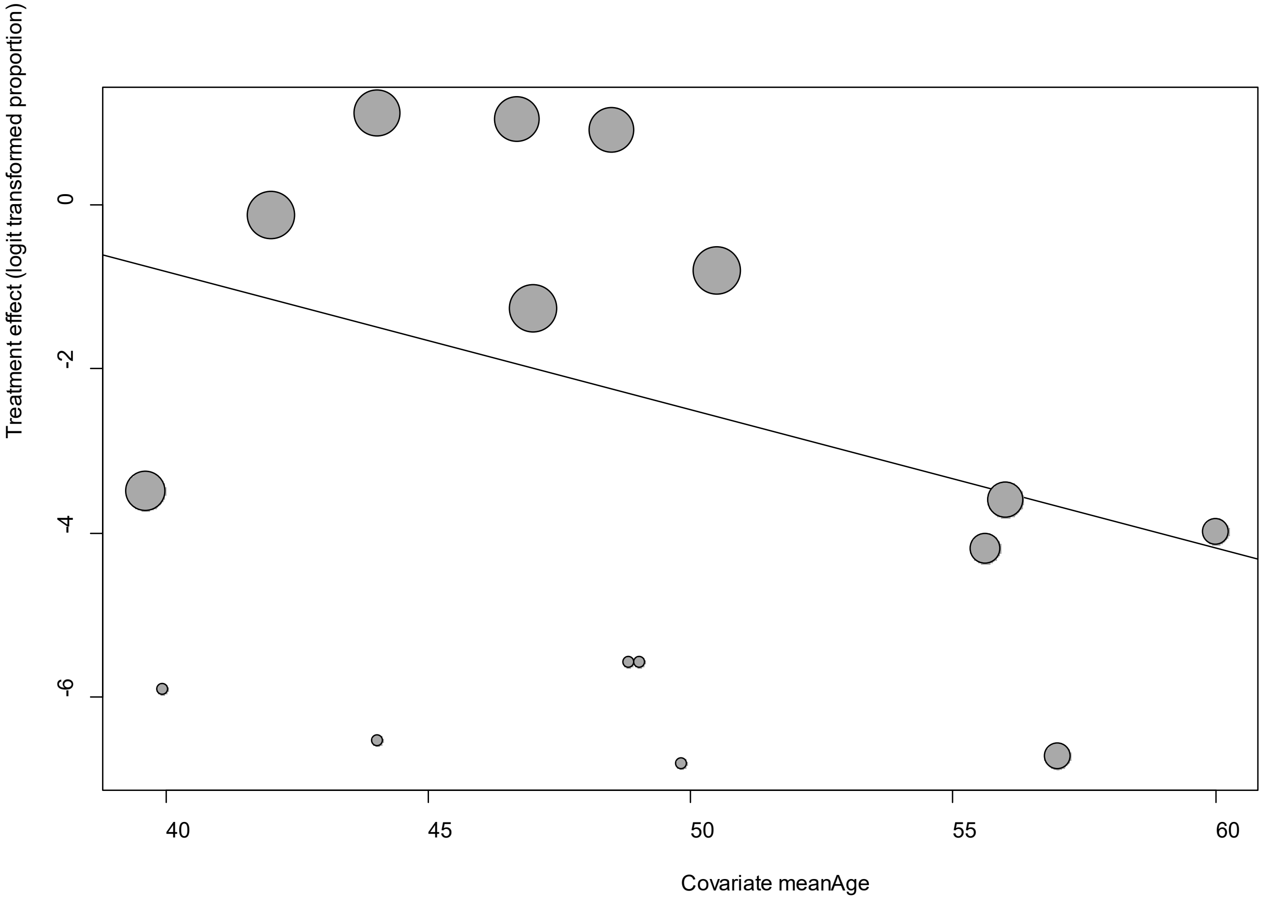


Point sizes are proportional to an inverse of the precision of the estimates.

The regression coefficient was -0.17 (95% confidence interval:-0.27—0.07).(*P*=0.0009).

### eFigure 21d. Bubble Plots (sex) on Fever

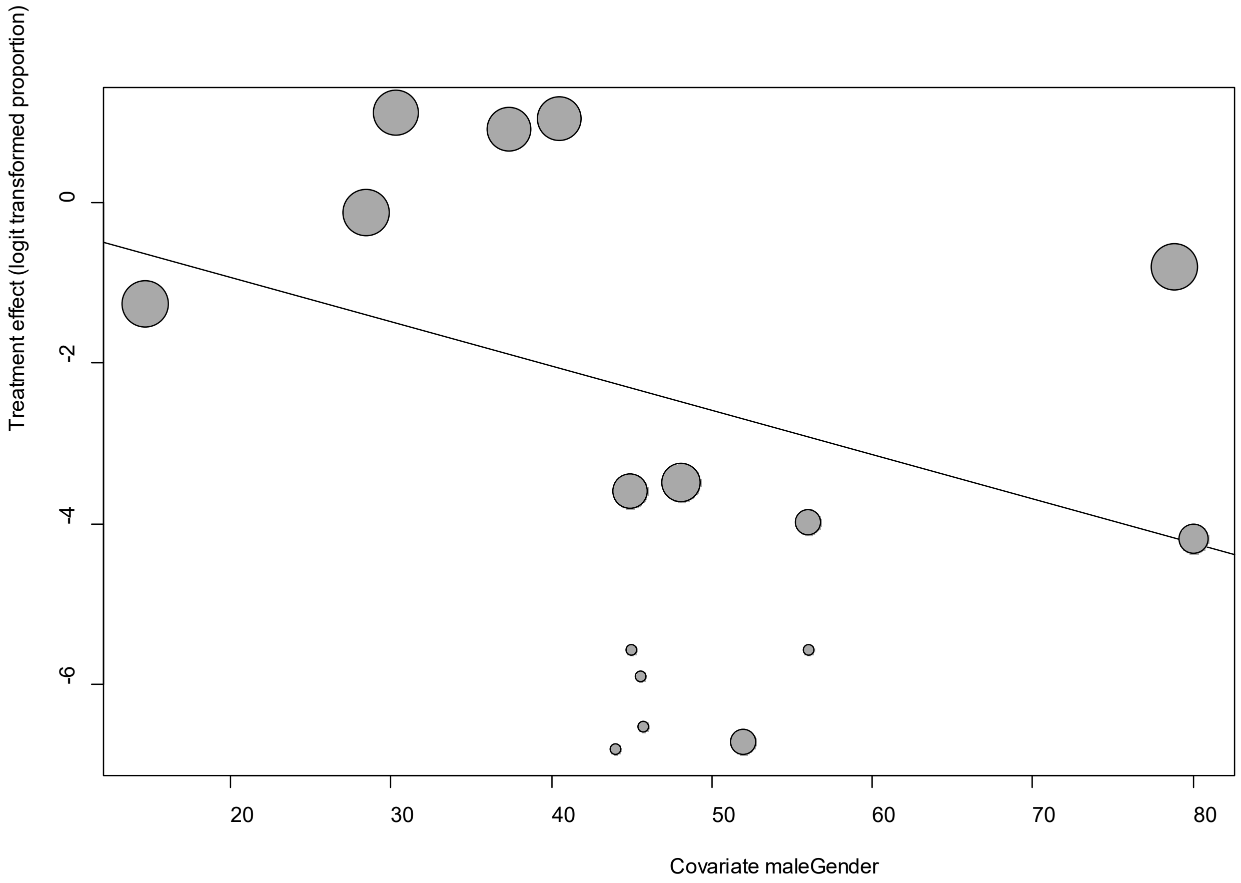


Point sizes are proportional to an inverse of the precision of the estimates.

The regression coefficient was -0.06 (95% confidence interval:-0.09—0.02).(*P*=0.0019).

**eFigure 21e. Funnel Plot of studies reporting on Fever**

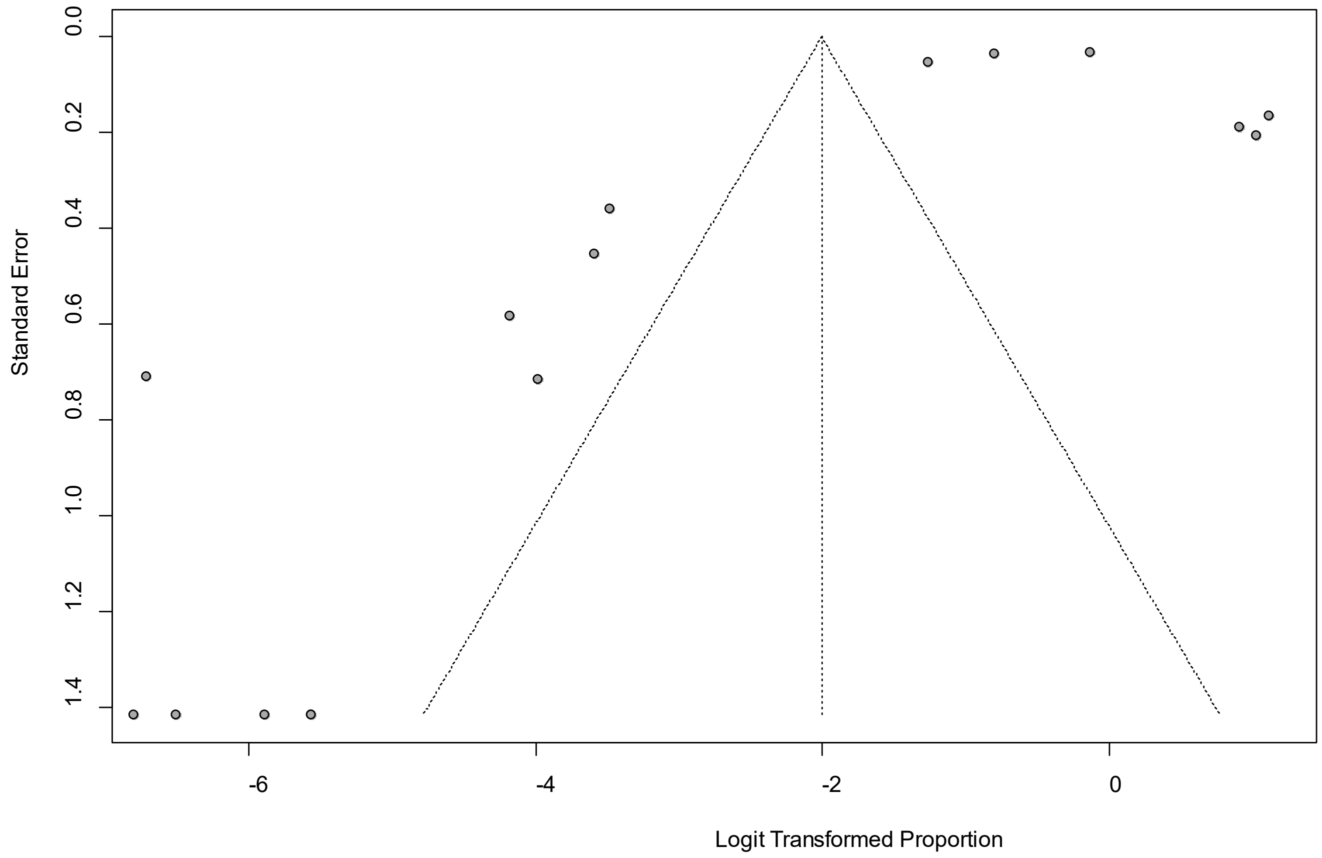


Egger’s *P* was 0.17, indicating the absence of publication bias.

### eFigure 22a. Forest Plot on Insomnia

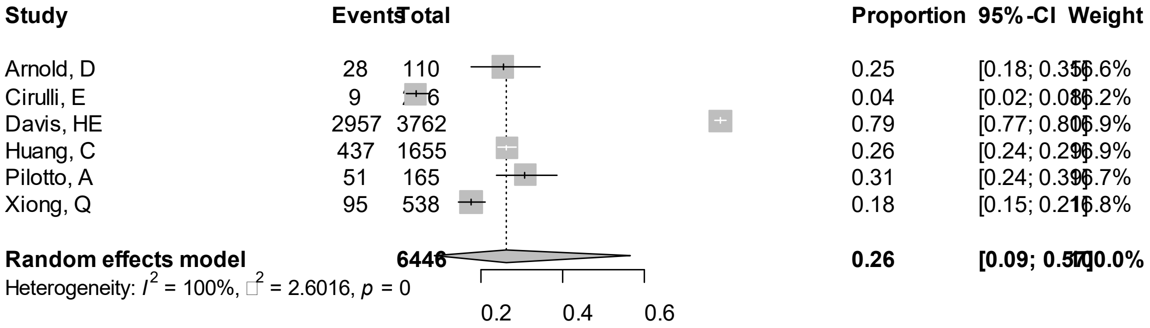


Point sizes are proportional to an inverse of the precision of the estimates.

### eFigure 22b. Bubble Plots (follow-up period) on Insomnia

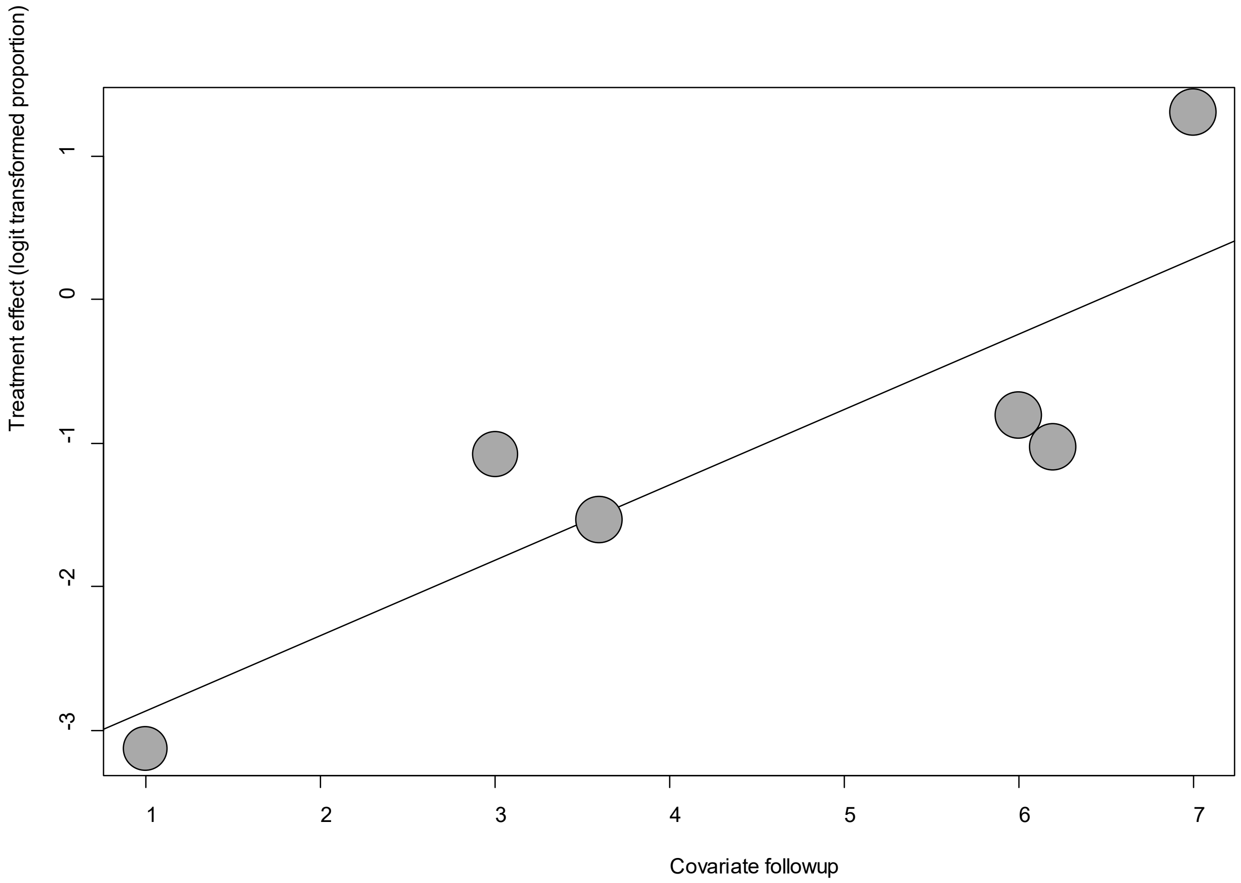


Point sizes are proportional to an inverse of the precision of the estimates.

The regression coefficient was 0.53 (95% confidence interval:0.06-0.99).(*P*=0.03).

### eFigure 22c. Bubble Plots (age) on Insomnia

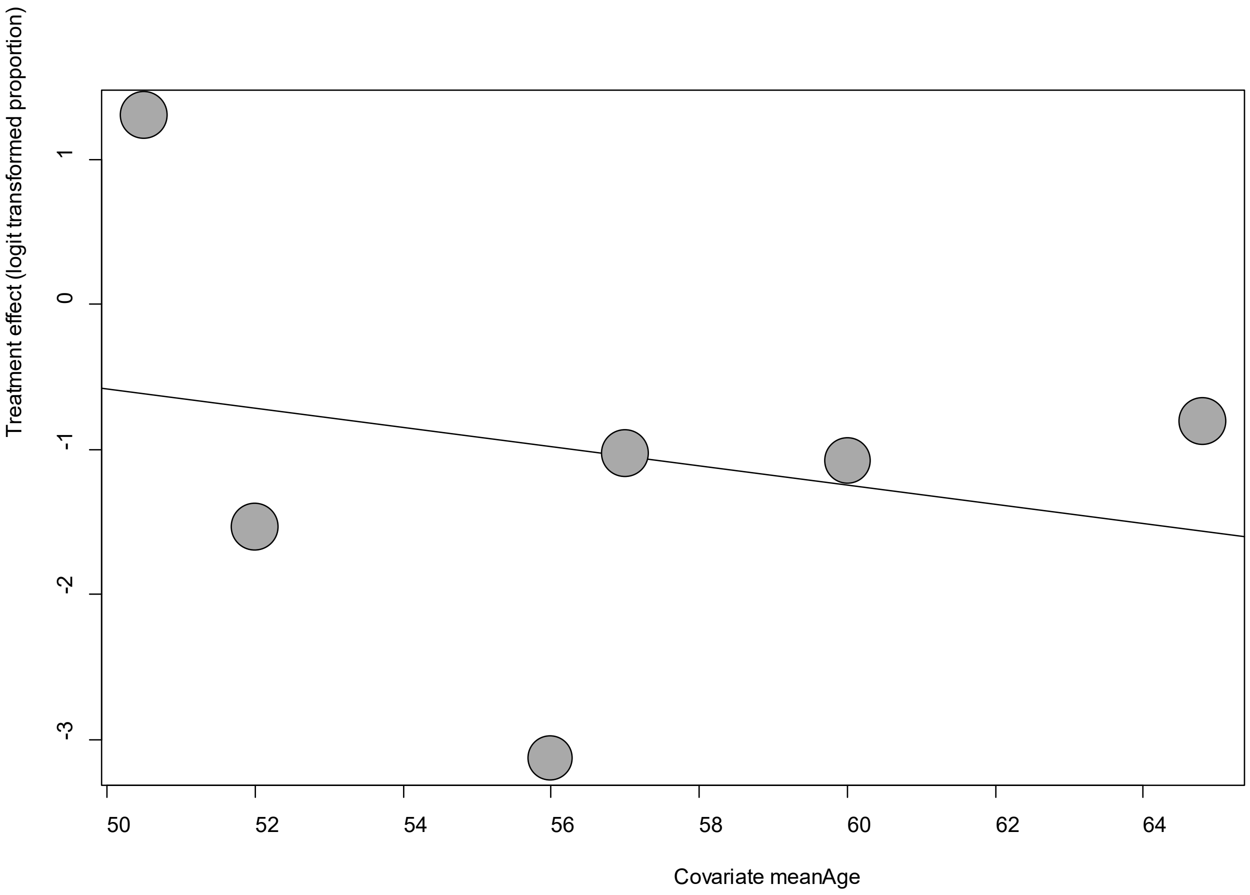


Point sizes are proportional to an inverse of the precision of the estimates.

The regression coefficient was -0.07 (95% confidence interval:-0.30-0.17).(*P*=0.58).

**eFigure 22d. Bubble Plots (sex) on Insomnia**

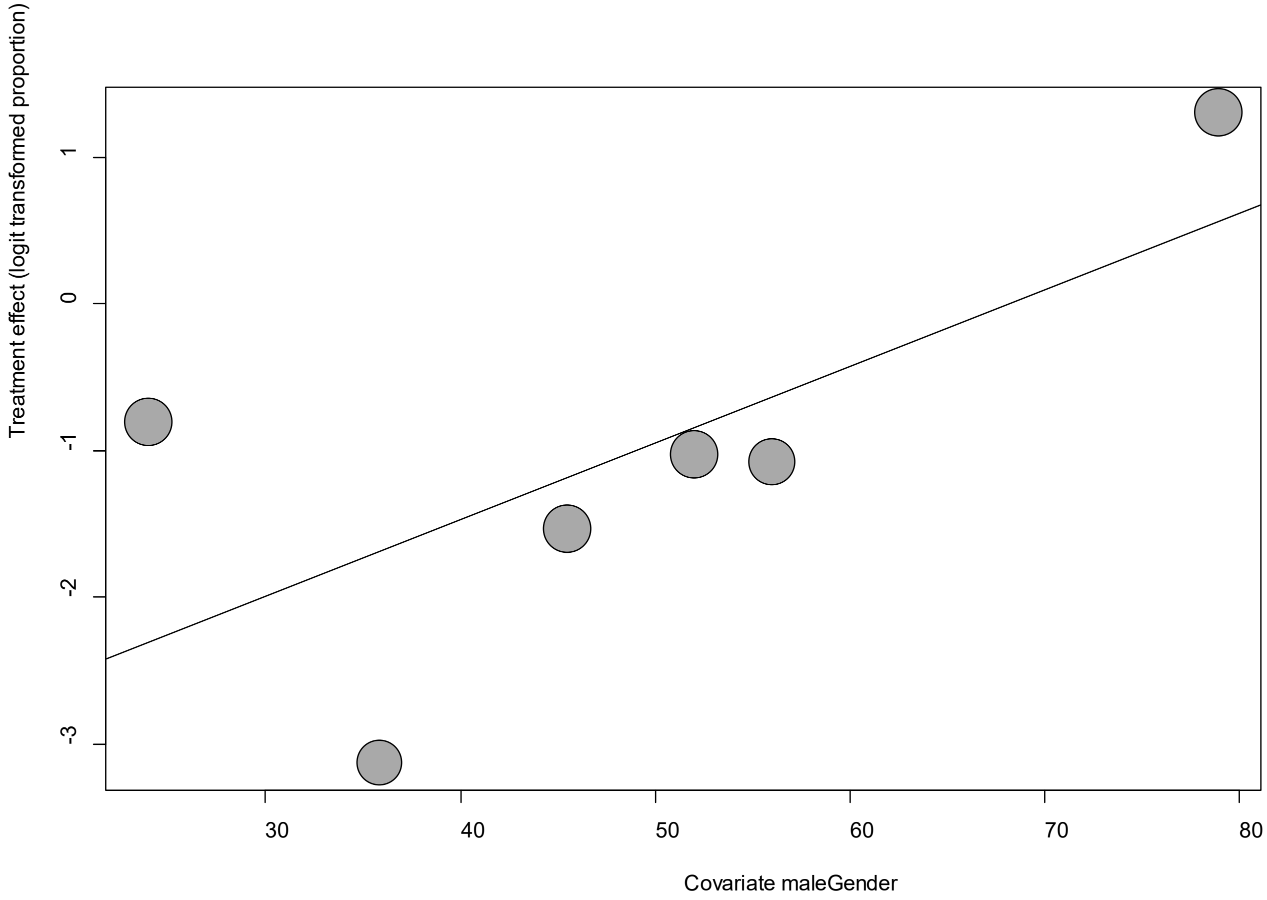


Point sizes are proportional to an inverse of the precision of the estimates.

The regression coefficient was 0.05 (95% confidence interval:0.01-0.09).(*P*=0.007).

**eFigure 22e. Funnel Plot of studies reporting on Insomnia**

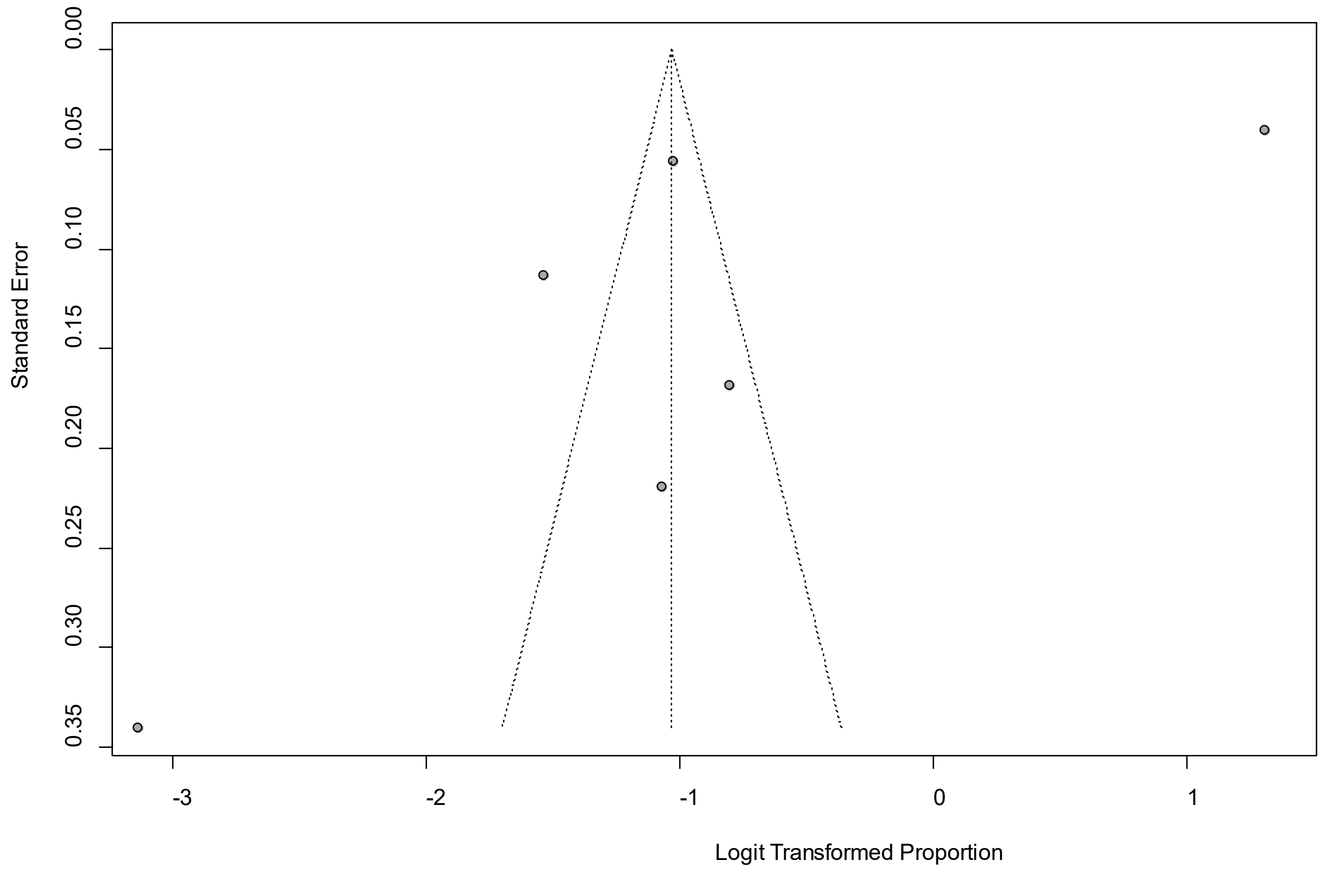


Egger's *P* was 0.20, indicating the absence of publication bias.

**eFigure 23a. Forest Plot on Memory impairment**

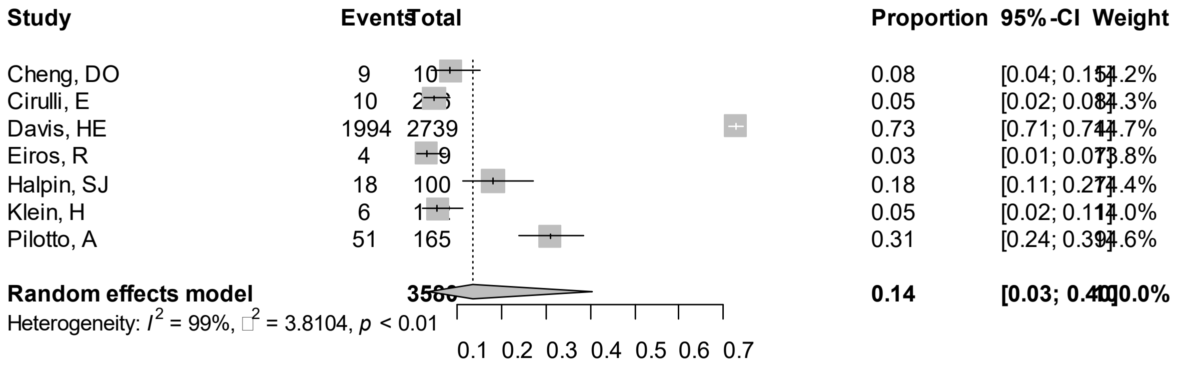


Point sizes are proportional to an inverse of the precision of the estimates.

**eFigure 23b. Bubble Plots (follow-up period) on Memory impairment**

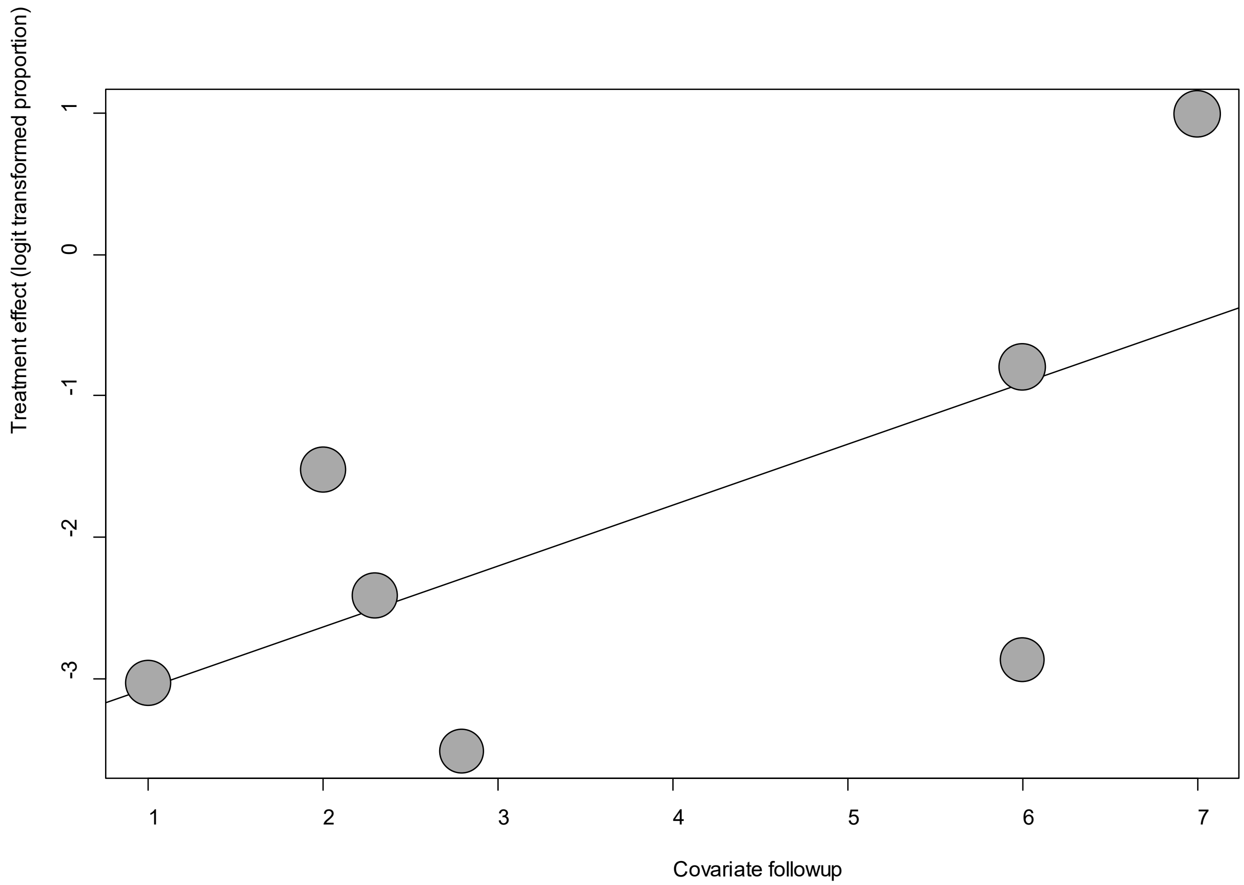


Point sizes are proportional to an inverse of the precision of the estimates.

The regression coefficient was 0.43 (95% confidence interval:0.04-0.82).(*P*=0.03).

**eFigure 23c. Bubble Plots (age) on Memory impairment**

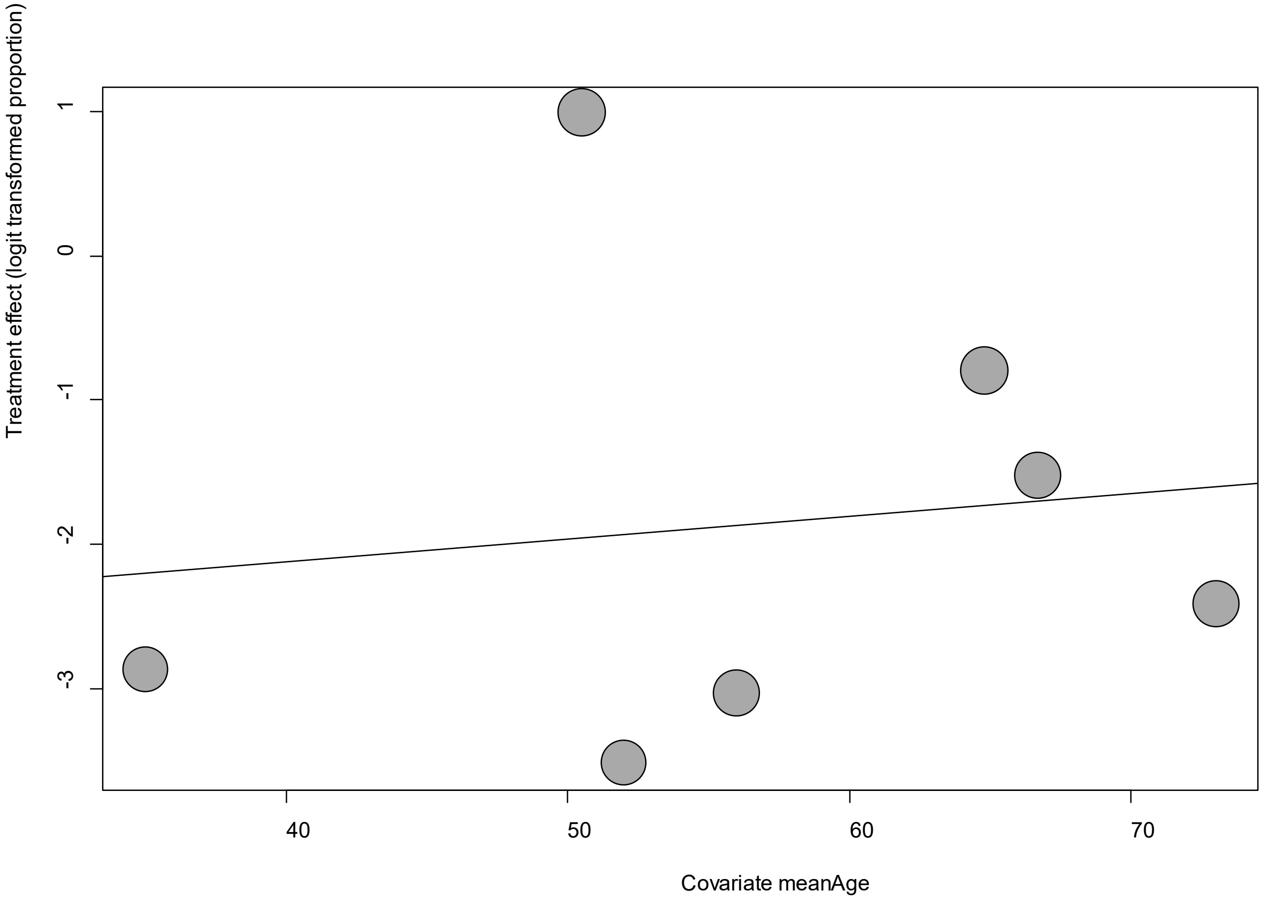


Point sizes are proportional to an inverse of the precision of the estimates.

The regression coefficient was 0.02 (95% confidence interval:-0.12-0.15).(*P*=0.81).

**eFigure 23d. Bubble Plots (sex) on Memory impairment**

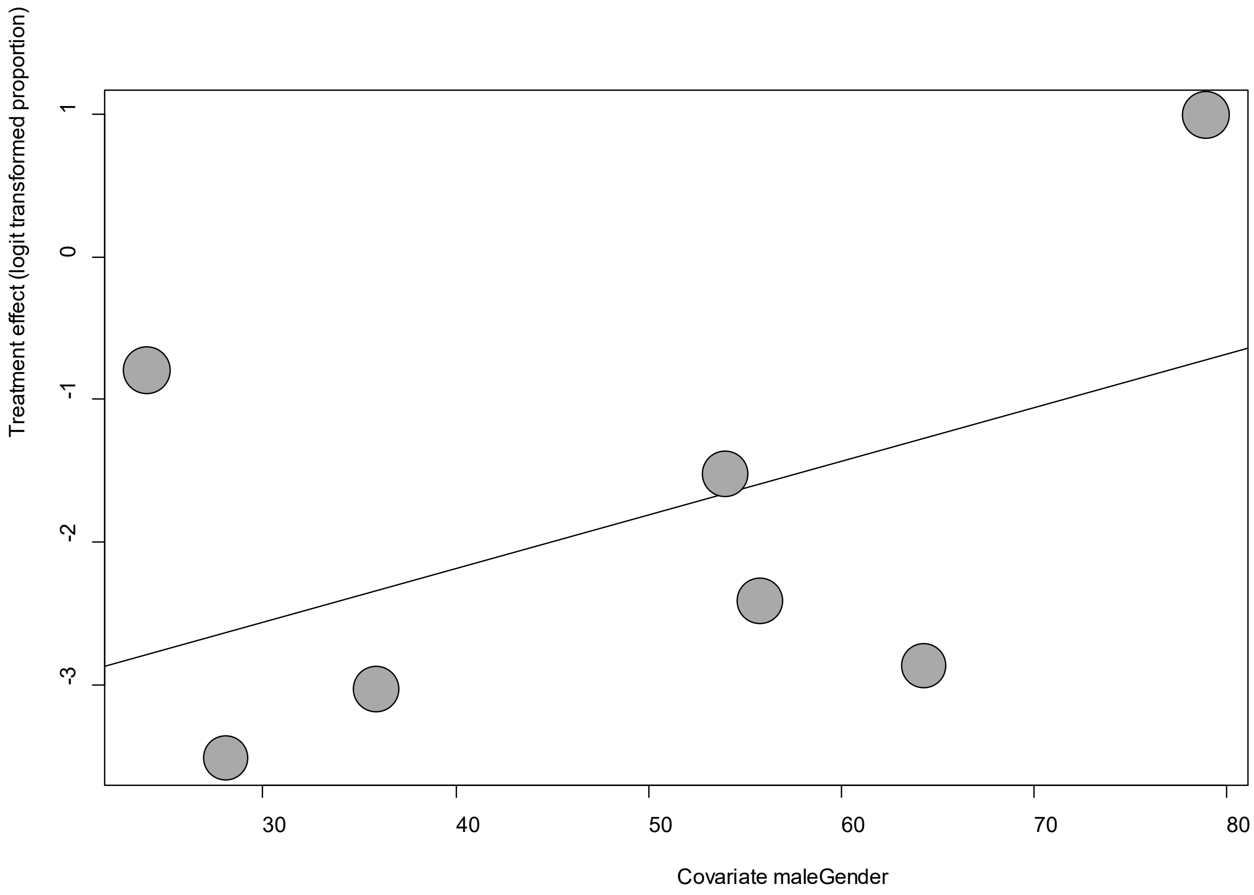


Point sizes are proportional to an inverse of the precision of the estimates.

The regression coefficient was 0.04 (95% confidence interval:-0.04-0.11).(*P*=0.31).

**eFigure 23e. Funnel Plot of studies reporting on Memory impairment**

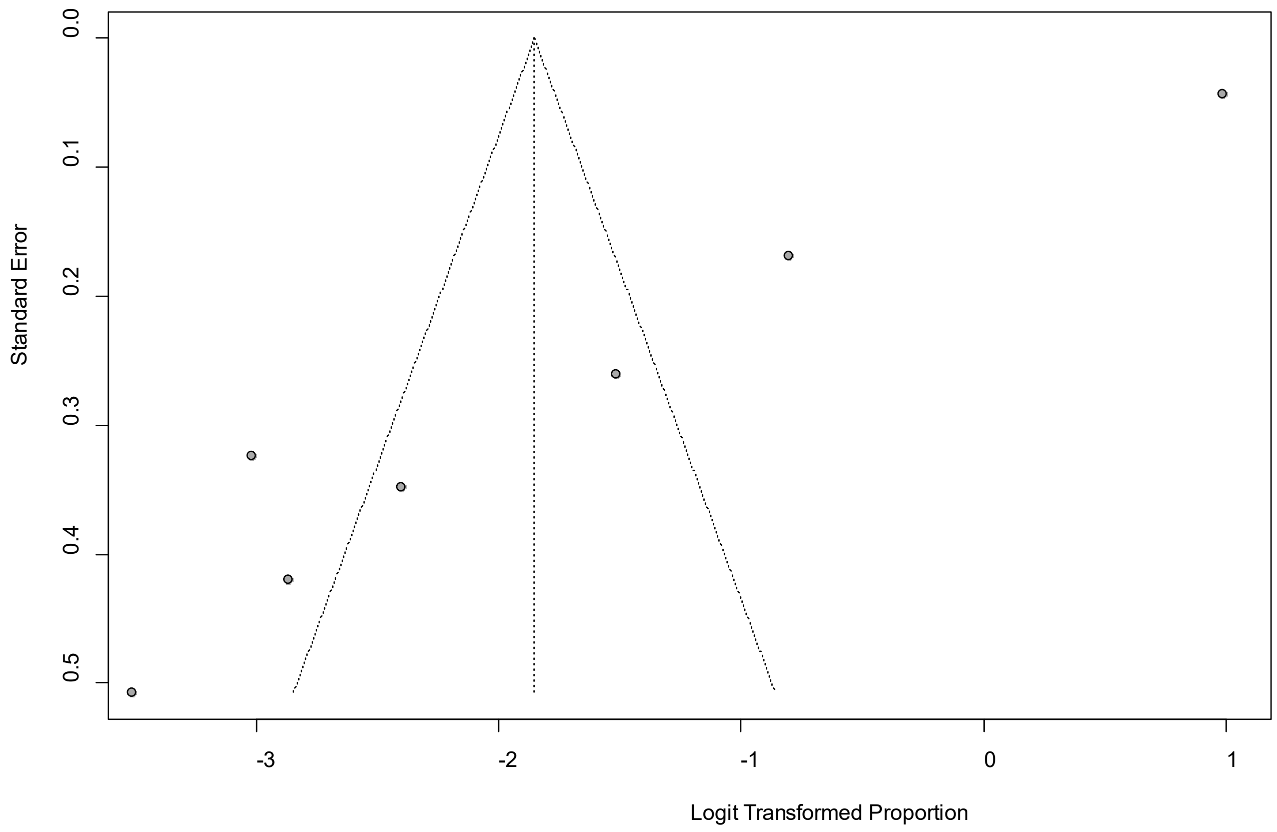


Egger's *P* was <0.0001, indicating the presence of publication bias.

### eFigure 24a. Forest Plot on Nasal blockage

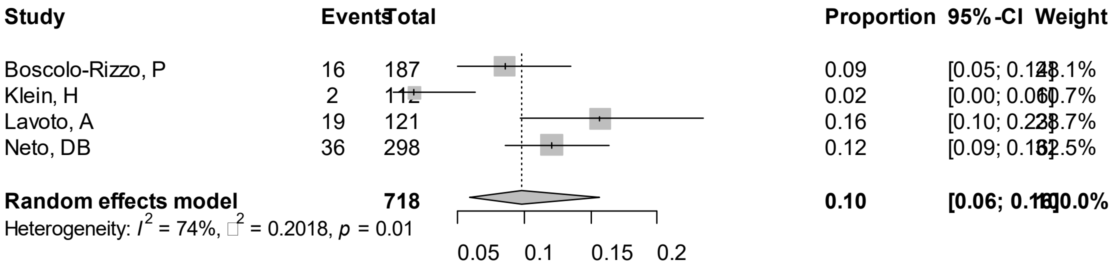


Point sizes are proportional to an inverse of the precision of the estimates.

**eFigure 24b. Bubble Plots (follow-up period) on Nasal blockage**

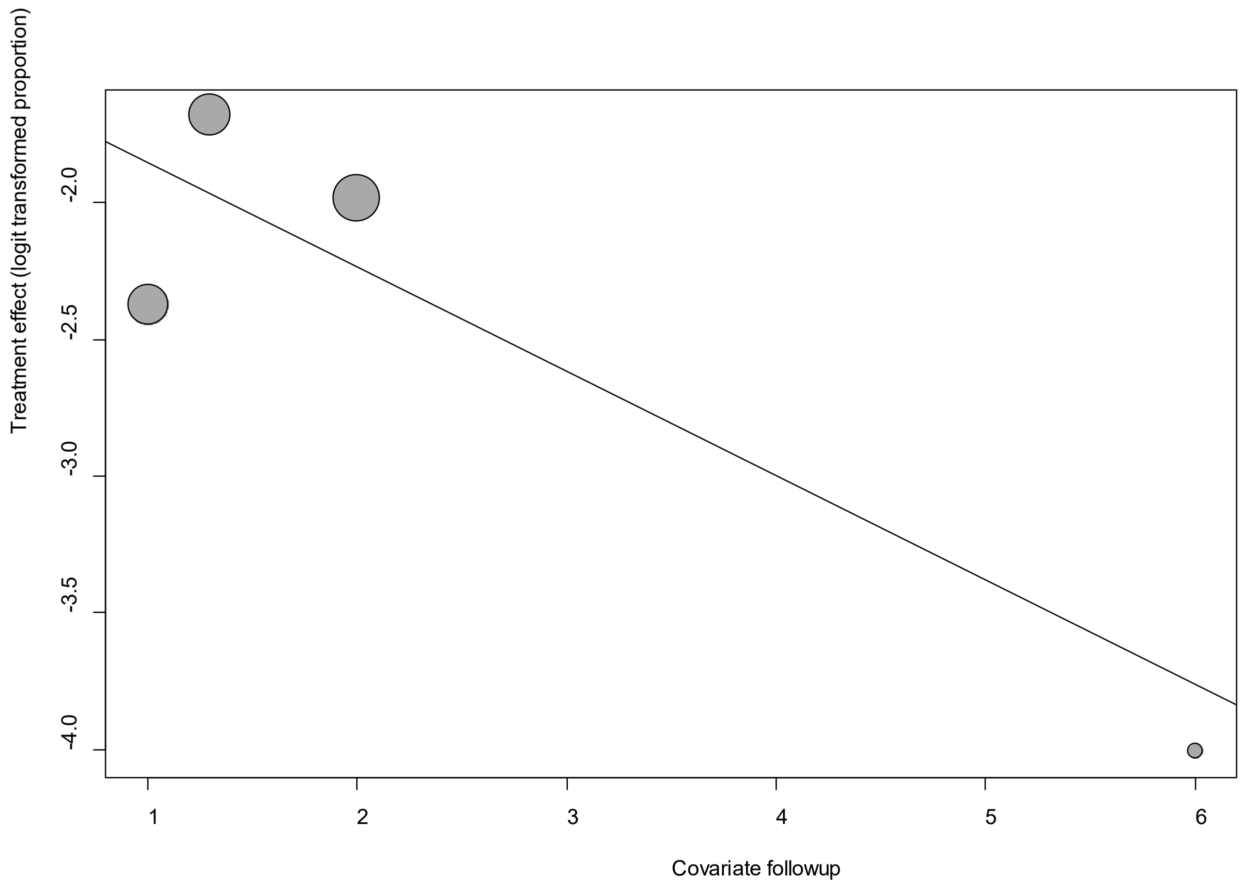


Point sizes are proportional to an inverse of the precision of the estimates.

The regression coefficient was -0.38 (95% confidence interval:-0.73—0.04).(*P*=0.03).

### eFigure 24c. Bubble Plots (age) on Nasal blockage

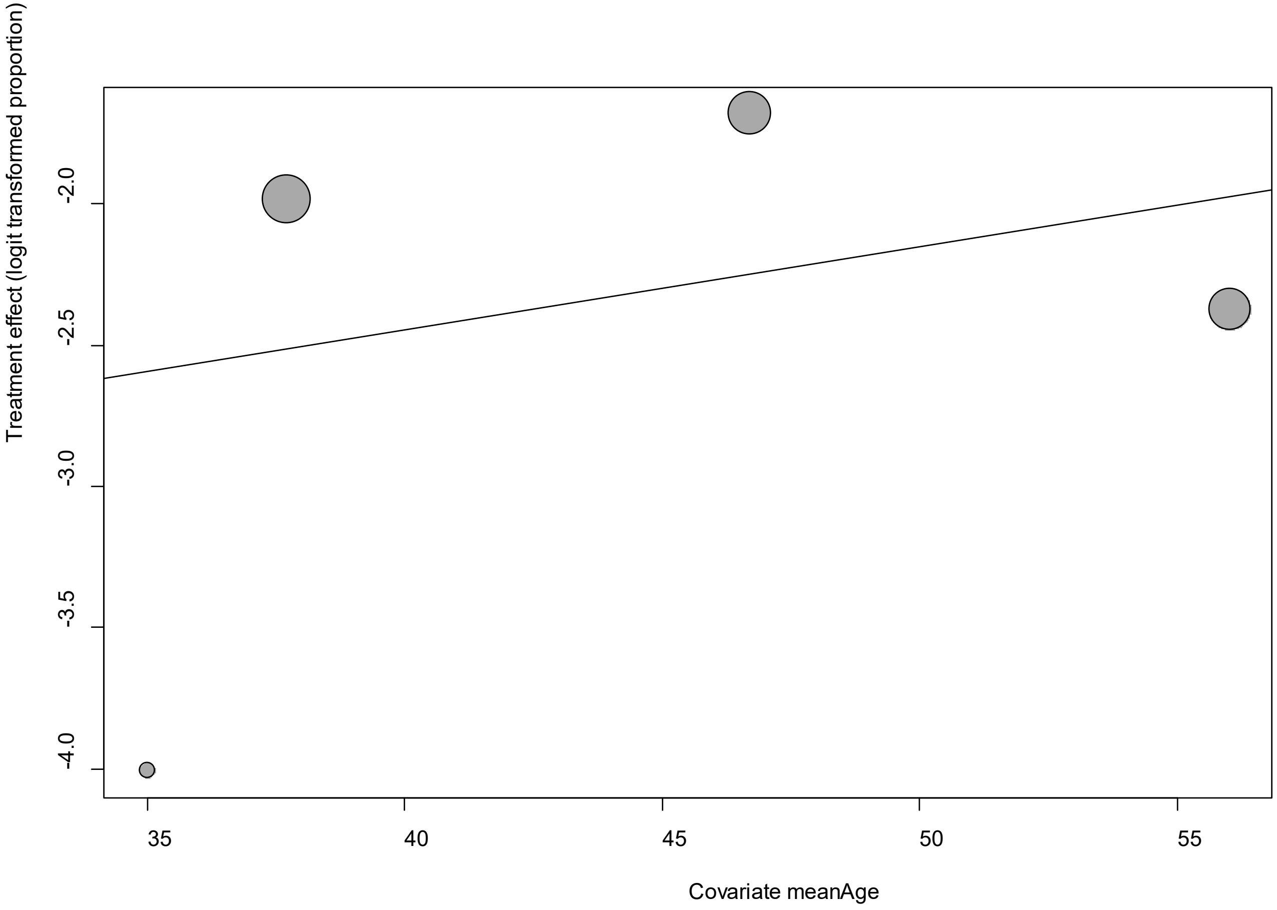


Point sizes are proportional to an inverse of the precision of the estimates.

The regression coefficient was 0.03 (95% confidence interval:-0.06-0.12).(*P*=0.52).

**eFigure 24d. Bubble Plots (sex) on Nasal blockage**

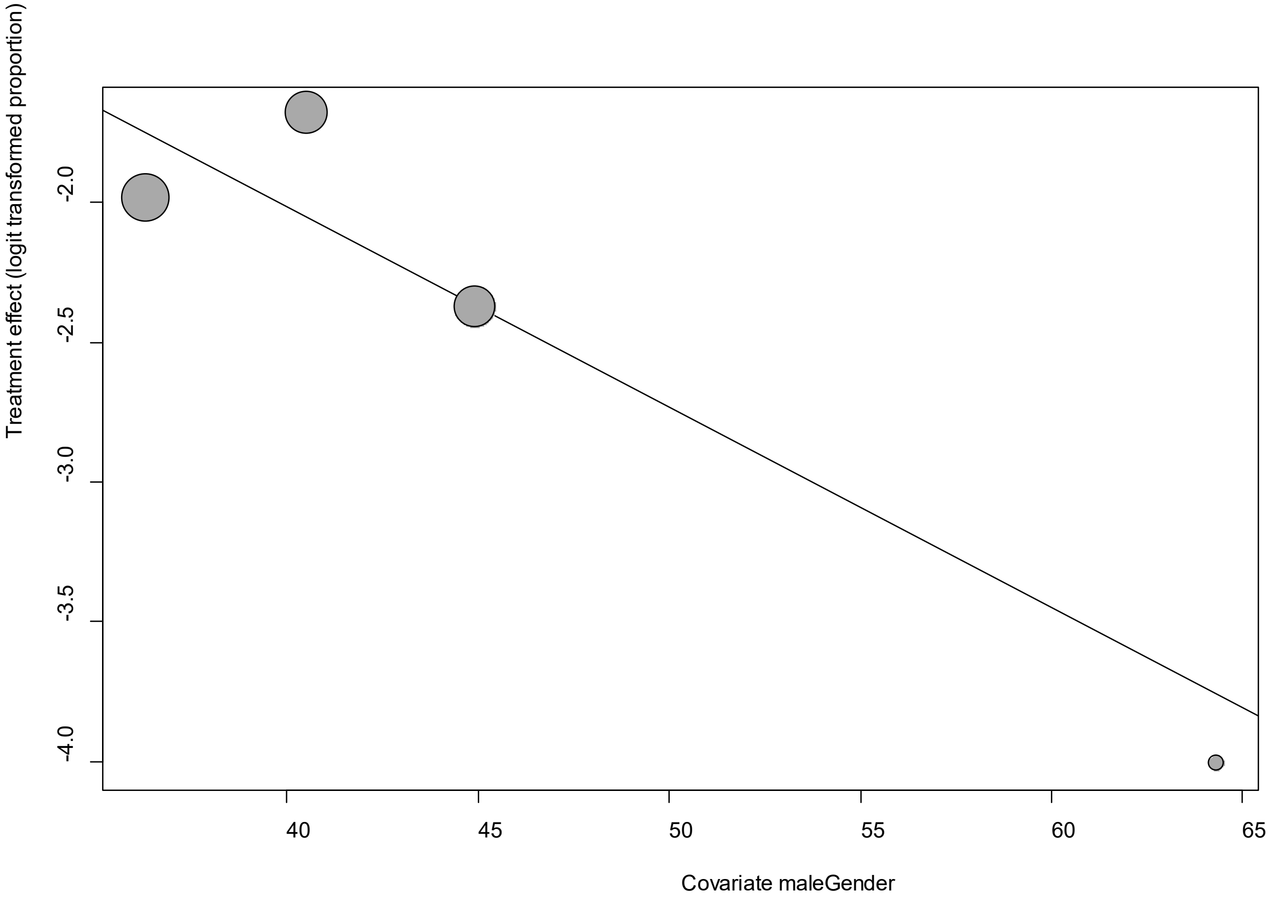


Point sizes are proportional to an inverse of the precision of the estimates.

The regression coefficient was -0.07 (95% confidence interval:-0.13—0.02).(*P*=0.0098).

**eFigure 24e. Funnel Plot of studies reporting on Nasal blockage**

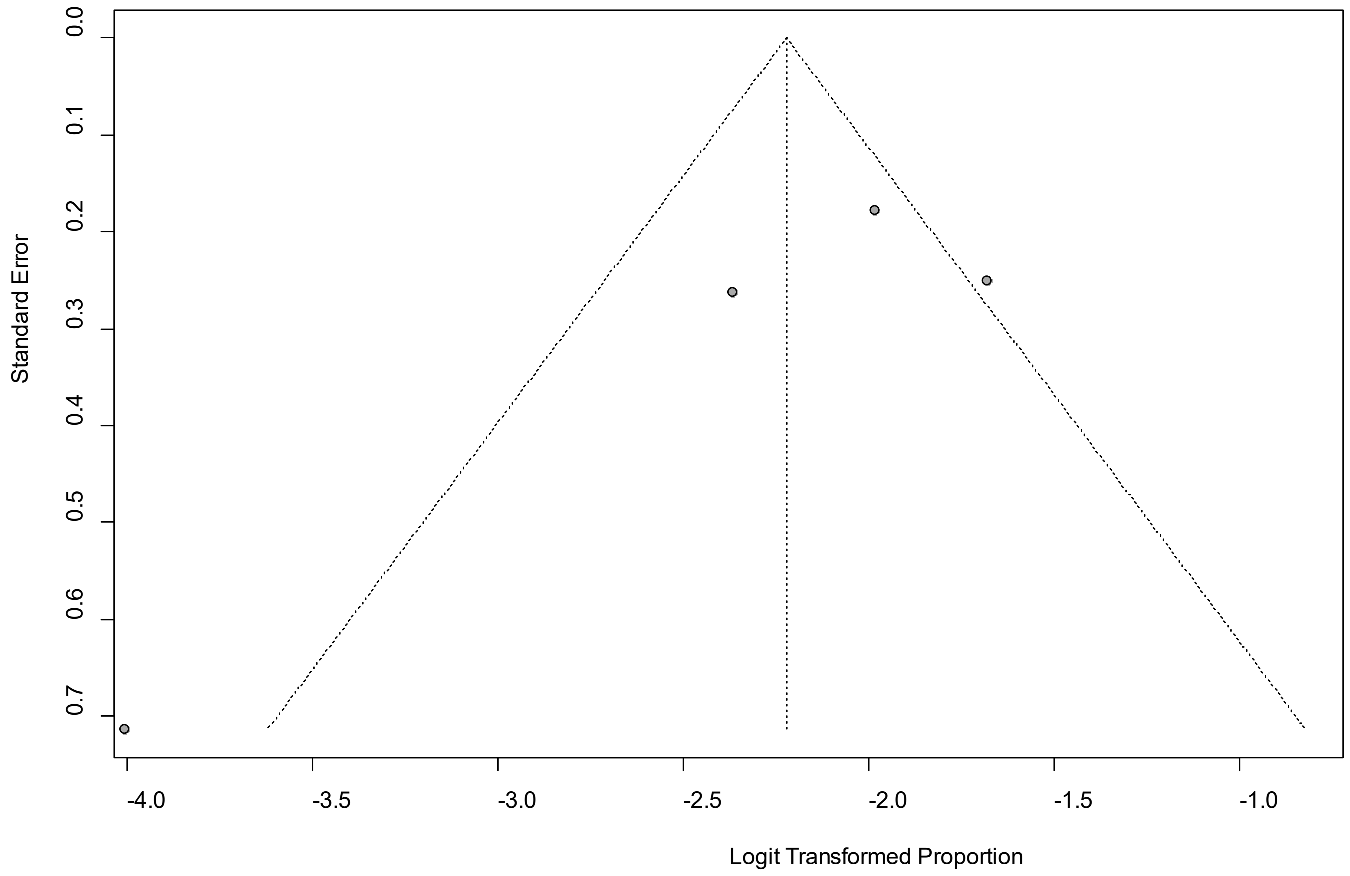


Egger's *P* was 0.21, indicating the absence of publication bias.

**eFigure 25a. Forest Plot on Nausea**

| Study | Events | Total | Proportion | 95%-CI | Weight |
|---|---|---|---|---|---|
| Arnold, D | 0 | [illegible] | 0.00 | [illegible] | [illegible] |
| Boscolo-Rizzo, P | 1 | [illegible] | 0.01 | [illegible] | [illegible] |
| Cirulli, E | 5 | [illegible] | 0.02 | [illegible] | [illegible] |
| Davis, HE | 1825 | 3762 | 0.49 | [0.47; 0.50] | 10.7% |
| Geortz, YNJ | 254 | 2113 | 0.12 | [0.11; 0.13] | 10.7% |
| Moradian, ST | 3 | [illegible] | 0.01 | [illegible] | [illegible] |
| Petersen, M | 7 | [illegible] | 0.04 | [0.02; 0.08] | 10.1% |
| Stavem, K | 8 | [illegible] | 0.02 | [0.01; 0.03] | 10.2% |
| Tenforde, MW | 27 | 270 | 0.10 | [0.07; 0.14] | 10.5% |
| Wang, X | 1 | [illegible] | 0.01 | [illegible] | [illegible] |
| Yan, N | 1 | [illegible] | 0.00 | [illegible] | [illegible] |
| **Random effects model** | | **[illegible]** | **0.03** | **[0.01; 0.08]** | **100.0%** |

Heterogeneity: $I^2 = 99\%$, $\tau^2 = 2.3786$, $p < 0.01$

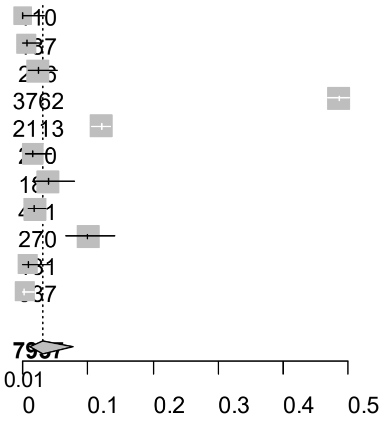


Point sizes are proportional to an inverse of the precision of the estimates.

### eFigure 25b. Bubble Plots (follow-up period) on Nausea

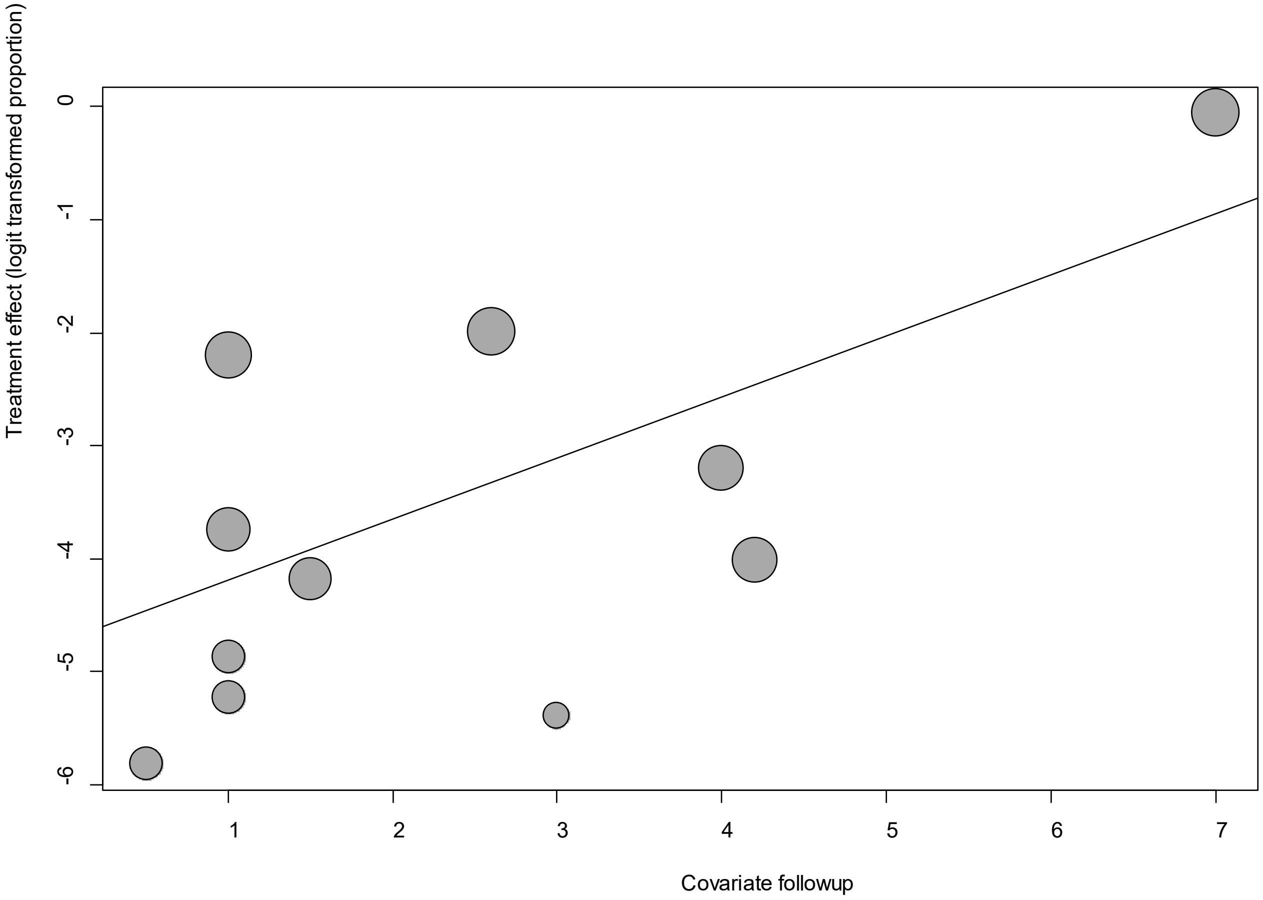


Point sizes are proportional to an inverse of the precision of the estimates.

The regression coefficient was 0.54 (95% confidence interval:0.19-0.89).(*P*=0.003).

**eFigure 25c. Bubble Plots (age) on Nausea**

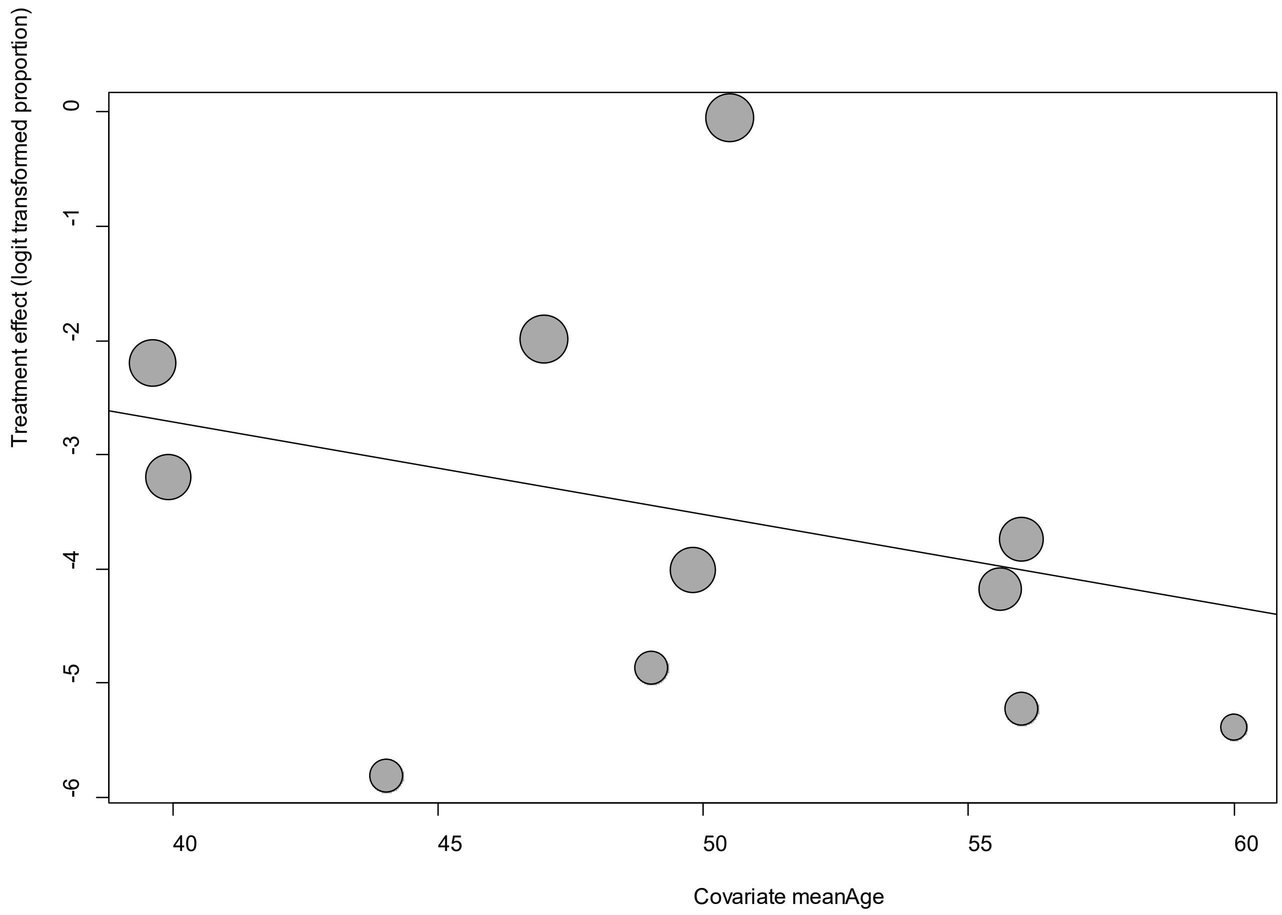


Point sizes are proportional to an inverse of the precision of the estimates.

The regression coefficient was -0.08 (95% confidence interval:-0.24-0.08).(*P*=0.31).

**eFigure 25d. Bubble Plots (sex) on Nausea**

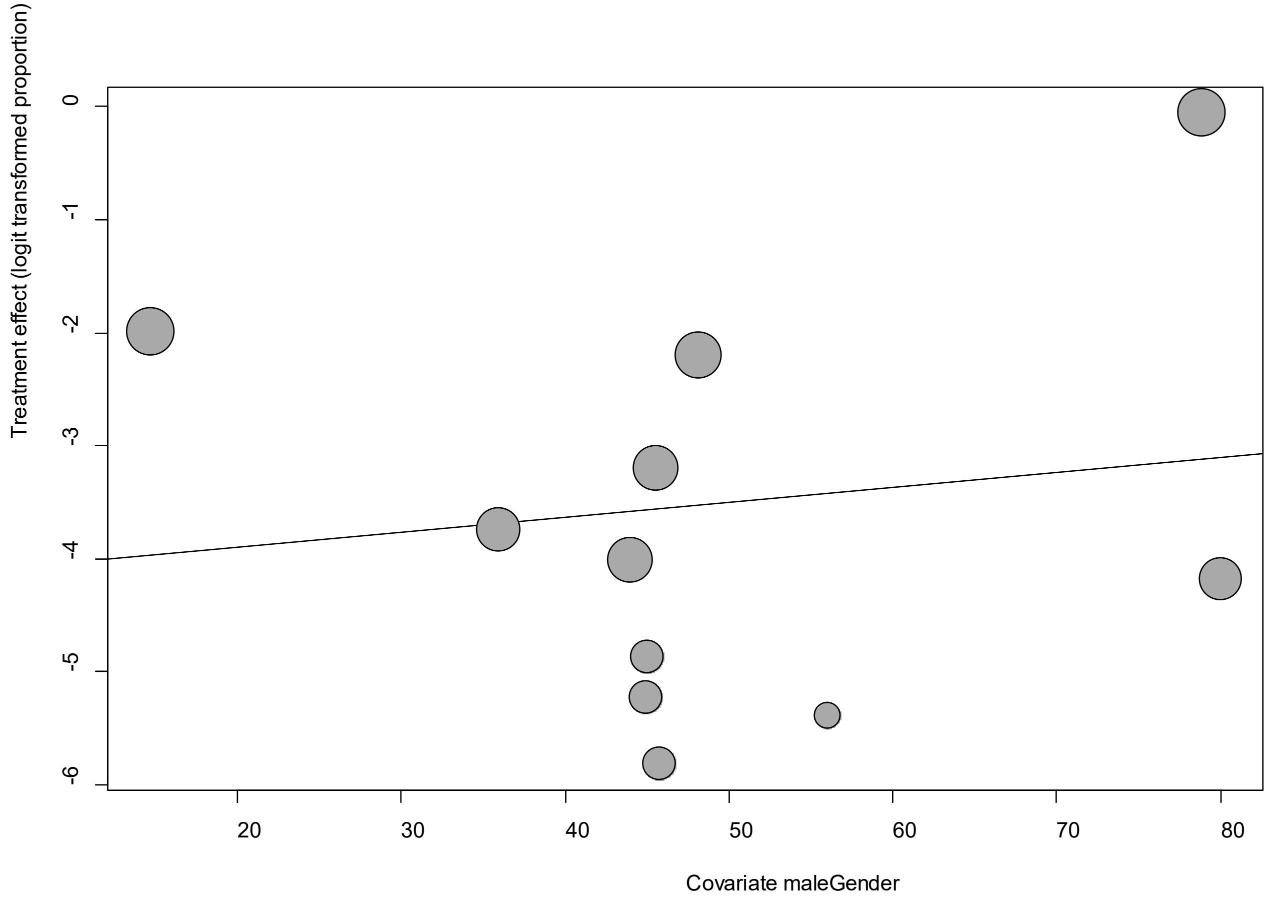


Point sizes are proportional to an inverse of the precision of the estimates.

The regression coefficient was 0.01 (95% confidence interval:-0.05-0.08).(*P*=0.68).

**eFigure 25e. Funnel Plot of studies reporting on Nausea**

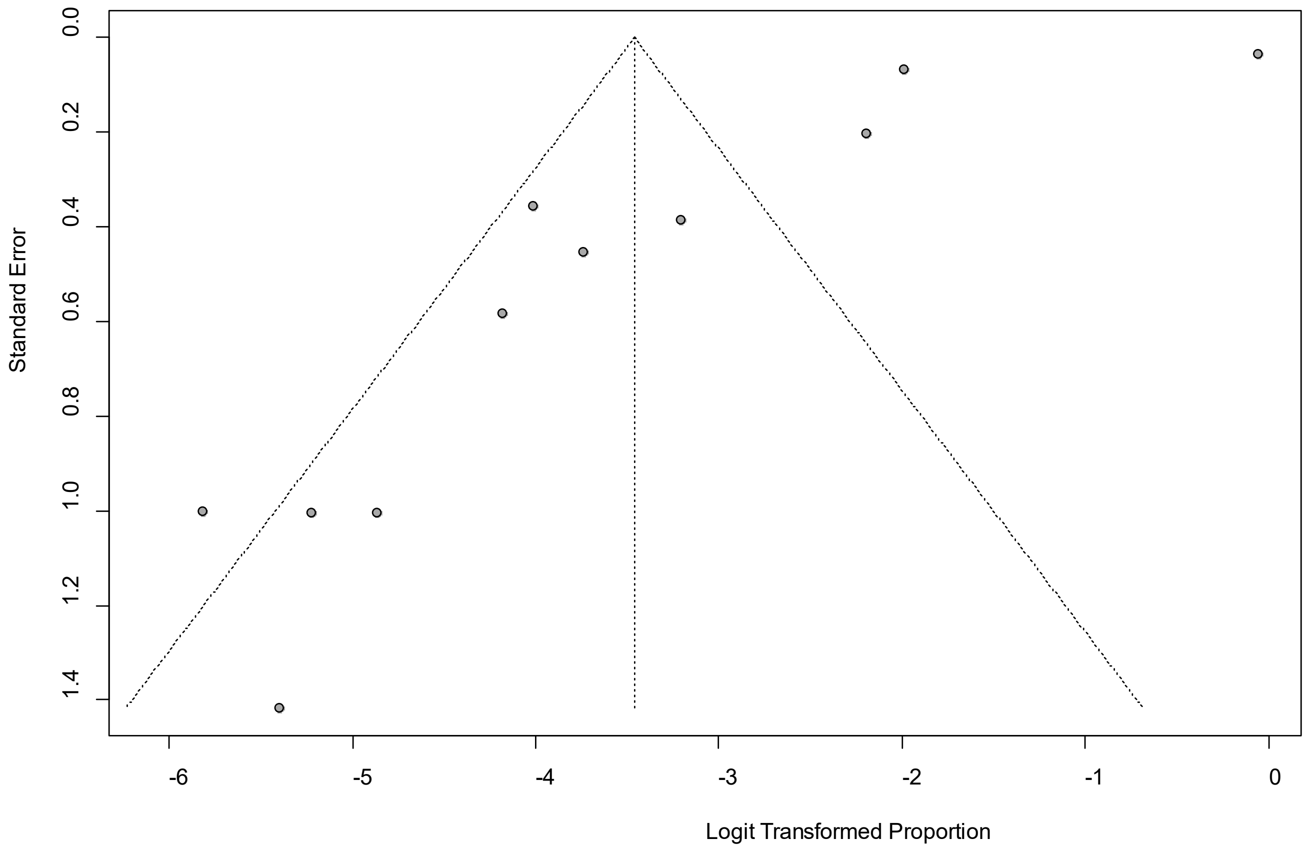


Egger's *P* was 0.018, indicating the presence of publication bias.

### eFigure 26a. Forest Plot on Palpitation

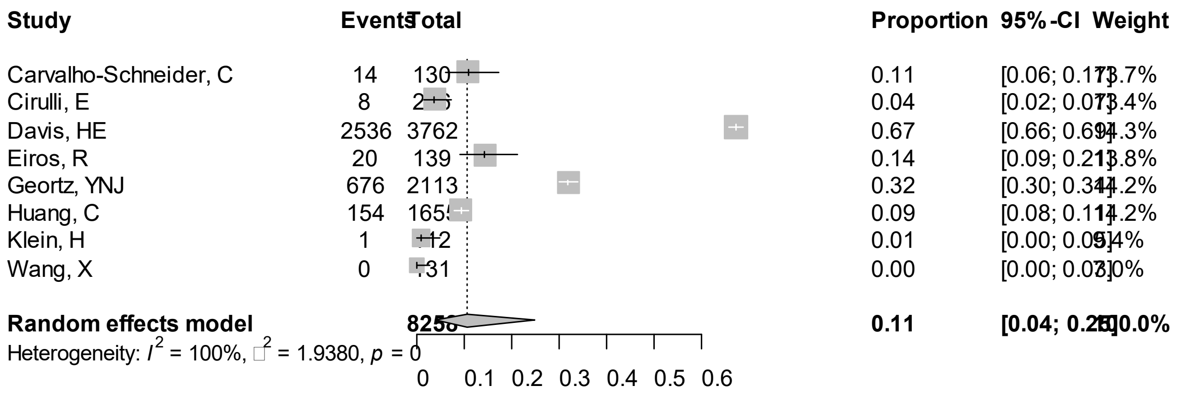


Point sizes are proportional to an inverse of the precision of the estimates.

**eFigure 26b. Bubble Plots (follow-up period) on Palpitation**

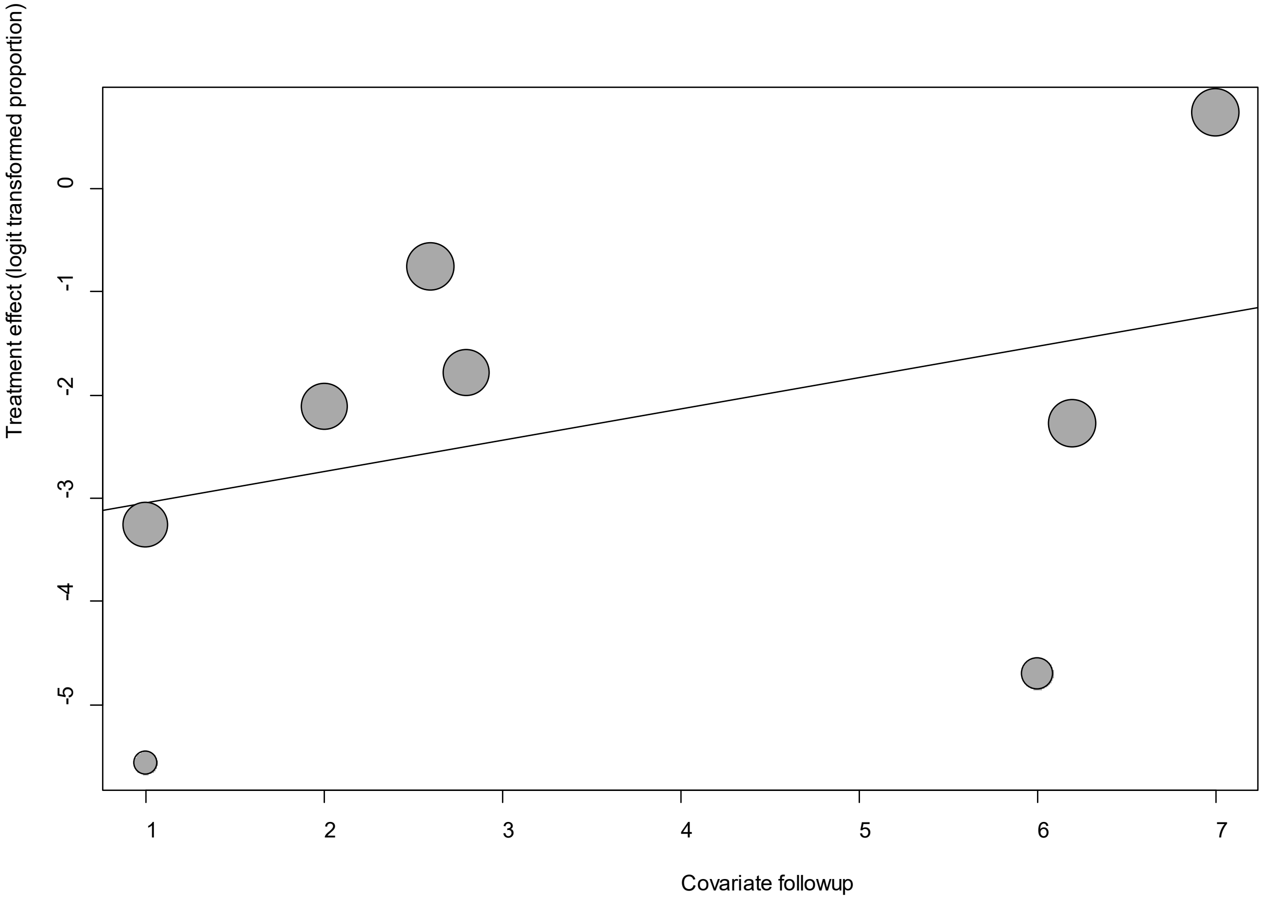


Point sizes are proportional to an inverse of the precision of the estimates.

The regression coefficient was 0.30 (95% confidence interval:-0.29-0.89).(*P*=0.32).

### eFigure 26c. Bubble Plots (age) on Palpitation

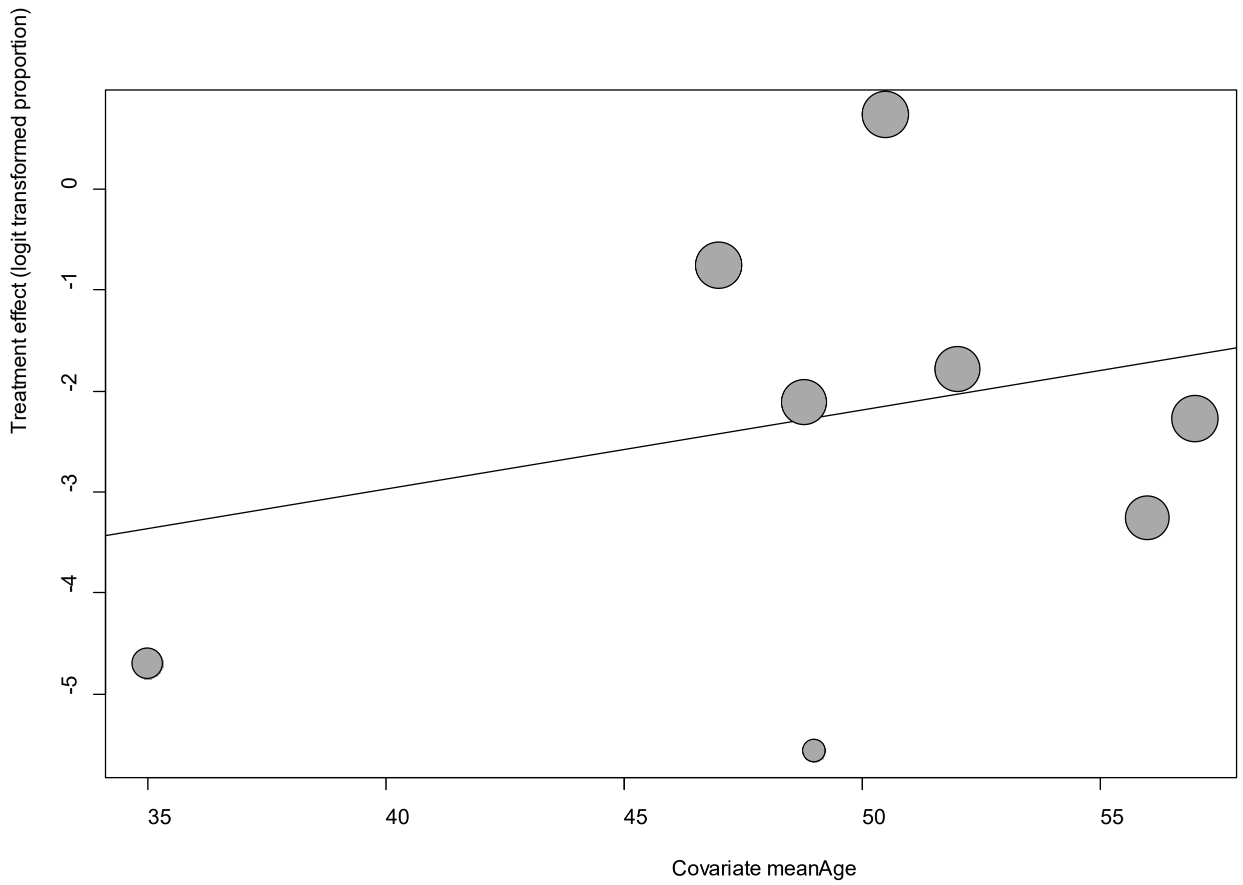


Point sizes are proportional to an inverse of the precision of the estimates.

The regression coefficient was 0.08 (95% confidence interval:-0.02-0.28).(*P*=0.44).

**eFigure 26d. Bubble Plots (sex) on Palpitation**

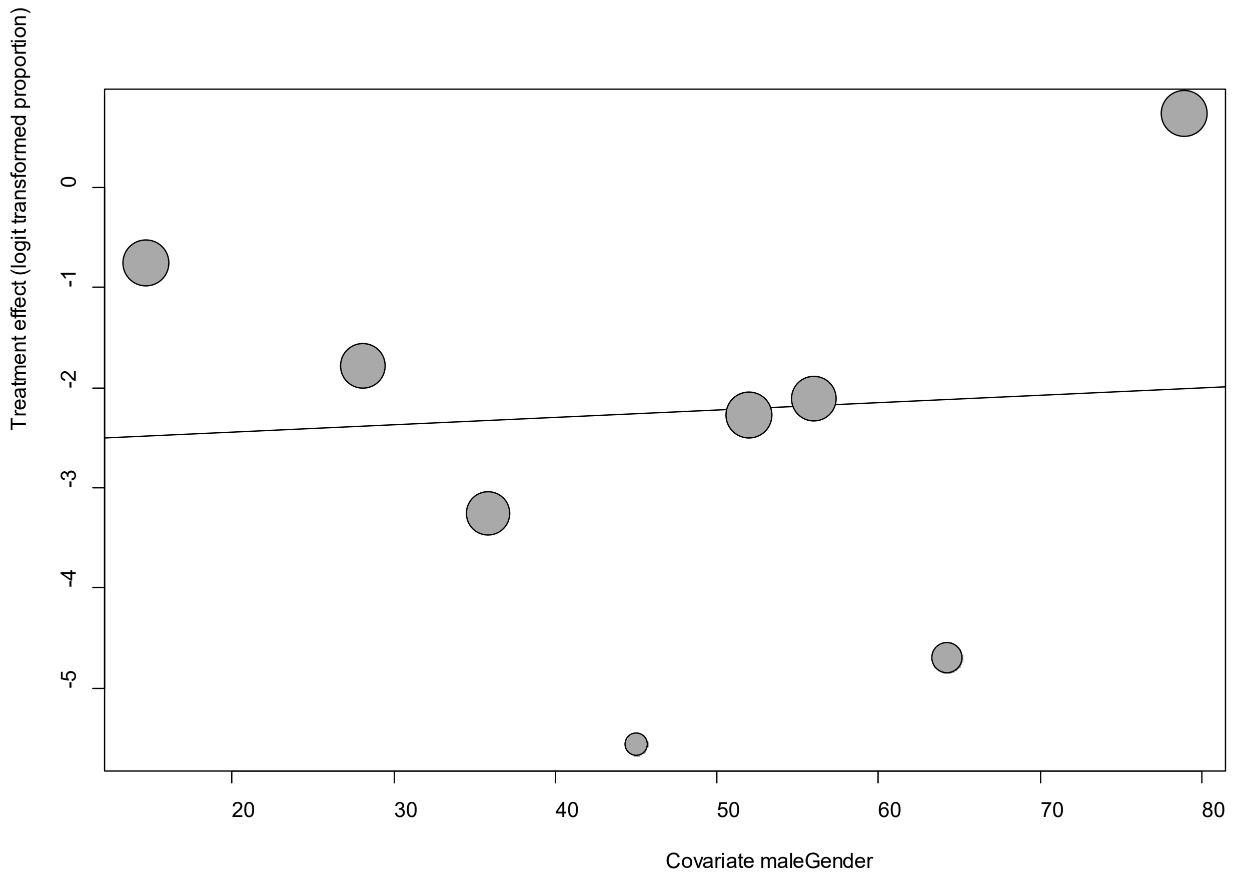


Point sizes are proportional to an inverse of the precision of the estimates.

The regression coefficient was -0.007 (95% confidence interval:-0.06-0.08).(*P*=0.84).

**eFigure 26e. Funnel Plot of studies reporting on Palpitation**

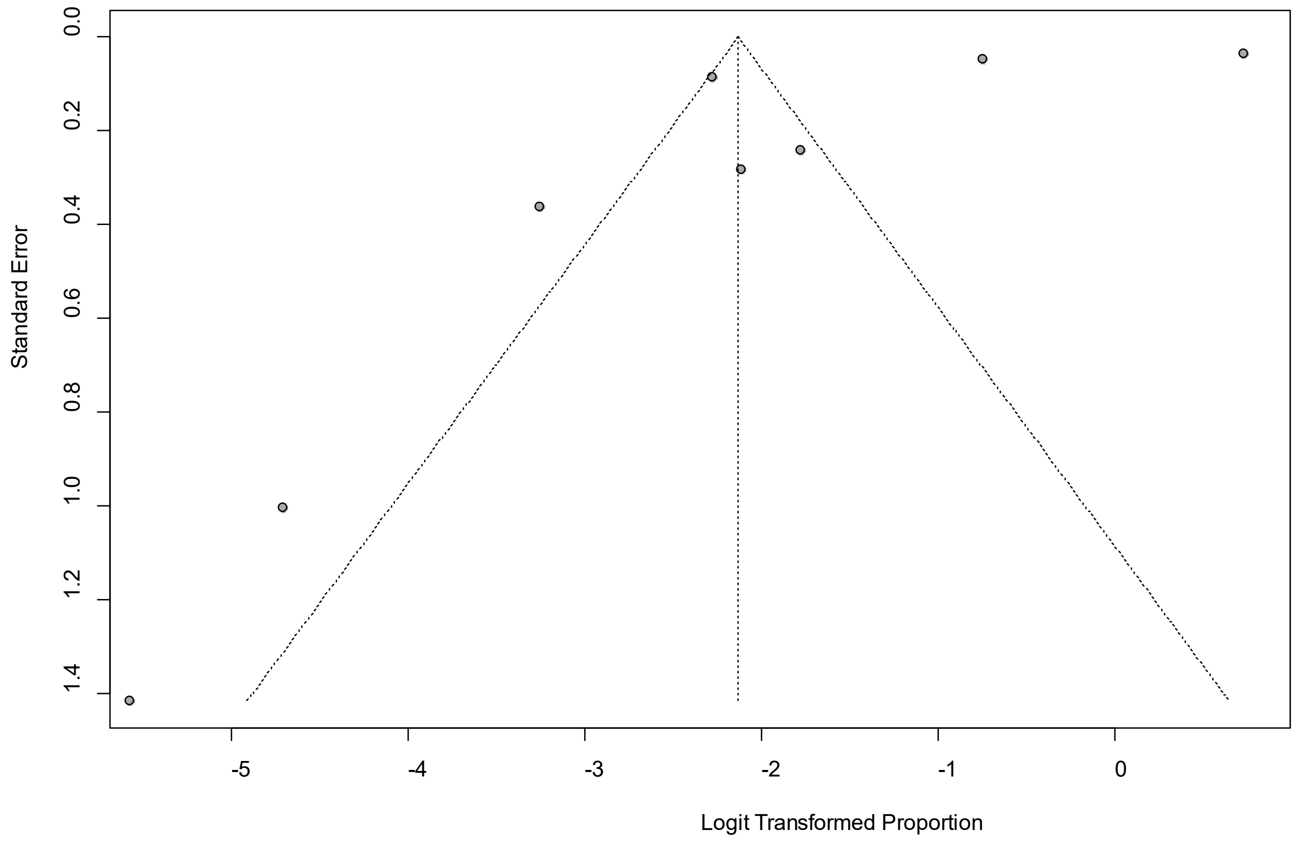


Egger's *P* was 0.15, indicating the presence of publication bias.

### eFigure 27a. Forest Plot on Rhinorrhea

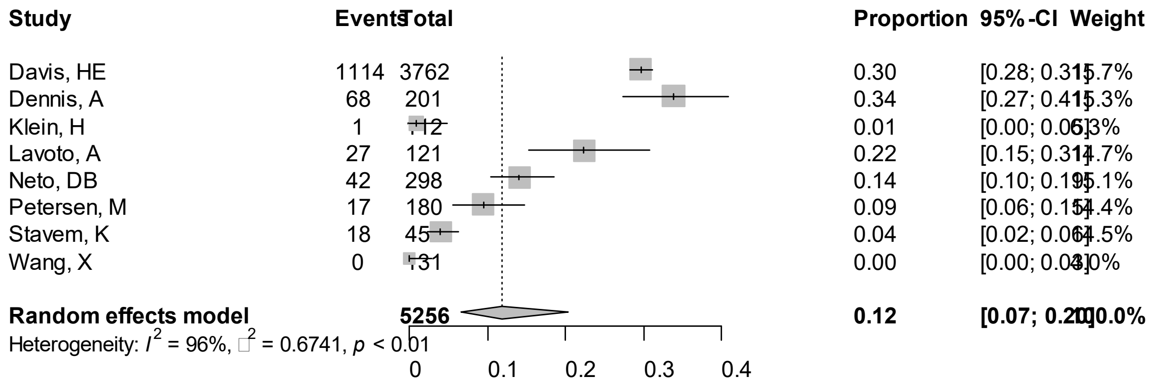


Point sizes are proportional to an inverse of the precision of the estimates.

### eFigure 27b. Bubble Plots (follow-up period) on Rhinorrhea

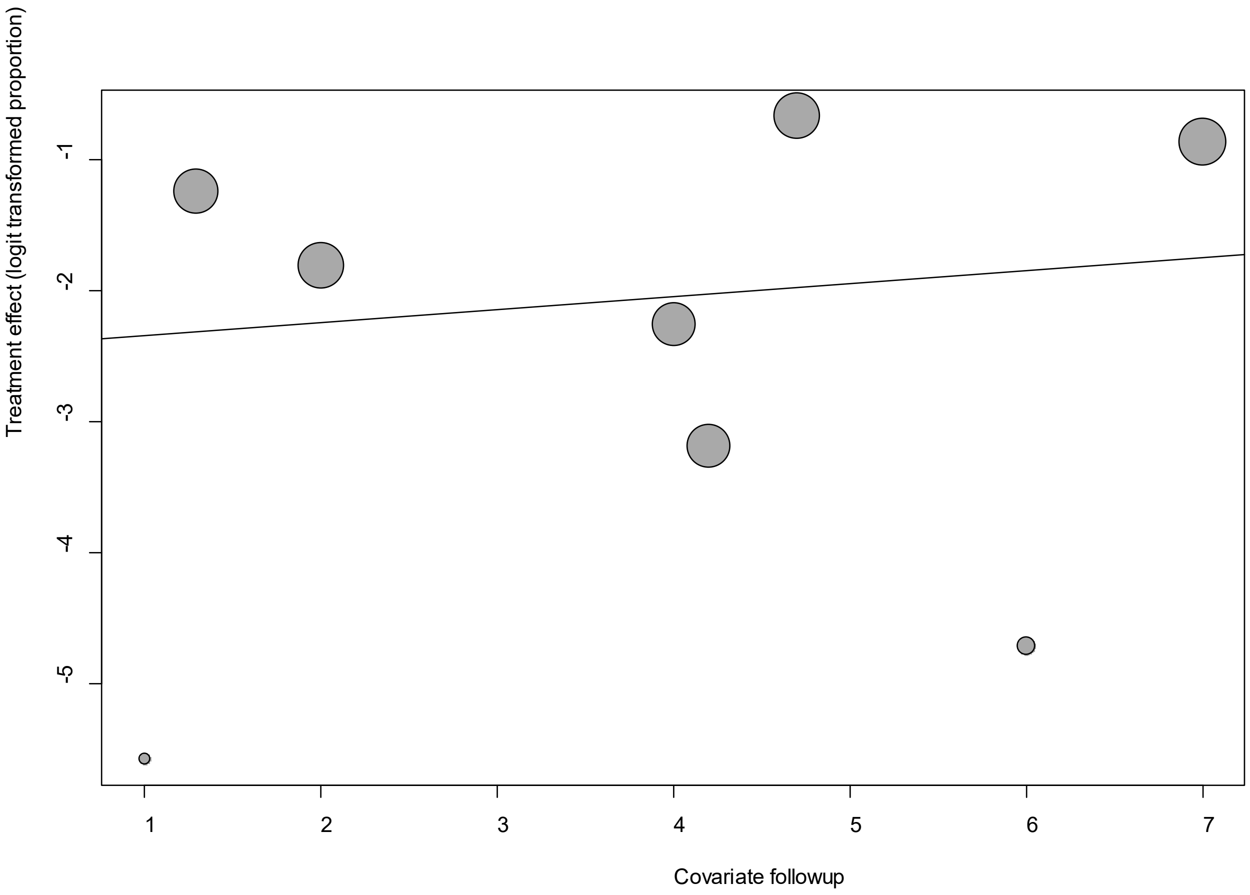


Point sizes are proportional to an inverse of the precision of the estimates.

The regression coefficient was 0.10 (95% confidence interval:-0.27-0.47).(*P*=0.59).

### eFigure 27c. Bubble Plots (age) on Rhinorrhea

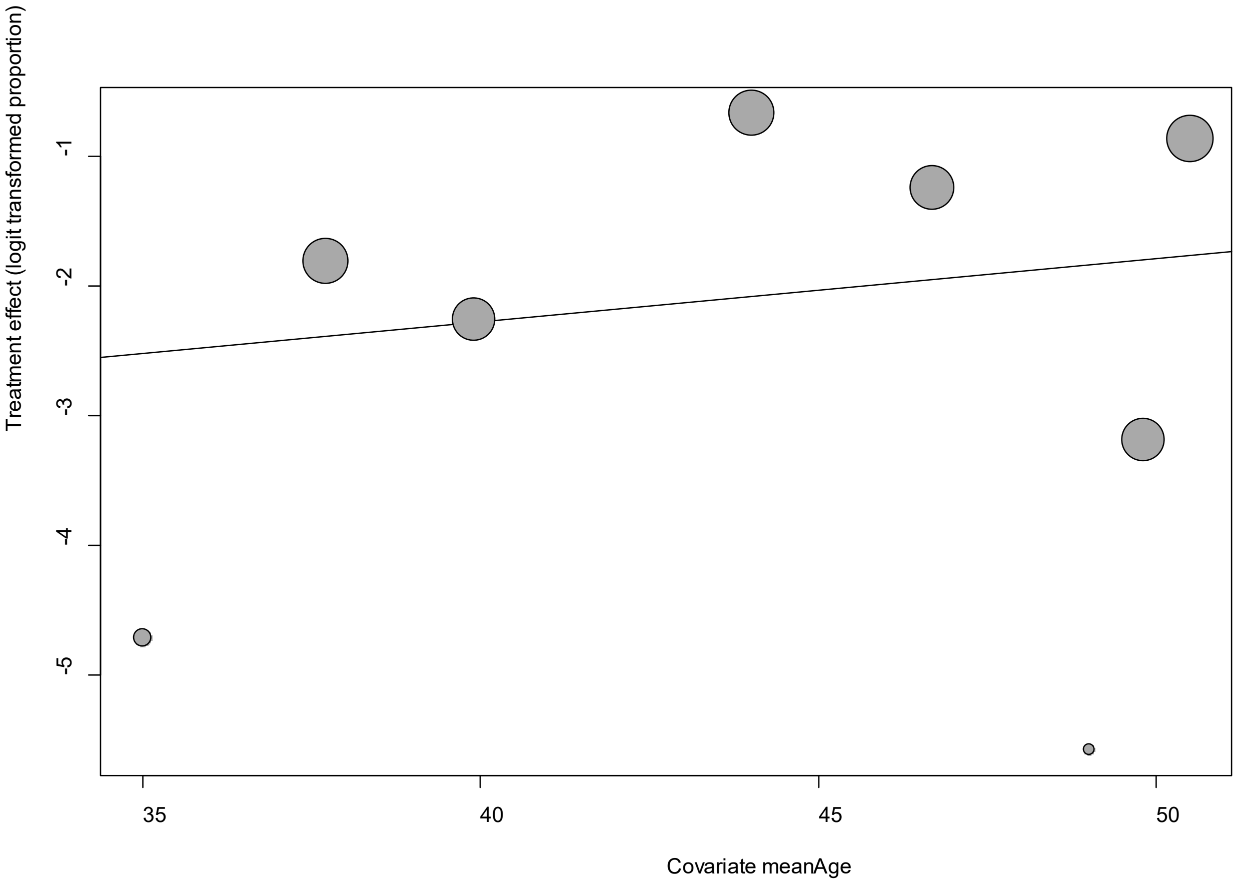


Point sizes are proportional to an inverse of the precision of the estimates.

The regression coefficient was 0.05 (95% confidence interval:-0.1-0.19).(*P*=0.50).

**eFigure 27d. Bubble Plots (sex) on Rhinorrhea**

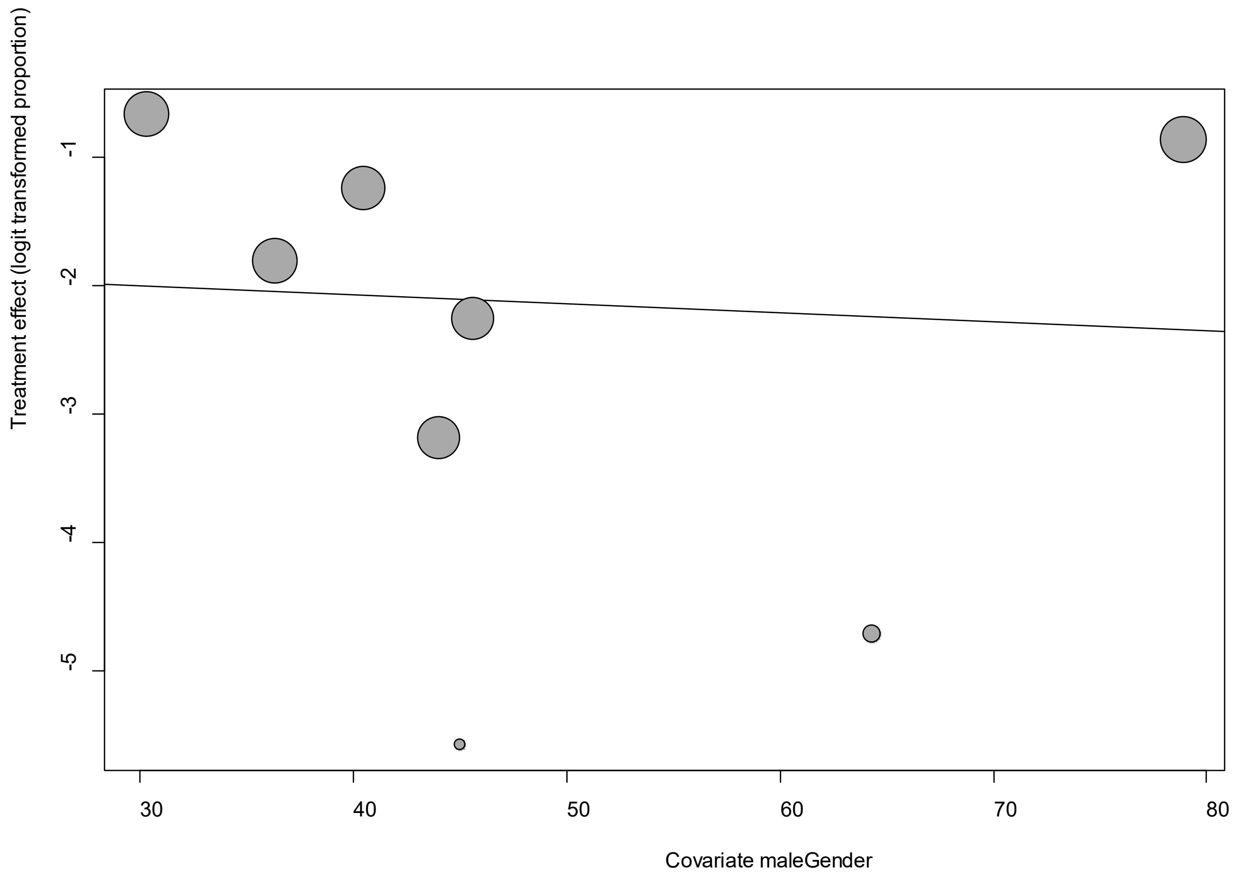


Point sizes are proportional to an inverse of the precision of the estimates.

The regression coefficient was -0.007 (95% confidence interval:-0.06-0.05).(*P*=0.80).

**eFigure 27e. Funnel Plot of studies reporting on Rhinorrhea**

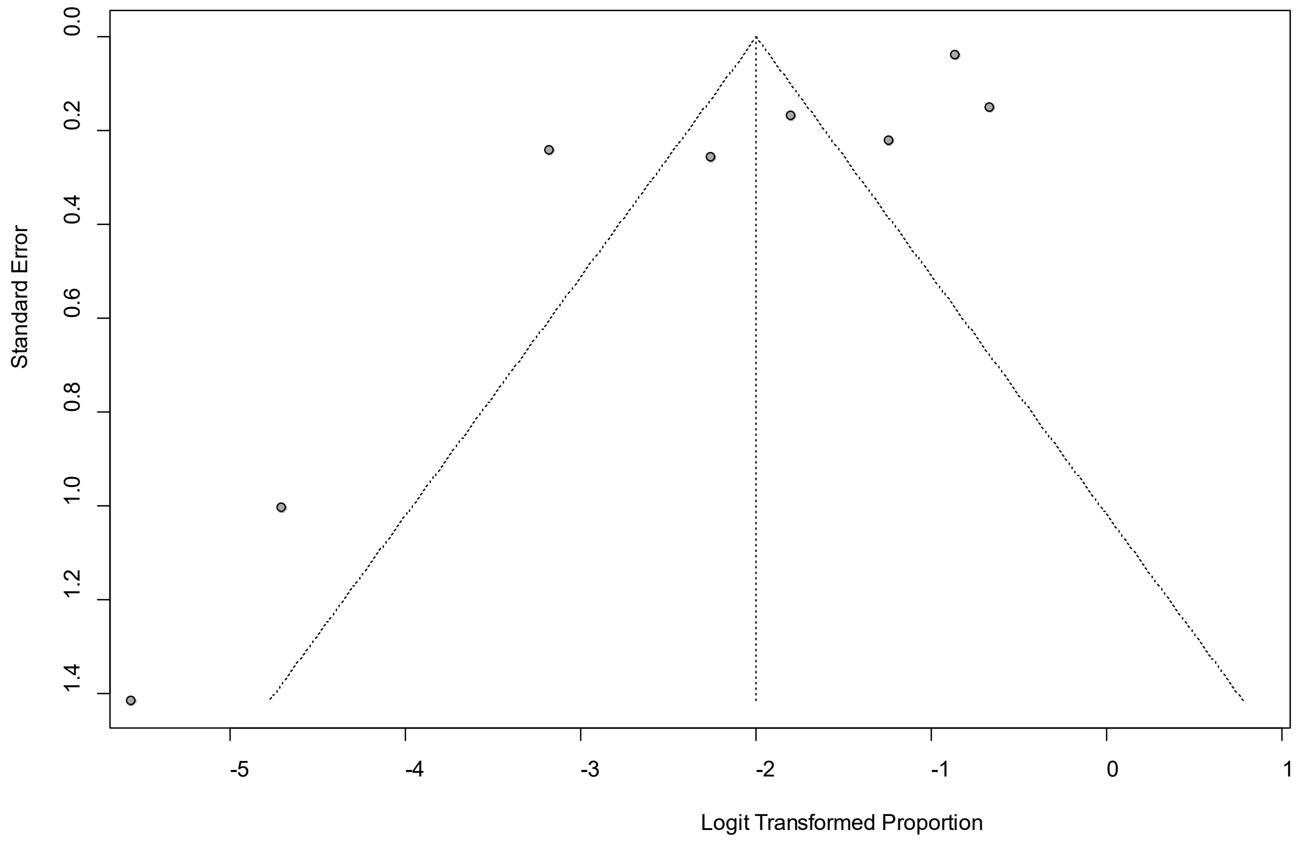


Egger's *P* was 0.02, indicating the presence of publication bias.

### eFigure 28a. Forest Plot on Sneezing

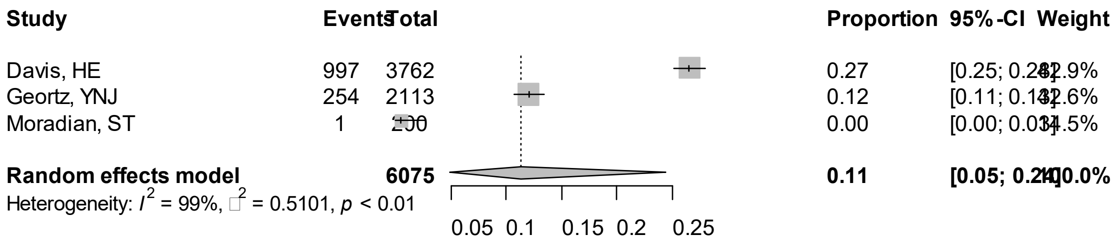


Point sizes are proportional to an inverse of the precision of the estimates.

**eFigure 28b. Bubble Plots (follow-up period) on Sneezing**

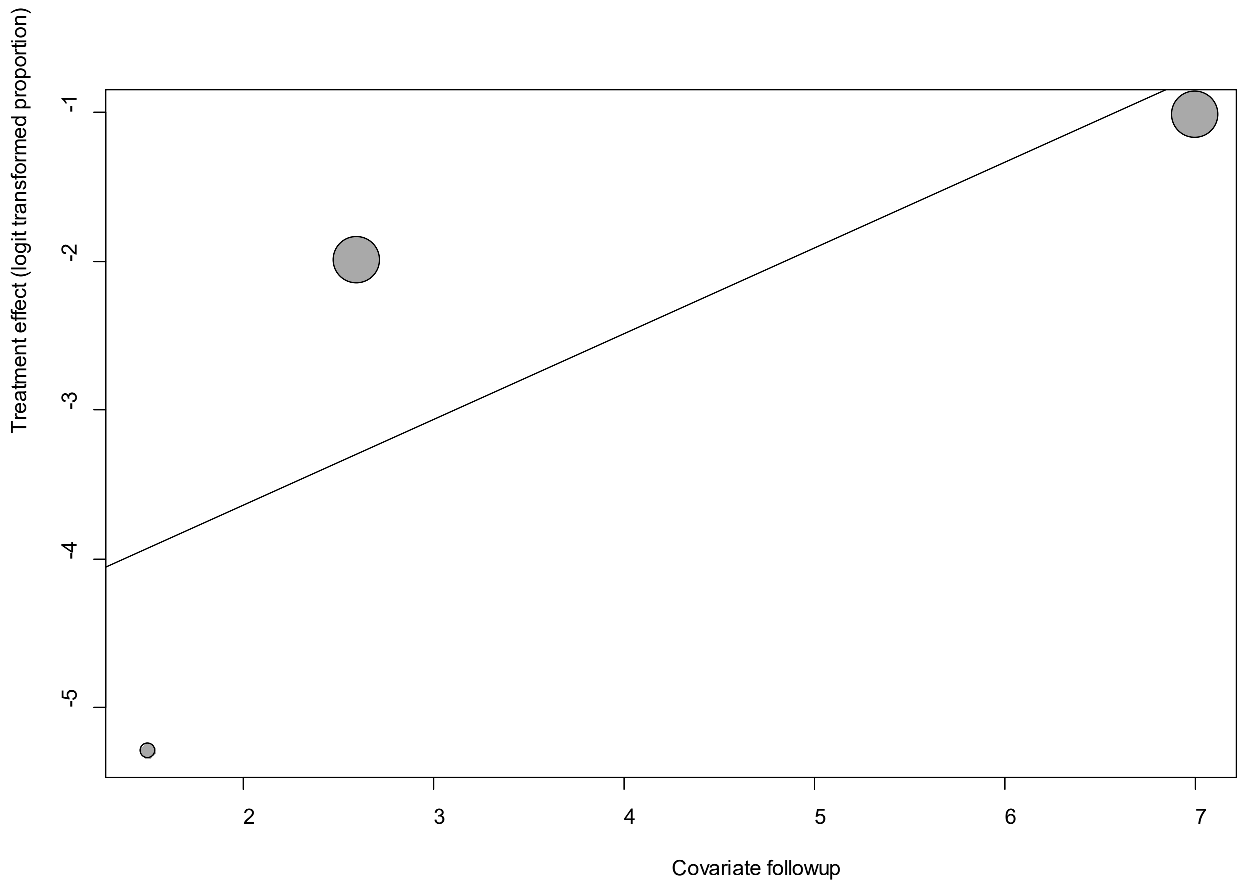


Point sizes are proportional to an inverse of the precision of the estimates.

The regression coefficient was 0.58 (95% confidence interval:-0.31-1.46).(*P*=0.20).

**eFigure 28c. Bubble Plots (age) on Sneezing**

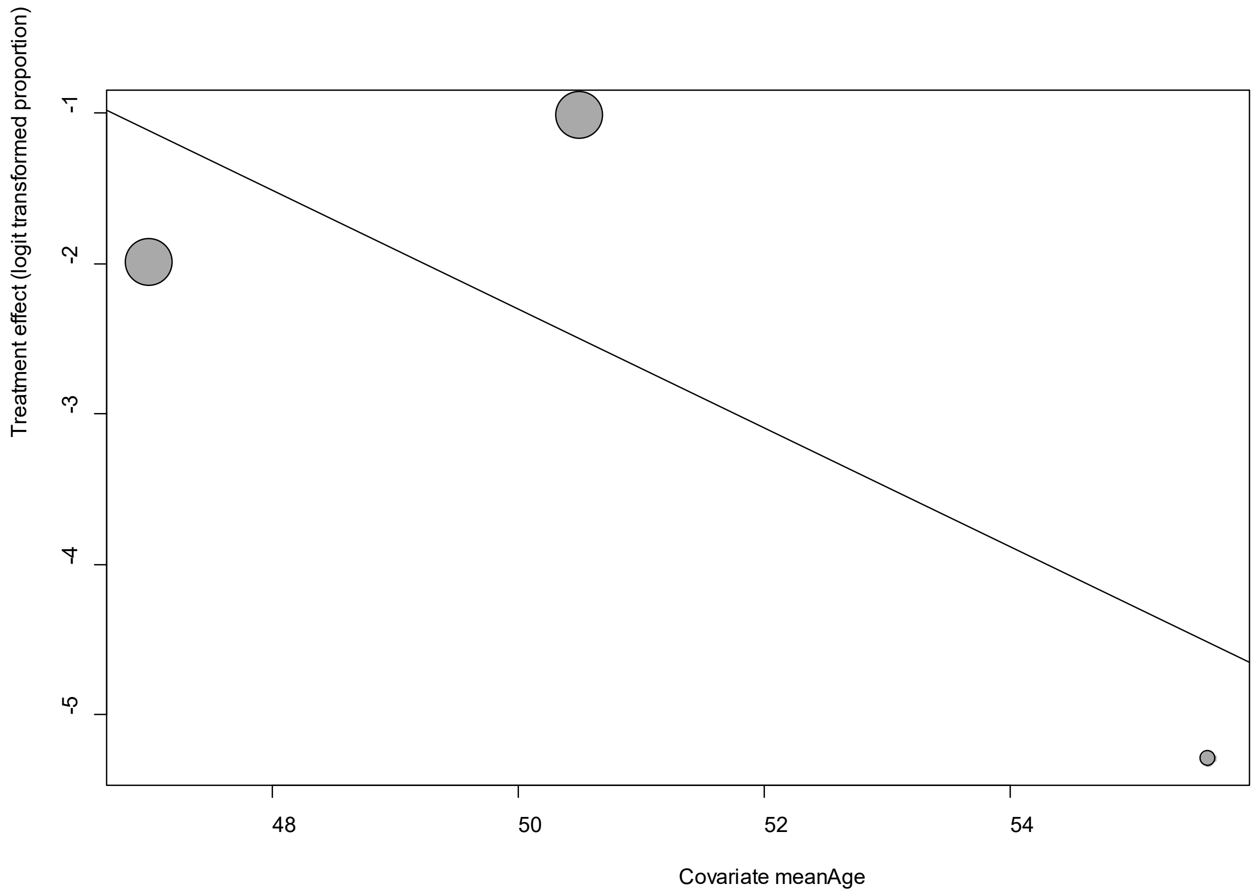


Point sizes are proportional to an inverse of the precision of the estimates.

The regression coefficient was -0.40 (95% confidence interval:-1.03-0.24).(*P*=0.22).

**eFigure 28d. Bubble Plots (sex) on Sneezing**

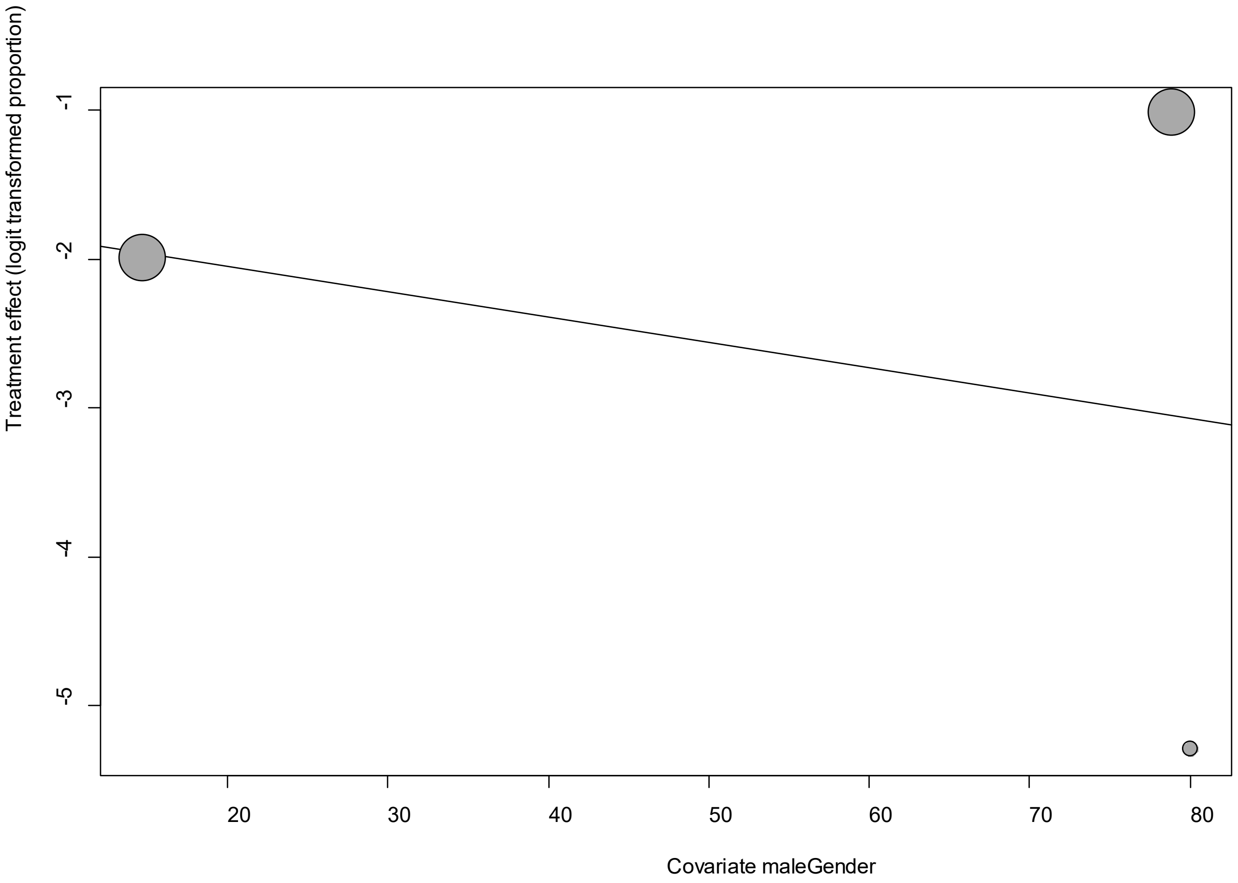


Point sizes are proportional to an inverse of the precision of the estimates.

The regression coefficient was -0.02 (95% confidence interval:-0.13-0.09).(*P*=0.76).

**eFigure 28e. Funnel Plot of studies reporting on Sneezing**

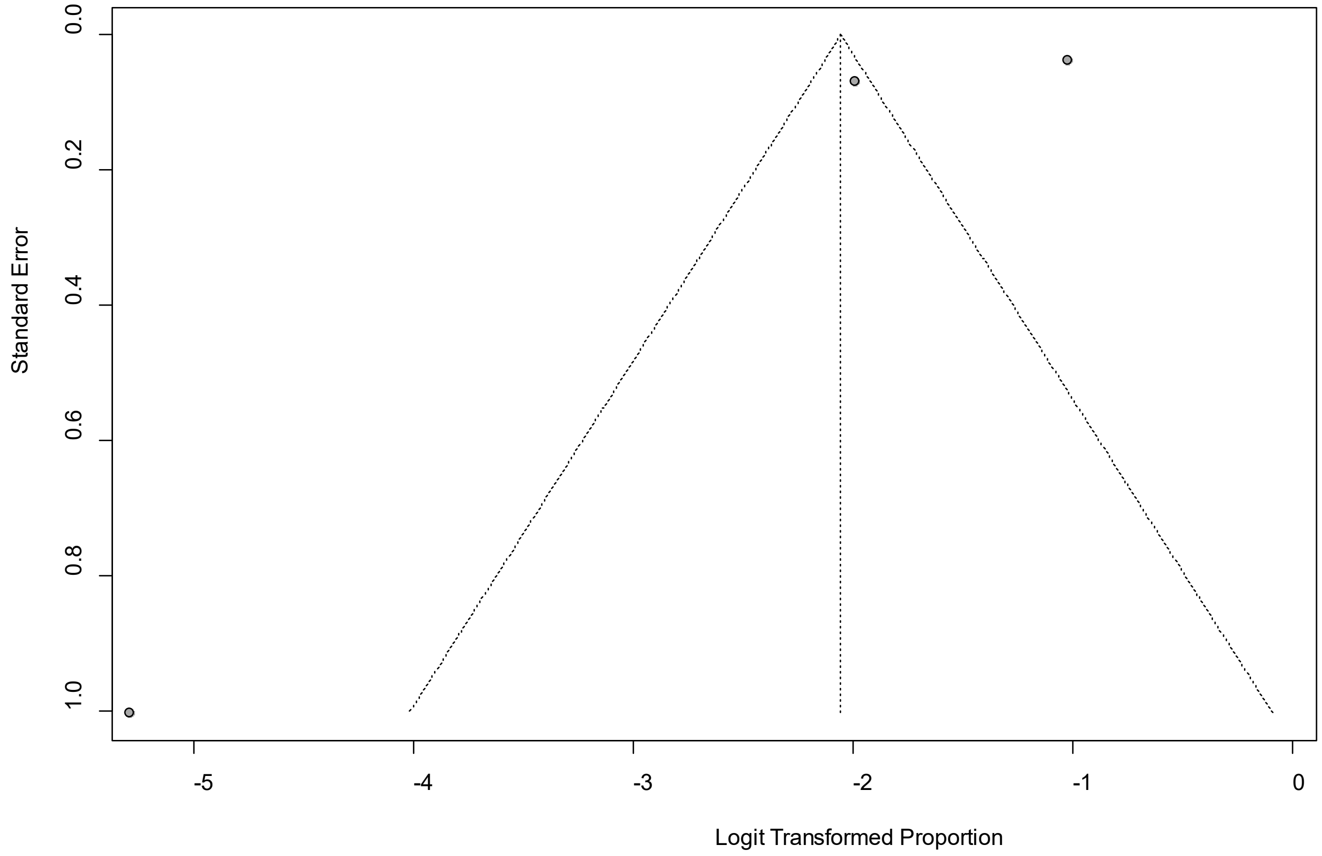


Egger's *P* was 0.56, indicating the absence of publication bias.

### eFigure 29a. Forest Plot on Sputum

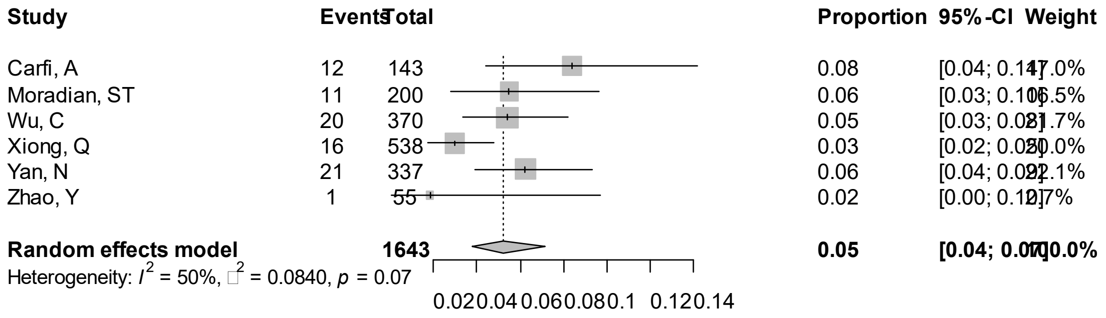


Point sizes are proportional to an inverse of the precision of the estimates.

**eFigure 29b. Bubble Plots (follow-up period) on Sputum**

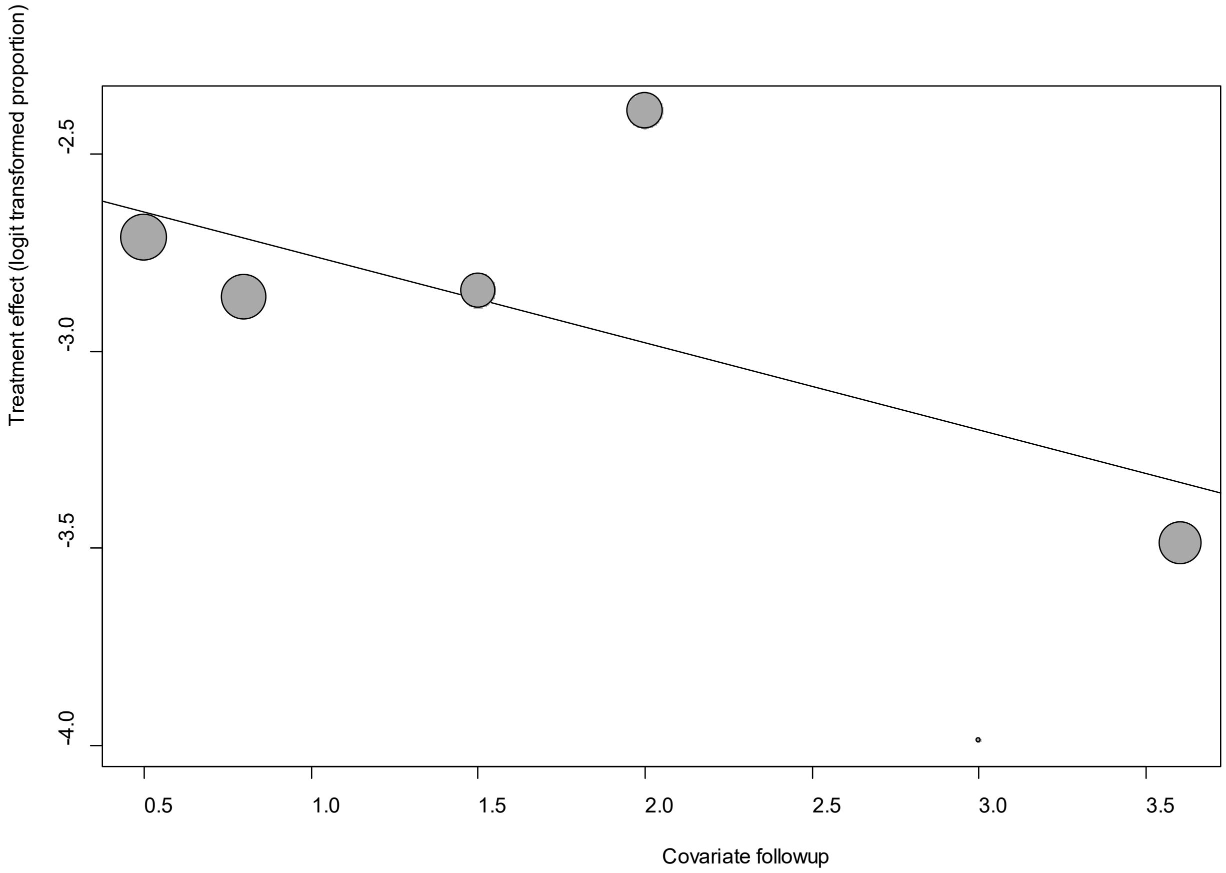


Point sizes are proportional to an inverse of the precision of the estimates.

The regression coefficient was -0.22 (95% confidence interval:-0.46-0.01).(*P*=0.07).

**eFigure 29c. Bubble Plots (age) on Sputum**

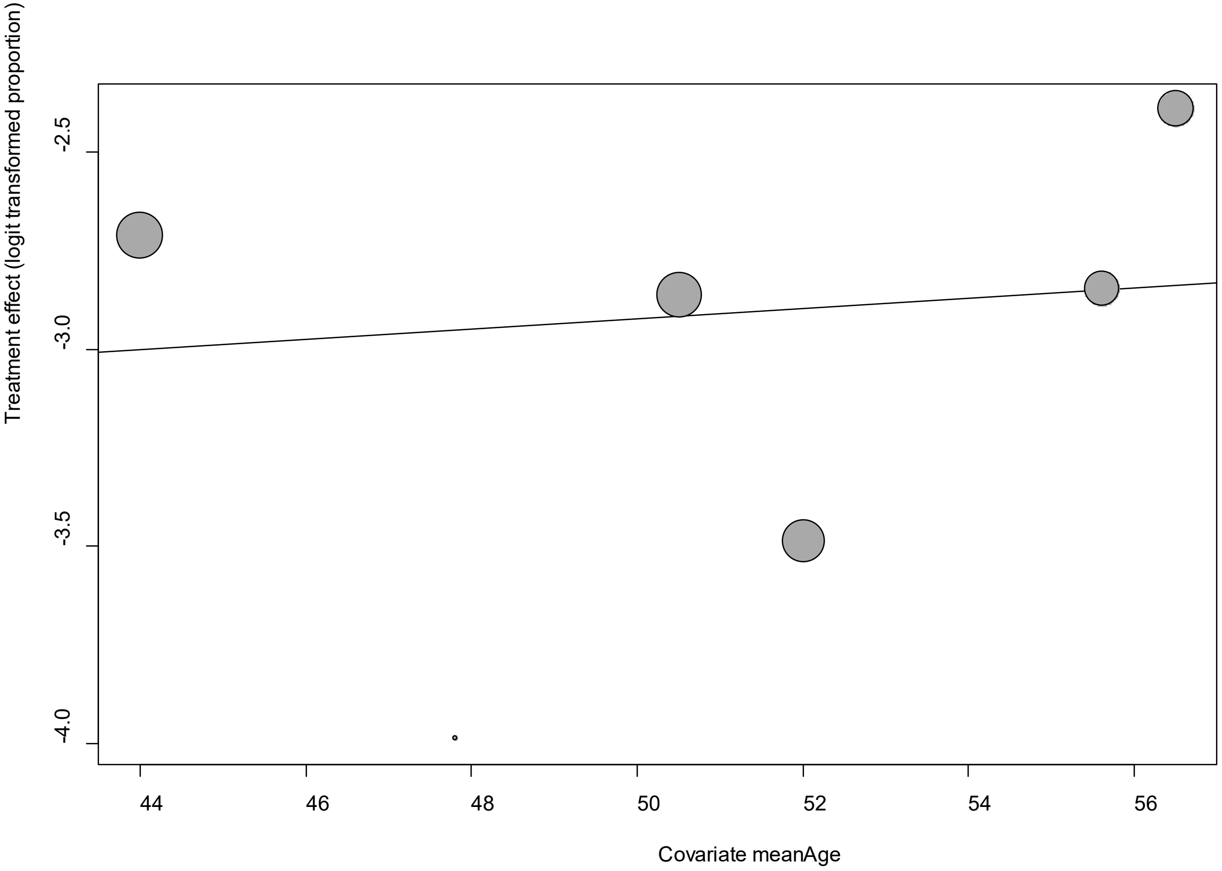


Point sizes are proportional to an inverse of the precision of the estimates.

The regression coefficient was 0.01 (95% confidence interval:-0.07-0.10).(*P*=0.78).

**eFigure 29d. Bubble Plots (sex) on Sputum**

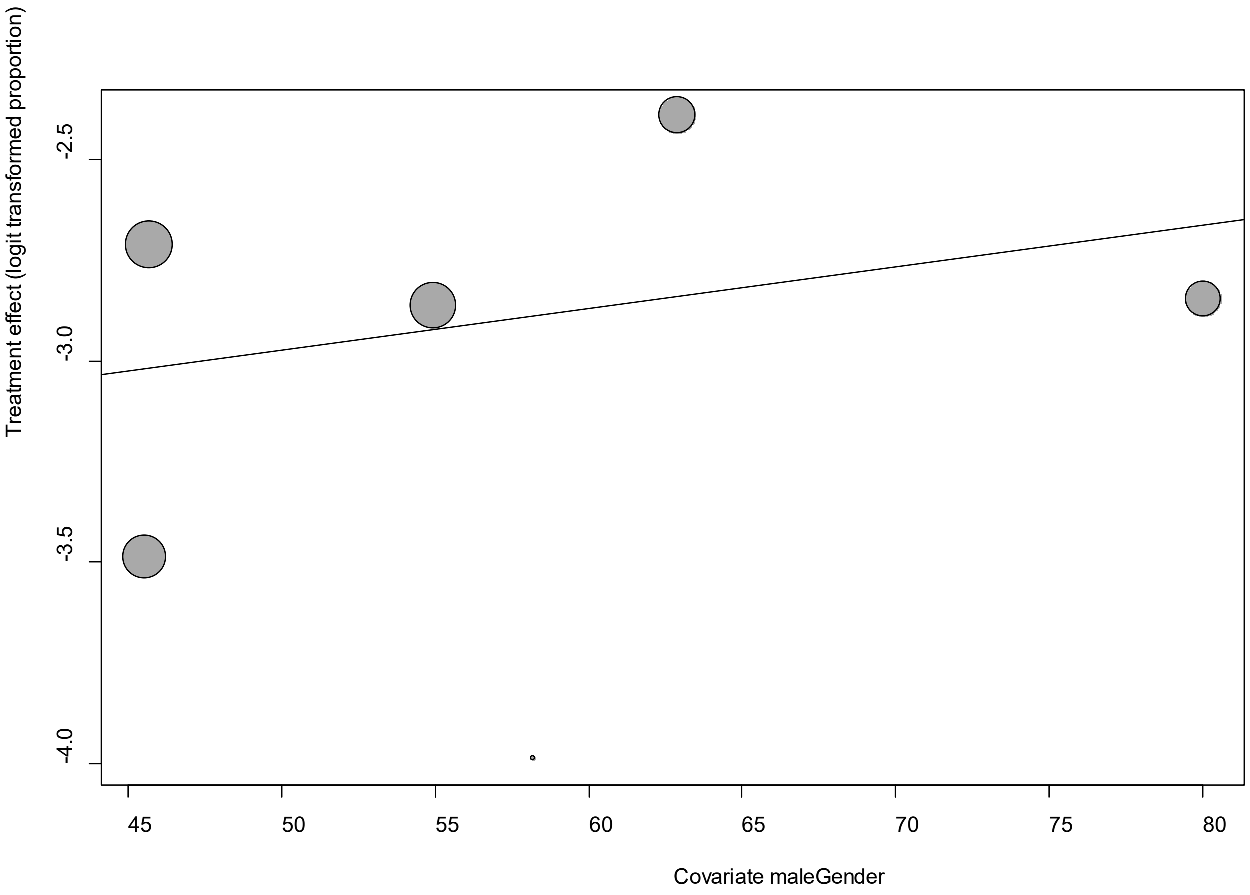


Point sizes are proportional to an inverse of the precision of the estimates.

The regression coefficient was 0.01 (95% confidence interval:-0.02-0.04).(*P*=0.49).

**eFigure 29e. Funnel Plot of studies reporting on Sputum**

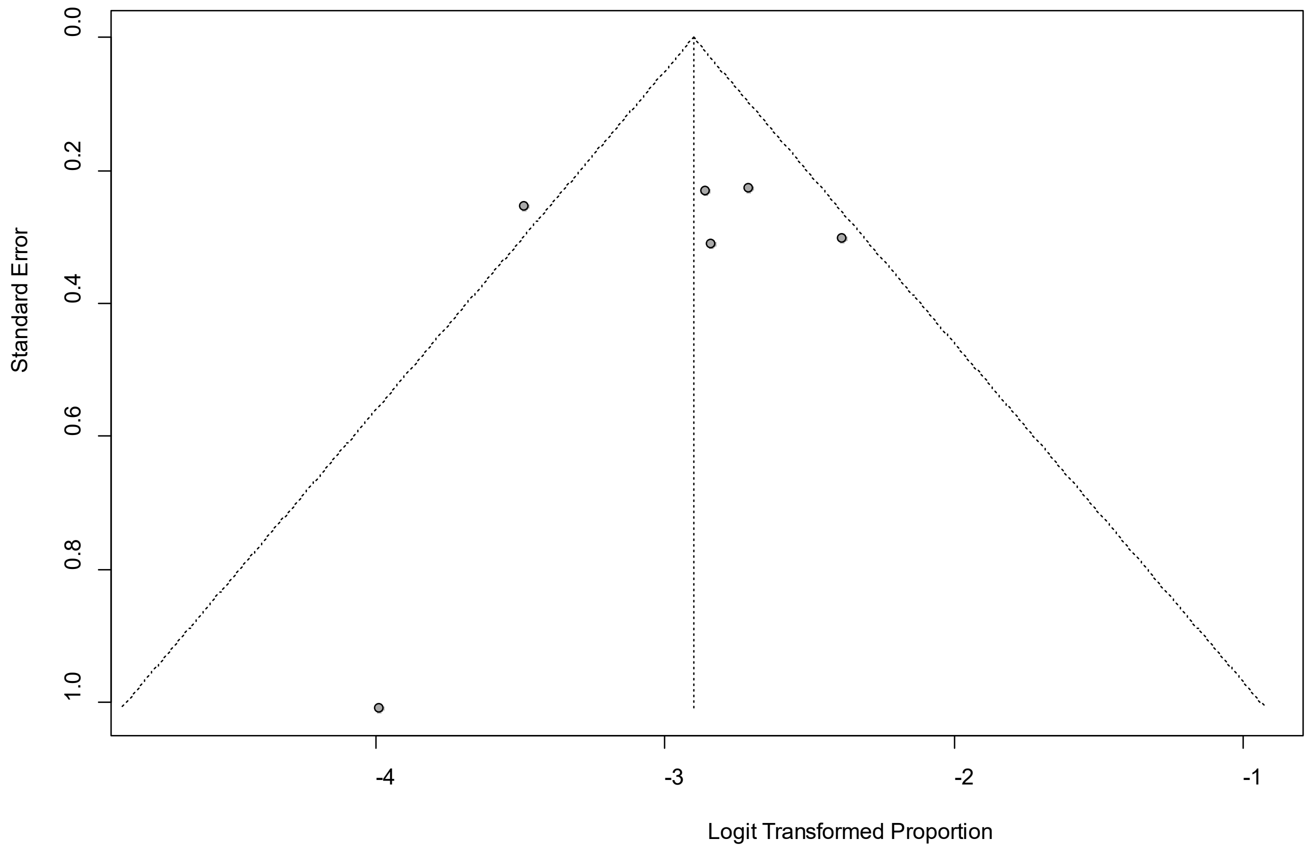


Egger's *P* was 0.63, indicating the absence of publication bias.

### eFigure 30a. Forest Plot on Vertigo (Dizziness)

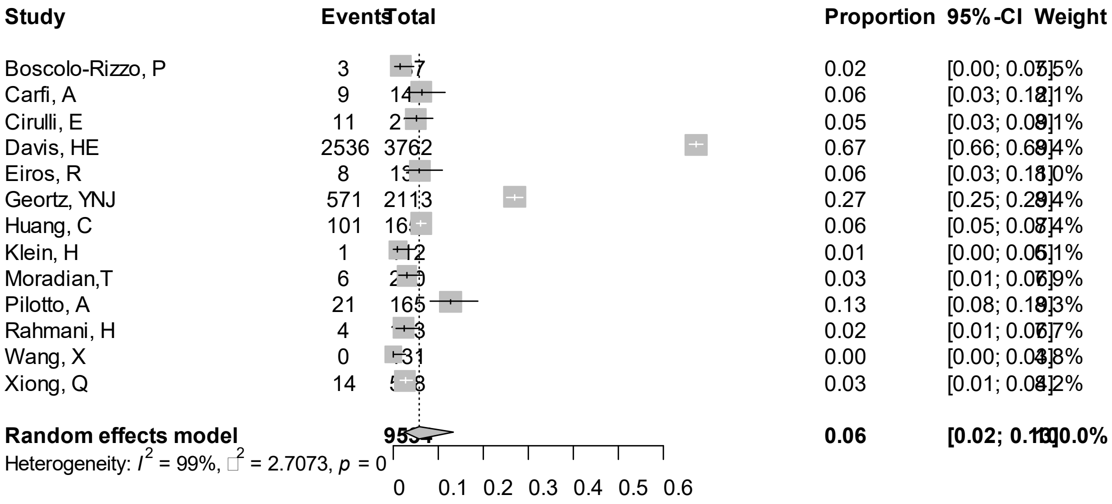


Point sizes are proportional to an inverse of the precision of the estimates.

**eFigure 30b. Bubble Plots (follow-up period) on Vertigo (Dizziness)**

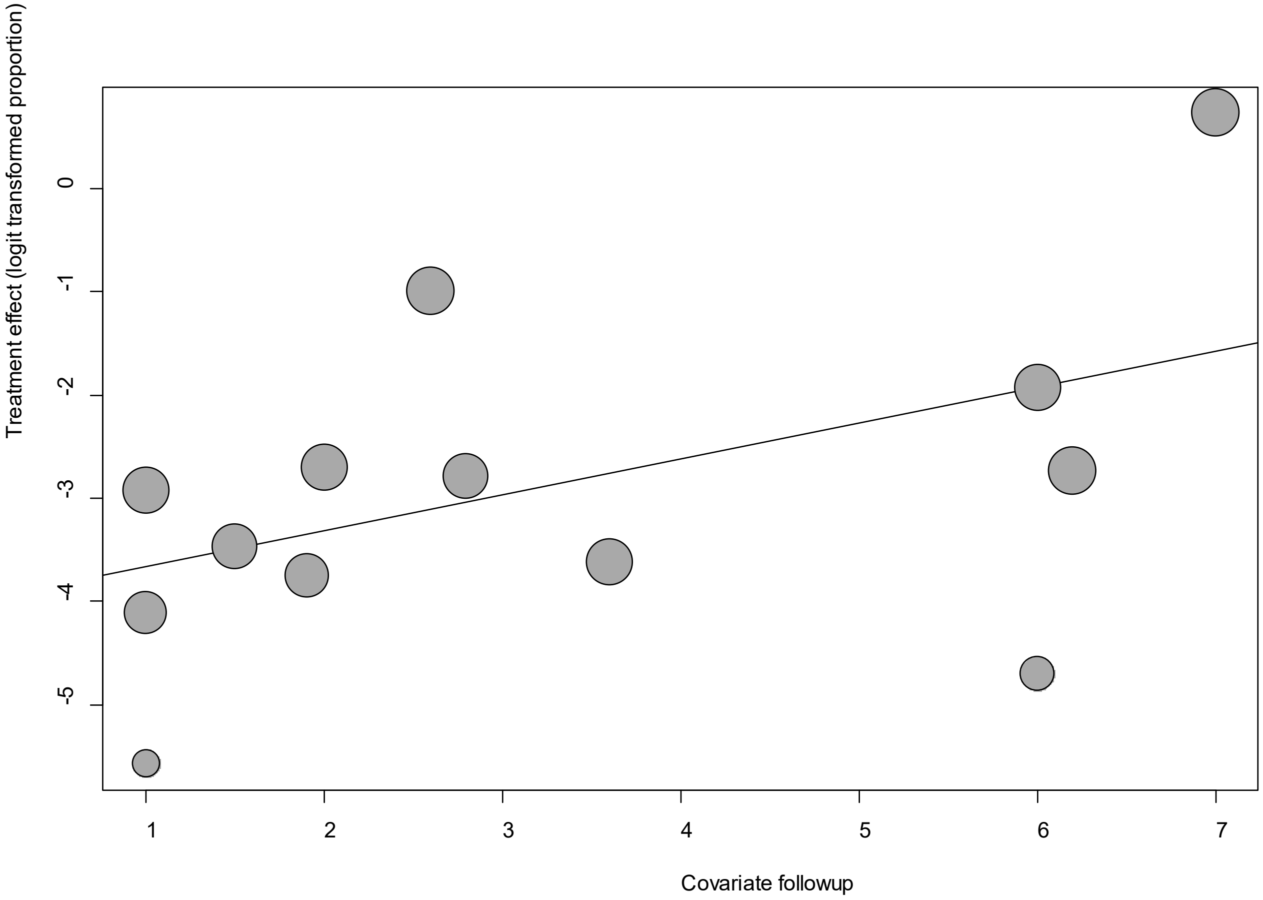


Point sizes are proportional to an inverse of the precision of the estimates.

The regression coefficient was 0.35 (95% confidence interval:-0.17-0.87).(*P*=0.19).

**eFigure 30c. Bubble Plots (age) on Vertigo (Dizziness)**

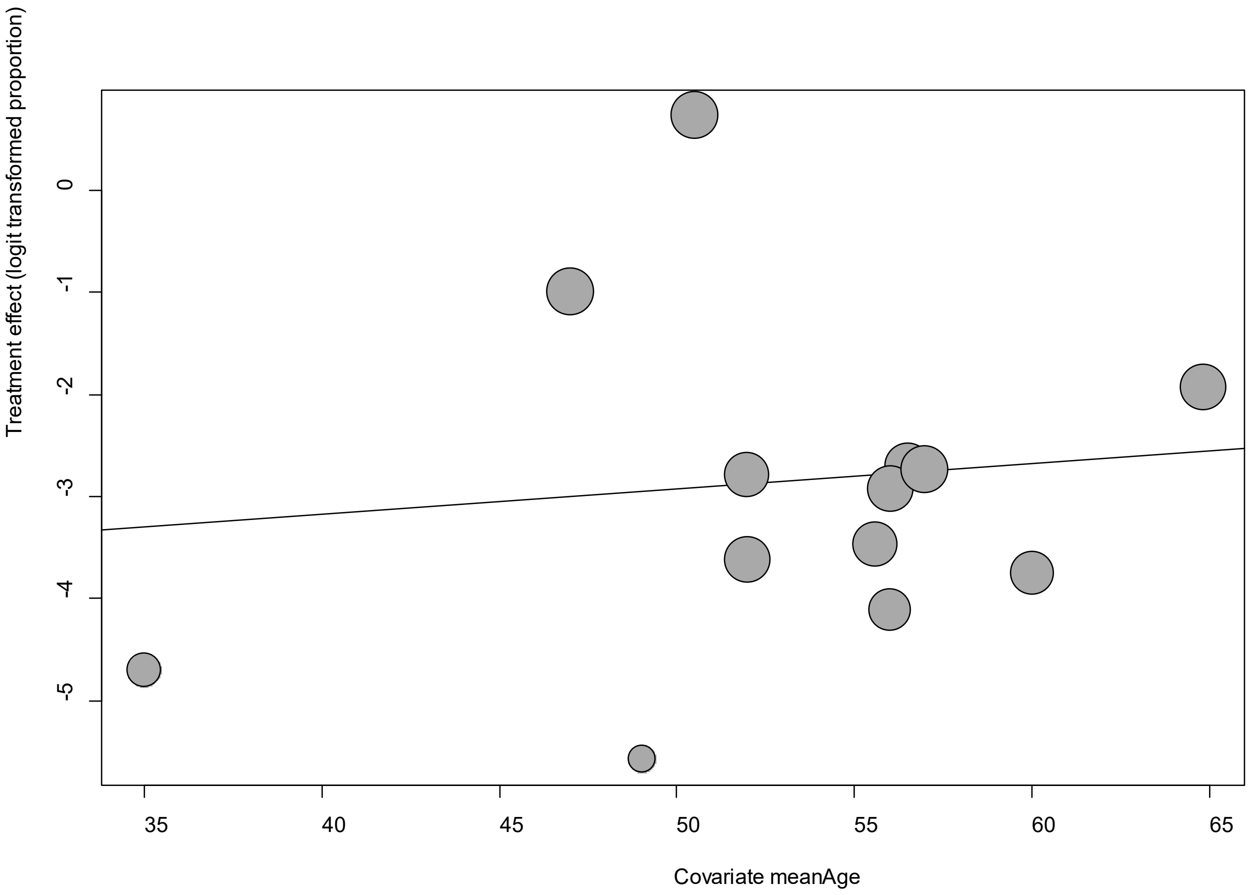


Point sizes are proportional to an inverse of the precision of the estimates.

The regression coefficient was 0.03 (95% confidence interval:-0.13-0.18).(*P*=0.75).

**eFigure 30d. Bubble Plots (sex) on Vertigo (Dizziness)**

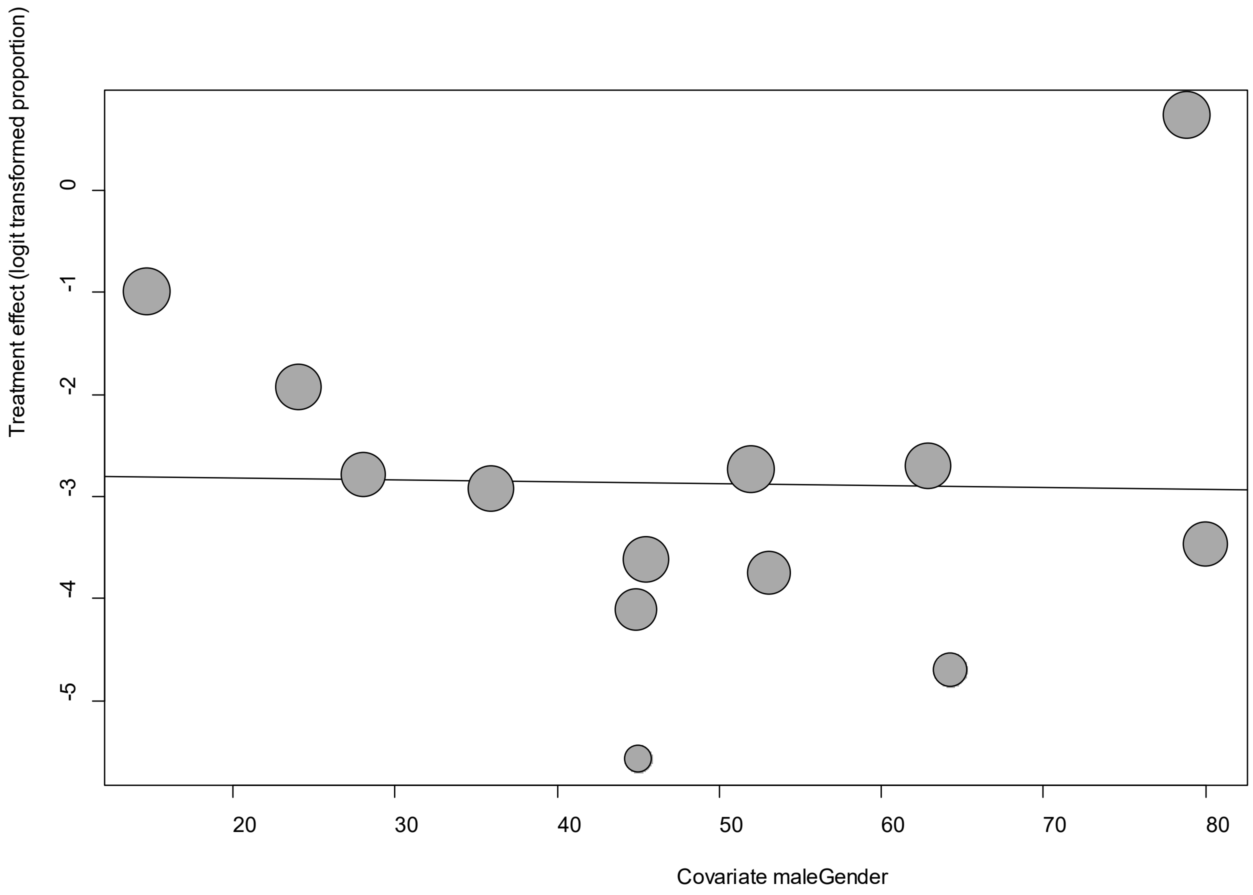


Point sizes are proportional to an inverse of the precision of the estimates.

The regression coefficient was -0.002 (95% confidence interval:-0.06-0.06).(*P*=0.96).

**eFigure 30e. Funnel Plot of studies reporting on Vertigo (Dizziness)**

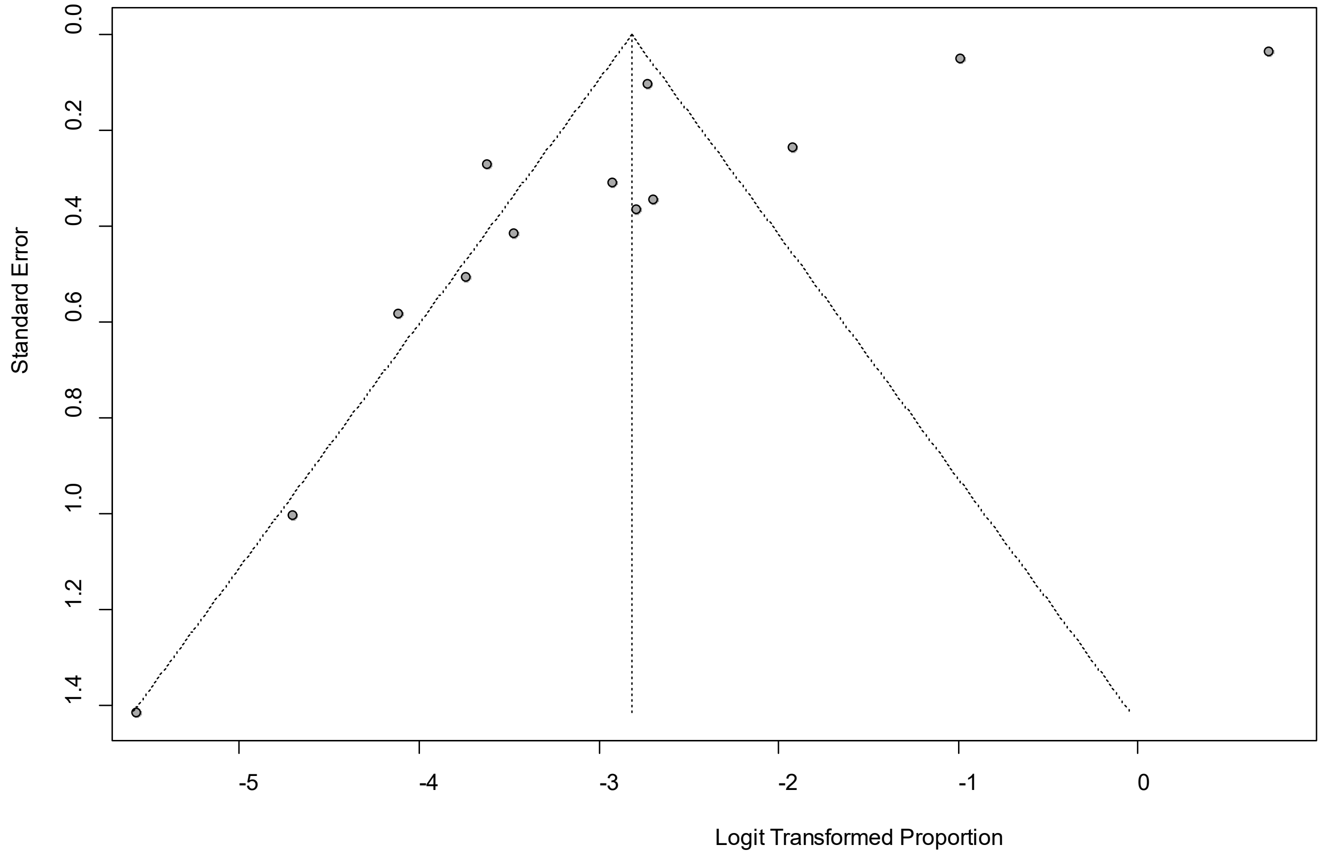


Egger's *P* was 0.014, indicating the absence of publication bias.

### eFigure 31a. Forest Plot on Vomiting

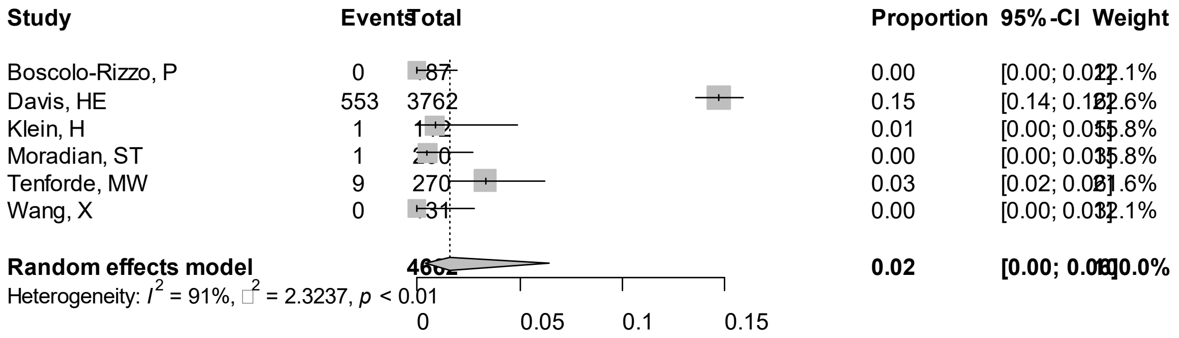


Point sizes are proportional to an inverse of the precision of the estimates.

**eFigure 31b. Bubble Plots (follow-up period) on Vomiting**

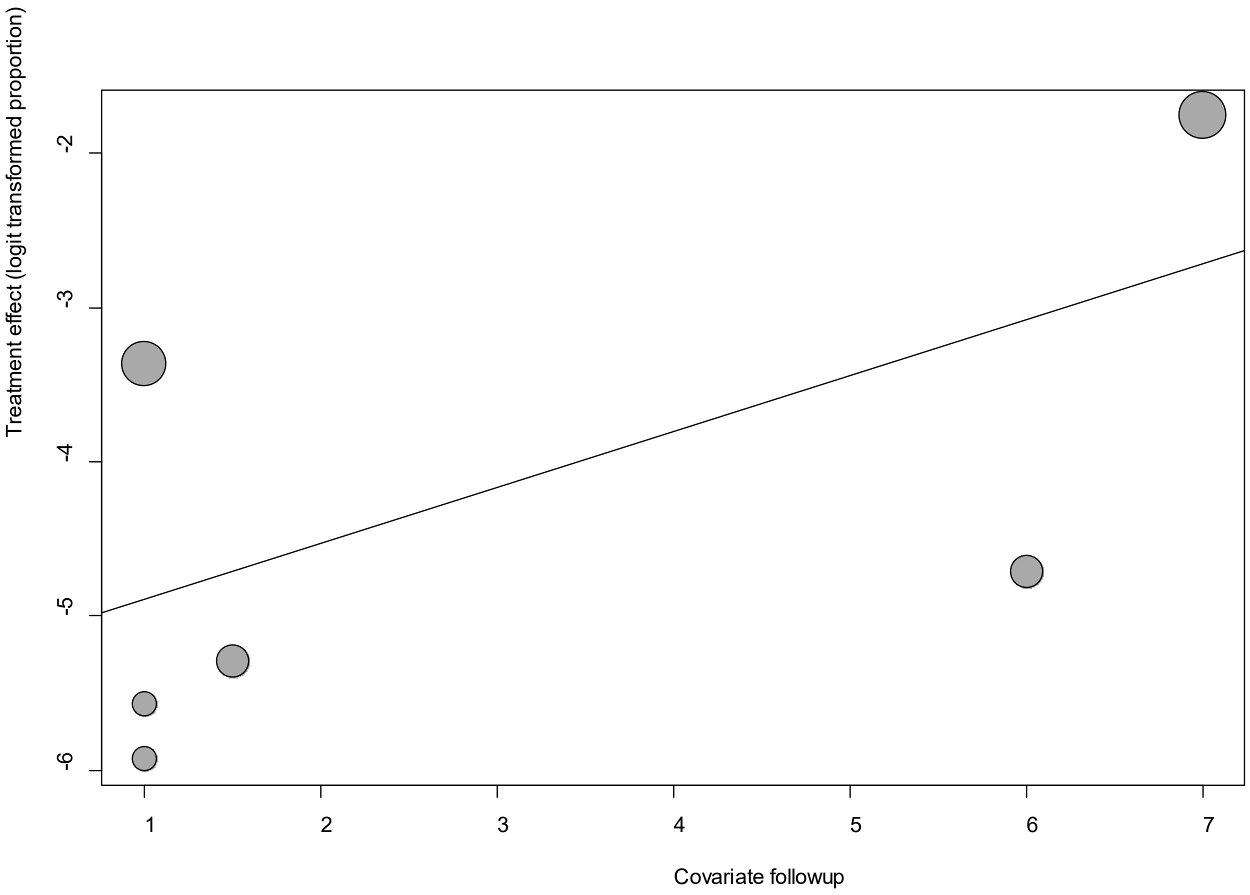


Point sizes are proportional to an inverse of the precision of the estimates.

The regression coefficient was 0.36 (95% confidence interval:-0.15-0.88).(*P*=0.17).

**eFigure 31c. Bubble Plots (age) on Vomiting**

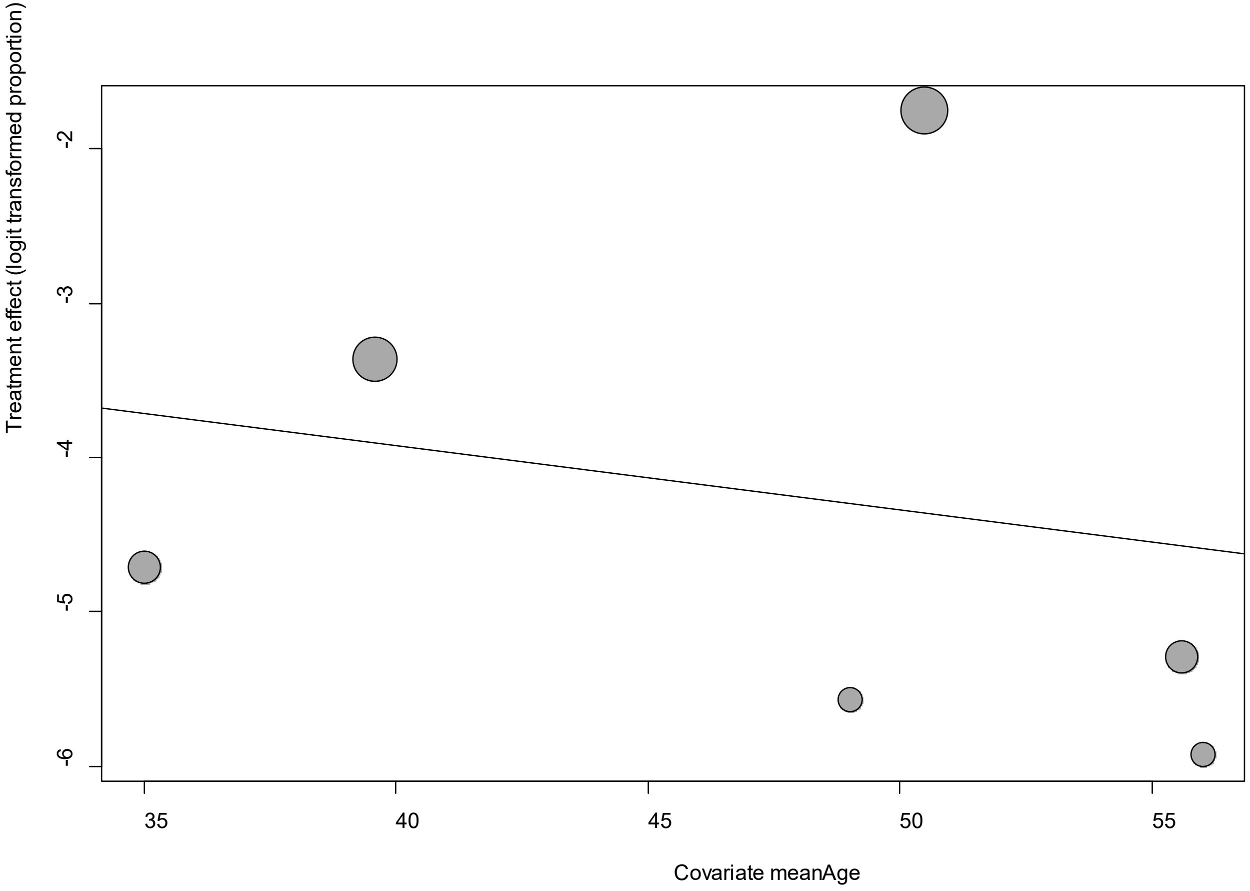


Point sizes are proportional to an inverse of the precision of the estimates.

The regression coefficient was -0.04 (95% confidence interval:-0.27-0.19).(*P*=0.72).

### eFigure 31d. Bubble Plots (sex) on Vomiting

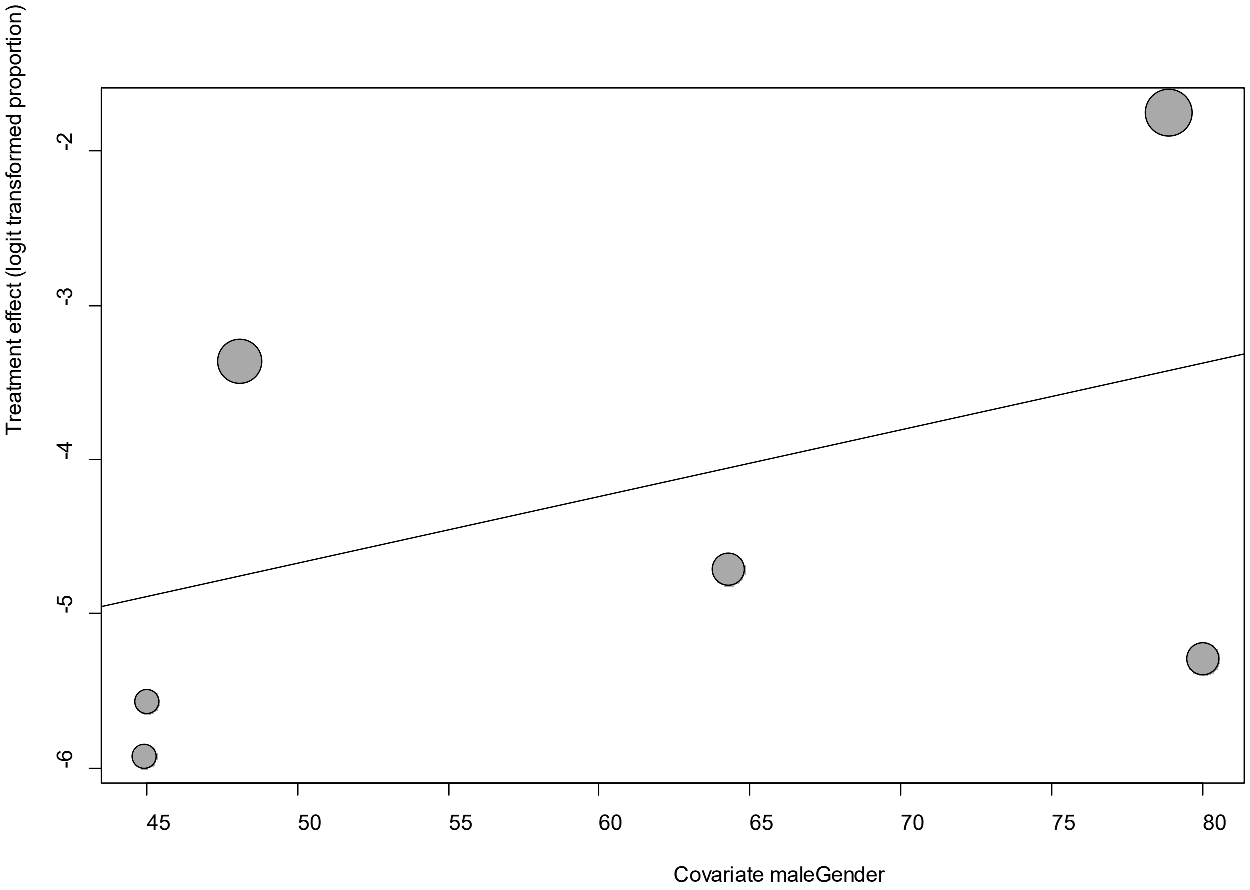


Point sizes are proportional to an inverse of the precision of the estimates.

The regression coefficient was 0.04 (95% confidence interval:-0.06-0.15).(*P*=0.42).

**eFigure 31e. Funnel Plot of studies reporting on Vomiting**

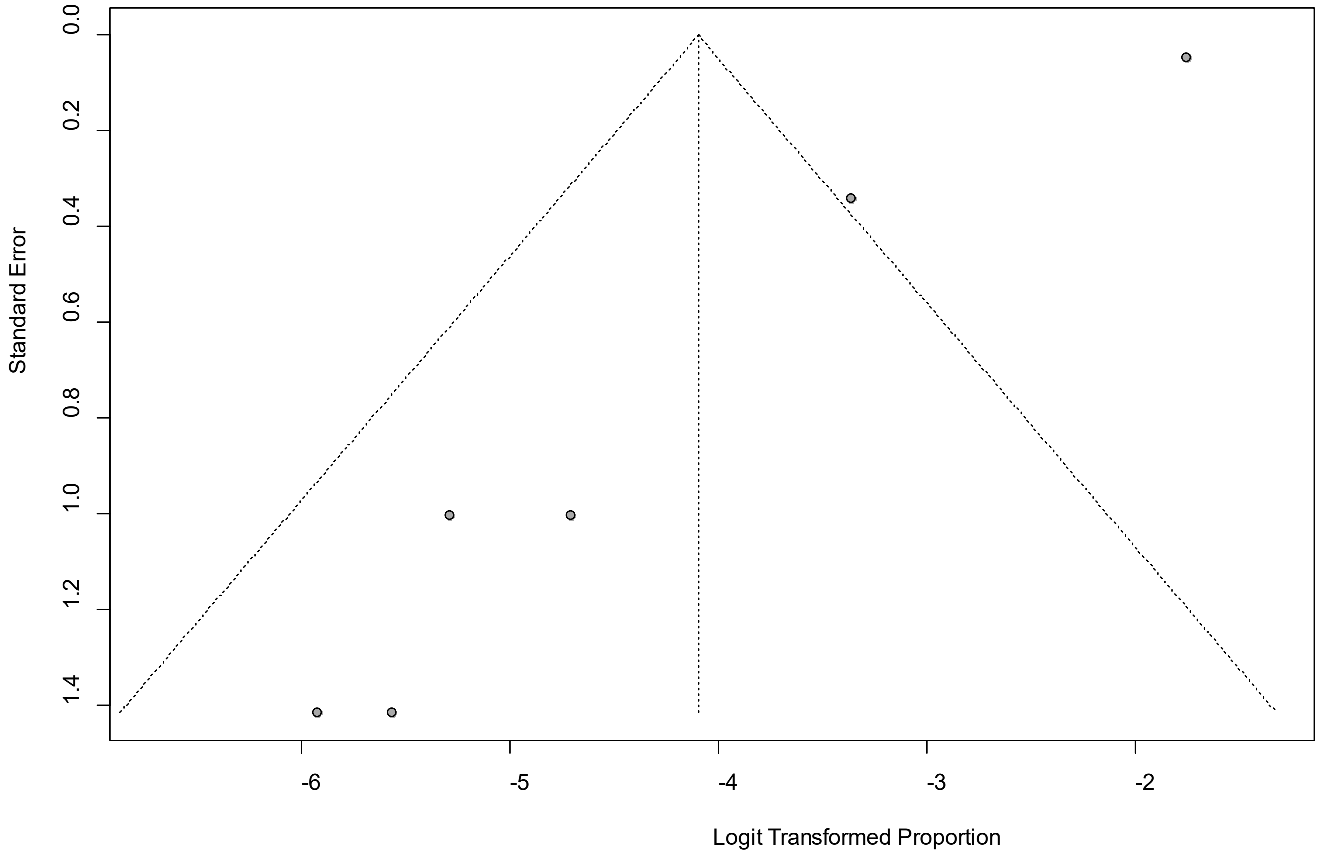


Egger's *P* was 0.002, indicating the presence of publication bias.

**eFigure 32a. Forest Plot on Weakness**

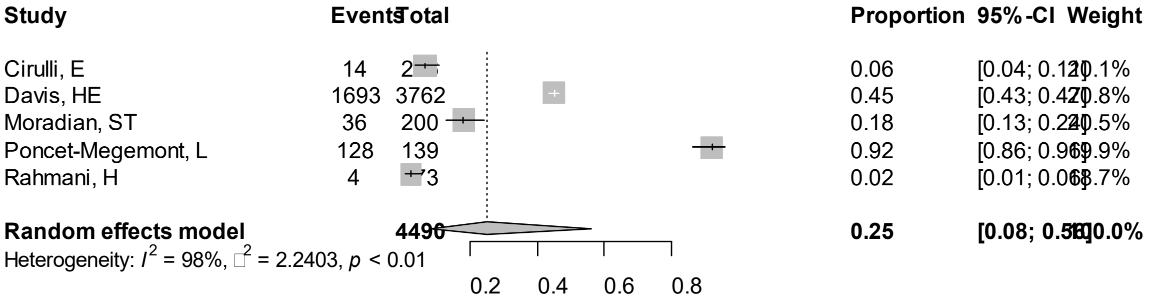


Point sizes are proportional to an inverse of the precision of the estimates.

**eFigure 32b. Bubble Plots (follow-up period) on Weakness**

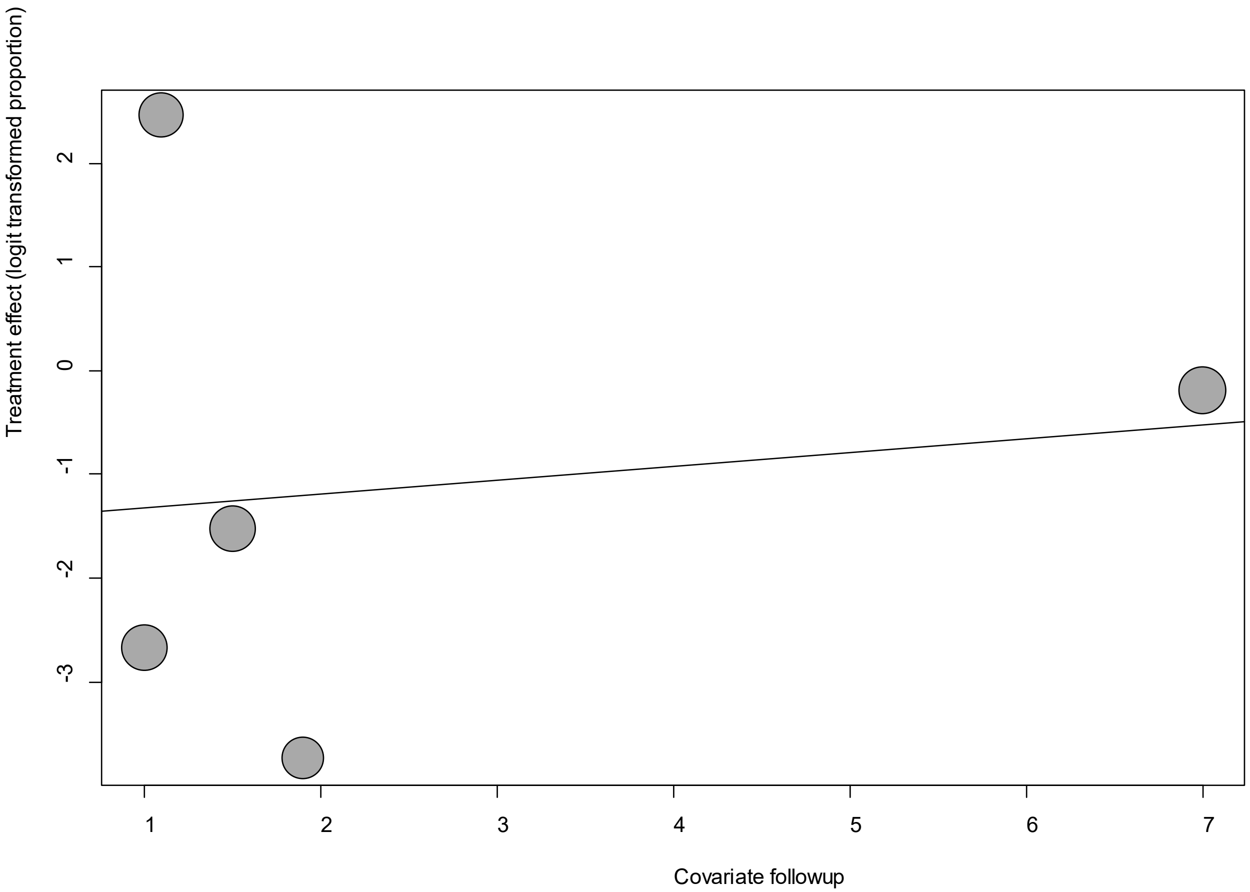


Point sizes are proportional to an inverse of the precision of the estimates.

The regression coefficient was 0.13 (95% confidence interval:-0.75-1.01).(*P*=0.76).

**eFigure 32c. Bubble Plots (age) on Weakness**

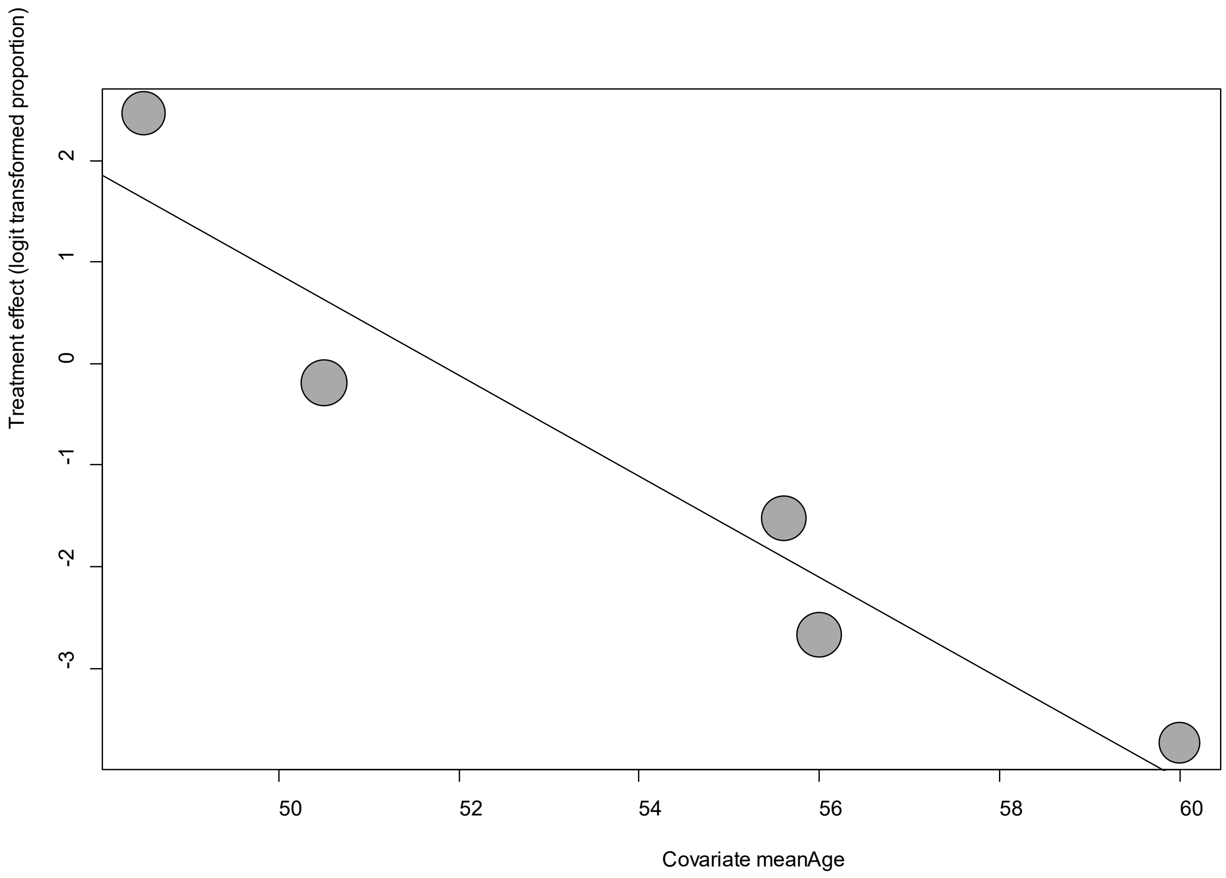


Point sizes are proportional to an inverse of the precision of the estimates.

The regression coefficient was -0.50 (95% confidence interval:-0.70—0.29).(*P*<0.001).

### eFigure 32d. Bubble Plots (sex) on Weakness

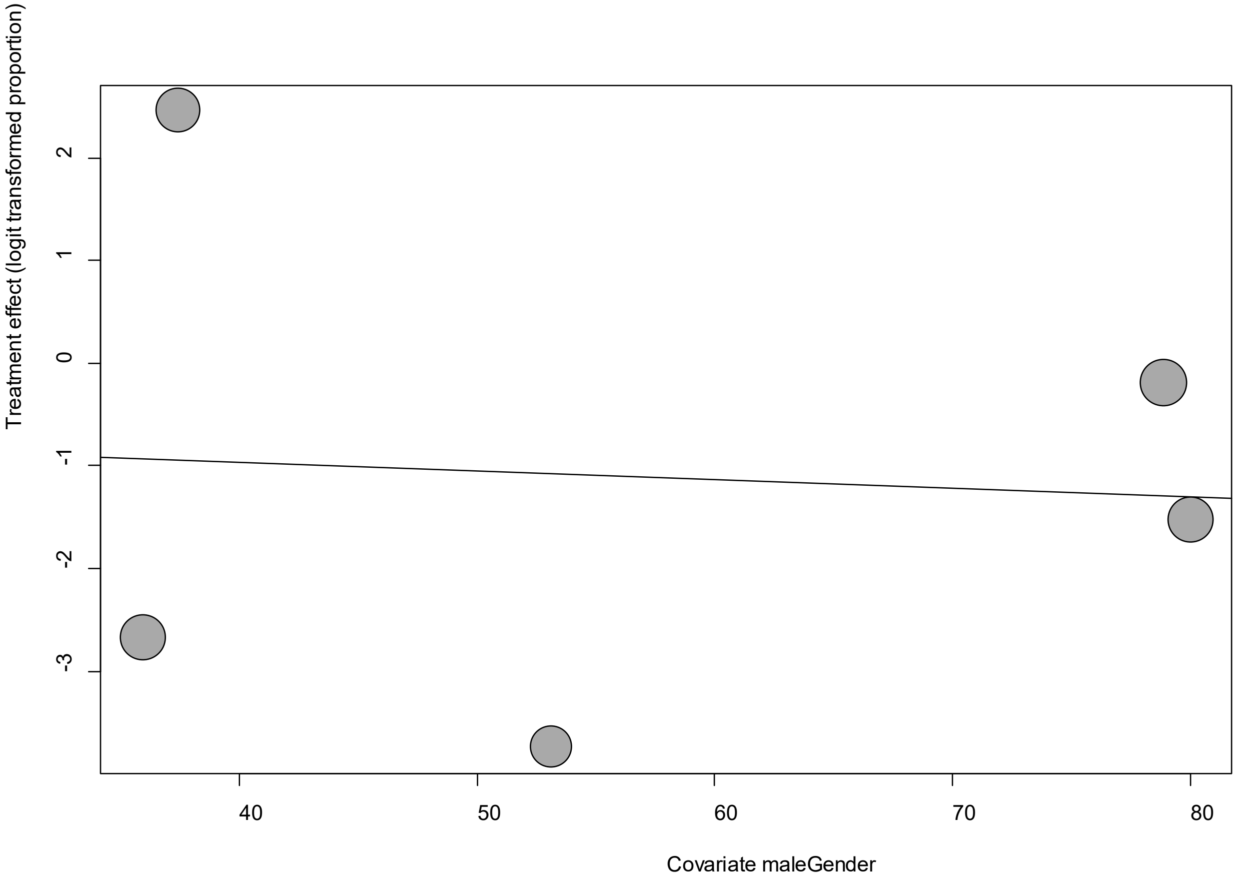


Point sizes are proportional to an inverse of the precision of the estimates.

The regression coefficient was -0.009 (95% confidence interval:-0.09-0.07).(*P*=0.84).

**eFigure 32e. Funnel Plot of studies reporting on Weakness**

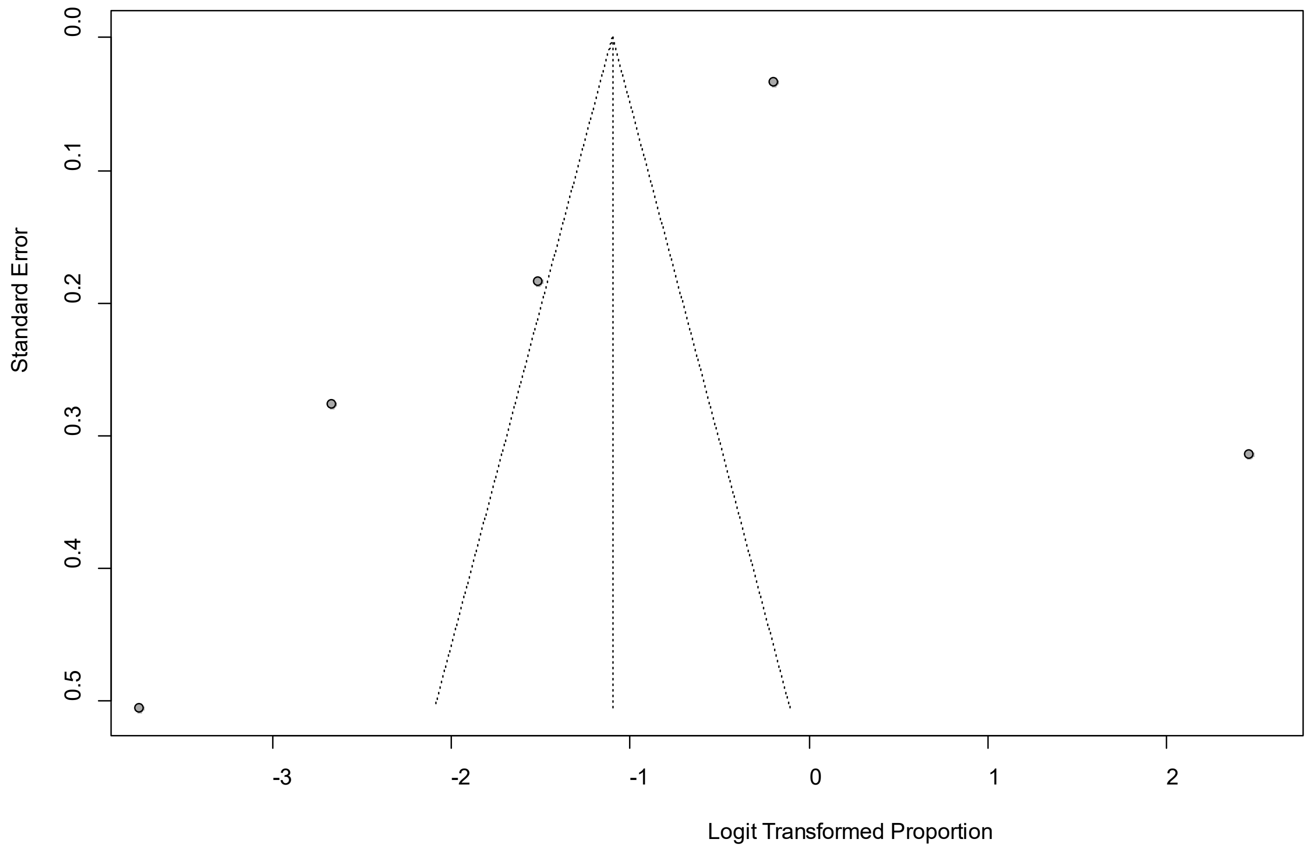


Egger's *P* was 0.46, indicating the absence of publication bias.

### eFigure 33a. Forest Plot on Weight loss

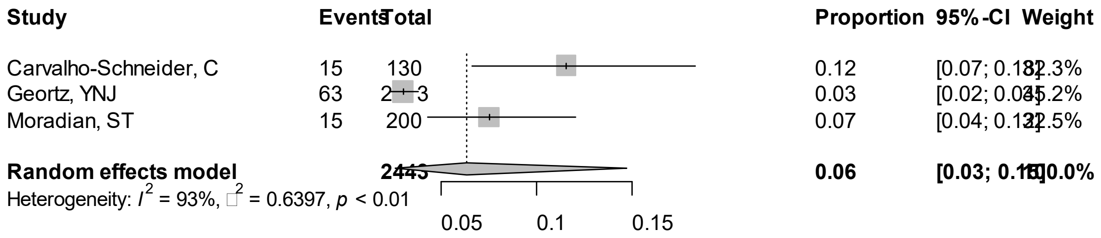


Point sizes are proportional to an inverse of the precision of the estimates.

**eFigure 33b. Bubble Plots (follow-up period) on Weight loss**

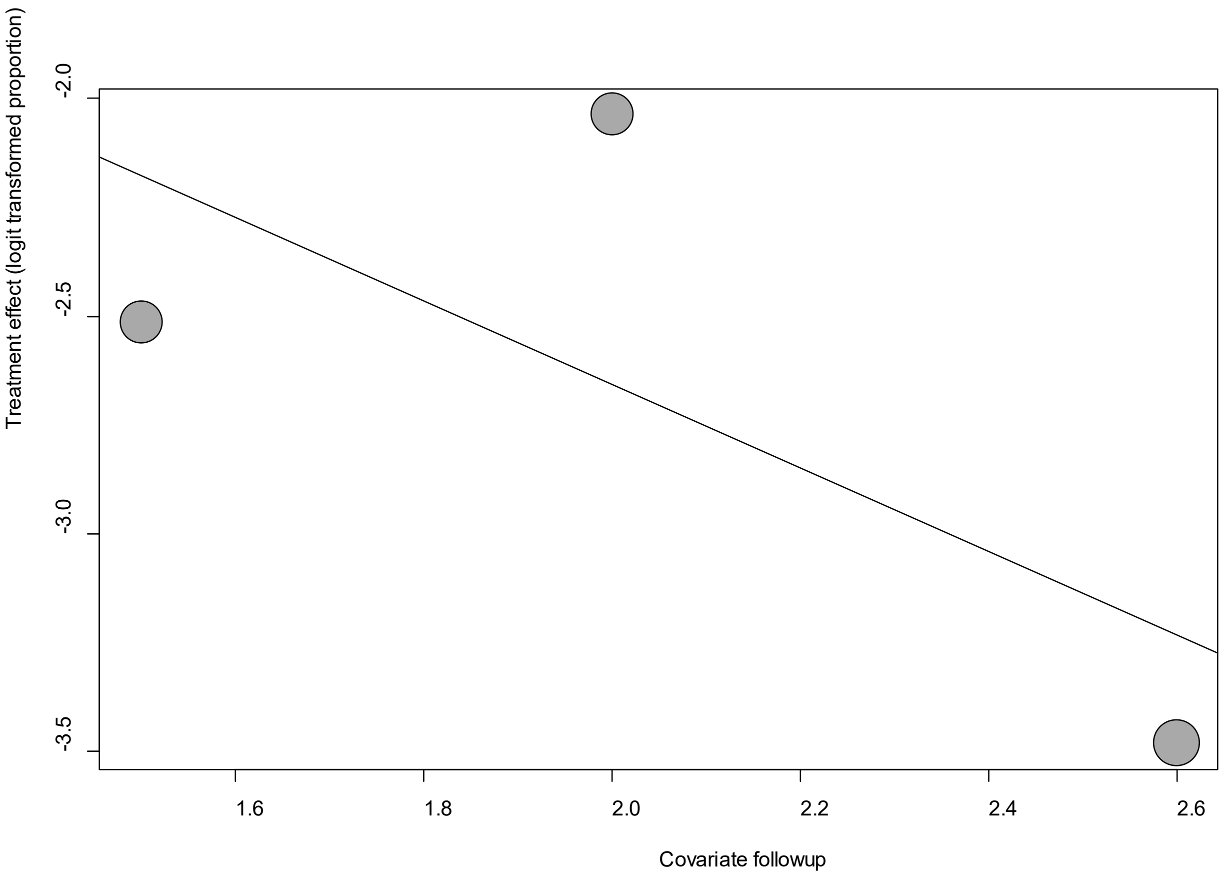


Point sizes are proportional to an inverse of the precision of the estimates.

The regression coefficient was -0.96 (95% confidence interval:-2.80-0.88).(*P*=0.31).

**eFigure 33c. Bubble Plots (age) on Weight loss**

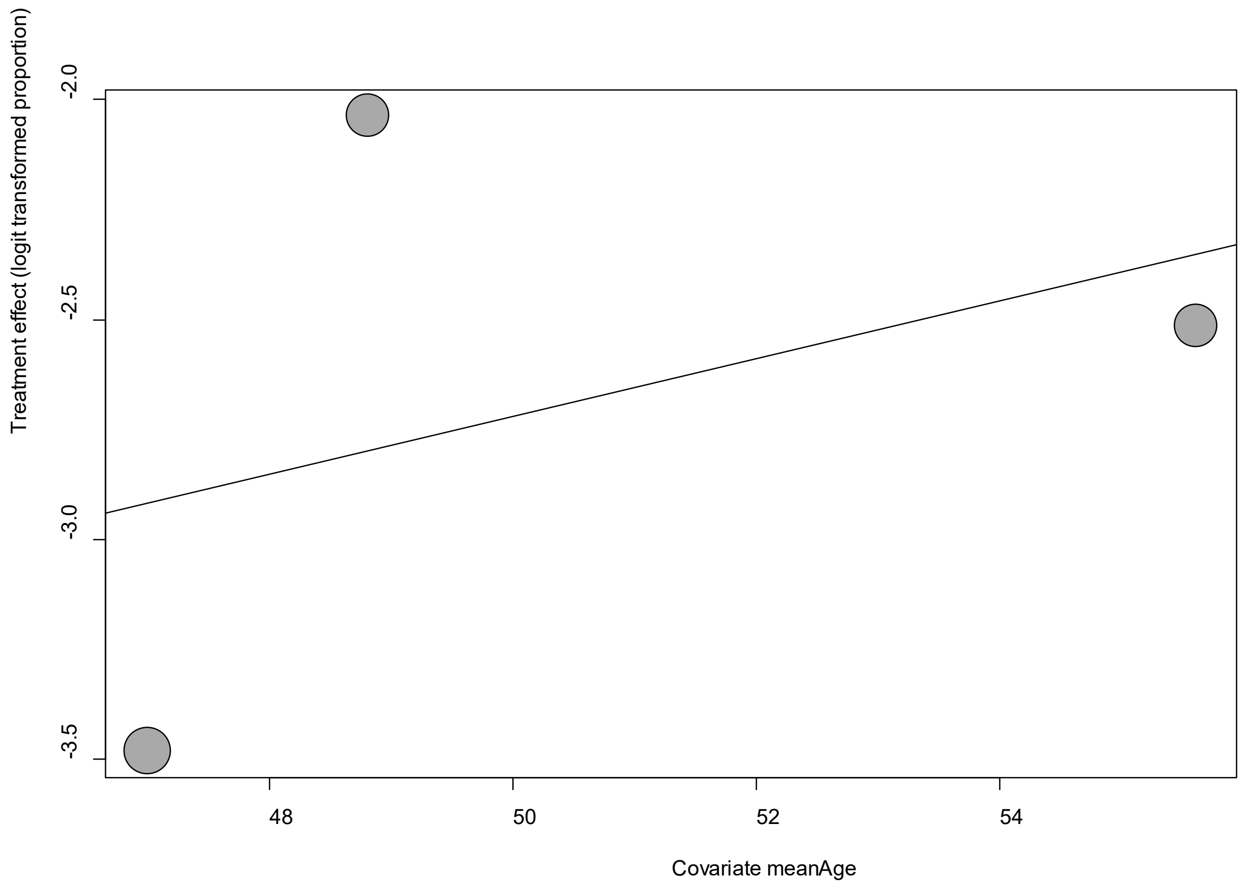


Point sizes are proportional to an inverse of the precision of the estimates.

The regression coefficient was 0.06 (95% confidence interval:-0.23-0.36).(*P*=0.66).

**eFigure 33d. Bubble Plots (sex) on Weight loss**

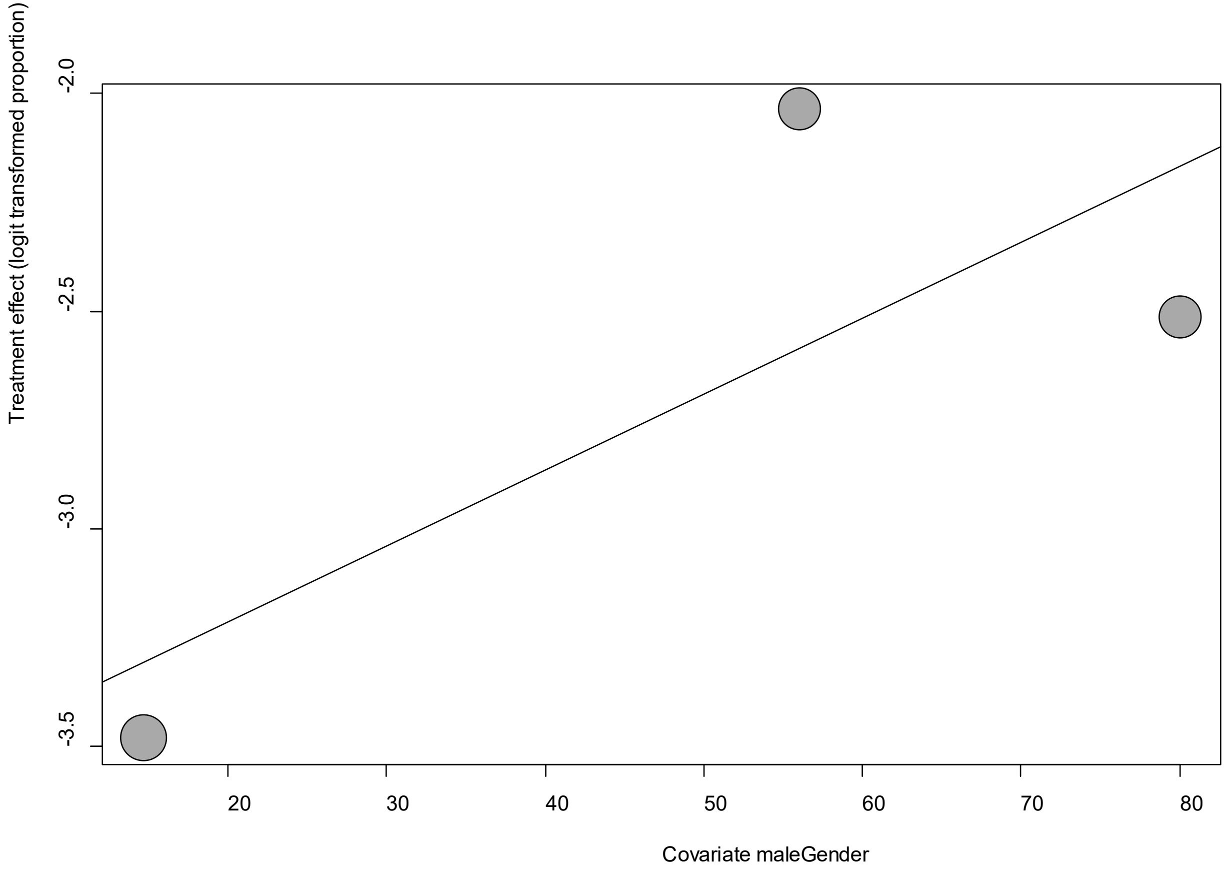


Point sizes are proportional to an inverse of the precision of the estimates.

The regression coefficient was 0.02 (95% confidence interval:-0.01-0.45).(*P*=0.21).

**eFigure 33e. Funnel Plot of studies reporting on Weight loss**

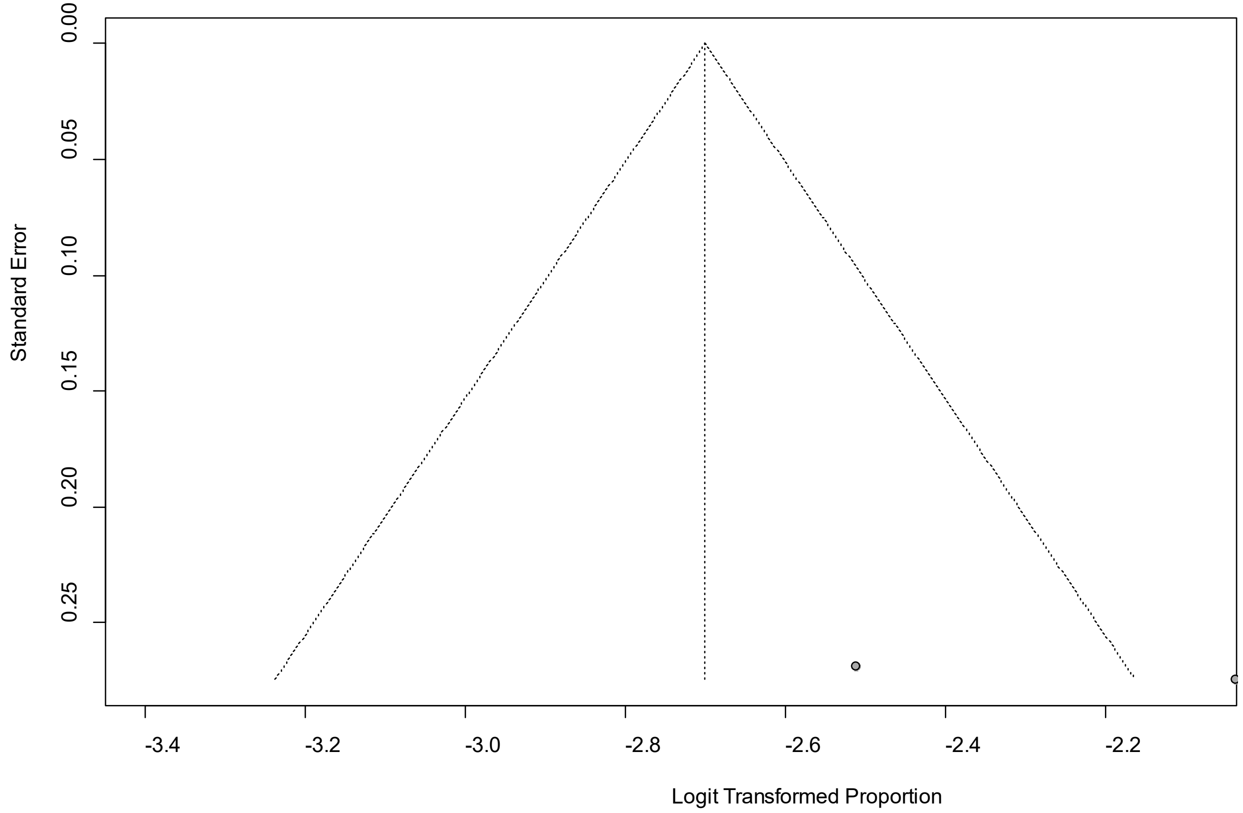


Egger's *P* was 0.13, indicating the absence of publication bias.
